# Rare Cholesterol Related Disorders – A Sterolomic Library for Diagnosis and Monitoring of Diseases

**DOI:** 10.1101/2025.06.23.25328695

**Authors:** Mohsen Ali Asgari, Eylan Yutuc, Jonas Abdel-Khalik, Peter J. Crick, Andrew A Morris, Simon A Jones, Arunabha Ghosh, Richard Curnock, Claire Hart, Ludger Schöls, Silke Matysik, Ioanna Laina, Evan Reid, Thomas Warner, Belen Gonzalez-Herrero, Rajith De Silva, W. Owen Pickrell, Stuart J Moat, William J Griffiths, Yuqin Wang

## Abstract

Cholesterol is an essential molecule in all animals, it can be made by *de novo* synthesis and can be taken up from the diet. Inherited disorders of cholesterol synthesis, metabolism and transport lead to disease, often with neurological signs. However, such disorders tend to have non-specific symptoms and can be difficult to diagnose. In addition, there is no single diagnostic test applicable to multiple disorders of cholesterol synthesis, metabolism and transport which can be used to suggest or confirm a diagnosis, resulting in a delay in treatment, particularly in the case of unknown genetic variants. Here, we present the first version of a mass spectrometry sterolomic library to aid the diagnosis of manifold cholesterol-related inherited disorders of metabolism. The library was generated using technology based on simple derivatisation chemistry exploiting the Girard P hydrazine reagent and utilising electrospray ionisation mass spectrometry in the positive and negative modes. The library includes data for 13 autosomal recessive disorders and predicted data for a further 8 disorders.

## INTRODUCTION

Inherited disorders of cholesterol synthesis, metabolism and transport tend to be autosomal recessive disorders where a gene variant is inherited from both parents. Mass spectrometry (MS) has been used for decades to diagnose such disorders and is usually performed in dedicated laboratories upon the request of a specialist clinician (1–5).

Of the inherited disorders of cholesterol biosynthesis, Smith-Lemli-Opitz syndrome (SLOS, OMIM: 270400) resulting from a deficiency in the enzyme 7-dehydrocholesterol reductase (DHCR7) is the most prevalent (Figure 1), despite being a rare disorder. SLOS often presents soon after birth and affects 1 in 20,000 to 1 in 60,000 new-borns, being most common in Europeans (6). A deficiency in DHCR7 will result in an accumulation of the enzyme substrate (7-dehydrocholesterol, 7-DHC and its isomer 8(9)-dehydrocholesterol, 8-DHC, see Supplemental Table S1 for a list of dysregulated metabolites, with abbreviations, for each disorder) and a reduced concentration of the enzyme product (cholesterol), in blood and tissues (7, 8). In a specialist clinical laboratory, this can be easily determined either by gas chromatography (GC) – MS or liquid chromatography (LC) – MS (7–9). At present there is no cure for SLOS, but there are treatments including dietary supplementation of cholesterol and oral bile acid therapy which may improve some of the symptoms (10, 11).

**Figure 1.**
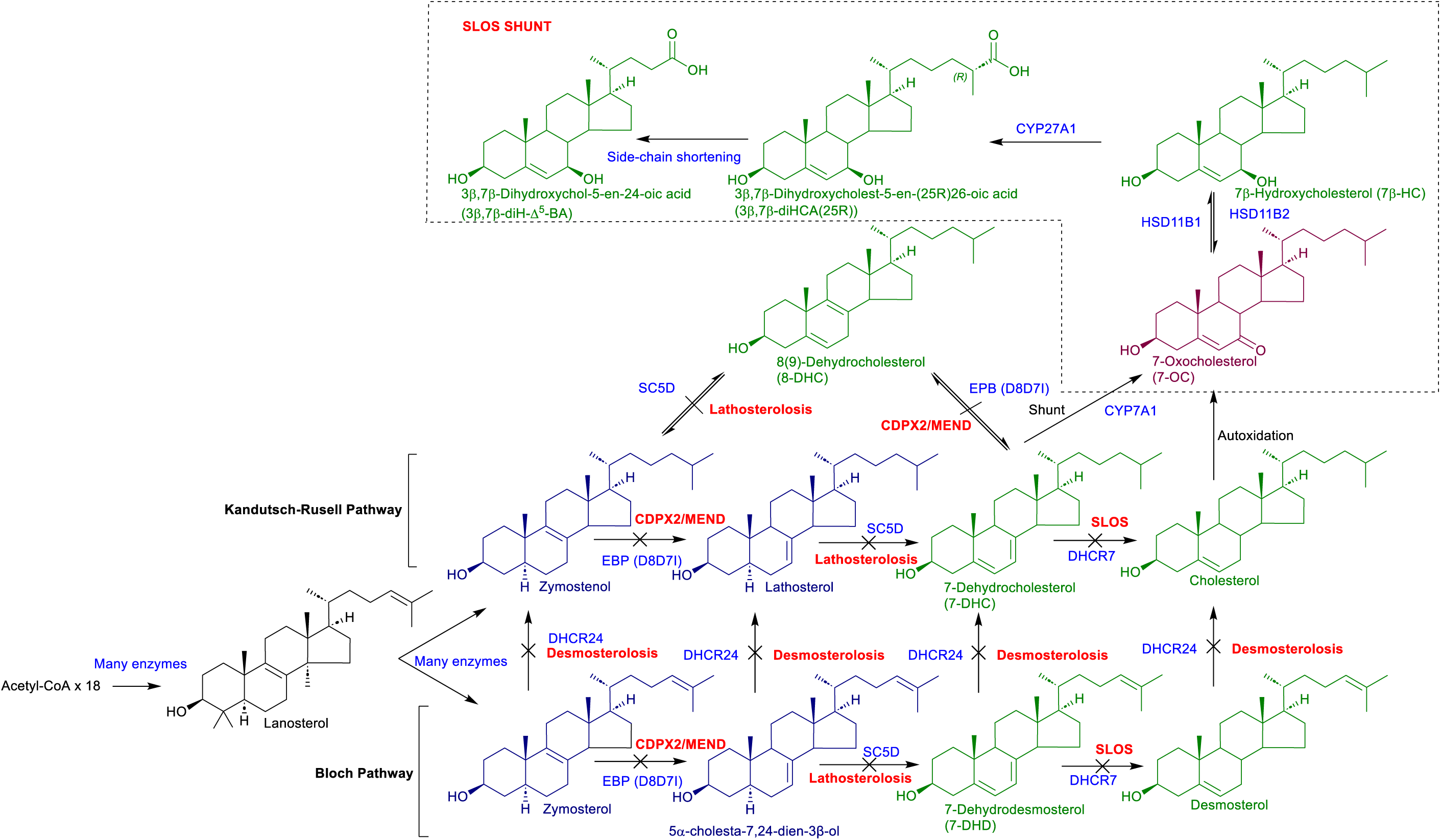
Simplified scheme of cholesterol biosynthesis beyond lanosterol and, shown in the dashed box, metabolism of 7-DHC following oxidation by CYP7A1 in a shunt pathway. Enzymes are shown in blue and metabolic disorders in bold red. Metabolites in blood (plasma/serum) observed by LC-MS(MS^n^) after treatment with cholesterol oxidase and derivatisation with GP reagent are shown in green, metabolites observed following GP derivatisation in the absence of cholesterol oxidase treatment are shown in claret. The metabolites shown in dark blue with a 3β-hydroxy-5α-hydrogen configuration are also substrates for cholesterol oxidase and GP derivatisation but are of low abundance in blood and only observed when greatly elevated. Enzyme abbreviations with synthesis disorders in parenthesis: EBP, emopamil-binding protein, (CDPX2, X-linked dominant chondrodysplasia punctata-2; MEND, male EBP disorder with neurological abnormalities); SC5D, sterol C-5 desaturase, (lathosterolosis); DHCR7, 7-dehydrocholesterol reductase, (SLOS, Smith-Lemli-Opitz syndrome); DHCR24, 24-dehydrocholesterol reductase, (desmosterolosis). Other enzymes: CYP7A1, cytochrome P450 family 7 subfamily A member 1; HSD11B1, hydroxysteroid 11-beta dehydrogenase 1; HSD11B2, hydroxysteroid 11-beta dehydrogenase 2; CYP27A1, cytochrome P450 family 27 subfamily A member 1. Abbreviations: GP, Girard P; LC-MS(MS^n^), liquid chromatography – mass spectrometry with multistage fragmentation.

Cerebrotendinous xanthomatosis (CTX, OMIM: 213700) was the first inherited disorder of cholesterol metabolism to be described in the medical literature (12), it is due to a deficiency of the enzyme sterol (25R)26-hydroxylase (also called sterol 27-hydroxylase, CYP27A1, Figure 2) (2, 3, 13, 14). Like SLOS, CTX is autosomal recessive. Its predicted prevalence ranges from 1 in 40,000 to 1 in 500,000 in different populations, but less than a thousand cases have been reported (15–18). It can be diagnosed by elevated 5α-cholestan-3β-ol (cholestanol) in plasma (whenever the noun plasma is used serum is equally applicable) and low concentrations of (25R)26-hydroxycholesterol (26-HC, also called 27-hydroxycholesterol, 27-HC) by GC-MS or by elevated concentrations of bile alcohol glucuronides (Figure 2, see CTX Shunt, see Supplemental Table S1 for biomarkers for different disorder) in plasma or urine by direct infusion (DI)-MS or LC-MS (1–3, 5, 15, 19, 20). Note, in this article we adopt systematic nomenclature and the name (25R)26-hydroxycholesterol (26-HC) (21). Unless stated otherwise stereochemistry about C-25 is assumed to be 25R. Where both or either epimers are specifically referred to, the (25R/S) descriptor is used. High concentrations of the oxysterols 7α-hydroxycholest-4-en-3-one (7α-HCO, also known as C4) and 7α,12α-dihydroxycholest-4-en-3-one (7α,12α-diHCO) in plasma, which can be measured by GC-MS or LC-MS, are also diagnostic of CTX (22–28). CTX can be treated by oral bile acid replacement therapy which may halt disease progression (2, 3, 29), emphasising the importance of early detection.

**Figure 2.**
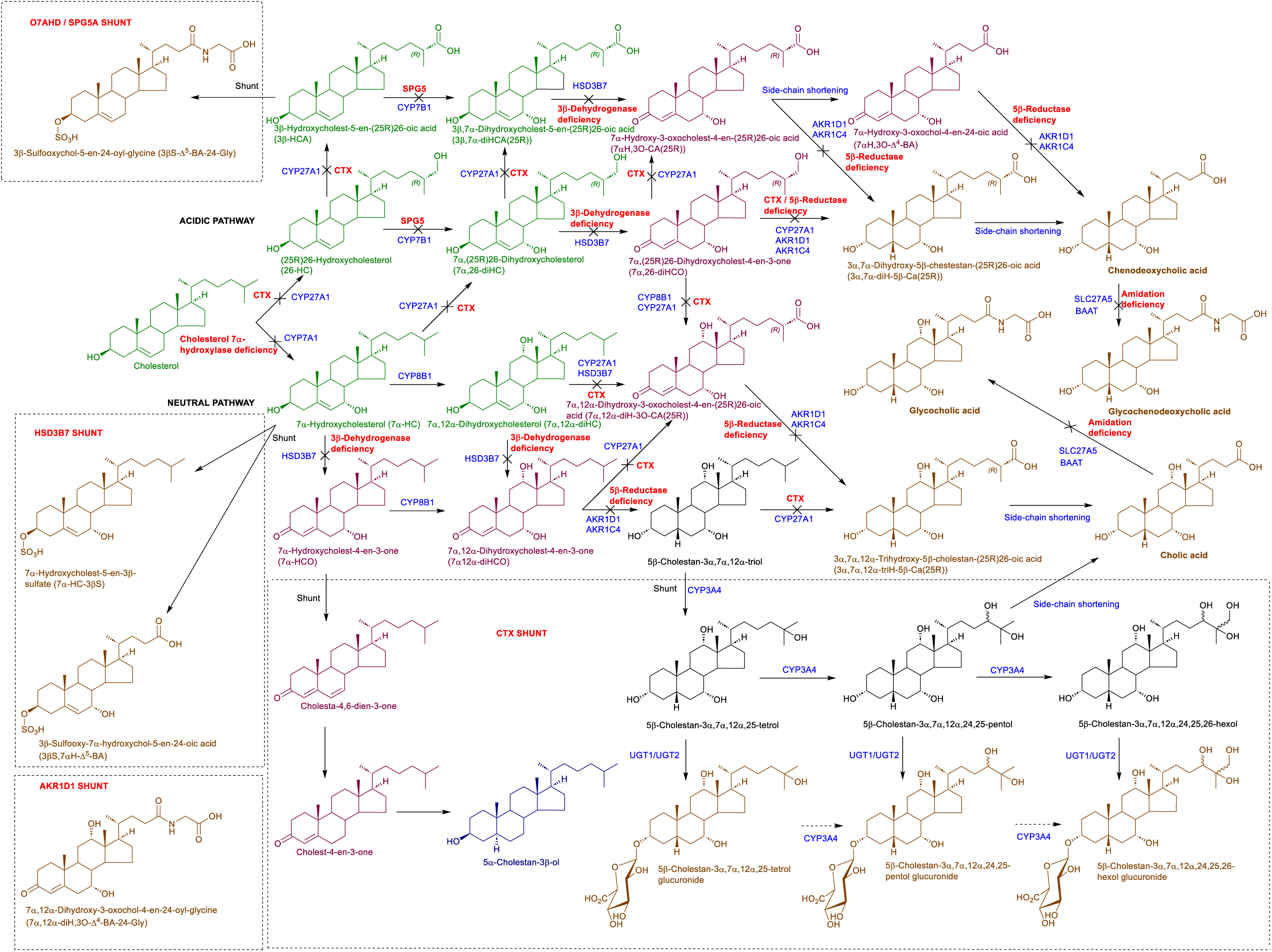
Simplified scheme of the neutral and acidic pathways of bile acid biosynthesis showing in dashed-boxes shunt pathways exploited in CTX, 3β-hydroxy-Δ^5^-C_27_-steroid oxidoreductase (HSD3B7) deficiency, Δ^4^-3-oxosteroid-5β-reductase (AKR1D1) and oxysterol 7α-hydroxylase (CYP7B1) deficiency. Enzymes are shown in blue and disorders in bold red. Where enzymes are suggested rather than known the reactions are indicated by broken arrows. Metabolites observed by LC-MS(MS^n^) after cholesterol oxidase treatment and derivatisation by GP reagent are shown in green, metabolites derivatised with GP reagent in the absence of cholesterol oxidase treatment are shown in claret. 5α-Cholestan-3β-ol, shown in dark blue, has a 3β-hydroxy-5α-hydrogen configuration and is a substrate for cholesterol oxidase and GP derivatisation but is normally of low abundance in blood. Acidic metabolites, shown in brown, are observed in negative-ion MS. Enzyme abbreviations with disorders shown in parenthesis: CYP27A1, cytochrome P450 family 27 subfamily A member 1 (CTX, cerebrotendinous xanthomatosis); CYP7B1, cytochrome P450 family 7 subfamily B member 1 (SPG5A, spastic paraplegia type 5A also oxysterol 7α-hydroxylase deficiency); HSD3B7, hydroxysteroid dehydrogenase 3B7 also called 3β-hydroxy-Δ^5^-C_27_-steroid oxidoreductase, (3β-dehydrogenase deficiency); AKR1D1, aldo-keto reductase family 1 member D1, (Δ^4^-3-oxosteroid-5β-reductase deficiency or 5β-reductase deficiency); SLC27A5, solute carrier family 27 member 5, (bile acyl-CoA ligase deficiency), note, SLC27A2 is the first enzyme in the side-change shortening mechanism acting on C_27_ acids; BAAT, bile acid-CoA:amino acid *N*-acyl transferase, (BAAT deficiency, both bile acid-CoA ligase and BAAT deficiencies result in amidation disorders); CYP7A1, cytochrome P450 family 7 subfamily A member 1, (cholesterol 7α-hydroxylase deficiency). Other enzymes: AKR1C4, aldo-keto reductase family 1 member C4; CYP8B1, cytochrome P450 family 8 subfamily B member 1. Details of side-chain shortening can be found in Supplemental Figure S2.

Considering sterol transport disorders, Niemann Pick type C (NPC) disease is a rare autosomal recessive disorder resulting from variants in either *NPC1,* 95% of case (OMIM: 257220), or *NPC2*, 4% of cases (OMIM: 607625). The clinical incidence is estimated to be about 1 in 100,000 (30, 31), but the incidence may be as high as 1 in 20,000 for late onset forms (32, 33). NPC shows variable age of onset extending from perinatal to adult and can be difficult to diagnose because of an absence of specific clinical signs. NPC can be diagnosed by LC-MS for plasma biomarkers including elevated cholestane-3β,5α,6β-triol (C-triol or 3β,5α,6β-triol) and 7-oxocholesterol (7-OC, also called 7-ketocholesterol, 7-KC) (34), and their down-stream metabolic products. These including the bile acid 3β-hydroxy-7β-*N*-acetylglucosaminyl-chol-5-en-24-oic acid (3βH,7β-GlcNAc-Δ^5^-BA), its glycine (3βH,7β-GlcNAc-Δ^5^-BA-24-Gly) and sulfuric acid (3βS,7β-GlcNAc-Δ^5^-BA) conjugate and multi conjugate (3βS,7β-GlcNAc-Δ^5^-BA-24-Gly) derived from 7-OC or 7β-hydroxycholesterol (7β-HC); and 3β,5α,6β-trihydroxycholan-24-oic acid (3β,5α,6β-triHBA) and its glycine conjugate (3β,5α,6β-triHBA-24-Gly) derived from 3β,5α,6β-triol (see Figure 3) (35–37). These biomarkers are similarly applicable to Niemann Pick type B (NPB, OMIM: 607616), also known as acid sphingomyelinase (*SMPD1*) deficiency (ASMD), and to lysosomal acid lipase (*LIPA*) deficiency (LALD, OMIM: 620151 and 278000) (30, 38). *N*-Palmitoyl-*O*-phosphocholineserine (PPCS), previously known as “lyso-sphingomyelin 509” is an additional plasma biomarker for NPC and NPB, while sphingosine-1-phosphocholine (SPC, SM(d18:1/0:0) or lysosphingomyelin) is particularly elevated in NPB (Figure 3) (39, 40). The ratio of palmitoyl-*O*-phosphocholineserine to sphingosine-1-phosphocholine has been suggested as a marker to differentiate NPC from NPB (31).

**Figure 3.**
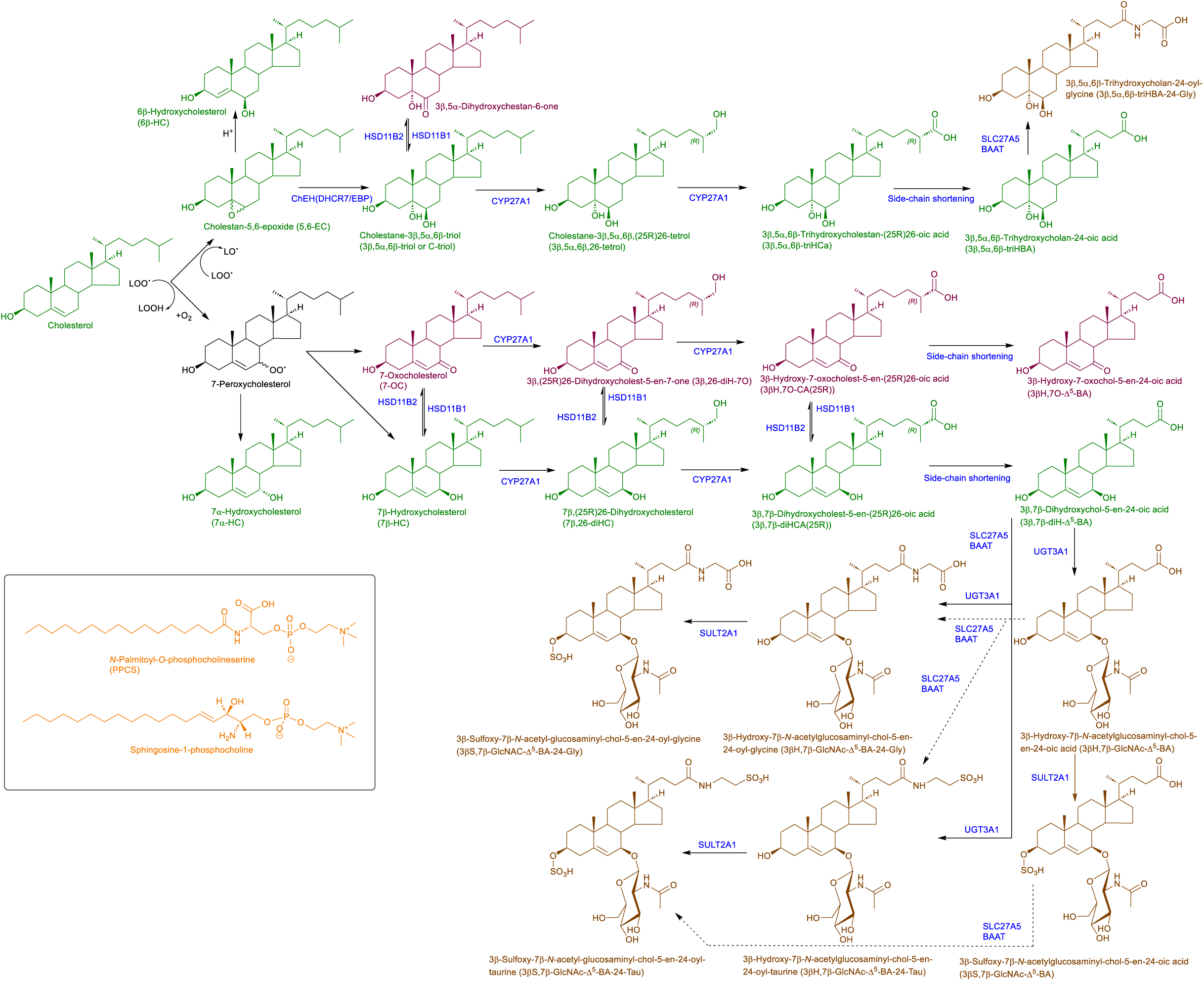
Metabolism of cholesterol to 3β,5α,6β-trihydroxycholan-24-oyl-glycine and 3β-hydroxy-7β-*N*-acetylglucosaminyl-chol-5-en-24-oyl-glycine via cholestane-3β,5α,6β-triol and 7-oxocholesterol / 7β-hydroxycholesterol. All five metabolites are biomarker of NPC, NPB and lysosomal acid lipase deficiency. In SLOS, 7-OC is formed enzymatically from 7-DHC by CYP7A1 (see Figure 1) and can be metabolised to 3β-hydroxy-7β-*N*-acetylglucosaminyl-chol-5-en-24-oic acid as shown in this Figure. Cholesterol metabolites are coloured as in earlier Figures. Phospholipids are in orange. Enzyme abbreviations: ChEH, cholesterol-5,6-oxide hydrolase; UGT3A1, UDP Glycosyltransferase Family 3 Member A1; SULT2A1, sulfotransferase family 2A member 1.

DI-MS or LC-MS have been used in the diagnosis of multiple cholesterol related disorders in a single analysis (1–3, 5, 36, 41) and recently Gelb and co-worker and Heywood and colleagues independently reported multiplexed LC-MS assays applicable to dried blood spots for the diagnosis of several lysosomal storage disorders (42, 43). Gelb’s assay is being expanded to include CTX (44). The multiplex concept is being pursued further by ScreenPlus, a newborn screening program run by Albert Einstein College of Medicine which will screen for 14 disorders including cholesterol-related conditions NPC, NPB, LALD and CTX. However, this and most other multiplexed DI-MS or LC-MS assays are not all embracing and are not ideal for the simultaneous diagnosis of other cholesterol-related inherited disorders of metabolism e.g. SLOS, cholesterol 25-hydroxylase (CH25H) deficiency, spastic paraplegia type 5A (SPG5A, OMIM: 270800) and oxysterol 7α-hydroxylase (CYP7B1) deficiency (O7AHD, OMIM: 613812) (8, 45–48).

Here we show the effectiveness of a plasma/serum based multiplexed LC-MS assay that can be used for the diagnosis of SLOS, lathosterolosis (OMIM: 607330), CTX, SPG5A, O7AHD, CH25H deficiency, 3β-hydroxy-Δ^5^-C_27_-steroid oxidoreductase (HSD3B7) deficiency (OMIM: 607765), branched-chain acyl-CoA oxidase (ACOX2) deficiency (OMIM: 617308) and in principle, based on mouse data, α-methylacyl-CoA racemase (AMACR) deficiency (OMIM: 614307). In addition, the method allows the identification of the lysosomal storage disorders NPC, NPB and LALD, although not their differentiation. The method will also aid the diagnosis of X-linked dominant chondrodysplasia punctata-2 (CDPX2, OMIM: 302960) and male EBP disorder with neurological abnormalities (MEND, OMIM: 300960), both deficiencies in emopamil-binding protein (EBP); desmosterolosis (DHCR24 deficiency, OMIM: 602398); Δ^4^-3-oxosteroid 5β-reductase (AKR1D1) deficiency (OMIM: 235555), and the amidation disorders bile acyl-CoA synthetase (BACS) deficiency and bile acid-CoA: amino acid *N*-acyl transferase (BAAT) deficiency (OMIM: 619232) but not their differentiation. The method also allows the diagnosis of sitosterolaemia-1 (ABCG8 deficiency, OMIM: 210250) and sitosterolemia-2 (ABCG5, OMIM: 618666) but not necessarily their differentiation. We have collated this data into a sterolomic library for the diagnosis and monitoring of cholesterol-related rare diseases (see Supplemental Tables S1 and S2).

## MATERIALS AND METHODS

### Ethical approval

All participants or their parent/guardian provided written informed consent in accordance with the Declaration of Helsinki. The study was conducted with institutional review board approval REC08/H1010/63 (Northwest Research Ethical Committee, UK), REC23/WA/0002 (Wales Research Ethics Committee 3), and vote 199/2011BO1 (regulatory board of the Medical Faculty at the University of Tübingen, Germany). Samples for validation of new methodology were used according to The Royal College of Pathologists “Guidance on the use of clinical samples retained in the pathology laboratory”.

### Enzyme-assisted derivatisation for sterol analysis - cholesterol precursors, oxysterols including bile acid precursors

The method adopted, “enzyme-assisted derivatisation for sterol analysis” (EADSA), is based on analysis of 100 μL of plasma, although smaller volumes can be used at the expense of quantifying fewer low abundance metabolites (41). The method involved single-phase extraction of sterols, oxysterols and bile acid precursors into ethanol containing isotope-labelled standards and, after dilution to 70% ethanol and centrifugation, separating oxysterols and bile acid precursors from highly abundant cholesterol on a reversed-phase C_18_ solid phase extraction (SPE) column (41). Oxysterols and bile acid precursors eluted in the flow through (SPE1-Fr1), and after a column-wash, cholesterol and similarly lipophilic components eluted in absolute ethanol (SPE1-Fr3, see Figure 4, exact details are given in Supplemental Information). Both SPE1-Fr1 and SPE1-Fr3 were divided into two equal subfractions giving SPE1-Fr1A, SPE1-Fr1B and SPE1-Fr3A, SPE1-Fr3B. Each sub-fraction was then lyophilised and re-suspended in 100 μL of propan-2-ol.

**Figure 4.**
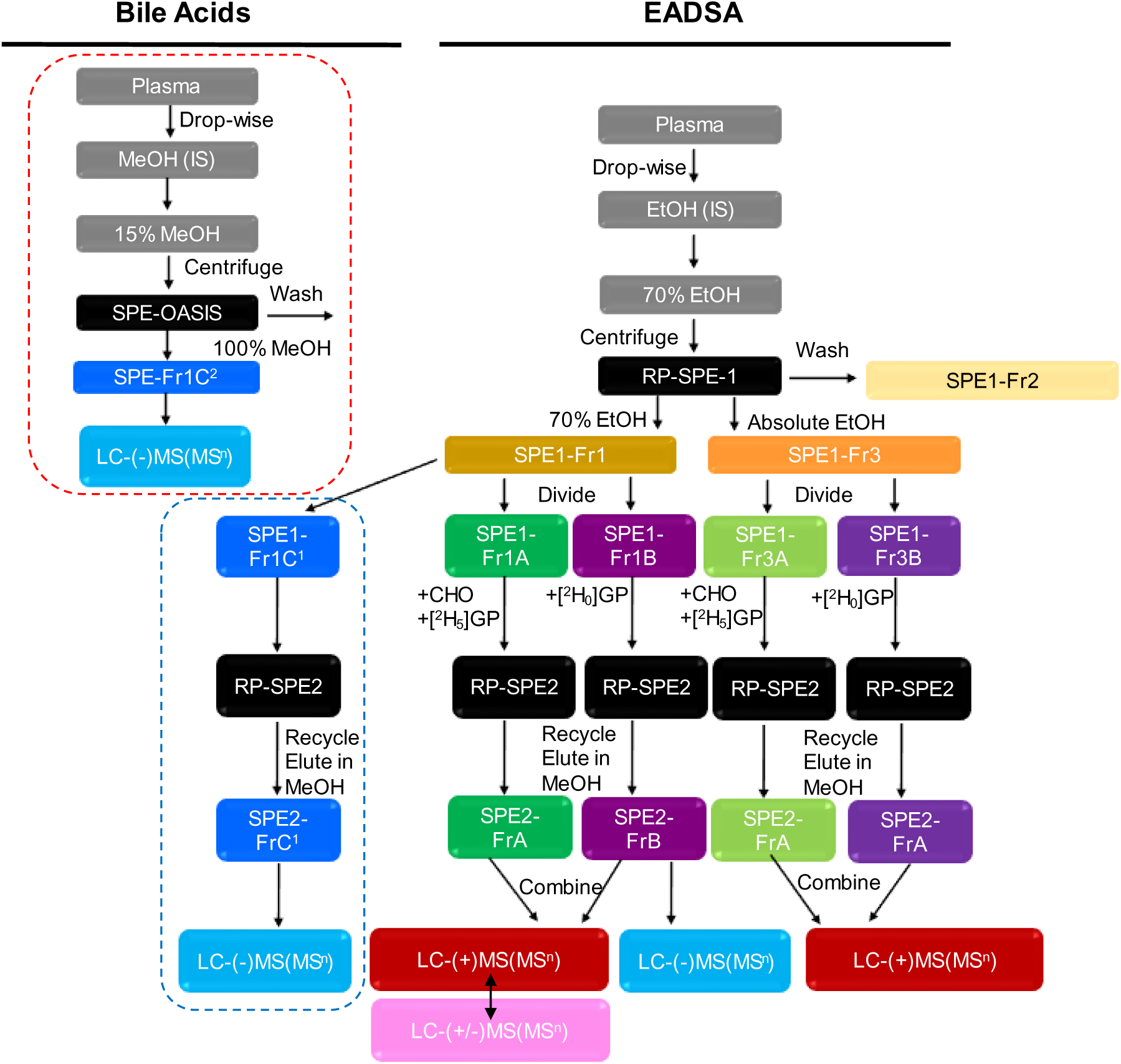
Scheme depicting sample preparation of sterols, oxysterols, bile acid precursors and bile acids for LC-MS(MS^n^) analysis. By combining SPE2-FrA and SPE2-FrB following derivatisation it is possible to simultaneously analyse oxysterols and bile acids precursors with a 3-oxo-4-ene or 3β-hydroxy-5-ene structure by positive-ion LC-MS(MS^n^). By switching polarity to negative-ion, underivatized non-amidated bile acids are detected. However, glycine, taurine and sulfate conjugates are lost to different extents. To observe the entire repertoire of bile acids and bile alcohol conjugated with amino acids, sugars and sulfuric acid, the SPE1-Fr1 eluate is divided into a third fraction SPE1-Fr1C^1^ which is then treated as fraction SPE1-Fr1B but in the absence of both cholesterol oxidase and GP reagent (see box with blue outline). Where bile acids or bile alcohols are of primary interest they are analysed by a simplified protocol shown in the red box involving only one SPE step. SPE columns are shown on a black background, fractions are colour coded, those treated with cholesterol oxidase and [^2^H_5_]GP reagent are on a green background, those treated by [^2^H_0_]GP in the absence of cholesterol oxidase on a claret/purple background and those without either treatment on a blue background. (-) indicates negative-ion MS analysis shown on a light blue background, (+) positive-ion MS analysis shown on a red background and (+/-) positive/negative-ion switching shown on a pink background. Abbreviations: CHO, cholesterol oxidase; GP, Girard P hydrazine.

### A-fractions

To the individual A-fractions cholesterol oxidase enzyme in phosphate buffer was added and the mixture incubated for 1 hr at 37°C. This efficiently converts C_27_ and C_24_ sterols, oxysterols and bile acid precursors with a 3β-hydroxy-5-ene function to their 3-oxo-4-ene equivalents (Supplemental Figure S1A) (49). Note, the enzyme will less efficiently catalyse the oxidation of 3β-hydroxy-5α-hydrogen and 3β-hydroxy-5α-hydroxy substrates to their 3-oxo counterparts. Next methanol and glacial acetic acid were added to give a solution of 5% acid in 69% organic. A large molar excess of [^2^H_5_]Girard P (GP) hydrazine reagent (0.8 mmole) was then added. GP hydrazine reagent readily react with the 3-oxo-4-ene function to give hydrazones (50, 51). As the GP reagent is positively charged this results in charge-tagging of the target analyte and a great improvement in sensitivity for LC-MS analysis of the tagged components (41, 52). Excess derivatisation reagent was then removed by a re-cycling method on a second reversed-phase SPE column yielding SPE2-FrA.

### B-fractions

Some sterols, oxysterols and bile acid precursors naturally possess an oxo group, e.g. 7-OC, 7α-hydroxy-3-oxocholest-4-en-(25R/S)26-oic acid (7αH,3O-CA(25R/S), Figure 2 & 3) (53), so will react with GP reagent in the absence of added cholesterol oxidase (see Supplemental Figure S1B). This leads to an extension in the protocol where the individual B-fractions were treated in an identical fashion to A-fractions as described above but in the absence of cholesterol oxidase and with [^2^H_0_]GP replacing its deuterated analogue (see Supplemental Figure S1B). After recycling on a second reversed-phase SPE column (41), an aliquot of the resulting fraction SPE2-FrB was combined with an equal volume aliquot of SPE2-FrA (Figure 4) and after dilution injected onto the LC-MS system. This procedure was the same for both the oxysterol/bile acid precursor fractions collected from SPE-1 (SPE1-Fr1) and the cholesterol rich fractions SPE1-Fr3.

### LC-MS(MS^n^)

LC separation was performed on a C_18_ reversed-phase column (Hypersil Gold C_18_, 2.1 x 500 mm, 1.9 μm for cholesterol metabolites; Ace C_18_ 10 cm x 2.1 mm, 2 µm for cholesterol and its precursors) using a water, methanol, acetonitrile, 0.1% formic acid gradient followed by electrospray ionisation (ESI)-MS analysis on an Orbitrap Elite, Orbitrap IDX or Orbitrap IQX (see Supplemental Information for further details). Mass spectra were typically recorded at 120,000 or 240,000 resolution (at *m/z* 400) in the positive-ion mode. Mass accuracy was consistently better than 5 ppm. In parallel to recording mass spectra in the Orbitrap, MS^3^ spectra were recorded in the linear ion trap (LIT) exploiting the transitions [M]^+^è[M-Py]^+^è, where [M]^+^ is the GP-derivatised molecule and [M-Py]^+^ its fragment ion where pyridine has been lost (see Supplemental Figure S1C & D). For each injection there were typically four to six MS^3^ scan events where different target *m/z* were fragmented. Alternatively, MS^2^ (tandem MS, MS/MS) spectra ([M]^+^è) were recorded on the IDX or IQX instruments using the quadrupole mass filter for precursor ion selection (0.8 u window), the ion routing multipole for fragmentation and the Orbitrap for *m/z* analysis (typical resolution 7,500 at *m/z* 400). Each sample was injected several times onto the LC-MS system to allow fragmentation spectra to be recorded for multiple target analytes without prescription of retention time. When data was acquired on the IDX or IQX instruments one such injection exploited positive-negative-ion switching, allowing recording of both positive- and negative-ion spectra in the same analytical run. Preferably, as sample quantities were not limiting, a separate LC-MS run was performed in the negative-ion mode (resolution 120,000 at *m/z* 400) on Fraction SPE2-FrB.

Sterols and oxysterols, including bile acid precursors, derivatised with [^2^H_0_]GP were observed as even *m/z* [M]^+^ ions in LC-MS analysis (see Supplemental Figure S1B). These were derived from B-fractions where the target molecules possessed a **natural** oxo group (e.g. 7-OC, 7αH,3O-CA(25R/S)). These metabolites were quantified directly against isotope-labelled internal standards also derivatised with [^2^H_0_]GP. Metabolites originating from A-fractions were derivatised with [^2^H_5_]GP and their [M]^+^ ions had an odd *m/z*. Such molecules could have a **natural** oxo group (e.g. 7αH,3O-CA(25R/S)) or one introduced by *ex vivo* treatment with cholesterol oxidase (e.g. 3β,7α-dihydroxycholest-5-en-(25R/S)26-oic acid, 3β,7α-diHCA(25R/S)), in which case the sum of the originating molecules was quantified against the isotope-labelled standard derivatised with [^2^H_5_]GP. In situations where [M]^+^ ions were derived from molecules containing a **natural** oxo group (e.g. 7αH,3O-CA(25R/S)) and those oxidised *ex vivo* to contain one (e.g. 3β,7α-diHCA(25R/S)), quantities determined from the B-fraction ([^2^H_0_]GP, even mass) were indicative of *only* the **natural** oxo containing molecules (7αH,3O-CA(25R/S)) and the quantitative difference between fraction-A (7αH,3O-CA(25R/S) + 3β,7α-diHCA(25R/S)) and fraction-B (7αH,3O-CA) gave the amounts of the molecules oxidised *ex vivo* to contain an oxo group (i.e. 3β,7α-diHCA(25R/S), see Eq 1).

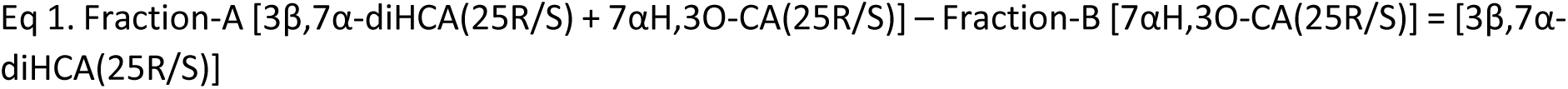

Using this protocol essentially all oxysterols and bile acid precursors present in plasma with a 3β-hydroxy-5-ene (substrate for cholesterol oxidase) or with a 3-oxo-4-ene function are detected (41, 54–56). Sterols with a 3β-hydroxy-5α-hydrogen or 3β-hydroxy-5α-hydroxy group are also substrates for cholesterol oxidase (38, 49, 56, 57), as are sterols with a 3β-hydroxy-5,7-diene, 3β-hydroxy-5,8-diene or 3β-hydroxy-4,6-diene structure (58). Besides reacting with 3-oxo groups, GP reagent will also react with oxo groups at carbon 6 and 7, and in C_18_, C_19_ and C_21_ steroids at carbons 17, 20 and 21 (51, 59). A significant advantage of Girard derivatisation is that upon LC-MS with multistage fragmentation (MS^n^, either MS^2^ or MS^3^) the resulting spectra are information rich providing security for compound identification (41, 52, 59, 60).

However, some bile acid precursors and primary bile acids themselves, possess a 3α-hydroxy rather than a 3β-hydroxy group and unless they have an oxo group at another location are not derivatised by the above protocol and are transparent to the EADSA method. Fortuitously, bile acids possess an acidic function (as do glucuronidated or sulfated bile alcohols) and are amenable to analysis by LC-MS in the negative-ion mode (61). This allows their analysis by exploiting positive-negative-ion switching or preferably adding extra injections and recording negative-ion scans to complement the positive-ion scans used for detection of the GP-derivatised analytes.

### Non-amidated bile acids

Non-amidated bile acids were analysed in the same LC-MS(MS^n^) run as oxysterols and bile acid precursors by performing positive-negative-ion switching, where bile acids were recorded in the negative-ion mode. Preferably, a separate aliquot of Fraction SPE2-FrB was injected on the column (Figure 4). SPE2-FrB is devoid of bile acids possessing an oxo group, now derivatised with GP reagent, but bile acids with a 3α- or 3β-hydroxy group are still present. There is, however, a disadvantage in that amidated and sulfated bile acids can be lost. Negative-ion scans were recorded on the IDX or IQX instrument at a resolution of 120,000 (at *m/z* 400) with mass accuracy typically better than 5 ppm. MS^2^ (MS/MS) spectra ([M-H]^-^è) were recorded using the quadrupole mass filter for precursor ion selection, the ion routing multipole for fragmentation and the Orbitrap for *m/z* analysis at a resolution of 7,500 at *m/z* 400 giving a mass accuracy of better than 20 ppm in most cases.

### Complete bile acid profile

When a detailed profile of bile acids was required, the eluent from SPE1-Fr1 was divided into three subfractions SPE1-Fr1A, SPE1-Fr1B and SPE1-Fr1C^1^, where SPE1-Fr1C^1^ was used for bile acid analysis and was processed in an identical fashion to SPE1-Fr1B but in the absence of added GP reagent (Figure 4, blue box). Where quantitative bile acid information was desired appropriate isotope-labelled internal standards (Cayman Chemical Company) were included during the single-phase liquid extraction step (see Supplemental Information for more details). When only bile acid information was required a simpler sample preparation protocol involving single-phase extraction into methanol, dilution to 15% methanol, centrifugation and a single reversed-phase SPE step was used (SPE1-Fr1C^2^), followed by LC-MS analysis in the negative-ion mode.

## RESULTS AND DISCUSSION

### Disorders of cholesterol synthesis

#### Smith-Lemli-Opitz Syndrome

In earlier studies exploiting EADSA and LC-MS(MS^n^), we showed, as expected, the plasma ratio of unresolved 7-DHC and 8-DHC to cholesterol to be elevated compared to controls (62). In the current study we have repeated the earlier work but with improved chromatographic resolution allowing the separation of 7-DHC and 8-DHC and confirmed the previous result (Figure 5, see also Supplemental Table S1, column M).

**Figure 5.**
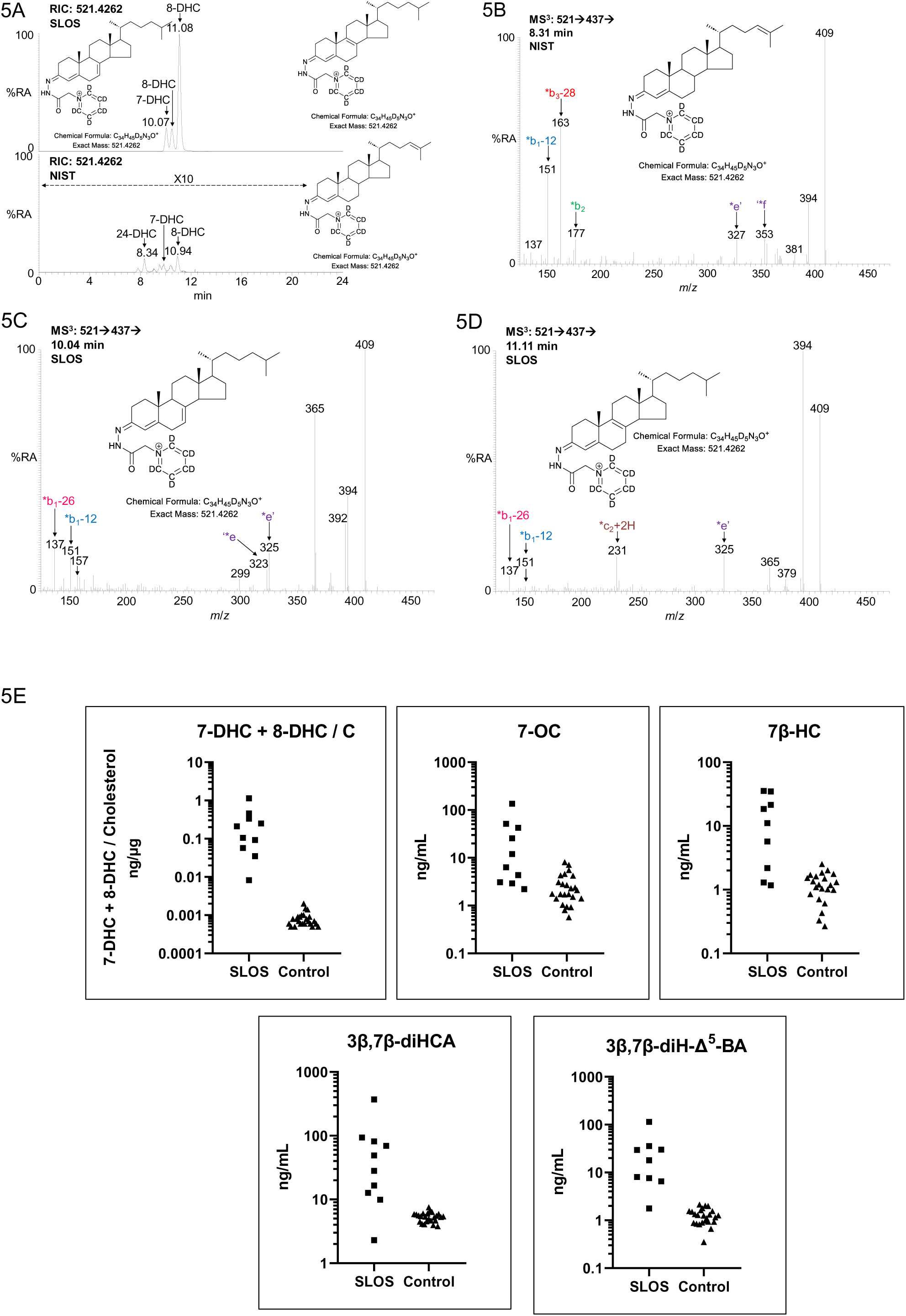
EADSA-LC-MS(MS^n^) analysis of plasma from a SLOS patient and of the NIST SRM 1950 standard reference material (65). (A) LC-MS reconstructed ion-chromatogram (RIC, ± 5ppm) for dehydrocholesterol isomers in an SLOS sample (upper panel) and in NIST SRM 1950 plasma (lower panel). For the NIST sample the y-axis has been magnified by a factor of 10. MS^3^ ([M]^+^è[M-Py]^+^è) spectra of (B) desmosterol (24-DHC); (C) 7-DHC; and (D) 8-DHC. Fragmentation nomenclature is described in (41). The spectra of 7-DHC and 8-DHC are from the SLOS sample, the spectrum of desmosterol is from the NIST sample. Data acquired on the Orbitrap IQX using an Ace C_18_ column. Note, GP derivatives give *cis-* and *trans*-configurations about the C=N bond, in some cases these are separated on the C_18_ column. (E) Concentrations of different metabolites in SLOS patients (n = 10) and adult controls (n = 24), 7-DHC+8-DHC/cholesterol; 7-OC; 7β-HC; 3β,7β-diHCA(25R/S); and 3β,7β-diH-Δ^5^-BA. The y-axis is shown on a log scale. Data in (E) previously reported in (58).

Metabolites of 7-DHC containing a 7-oxo or 7β-hydroxy group are often observed at elevated concentrations in plasma from SLOS patients e.g. 7β-HC, 3β,7β-dihydroxycholest-5-en-(25R/S)26-oic acid (3β,7β-diHCA(25R/S)) and 3β,7β-dihydroxychol-5-en-24-oic acid (3β,7β-diH-Δ^5^-BA), see shunt pathway in Figure 1 and data Supplemental Table S1 (58). However, these latter metabolites have to date been measured in very few patients (58) and are also elevated in other disorders where concentrations of 7-OC or 7β-HC are high e.g. NPC, ASMD, LALD (38, 57) indicating that the 7-DHC plus 8-DHC to cholesterol ratio is the optimal diagnostic marker.

#### Lathosterolosis

In this study we analysed plasma from three patient suffering from lathosterolosis (OMIM: 607330), a disorder which results from a deficiency of the enzyme sterol C-5 desaturase (SC5D, Figure 1) and elevation in plasma lathosterol (63). Although lathosterol was shown to be elevated by GC-MS (e.g. Anderson et al (64)), we were unable to resolve lathosterol on our LC-MS system from its much more abundant isomer, cholesterol (Supplemental Figure S3). However, in more severe cases of lathosterolosis there is a possibility of observing this change using EADSA and LC-MS(MS^n^).

While elevated lathosterol is the classical diagnostic for lathosterolosis, the enzyme SC5D is also required for the formation of 8-DHC and of desmosterol (Figure 1), two abundant cholesterol precursors normally observed in plasma by the EADSA-LC-MS(MS^n^) assay. In the three lathosterolosis patients studied here the concentrations of 8-DHC and of desmosterol were reduced by factors between 3 and 20 and 5 to 20, respectively, compared to the NIST SRM 1950, a standard reference material reflective of the adult US population (see Supplemental Figure S3D and Supplemental Table S1) (65), and in the case of desmosterol and in two of the three patients for 8-DHC well below the 2.5^th^ percentile for a control group of more than 80 adults routinely used to provide a reference range in our laboratory. Anderson et al also observed desmosterol to be at low concentration in plasma of one of these patients (64). Low concentrations of 8-DHC and desmosterol can be used to support a biochemical diagnosis of lathosterolosis made by GC-MS. Interestingly, in our study and that of Anderson et al, and in an earlier study made on different patients by Ho et al, 7-DHC concentrations were found in the normal range or even elevated (64, 66). This indicates that despite a deficiency in SC5D the Kandutsch-Russel pathway of cholesterol biosynthesis is intact.

#### Other disorders of cholesterol synthesis

Very few cases of desmosterolosis (OMIM: 602398) have been reported, only nine cases to date (6, 67, 68). Here there is a deficiency in the enzyme DHCR24 (see Figure 1), leading to elevated tissue and plasma desmosterol, which may be 100 times the normal concentration (see Supplemental Table S1, column M). Although we do not report a case of desmosterolosis here, a 100-fold increase in plasma desmosterol concentration is evident upon EADSA-LC-MS(MS^n^) analysis.

Deficiency in EBP, also called Δ^8^-Δ^7^ sterol isomerase (D8D7I), leads to the disorder CDPX2 (OMIM: 302960) in females and to MEND (OMIM: 300960) in males (4, 6). EBP catalyses the conversion of zymostenol to lathosterol, zymosterol to 5α-cholesta-7,24-dien-3β-ol and 8-DHC to 7-DHC, all involving Δ^8^-Δ^7^ isomerisation (see Figure 1). Biochemically this leads to an increase in plasma zymostenol and 8-DHC which have been measured in the past by GC-MS to diagnose the disorder, where 8-DHC was found to be elevated by more than 500-fold (see Supplemental Table S1, column M) (69–71). Although zymostenol, an isomer of cholesterol, is difficult to resolve from cholesterol by EADSA-LC-MS, 8-DHC is readily resolved from both 7-DHC and desmosterol (see Supplemental Figure S3D), so CDPX2 and MEND will be diagnosed by EADSA-LC-MS(MS^n^).

### Disorders of bile acid biosynthesis

#### Cerebrotendinous Xanthomatosis

CTX (OMIM: 213700) results from a deficiency of the first enzyme, CYP27A1, in the acidic pathway of bile acid synthesis (Figure 2). However, cholic acid is still synthesised via an additional pathway not involving CYP27A1 catalysed (25R)26-hydroxylation of the steroid side-chain but rather through hydroxylation at C-25 (Figure 2, see CTX Shunt Pathway) (72). Data from analysis of plasma from the *Cyp27a1^-/-^* mouse (73) and from human CTX plasma and CSF (28, 74) indicates that there is a further pathway to primary bile acids proceeding via (25S)26-hydroxylation of 7α-hydroxycholesterol (7α-HC) to 7α,(25S)26-dihydroxycholesterol (7α,(25S)26-diHC) and on to 7α-hydroxy-3-oxocholest-4-en-(25S)26-oic (7αH,3O-CA(25S)) and 7α,12α-dihydroxy-3-oxocholest-4-en-(25S)26-oic (7α,12α-diH,3O-CA(25S)) acids (Supplemental Figure S4), where the 25S-acids are intermediates in side-chain shortening mechanisms of the of bile acid biosynthesis pathways (see also Supplemental Figure S2). CTX can be diagnosed by GC-MS from high concentrations of plasma cholestanol (5α-cholestan-3β-ol) and 7α-HCO, or low concentrations of 26-HC (15, 22, 28), and by LC-MS from elevated concentrations of 7α,12α-diHCO (24–26). It can also be diagnosed from elevated concentrations of bile alcohol glucuronides by LC-MS or DI-MS (1, 3, 5, 20, 44, 75). However, CTX cases with atypical biochemistry may be difficult to diagnose from measurements of 5α-cholestan-3β-ol, 7α-HCO or bile alcohol glucuronides alone, in which case measurements of multiple metabolites simultaneously is desirable (16).

#### Oxysterols in CTX

As can be seen from Figure 2, a deficiency of CYP27A1 will reduce the abundance of not only 26-HC but also 3β-hydroxycholest-5-en-(25R)26-oic acid (3β-HCA) and 7αH,3O-CA(25R/S) (Figure 6). The acids normally have concentrations of about 75 – 200 ng/mL in plasma, while 26-HC has a concentration of 25 – 75 ng/mL, but in CTX their concentrations are usually less than 1 ng/mL. In Figure 7 and Supplemental Table S1 we summarise EADSA-LC-MS(MS)^n^ data from previous studies for these CYP27A1 products along with new data for other metabolites not consistently reported (25, 28, 41, 46, 56, 74, 76). For simplicity of measurement the combination of 7αH,3O-CA(25R) and 7αH,3O-CA(25S) is reported. Also displayed in Figure 7 is EADSA-LC-MS(MS^n^) data for 7α-HC, 7α-HCO and 7α,12α-diHCO which are usually elevated in CTX, although both 7α-HC and 7α-HCO can be normalised upon treatment (25, 28). The elevation in plasma 7α,12α-diHCO in CTX is extreme, rising from less than 1 ng/mL in controls to more than 100 of ng/mL in many cases of CTX (28). The advantage of analysing multiple metabolites by EADSA-LC-MS(MS)^n^ can be exploited through concentration ratios of 7α,12α-diHCO to 26-HC, 7α,12α-diHCO to 3β-HCA and 7α,12α-diHCO to 7αH,3O-CA illustrated in Figure 7B, where the numerator metabolite is elevated in CTX while the denominator is reduced giving clear discrimination between CTX samples and controls. These ratios should be of value in defining CTX where a single marker gives an equivocal result and where genetic testing is not definitive e.g. variants of uncertain significance.

**Figure 6.**
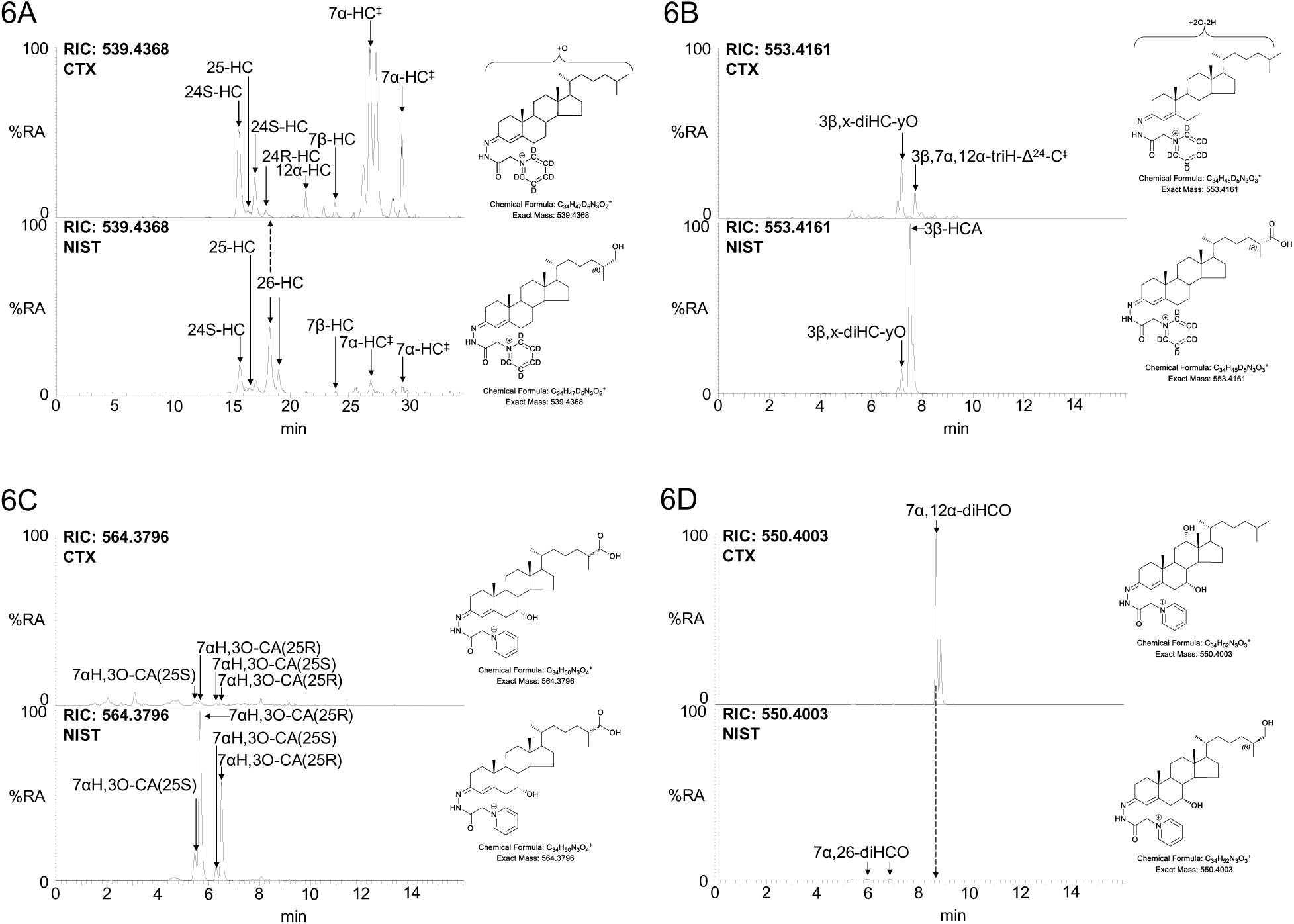
Analysis of CTX (upper panels) and NIST SRM1950 (lower panels) plasma by EADSA-LC-MS(MS^n^). The upper and lower panels are plotted on the same y-axis. (A) LC-MS RIC of *m/z* 539.4368 ± 5 ppm corresponding to mono-hydroxycholesterols and hydroxycholestenones. 7α-HC^‡^ indicates a combination of 7α-HC and 7α-HCO. (B) LC-MS RIC of *m/z* 553.4161 ± 5 ppm corresponding to 3β-HCA and its isomers. 3β,7α,12α-triH-Δ^24^-C^‡^ indicates a combination of 3β,7α,12α-triH-Δ^24^-C and 7α,12α-diH-Δ^24^-CO. (C) LC-MS RIC of *m/z* 564.3796 ± 5 ppm corresponding to 7αH,3O-CA(25R/S). See Figure S5A for the acids shown on a magnified scale from the CTX plasma. (D) LC-MS RIC of *m/z* 550.4003 ± 5 ppm corresponding to dihydroxycholestenones. The CTX patient had been treated with chenodeoxycholic acid (CDCA). Data acquired on the Orbitrap Elite using a Hypersil Gold C_18_ column. 3β,7α,12α-triH-Δ^24^-C and 7α,12α-diH-Δ^24^-CO correspond to cholesta-5,24-diene-3β,7α,12α-triol and 7α,12α-dihydroxycholesta-4,24-dien-3-one, respectively, other compound abbreviations can be found in Supplemental Table S1 or in reference (41). Data for NIST SRM1950 plasma has been previously reported in reference (41).

**Figure 7.**
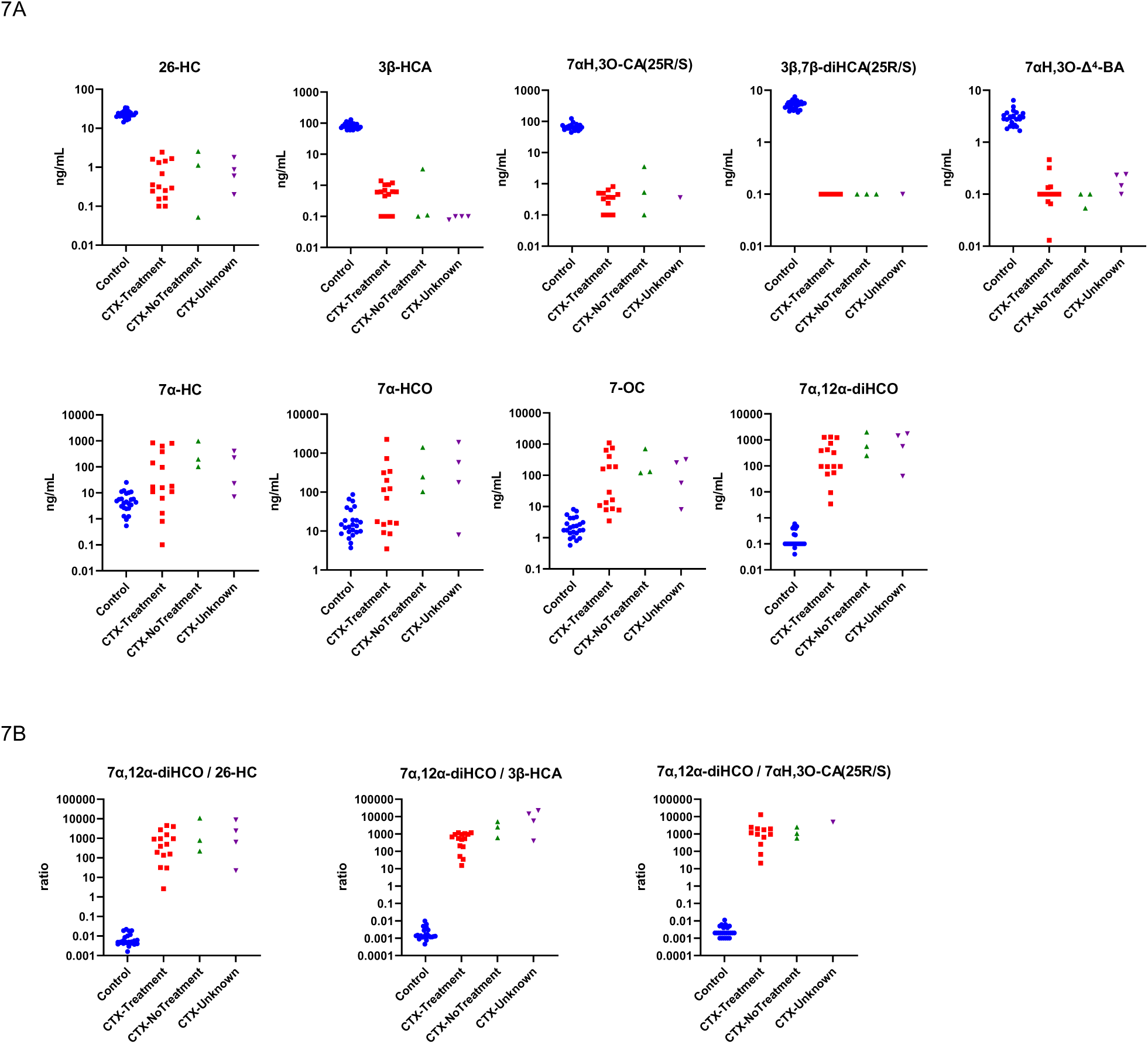
(A) Concentration of bile acid precursors in control (n = 24) and CTX plasma (total n = 22, patients on treatment n = 15, not on treatment n = 3, treatment not known n = 4). For the quantification of 7αH,3O-CA(25R/S) in the CTX group the total number of samples was n = 16. For simplicity of measurement the combined concentration of 7αH,3O-CA(25R) and 7αH,3O-CA(25S) is reported. (B) Concentration ratios of 7α,12α-diHCO/26-HC, 7α,12α-diHCO/3β-HCA, 7α,12α-diHCO/7αH,3O-CA(25R/S). To avoid concentration ratios of infinity, where undetected, concentrations of the denominator are set at 0.1 ng/mL. Data for some metabolites previously reported in (28, 74). Control data from (54). Figure 7B modified from (123).

The metabolites 7α,12α-diHCO and 7αH,3O-CA(25R/S) can be measured using GP-derivatisation without the use of cholesterol oxidase, this means that if diagnosis of CTX is the primary goal of the study, the method can be simplified to only include preparation of fraction-B (omit incubation with cholesterol oxidase, see Figure 4 and Supplemental Table S2, row 26 – 29, column G). The method is also applicable for dried blood spots as illustrated in Supplemental Figure S6.

#### Bile alcohol glucuronides in CTX

Elevated concentrations of bile alcohol glucuronides i.e. glucuronides of cholestanetetrol, cholestanepentol and cholestanehexol in urine and plasma have been wildly used to diagnose CTX (1, 3, 5, 20) and the potential of cholestanetetrol glucuronides, particularly 5β-cholestane-3α,7α,12α,25-tetrol-3-*O*-β-D-glucuronide (7α,12α,25-trihydroxy-5β-cholestane-3α-*O*-β-D-glucuronide) (77, 78), have been assessed as a CTX marker for newborn screening (44, 75, 78). These glucuronides can be analysed in the same LC-MS run as the GP-derivatised metabolites by incorporating positive-negative-ion switching (GP-derivatives in positive-ion mode, bile alcohol glucuronides in negative-ion mode), or preferably if time allows by just analysing fraction-B (or fraction-C, a dedicated bile acid/alcohol preparation, see Figure 4) in the negative-ion mode in a separate injection (Figure 8). This analysis also shows the almost complete absence of chenodeoxycholic acid (CDCA). If bile acids/alcohols and their conjugates are of primary interest, then as in shown in Figure 4, GP-derivatisation can be omitted and the glucuronides analysed after SPE (see Supplemental Figure S7). We do not have access to authentic standards of bile alcohol glucuronides but measured *m/z*, MS^2^ (MS/MS) spectra recorded at high-resolution, and chromatographic retention time and pattern (Figures 8 & S7, cf. reference (78)) are compatible with cholestanetetrol glucuronides. In the absence of bile alcohol glucuronide standards we have not attempted absolute quantification, but as can be seen in Figure 8 the concentration of cholestanetetrol glucuronides in the two patient samples shown is at least 100 fold greater than in the NIST SRM 1950.

**Figure 8.**
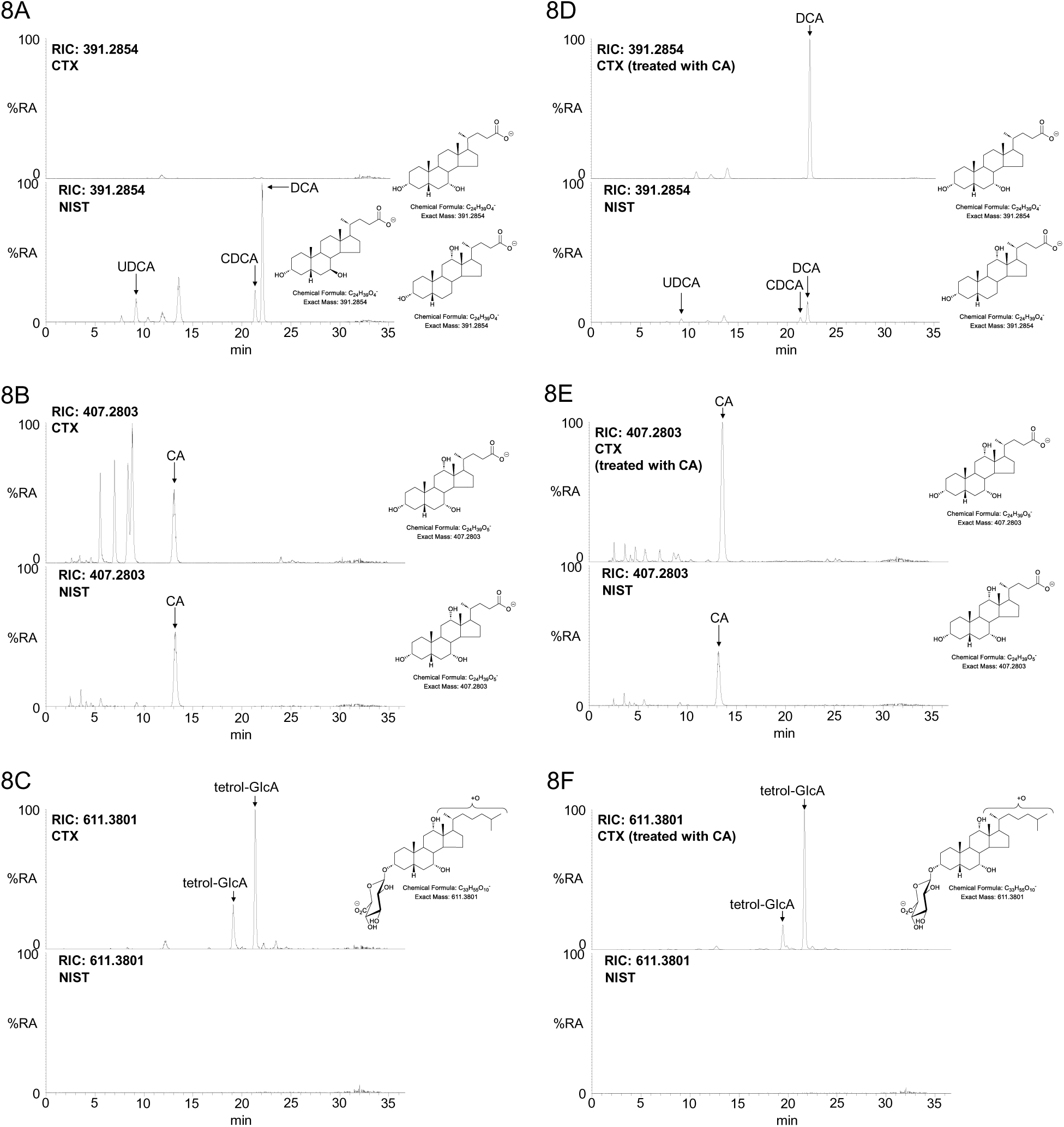
Analysis of CTX (upper panels) and NIST SRM1950 (lower panels) plasma by negative-ion LC-MS of fraction-B (no cholesterol oxidase treatment). The upper and lower panels are plotted on the same y-axis. LC-MS RICs of (A) *m/z* 391.2854 ± 5 ppm, corresponding to dihydroxycholanoic acids; (B) *m/z* 407.2803 ± 5 ppm, corresponding to trihydroxycholanoic acids; and (C) 611.3801 ± 5 ppm corresponding to cholestanetetrol glucuronides. The CTX patient was untreated. (D) LC-MS RICs of *m/z* 391.2854 ± 5 ppm; (E) *m/z* 407.2803 ± 5 ppm; and (F) 611.3801 ± 5 ppm, from a different CTX patient treated with cholic acid (CA). Glucuronidation is drawn at C-3. Data acquired on the Orbitrap IQX using a Hypersil Gold C_18_ column. MS/MS spectra are shown in Supplemental Figure S7. Note, cholic acid is produced in CTX patients despite a deficiency in CYP27A1 (see Figure 2).

Bile alcohol glucuronides only give minor signals from control plasma but can be elevated in e.g. Zellweger spectrum disorders and in cholestasis, in which case the ratio of taurotrihydroxycholestanoic acid (elevated in Zellweger and cholestasis, but not CTX) to cholestanetetrol glucuronide can be used to distinguish the disorders (44, 75). In Zellweger syndrome and in cholestasis both 26-HC and 3β-HCA are produced, so confusion of these disorders with CTX is not a problem using EADSA-LC-MS.

#### Cholesterol 7α-hydroxylase (CYP7A1) deficiency

There is only one report of *CYP7A1* deficiency (79). A kindred with three homozygous subjects with a frame-shift mutation abolishing CYP7A1 activity was found, and CYP7A1-deficiency was linked to hypercholesterolemia in these patients. Biochemical measurements of bile acids were made on plasma, but the results were not consistent although in one patient where stool bile acids were measured, there was a large reduction in bile acid excretion. Liver biopsy in this patient revealed elevated CYP27A1 activity. To confirm the link between CYP7A1 deficiency and hypercholesterolemia there is a need to uncover further patients. EADSA analysis of plasma from such patients should confirm a lack of CYP7A1 activity from an absence of 7α-HCO and low concentrations of 7α-HC, while hypercholesterolemia will be revealed by elevated cholesterol. It is likely that some 7α-HC will still be present in plasma formed via non-enzymatic mechanisms.

#### Oxysterol 7α-hydroxylase deficiency and spastic paraplegia type 5A

A deficiency in the enzyme CYP7B1 can result in two different disorders, Oxysterol 7α-hydroxylase deficiency (O7AHD, OMIM: 613812) or SPG5A (OMIM: 270800). These inherited metabolic disorder were initially found as O7AHD by Setchell et al and later by Ueki et al in infants presenting with jaundice and hepatosplenomegaly (45, 80). They were found to have high urinary and serum bile acids with a 3β-hydroxychol-5-en-24-oic acid (3βH-Δ^5^-BA) structure conjugated with glycine and sulfuric acid and an absence of normal bile acids (45, 80). These infants also showed very high concentrations of serum 26-HC and 3β-HCA (45, 46). A further infant suffering from the same deficiency was found by Dai et al whose condition rapidly improved upon treatment with CDCA (81). This patient also presented with high urinary concentrations of taurine conjugated 3βH-Δ^5^-BA and also of the glycine and sulfuric acid double conjugate and high plasma concentrations of 26-HC and 3β-HCA (81). Supplemental Figure S8 shows the shunt pathway for O7AHD and SPG5A responsible for the elevation of these metabolites.

The disorder SPG5A, like O7AHD, results from deficiency in CYP7B1 but patients present with spastic paraplegia, usually as adolescents (82). SPG5A can be diagnosed by the high concentrations of plasma 25-hydroxycholesterol (25-HC) and 26-HC but unlike in infants presenting with O7AHD, adults with SPG5A present with normal bile acids (48, 83). The absence of normal bile acids in the infant but their presence in the adult indicates that CYP7B1 is most important for bile acid formation via the acidic pathway in early life.

#### Oxysterols in O7AHD and SPG5A

Both O7AHD and SPG5A are readily diagnosed by EADSA-LC-MS(MS^n^) where besides 25-HC and 26-HC, the acids 3β-HCA and 3βH-Δ^5^-BA are found to be elevated (Figure 9, see also Supplemental Table S1). In contrast, 3β,7α-diHCA(25R/S) is diminished although not absent on account of continued formation by the neutral pathway of bile acid synthesis. In Figure 10 we summarise data from three new patients in combination with data from previous studies with re-mining of data for metabolites not previously reported (46, 74). Exploiting concentration ratios, particularly of 25-HC or 3β-HCA to 3β,7α-diHCA(25R/S), the diagnostic value of the methods is accentuated. Notably, for the 3β-HCA to 3β,7α-diHCA(25R/S) ratio, the lowest ratio for SPG5 was more than three-fold higher than the highest ratio for controls, while for 25-HC to 3β,7α-diHCA(25R/S) there was a more than four-fold difference. The difference was even more stark for the infants with the more severe O7AHD.

**Figure 9.**
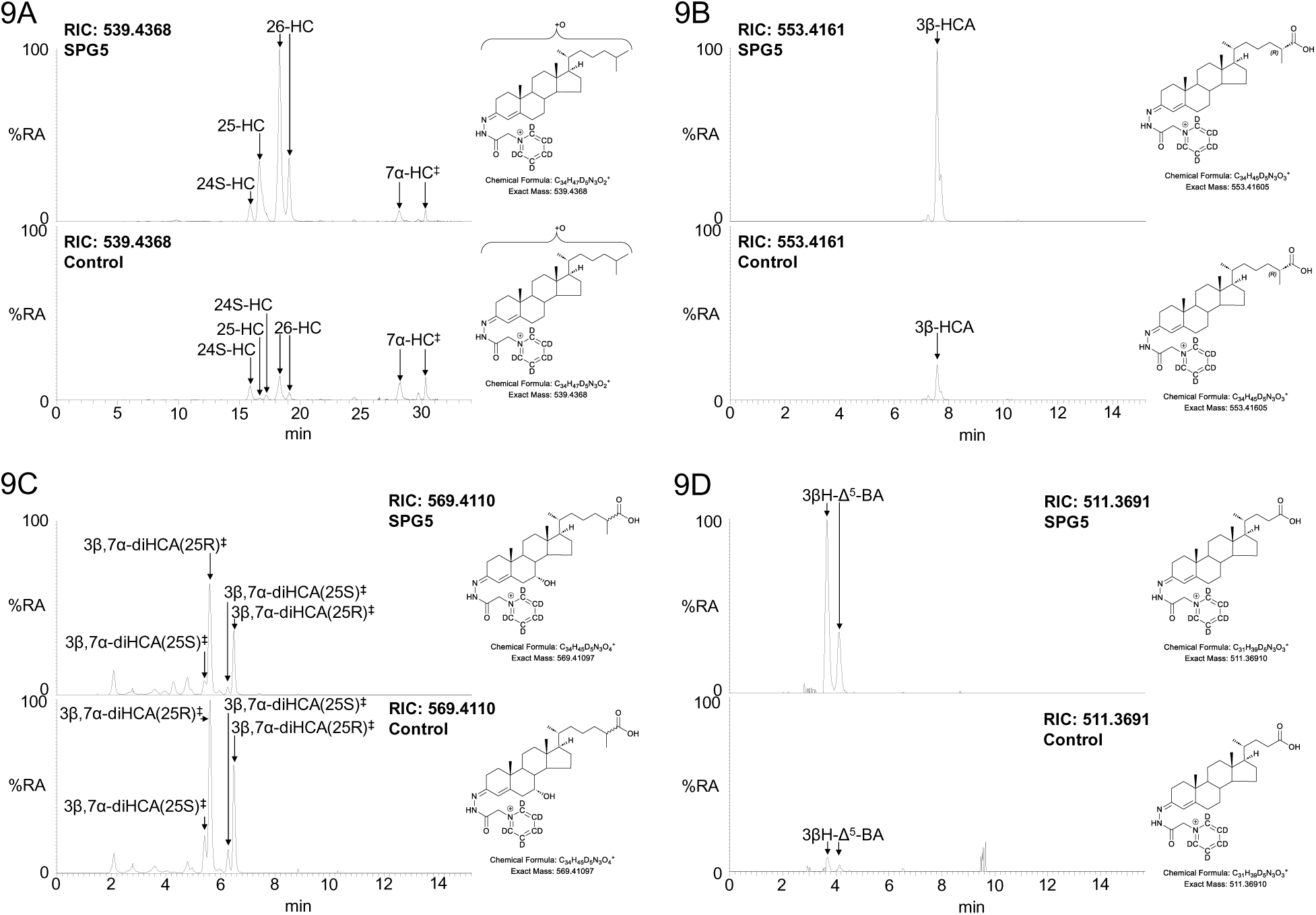
Analysis of SPG5A (upper panels) and control (lower panels) plasma by EADSA-LC-MS(MS^n^). The upper and lower panels are plotted on the same y-axis. (A) LC-MS RIC of *m/z* 539.4368 ± 5 ppm corresponding to mono-hydroxycholesterols plus hydroxycholestenones. 7αHC^‡^ indicates the sum of 7α-HC and 7α-HCO. (B) LC-MS RIC of *m/z* 553.4161 ± 5 ppm corresponding to 3β-HCA. (C) LC-MS RIC of *m/z* 569.4110 ± 5 ppm, 3β,7α-diHCA(25R/S)^‡^ indicates the sum of 3β,7α-diHCA(25R/S) and 7αH,3O-CA(25R/S). (D) LC-MS RIC of *m/z* 511.3691 ± 5 ppm corresponding to 3βH-Δ^5^-BA. Data acquired on the Orbitrap IQX using a Hypersil Gold C_18_ column. Control data is from (54).

**Figure 10.**
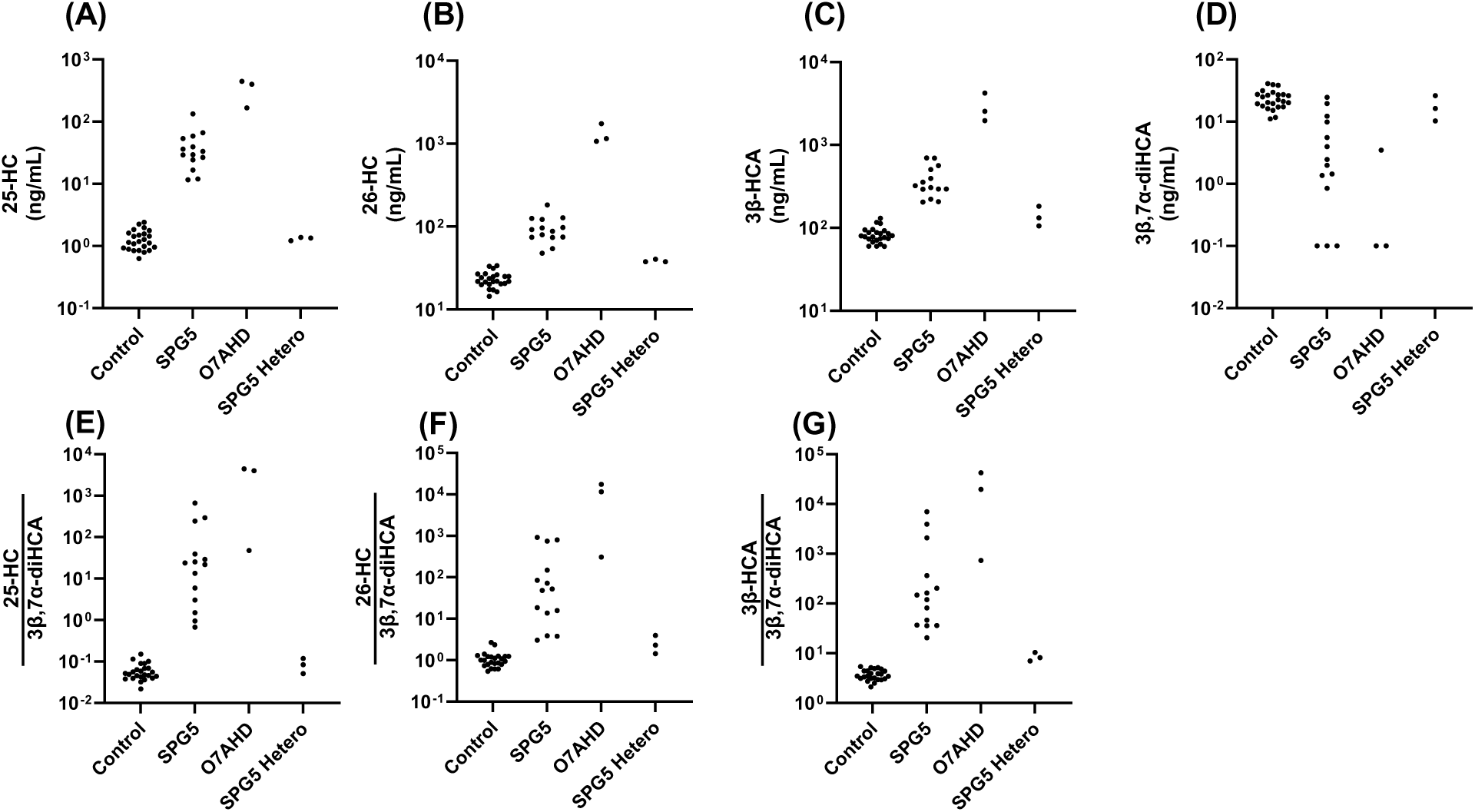
Oxysterols and bile acid precursors in adult controls (n = 24), patients with SPG5A (n = 14), O7AHD (n = 3) and heterozygote individuals (n = 3) measured by EADSA-LC-MS(MS^n^). For simplicity the 25R- and 25S-epimers of 3β,7α-diHCA were measured in combination. (A) 25-HC; (B) 26-HC; (C) 3β-HCA; (D) 3β,7α-diHCA(25R/S); (E) 25-HC/3β,7α-diHCA(25R/S); (F) 26-HC/3β,7α-diHCA(25R/S); and (G) 3β-HCA/3β,7α-diHCA(25R/S). Data from the current study and historical data for O7AHD and SPG5A from (46, 74), control data from (54).

Unfortunately, the sulfates of 3βH-Δ^5^-BA are lost in the derivatisation process, but these were observed by preparing a sample without treatment with cholesterol oxidase or GP reagent (Fraction-C in Figure 4). In this study we did not include a bile acid sulfate internal standard but the elevation of unsaturated and sulfated bile acids is evident by comparison of an SPG5 sample to NIST SRM 1950 plasma as shown in Supplemental Figure S9.

#### 3β-Hydroxy-Δ5-C27-steroid oxidoreductase (HSD3B7) deficiency

The disorder 3β-hydroxy-Δ^5^-C_27_-steroid oxidoreductase deficiency also called 3β-dehydrogenase deficiency (OMIM: 607765) was first defined by Clayton and colleagues (84, 85). It is the most common bile acid synthesis disorder. The affected gene was later identified by Russell and colleagues to be *HSD3B7* (see Figure 2) (86, 87). HSD3B7 deficiency frequently presents with neonatal cholestasis but it has also been observed in the absence of liver disease (3, 88). HSD3B7 deficiency can be effectively treated by bile acid replacement therapy (2, 3, 85). A deficiency in HSD3B7 leads to an elevation in cholesterol metabolites with a 3β,7α-dihydroxy-5-ene structure and a relative decrease in those with a 3-oxo-4-ene structure.

#### Oxysterols and bile acid precursors in HSD3B7 deficiency

The fall in the relative abundance of metabolites with a 3-oxo-4-ene structure and increase in those with a 3β,7α-dihydroxy-5-ene structure is evident in the ratio of 7αH,3O-CA(25R/S) to 3β,7α-diHCA(25R/S) which in control plasma is about 2.5 but in patient samples where HSD3B7 is deficient can be 10-fold smaller (Supplemental Figure S10 and Supplemental Table S1).

Deficiency in HSD3B7 leads to the production of increased amounts of bile acid precursors with a 3β,7α-dihydroxychol-5-en-24-oic acid (3β,7α-diH-Δ^5^-BA) and 3β,7α,12α-trihydroxychol-5-en-24-oic acid (3β,7α,12α-triH-Δ^5^-BA) structure conjugated with glycine or sulfuric acid or doubly conjugated with glycine and sulfuric acid (see Supplemental Table S1) (1, 2, 36). To observe sulfated bile acids it is necessary to analyse plasma in the absence of Girard derivatisation (blue and red boxes in Figure 4). The degree and type of conjugation appears to vary from patient to patient with *N*-acetylglucosamine (GlcNAc) conjugates of dihydroxycholenoic acid (diH-Δ^5^-BA), and also the glycine, glucuronic acid (GlcA) and GlcNAc triple conjugate being observed in plasma in one patient (89), although the sulfuric acid conjugate and the glycine, sulfuric acid double conjugate of dihydroxycholenoic and trihydroxycholenoic acids, probably with 3β,7α-diH-Δ^5^-BA and 3β,7α,12α-triH-Δ^5^-BA structures, dominate in others (85). The common theme for diagnosis of a deficiency in HSD3B7 is an elevation of metabolites with a 3β-hydroxy-5-ene structure, mostly a 3β,7α-dihydroxy-5-ene structure. This is also evident in oxysterol conjugates where double conjugates of hydroxycholesterols, dihydroxycholesterols and trihydroxycholesterols with two sulfuric acids or sulfuric acid and GlcA have been observed (Supplemental Figure S11) (90). Double conjugates provide a complication for ESI-MS analysis as the ratio of doubly to singly charged ions is dependent on the ion source conditions and often doubly charged ions are missed as they are frequently below the *m/z* range scanned.

#### Δ^4^-3-Oxosteroid-5β-reductase (AKR1D1) deficiency

A deficiency in the enzyme Δ^4^-3-oxosteroid-5β-reductase (5β-reductase deficiency) was first reported in 1988 (91, 92). However, this deficiency can be a secondary phenomenon in patients with severely damaged liver function (3). The disorder results in an increase in bile acid precursors with a 3-oxo-4-ene structure (Figure 2). It was not until 2003, following sequencing of the *AKR1D1* gene, that Lemonde et al identified the first patients with variants in *AKR1D1* and *bona fid*e Δ^4^-3-oxosteroid-5β-reductase deficiency (OMIM: 235555) (93). *AKR1D1* deficiency can be successfully treated by bile acid supplementation, providing the treatment starts before irreversible liver damage (3, 94). Amongst the patients identified by Lemonde et al was one particularly interesting individual described earlier by Clayton et al (95) who was initially treated with a combination of CDCA and cholic acid, but was found to self-correct for the disorder even in the absence of medication (94). Instead of the normal 5β(H)-bile acids, 5α(H)- (*allo*-bile) acids were produced being formed by a 5α-reductase and plasma concentrations of 3-oxo-4-ene bile acid precursors were found to have fallen to normal concentrations (94). It is noteworthy that Setchell et al reported increased synthesis of *allo*-bile acids in their 1988 paper on Δ^4^-3-oxosteroid-5β-reductase deficiency (92). The presence of allo-bile acids can be used to distinguish *bona fid*e Δ^4^-3-oxosteroid-5β-reductase deficiency from the secondary phenomena also presenting with elevated 3-oxo-4-ene bile acid precursors. However, unless the chromatography has been optimised the allo and conventional bile acid may overlap.

#### Oxysterols and bile acid precursors in AKRD1 deficiency

Lemonde et al found plasma concentrations of 7α-hydroxy-3-oxochol-4-en-24-oic (7αH,3O-Δ^4^-BA) and 7α,12α-dihydroxy-3-oxochol-4-en-24-oic (7α,12α-diH,3O-Δ^4^-BA) acids to be elevated in patients with AKR1D1 deficiency (93). Although we have not reported data for a patient with *bona fid*e AKR1D1 deficiency both these acids are observed by EADSA-LC-MS(MS^n^) as is 7αH,3O-CA(25R/S) and downstream metabolites 7α,12α-diH,3O-CA(25R) and 7α-hydroxy-3,24-*bis*oxocholest-4-en-(25R)26-oic acid (7αH,3,24-diO-CA(25R)) which should also be elevated in patient plasma (See Supplemental Table S1). Note, 7αH,3,24-diO-CA(25R) is also observed in EADSA-LC-MS(MS^n^) as the decarboxylation product 7α-hydroxy-27-*nor*-cholest-4-en-3,24-dione (7αH-27-*nor*-C-3,24-diO) (41). We have also found these metabolites to be elevated in patients with no observable variants in *AKR1D1* and with elevated bile acid plasma concentrations and concluded the 3-oxo-4-ene metabolic pattern to be a secondary phenomenon (Supplemental Figure S12).

Deficiency in Δ^4^-3-oxosteroid-5β-reductase leads to accumulation of conjugated bile acid precursors with a 3-oxo-4-ene structure, these include glycine and taurine conjugates of 7αH,3O-Δ^4^-BA and 7α,12α-diH,3O-Δ^4^-BA and in some cases of 7α,12α-diH,3O-CA. The production of CDCA and cholic acid is reduced and these are replaced by *allo*-bile acids. This can lead to confusion in LC-MS analysis unless these isomers are resolved. While non-amidated bile acids can be analysed following the EADSA protocol by LC-MS(MS^n^) and positive-negative ion switching, or preferably by negative-ion analysis of fraction-B alone in an extra LC-MS(MS^n^) injection, analysis of the glycine and taurine conjugates requires sample preparation in the absence of GP-reagent (Figure 4).

### Disorders of side-chain shortening

#### Bile acyl-CoA synthestase (BACS) deficiency

Side-chain shortening proceeds predominantly in the peroxisome (see Supplemental Figure S1). The first step is conversion of the C_27_-acid substrate into its CoA-thioester. This is achieved by the enzymes very long chain acyl-CoA synthetase (VLACS) and VLCS-homolouge2 (VLCSH2), also called BACS, coded by the genes *SLC27A2* and *SLC27A5*, respectively (96). Both enzymes are microsomal with VLACS also expressed in the peroxisome. These enzymes also activate long chain fatty acids. BACS will also use C_24_-bile acids as substrates and is responsible along with BAAT for the amidation of primary bile acids. Chong et al identified an infant with homozygous variants in a highly conserved residue in *SLC27A5* who presented with infantile cholestasis and with unconjugated bile acids in plasma and urine, indicating an amidation disorder (97). The fact that normal un-conjugated bile acids were synthesised indicates that side-chain shortening can proceed in the absence of BACS, but that C_24_ acids (formed following bacterial de-conjugation) cannot be efficiently re-amidated. As an infant this child was treated with ursodeoxycholic acid (UDCA), but after UDCA was withdrawn, and by the age of six the child was found to be asymptomatic. A sister with the same variants in *SLC27A5* is without clinical disease, but also shows the same deficiency in amidated bile acids.

We have not encountered a case of BACS deficiency but elevated concentrations of unconjugated bile acids and reduced amounts of glycine and taurine conjugated bile acids will be evident in samples analysed LC-MS prepared in the absence of derivatisation (red and blue boxes in Figure 4, see also Supplemental Table S1).

#### Alpha-methyl-acyl-CoA racemase (AMACR) deficiency

AMACR is required for peroxisomal side-chain shortening of C_27_ bile acid precursors (see Supplemental Figure S1). It converts the Co-A thioesters of cholestanoic and cholestenoic acids of 25R stereochemistry (derived from 25R-specific oxidation by CYP27A1) to their 25S-isomers, the required stereochemistry for substrates for the next enzyme, ACOX2, in the side-chain shortening pathway (98). Deficiency in AMACR (OMIM: 614307) can present as liver disease and cholestasis in childhood and in adults as progressive neurological symptoms (99–101). A deficiency in AMACR leads to a build-up of C_27_-bile acid precursors with 25R-stereochemistry in plasma and urine (5, 100, 101).

#### Oxysterols in AMARC deficiency

The cholestenoic acid 7αH,3O-CA is observed as the 25R- and 25S-isomers in human plasma, with the 25R-isomers being about 5-fold more abundant than the 25S-isomers (see Figure 6C, lower panel) (41). We have not reported a patient with AMACR deficiency but we have analysed plasma from the *Amacr^-/-^* mouse and compared this to plasma from the wild type animal (102). In the *Amacr^-/-^* mouse the concentration of the 25R-isomer is elevated, and the 25S-isomer no longer observed (see Supplemental Figure S13A), just as predicted by a deficiency of the racemase enzyme (102). In the current study we have re-analysed the mouse EADSA-LC-MS(MS^n^) data in greater detail and identified the further hydroxylated acid, 7α,12α-diH,3O-CA(25R), normally only a minor metabolite in human and mouse plasma, to be greatly elevated in plasma from the *Amacr^-/-^* mouse (Supplemental Figure S13B). In combination, this data indicates that EADSA-LC-MS(MS^n^) will be able to assist in the diagnosis of AMACR deficiency (see Supplemental Table S1).

#### Acyl-CoA oxidase 2 (ACOX2) deficiency

The enzyme coded by *ACOX2*, also known as branched-chain acyl-CoA oxidase, reduces the 25S-epimers of the CoA-thioesters of cholestanoic and cholestenoic acids by the introduction of a double bond at C-24 leading to *E* (trans) stereochemistry (Supplemental Figure S2) (98). The first case of ACOX2 deficiency (OMIM: 617308) was found in an adolescent presenting with liver fibrosis, ataxia and cognitive impairment and showing elevated plasma concentrations of di- and trihydroxycholestanoic acids and their taurine and to a lesser extent glycine conjugates (103). Other patients have since been found, a number of whom presented with hypertransaminasemia (104–106).

#### Oxysterols and bile acid precursors in ACOX2 deficiency

Elevated concentrations of both 25R- and 25S- epimers of 3β,7α-diHCA and 7αH,3O-CA are observed in ACOX2 deficiency, as are increased concentrations of 3β-HCA (Figure 11, see Supplemental Table S1) (41). The cholic acid precursor 7α,12α-diH,3O-CA is usually only observed as the 25R-epimer in plasma, however, deeper mining of our data performed in the current study, shows the ACOX2 substrate 7α,12α-diH,3O-CA(25S) is also observed in ACOX2 enzyme deficiency (Figure 11F). It is notable that both 25R- and 25S-epimers of 3β,7β-diHCA are elevated in ACOX2 plasma indicating that metabolism of the 25S-epimer is blocked by a deficiency of ACOX2, supporting the concept of endogenous biosynthesis of UDCA from 3β,7β-diHCA in human. Both 3β,5α,6β-trihydroxycholestan-26-oic acid (3β,5α,6β-triHCa) and 3β-hydroxy-7-oxocholest-5-en-(25R/S)-oic acid (3βH,7O-CA(25R/S)) are very minor acids in normal plasma (41), but as shown here Figure 11D & E their concentrations are elevated in ACOX2 deficiency compared to the NIST SRM 1950 reference material indicating that they are substrates of ACOX2.

**Figure 11.**
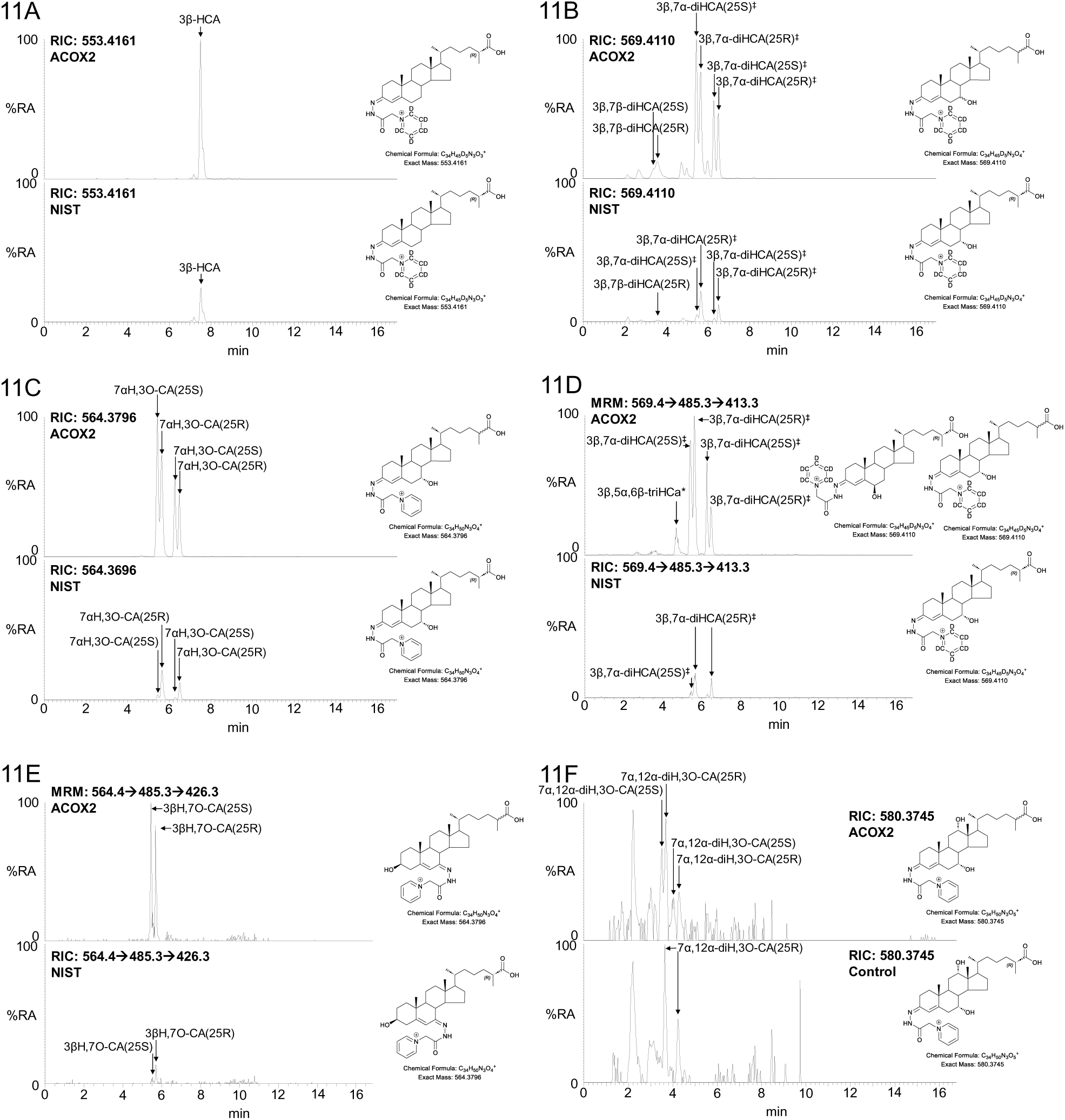
Analysis of ACOX2 (upper panels) and NIST SRM 1950 (lower panels) plasma by EADSA-LC-MS(MS^n^). The upper and lower panels are plotted on the same y-axis. (A) LC-MS RICs of *m/z* 553.4161 ± 5 ppm corresponding to 3β-HCA. (B) LC-MS RICs of *m/z* 569.4110 ± 5 ppm corresponding to 3β,7β-diHCA(25R/S) and 3β,7α-diHCA(25R/S) plus 7αH,3O-CA(25R/S). (C) LC-MS RICs of *m/z* 564.3796 ± 5 ppm corresponding to 7αH,3O-CA(25R/S). (D) MRM-like chromatograms *m/z* 569.4è485.3è413.3 targeting dehydrated 3β,5α,6β-triHCa (3β,5α,6β-triHCa*). The fragment ion at *m/z* 413.3 is enriched but not unique to the MS^3^ spectrum of dehydrated 3β,5α,6β-triHCa (57). (E) MRM-like chromatograms *m/z* 564.4è485.3è426.3 targeting 3βH,7O-CA(25R/S) (57). (F) LC-MS RIC of *m/z* 580.3745 ± 5 ppm corresponding to 7α,12α-diH,3O-CA(25R/S). Data acquired on the Orbitrap Elite using a Hypersil Gold C_18_ column. 3β,7α-diHCA^‡^ in (B) & (D) indicates that 3β,7α-diHCA and 7αH,3O-CA are measured in combination. Control and NIST SRM 1950 data were previously reported in (41, 54). Relevant MS^3^ spectra are shown in Supplemental Figure S14. The data in chromatogram (B) was previously reported in (41). The symbol (*) indicates the dehydrated molecule.

Besides the unsaturated C_27_ bile acids revealed by EADSA-LC-MS(MS^n^), negative-ion mode LC-MS analysis of plasma from ACOX2 patients shows patterns of elevated saturated di- and tri-hydroxycholestanoic acids and their glycine and taurine conjugates where both the 25R- and 25S-isomers are observed (Supplemental Table S1), but in most cases where the 25S-isomer is dominant (Supplement Figure S15). To observe the glycine and taurine conjugates it was necessary to prepare the samples in the absence of Girard reagent (red and blue boxes in Figure 4).

#### D-Bifunctional protein (DBP) deficiency

DBP hydrates the 24(*E*) double bond in cholest-24*E*-en-26-oyl-CoA substrates (see Figure S2) then oxidises the resulting 24R-hydroxy group to a ketone (5, 107). It also plays an essential role in the peroxisomal degradation of long-chain and branched-chain fatty acids. Deficiency in DBP (OMIM: 261515) can lead to three types of disorder. Type I is where there is deficiency in both hydratase and dehydrogenase activity, type II is where there is only hydratase deficiency, and type III is a deficiency in dehydrogenase activity alone. The clinical presentation of DBP deficiency is usually severe, with few patients showing developmental progress, but some patients have been identified with a relatively mild phenotype correlating with residual enzyme activity (108).

DBP deficiency leads to elevated plasma concentration of 3α,7α-di- and 3α,7α,12α-trihydroxy-5β-cholestan-26-oic acids (3α,7α-diHCa and 3α,7α,12α-triHCa), presumably both 25R and 25S-epimers, and their taurine conjugates (3α,7α-diHCa-(25R/S)26-Tau and 3α,7α,12α-triHCa-(25R/S)26-Tau) (see Supplemental Table S1) (5, 107). When hydratase activity is maintained but dehydrogenase activity deficient (type III) then there is a build-up in 3α,7α,12α,24R-tetrahydroxy-5β-cholestan(25R)26-oic acid (3α,7α,12α,24R-tetraHCa(25R)), while its diastereoisomer 3α,7α,12α,24S-tetrahydroxy-5β-cholestan(25S)26-oic acid (3α,7α,12α,24S-tetraHCa(25S)), formed via LBP activity, is elevated in types I and II disease (109, 110). There are similar elevations in the corresponding taurine conjugates. In all three types of the disease there is an elevation in 3α,7α,12α-trihydroxycholest-24*E*-en-26-oyl-taurine (3α,7α,12α-triH-Δ^24*E*^-Ca-26-Tau) (5, 109).

We have not reported analysis of a plasma sample from a patient with DBP deficiency but based on the published patterns of bile acid precursors (5, 109–111), we would expect to see, using EADSA-LC-MS(MS^n^), elevated concentrations of 3β,7α-diHCA(25S), 7αH,3O-CA(25S) (cf. Figure 11B & C), 3β,7α-dihydroxycholesta-5,24*E*-dien-26-oic (3β,7α-diH-Δ^24*E*^-CA) and 7α-hydroxy-3-oxocholesta-4,24*E*-dien-26-oic (7αH,3O-Δ^24*E*^-CA) acids in all three types of disease; elevated 3β,7α,24S-trihydroxycholest-5-en-(25S)26-oic (3β,7α,24S-triHCA(25S)) and 7α,24S-dihydroxy-3-oxocholest-4-en-(25S)26-oic (7α,24S-diH,3O-CA(25S)) acids and in types in types I and II deficiency; and 3β,7α,24R-trihydroxycholest-5-en-(25R)26-oic (3β,7α,24R-triHCA(25R)) and 7α,24R-dihydroxy-3-oxo-cholest-5-en-(25R)26-oic (7α,24R-diH,3O-CA(25R)) acids in type III deficiency (see supplemental Figure S2 and Supplemental Table S1). By switching to negative-ion mode the unconjugated bile acid precursors described above are observed, but glycine and taurine conjugates would only be observed following sample preparation in the absence of GP-reagent.

#### Sterol carrier protein 2 (SCP2) deficiency

SCP2, also called SCPx, catalyses the last step in the side-chain shortening process (Supplemental Figure S2) (5, 107). SCP2 deficiency (OMIM: 613724) is very rare with only three cases reported. The first two cases showed homozygous or compound heterozygous variants in *SCP2* (112, 113), while the third showed a heterozygous variant (114). All three patients show neurological abnormalities and biochemically elevated phytanic and pristanic acids, although to a lower degree in the third patent. Bile acid precursors were only analysed in the first study (112), where findings were consistent with studies of the *Scp2* knock-out mouse (115). In the knock-out mouse a deficiency in SCP2 enzyme leads to elevated plasma concentrations of 3α,7α,12α-trihydroxy-27-*nor*-5β-cholestan-24-one (3α,7α,12α-triH-27-*nor*-C-24O), a decarboxylation product of 3α,7α,12α-trihydroxy-24-oxo-5β-cholestan-(25R)26-oic acid (3α,7α,12α-triH,24O-Ca(25R)), which would be expected to be elevated in the absence of SCP2 activity (see Supplemental Figure S2, SCP2 shunt). Although, 3α,7α,12α-triH-27-*nor*-C-24O was not observed in human plasma, ESI-MS data suggested the presence of glucuronides of tetrahydroxy-27-*nor*-5β-cholestan-24-one, pentarahydroxy-27-*nor*-5β-cholestan-24-one and hexahydroxy-27-*nor*-5β-cholestan-24-one in urine (112).

Although EADSA-LC-MS(MS^n^) has not been used to analyse plasma from patients with SCP2 deficiency, 7α-hydroxy-27-*nor*-cholest-4-en-3,24-dione (7αH-27-*nor*-C-3,24-diO) is a normal, but minor, component of human plasma formed through decarboxylation of 7α-hydroxy-3,24-*bis*oxocholest-4-en-(25R)26-oic acid (7αH,3,24-diO-CA(25R)) via a shunt to the acidic pathway of bile acid biosynthesis (see supplemental Figure S2 and Figure S12A) (41), and should be elevated in SCP2 deficiency. Switching to negative-ion mode should allow detection of the 27-*nor*-5β-cholestanone glucuronides (see Supplemental Table S1).

#### Disorders of amidation

BACS (VLCSH2, *SLC27A5*) converts both C_27_ and C_24_ bile acids to the corresponding acyl-CoAs. VLACS (*SLC27A2*) has similar activity towards C_27_ bile acids. Bile acyl-CoAs are substrates for BAAT which catalyses amidation with glycine and taurine. A deficiency in BAAT (OMIM: 619232) results in high concentrations of unconjugated bile acids leading to fat malabsorption (116). In serum besides high concentrations of cholic acid, high concentrations of sulfated monohydroxycholenoic, dihydroxycholanoic and trihydroxycholanoic acids are observed along with glucuronides of dihydroxycholanoic and trihydroxycholanoic acids, predominantly of CDCA and cholic acids (3, 116).

Negative-ion analysis is most efficient for diagnosis of BAAT as the diagnostic ions are transparent to EADSA.

#### Lysosomal storage disorders

The lysosomal storage disorders NPC1 (OMIM: 257220), NPC2 (OMIM: 607625), ASMD (NPB, OMIM: 607616), Wolman disease (LALD, OMIM; 620151) and cholesterol ester storage disease (CESD, lysosomal acid lipase deficiency partial, OMIM: 278000) all show elevated metabolites derived from non-enzymatic oxidation of cholesterol (34–38, 57). In NPC1, NPC2 and ASMD disorders non-esterified cholesterol is trapped in the lysosome, while in Wolman disease and CESD the enzyme lysosomal acid lipase (LAL, *LIPA*) is deficient resulting in a build-up of cholesterol esters in the lysosome. A consequence of these biochemical deficiencies is that the normal negative-feedback mechanism on cholesterol synthesis following receptor mediated endocytosis of low density lipoprotein particles (LDL) is not activated (117). The biochemical result of this is enhanced non-enzymatic oxidation of cholesterol (Figure 3). This opens up three pathways of metabolism starting from the autoxidation products, 7β-HC, 7-OC and cholestane-5,6-epoxide (5,6-EC), proceeding through to 3β,7β-diH-Δ^5^-BA, 3β-hydroxy-7-oxochol-5-en-24-oic acid (3βH,7O-Δ^5^-BA) and 3β,5α,6β-triHBA (via 3β,5α,6β-triol), respectively. Elevation of the initial oxysterols, the pathway intermediates and C_24_ bile acids in plasma are detected by EADSA-LC-MS(MS^n^) (38, 57).

#### Oxysterols, bile acid precursors and bile acids in NPC, ASMD and Wolman disease

The oxysterols 7β-HC and 3β,5α,6β-triol are usually minor oxysterols in plasma (<5 ng/mL) (41). Both are measured by EADSA-LC-MS(MS^n^), with the triol being measured as the dehydrated molecule (41). In the case of NPC, ASMD and Wolman disease, 7β-HC, 7-OC and 3β,5α,6β-triol are often elevated by a factor of more than ten (Figure 12A,B and Supplemental Figure S17A,B) (38). A disadvantage of using these three oxysterols in the diagnosis of a lysosomal storage disorder is that they can also be formed via *ex vivo* oxidation in air during sample handling, storage and work-up. However, only endogenous molecules become enzymatically metabolised to e.g. the C_27_ acid 3β,7β-diHCA (measured as the 25R and 25S-epimers combined) usually elevated from about 10 ng/mL in controls to 50 – 100 ng/mL or more in patient samples (Figure 12C & Supplemental Figure S17C). The C_27_ acids 3βH,7O-CA(25R/S) and 3β,5α,6β-triHCa (measured as the dehydrated ion) are more difficult to analyse than 3β,7β-diHCA(25R/S) as they elute close to other isomeric compounds and require a multiple reaction monitoring (MRM)-like approach for quantification. The MRM transition [M]^+^è[M-Py]^+^è[M-Py-59]^+^ (564.4è485.3è426.3) is unique to 3βH,7O-CA(25R/S) allowing its relative quantification, while the MRM transition [M-18]^+^è[M-18-Py]^+^è[M-18-Py-72] (569.4è485.3è413.3), although not unique to 3β,5α,6β-triHCa, can provide its relative quantification as 3β,5α,6β-triHCa is chromatographically resolved from other sterols showing this transition (Figure 12D,E & Supplemental Figure S17D,E). In patient samples both acids are usually elevated by a factor of ten compared to controls (see Supplemental Table S1) (38). The down-stream C_24_ bile acids 3β,5α,6β-triHBA (measured as the dehydrated ion) and 3βH,7O-Δ^5^-BA are either absent or present at very low concentrations in control plasma (<1 ng/mL), while 3β,7β-diH-Δ^5^-BA is a normal, although minor, component of control plasma (<5 ng/mL) (41). In patient samples 3β,5α,6β-triHBA becomes detectable, while both 3βH,7O-Δ^5^-BA are 3β,7β-diH-Δ^5^-BA, are in many patient samples, elevated by a factor of ten compared to controls (Figures 12F,G and Supplemental Figure S17F,G). Note, 7-oxo metabolites are quantified from fraction-B (Figure 4, even *m/z*), this avoids the complication of incomplete oxidation by cholesterol oxidase (in fraction-A) due of the presence of the 7-oxo group and also the formation of double derivatives from the 3,7-dione. While Wolman disease is characterised by a complete lack of LAL activity, CESD is a milder later onset disorder where some LAL activity is maintained. We have not reported the oxysterol profile of CESD but predict a similar pattern to Woman disease, but with a lesser elevation of 7β-hydroxy, 7-oxo and 3β,5α,6β-triol metabolites of cholesterol.

**Figure 12.**
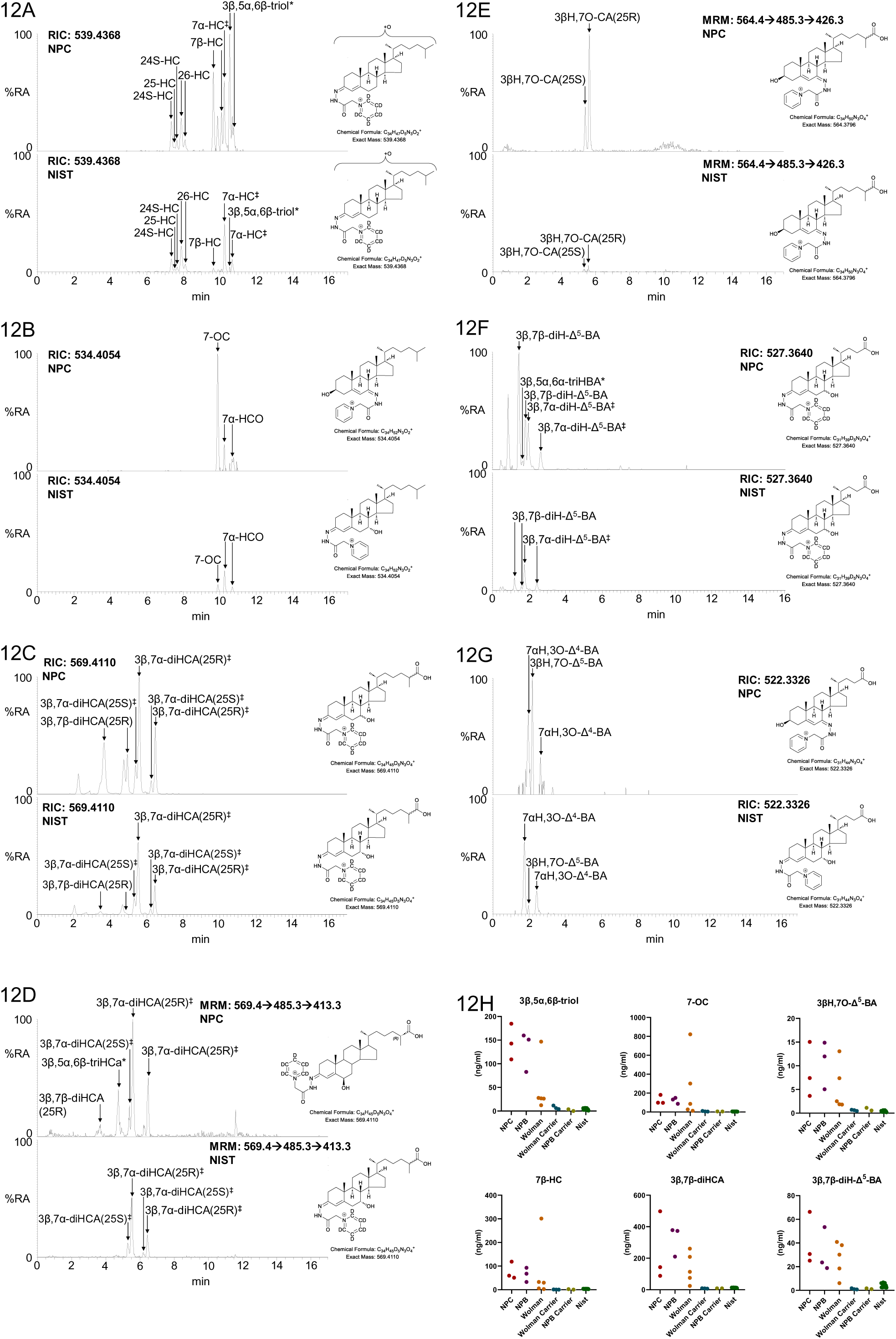
Analysis of NPC (upper panels) and NIST SRM 1950 (lower panels) plasma by EADSA-LC-MS(MS)^n^. The upper and lower panels are plotted on the same y-axis. (A) LC-MS RICs of *m/z* 539.4368 ± 5 ppm corresponding to monohydroxycholesterols including 7β-HC and dehydrated 3β,5α,6β-triol (3β,5α,6β-triol*). (B) LC-MS RICs of *m/z* 534.4054 ± 5 ppm corresponding to monohydroxycholestenones including 7-OC. (C) LC-MS RICs of *m/z* 569.4110 ± 5 ppm corresponding to isomers of 3β,7β-diHCA(25R/S) and 3β,7α-diHCA(25R/S). (D) MRM-like chromatograms *m/z* 569.4è485.3è413.3 targeting dehydrated 3β,5α,6β-triHCa (3β,5α,6β-triHCa*). The fragment ion at *m/z* 413.3 is enriched but not unique to the MS^3^ spectrum of dehydrated 3β,5α,6β-triHCa (57). (E) MRM-like chromatogram *m/z* 564.4è485.3è426.3 targeting 3βH,7O-CA(25R/S) (57). (F) LC-MS RIC of *m/z* 527.3640 ± 5 ppm corresponding to isomers of 3β,7β-diH-Δ^5^-BA, 3β,7α-diH-Δ^5^-BA and dehydrated 3β,5α,6β-triHBA (3β,5α,6β-triHBA*). (G) LC-MS RIC of *m/z* 522.3326 ± 5 ppm corresponding to hydroxyoxocholenoic acids. Data acquired on the Orbitrap Elite using a Hypersil Gold C_18_ column. (H) Concentrations of 7β-HC, 7-OC, 3β,5α,6β-triol* and their downstream metabolites in NPC, NPB (ASMD) and Wolman patient plasma samples compared to carrier individuals and NIST SRM1950. The symbol (^‡^) indicates that 3β-hydroxy-5-ene and 3-oxo-4-ene metabolites are measured in combination. The symbol (*) indicates the dehydrated molecule.

EADSA-LC-MS(MS^n^) is conducted in the positive-ion mode, this also allows the detection of PPCS (*N*-hexadecanoyl-*O*-phosphocholine-L-serine, see Figure 3), also called Lyso-SM509, which is elevated in NPC and NPB (39) and sphingosine-1-phosphocholine (sphing-4-enine-1-phosphocholine, SM(d18:1/0:0) also called lysosphingomyelin. The ratio of palmitoyl-*O*-phosphocholineserine to sphingosine-1-phosphocholine has been suggested as a marker to differentiate NPC from NPB, the latter analyte not being consistently elevated in NPC (40). Lysophospholipids co-purify with oxysterols and cholestenoic acids and are usually regarded as unwanted contaminants, but for NPC and NPB become target analytes. Insufficient data has been acquired to assess whether they may be biomarkers for Wolman disease or CESD.

There is a growing realisation of the potential for monitoring unusual bile acids as biomarkers for NPC in newborn screening (36, 37, 118). The atypical bile acid 3β,5α,6β-triHBA-24-Gly is the preferred bile acid biomarker for NPC (Figure 13B,C) (33, 36, 37). Clayton and colleagues have suggested that plasma concentrations of 3β,5α,6β-triHBA-24-Gly above 90 nM (40 ng/mL) as diagnostic for NPC (119). In addition to the glycine conjugate, Ory and colleagues have shown that the unconjugated acid, 3β,5α,6β-triHBA, and its taurine conjugate 3β,5α,6β-trihydroxycholan-24-oyl-taurine (3β,5α,6β-triHBA-24-Tau) are elevated in NPC (Figure 13A,D, Supplemental Table S1) (37). This series of bile acids is not unique to NPC, the unconjugated acid is observed at increased concentrations in Wolman disease and ASMD (NPB), although an insufficient number of samples have been analysed to confirm if the concentrations of this acid and its conjugates are similar to those seen in NPC (Supplemental Figure S19 & S20). Care should be taken when interpreting bile acid data, as cholestasis will also result in elevated concentrations of these and other unusual bile acids. The first bile acids to be suggested to be biomarkers for NPC were GlcNAc conjugates of 3β,7β-diH-Δ^5^-BA (i.e. 3βH,7β-GlcNAc-Δ^5^-BA), its glycine (3βH,7β-GlcNAc-Δ^5^-BA-24-Gly), sulfate (3βS,7β-GlcNAc-Δ^5^-BA), glycine and sulfate (3βS,7β-GlcNAc-Δ^5^-BA-24-Gly), and taurine and sulfate (3βS,7β-GlcNAc-Δ^5^-BA-24-Tau) conjugates (see Figure 3) (35). The current study revealed a similar pattern in some patients with ASMD (n = 3) and Wolman disease (n = 2), although very few patients were investigated, it is also probable that patients with CESD will show this pattern of GlcNAc conjugates (Figure 14, Supplemental Figure S22 and Dupplemental Table S1). However, the enzyme UDP *N*-acetylglucosaminyl transferase 3A1 (UGT3A1) which introduces the GlcNAc group at C-7β, can be inactive in 20% of Asians and Caucasians due to a common homozygous polymorphism, so these conjugates are not ideal biomarkers of NPC, ASMD or Wolman disease (36). While non-amidated bile acids are analysed by LC-MS following the standard EADSA protocol exploiting negative-ion analysis, glycine, taurine and or sulfate conjugates are lost in the derivatisation process and require sample preparation in the absence of GP-reagent.

**Figure 13.**
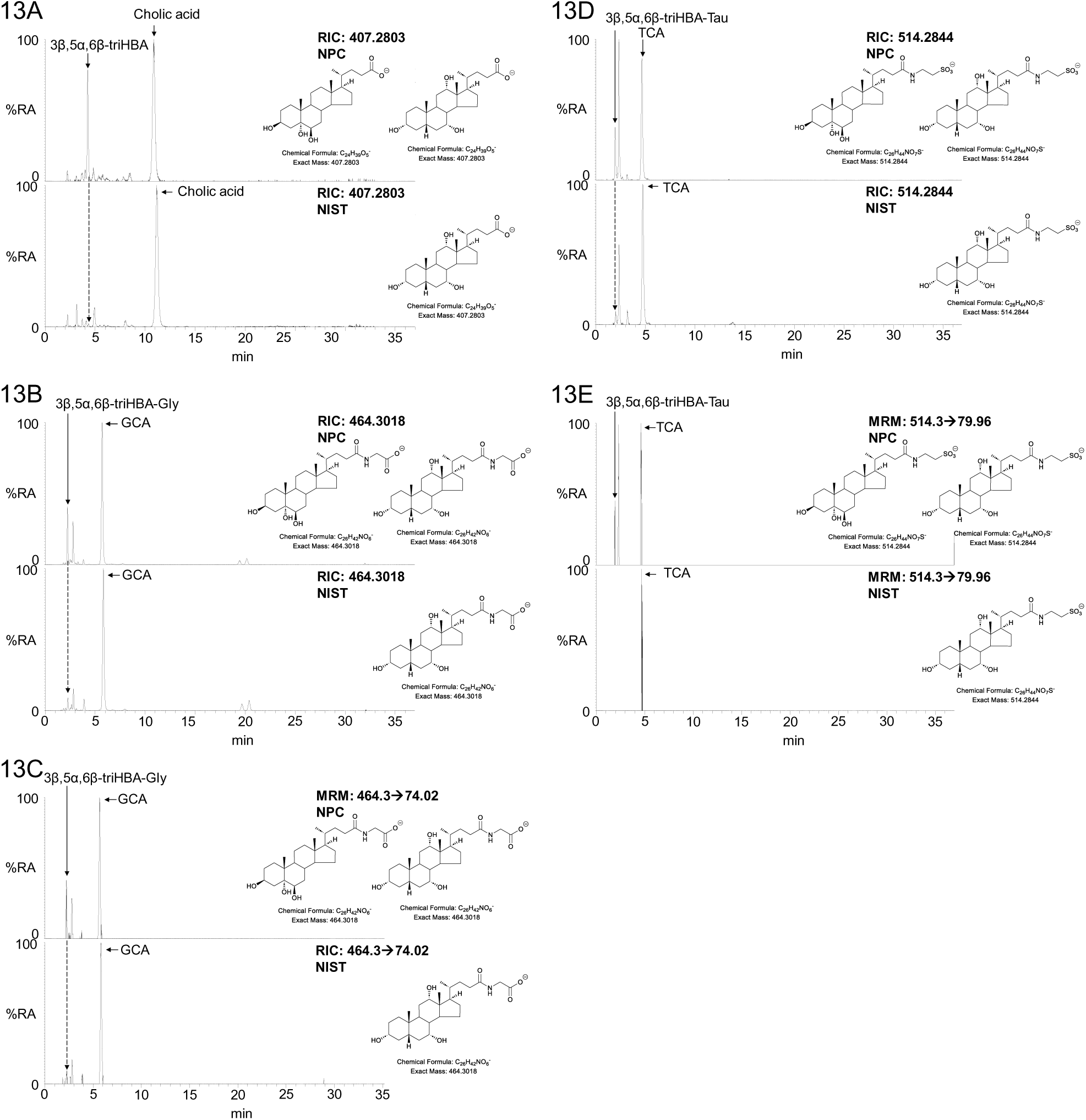
Analysis of NPC (upper panels) and NIST SRM 1950 (lower panels) plasma by negative-ion LC-MS(MS/MS) without enzyme or GP-treatment. LC-MS RICs of (A) *m/z* 407.2803 ± 5 ppm corresponding to trihydroxycholanoic acids including 3β,5α,6β-triHBA and cholic acid; (B) *m/z* 464.3018 ± 5 ppm corresponding to isomers of trihydroxycholanoyl-glycine including 3β,5α,6β-triHBA-24-Gly and glycocholic acid (GCA). (C) MRM-like transition [M-H] → 74.0248 ± 20 ppm giving the fragment ion NH_2_CH_2_CO ^-^ from trihydroxycholanoyl-glycine isomers. (D) RICs of *m/z* 514.2844 ± 5 ppm corresponding to trihydroxycholanoyl-taurine including 3β,5α,6β-triHBA-24-Tau and taurocholic acid (TCA). (E) MRM-like transition [M-H] → 79.9574 ± 20 ppm giving the fragment ion SO ^-^ from trihydroxycholanoyl-taurine isomers. Note labelling of the numeral indicating the position of amidation is omitted. Data acquired on the Orbitrap IQX using a Hypersil Gold C_18_ column. Fragmentation spectra can be found in Supplemental Figure S18.

**Figure 14.**
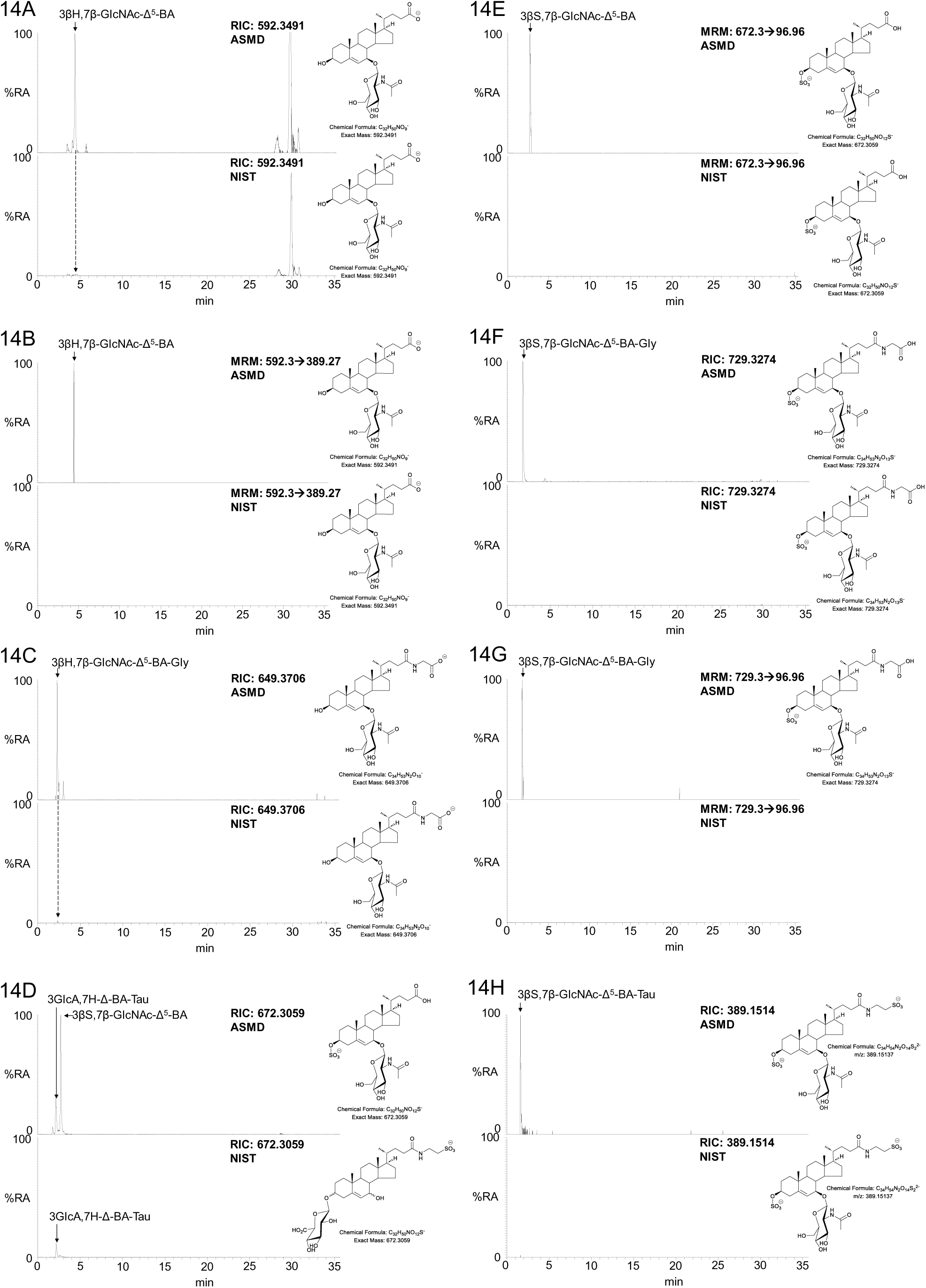
Analysis of ASMD (NPB, upper panels) and NIST SRM 1950 (lower panels) plasma by negative-ion LC-MS(MS/MS) without enzyme or GP-treatment. The upper and lower panels are plotted on the same y-axis, allowing for differences in injected plasma volume. (A) LC-MS RICs of *m/z* 592.3491 ± 3 ppm corresponding to 3βH,7β-GlcNAc-Δ^5^-BA; (B) MRM-like transition [M-H] → [M-H-203.0794]^-^ ± 20 ppm corresponding to the loss of the GlcNAc conjugating group from 3βH,7β-GlcNAc-Δ^5^-BA; (C) LC-MS RICs of *m/z* 649.3706 ± 5 ppm corresponding to 3βH,7β-GlcNAc-Δ^5^-BA-24-Gly; (D) LC-MS RIC of *m/z* 672.3059 ± 5 ppm corresponding to 3βS,7β-GlcNAc-Δ^5^-BA; (E) MRM-like transition [M-H] → 96.9601 ± 20 ppm giving the HSO ^-^ ion from 3βS,7β-GlcNAc-Δ^5^-BA; (F) LC-MS RICs of *m/z* 729.3274 ± 5 ppm corresponding to 3βS,7β-GlcNAc-Δ^5^-BA-24-Gly; (G) MRM-like transition [M-H]^-^è 96.9601 ± 20 ppm giving the HSO ^-^ ion from 3βS,7β-GlcNAc-Δ^5^-BA-24-Gly and (H) LC-MS RICs of *m/z* 389.1514 ± 5 ppm corresponding to doubly charged 3βS,7β-GlcNAc-Δ^5^-BA-24-Tau. Data acquired on the Orbitrap IQX using a Hypersil Gold C_18_ column. Fragmentation spectra can be found in Supplemental Figure S21.

The *CH25H* (*OMIM: 604551*) and *LIPA* (*OMIM: 613497*) genes are consecutive genes on chromosome 10. Goenka et al discovered a cluster of infants with a biallelic deletion of these two genes (47). Interestingly, of four patients who received BCG vaccination, three showed localised BCG abscess. In contrast, no BCG abscesses were reported in a group of eight children with only *LIPA* deficiency (47).

The plasma oxysterol patterns as determined by EADSA-LC-MS(MS^n^) of these patients was typical of Wolman disease, showing elevated concentrations of 3β,5α,6β-triol, 7β-HC and 7-OC; their C_27_ acids 3β,5α,6β-triHCa, 3β,7β-diHCA and 3βH,7O-CA and C_24_ acids 3β,5α,6β-triHBA, 3β,7β-diH-Δ^5^-BA and 3βH,7O-Δ^5^-BA, however, 25-HC was absent (47, 120). It should be noted however, that low concentrations of 25-HC are often a feature of Wolman disease even in the presence of functional CH25H. Surprisingly, 7α,25-dihydroxysterols were present in plasma of these children, indicating the presence of a second sterol 25-hydroxylase. The bile acid profiles in plasma from the children with the *CH25H* gene deletion is indistinguishable from that of children with classical Wolman disease.

#### Phytosterolaemia

Phytosterolaemia, also called sitosterolaemia, is a rare autosomal recessive disorder resulting from homozygous or compound heterozygous variants in either of the genes *ABCG5* (sitosterolemia 2, OMIM: 618666) or *ABCG8* (sitosterolemia 1, OMIM: 210250) coding the heterodimeric ABCG5/ABCG8 intestinal and hepatic efflux transporter (121). The disorder presents with an accumulation of phytosterols in plasma (122). Phytosterols are readily quantified by EADSA-LC-MS(MS^n^) and here we present new data from two children with the disorder where the dominant phytosterols sitosterol and campesterol were elevated by factors of more than 50 and 15, respectively (see Figure 15 and Supplemental Table S1). Other phytosterols including stigmasterol and brassicasterol were elevated by similar degrees. Analysis of phytosterols is important as phytosterolaemia can, like CTX, present with xanthoma. However, EADSA-LC-MS(MS^n^) easily differentiates between the two disorders.

**Figure 15.**
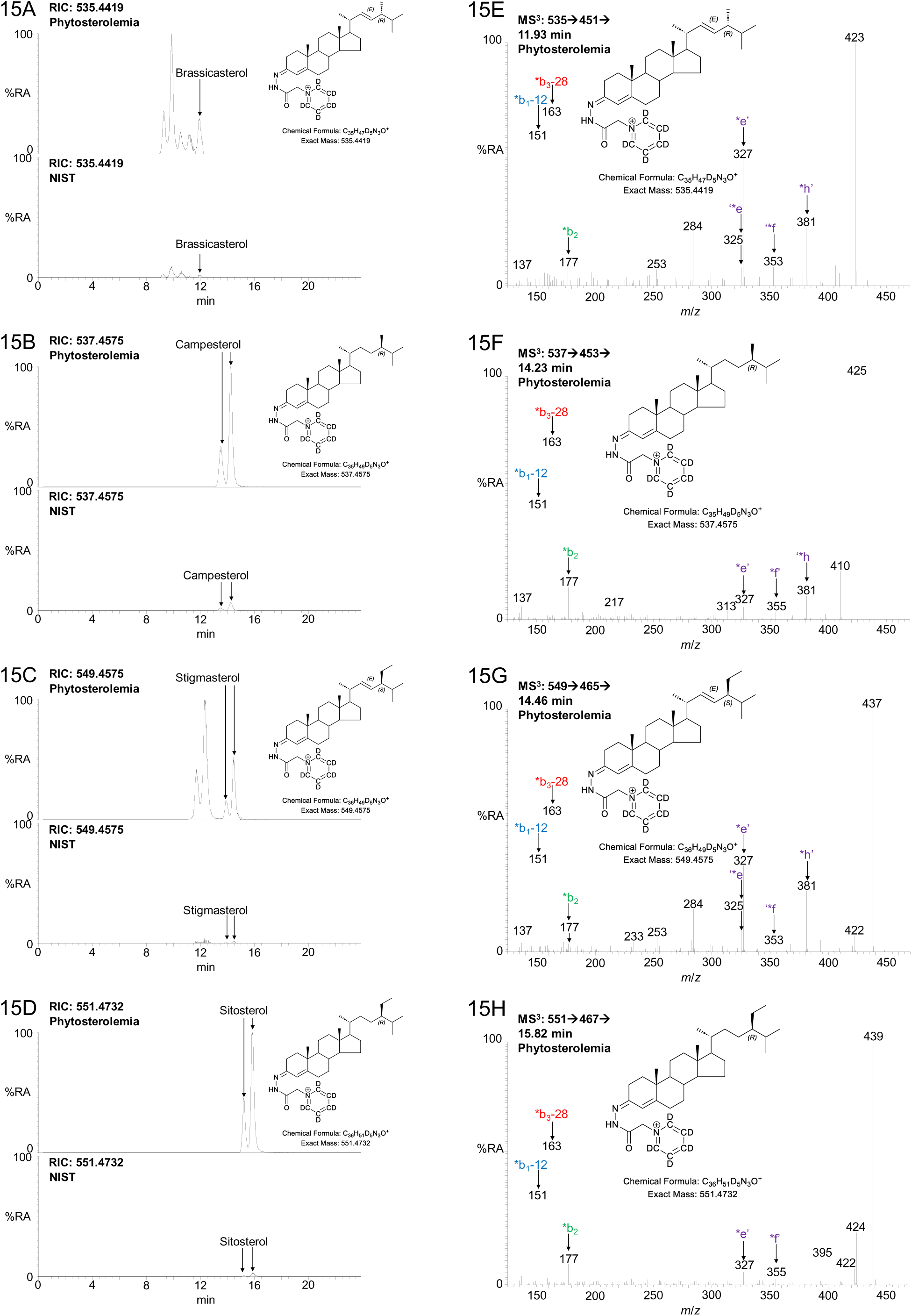
Figure 5. EADSA-LC-MS(MS^n^) analysis of plasma from a phosterolemia patient and of the NIST SRM 1950 standard reference material (65). LC-MS RICs ± 5 ppm for the phytosterols (A) brassicaterol; (B) campesterol; (C) stigmasterol; and (D) sitosterol, from patient plasma (upper panel) and in NIST SRM 1950 plasma (lower panel). The upper and lower panels are plotted on the same y-axis. MS^3^ ([M]^+^ → [M-Py]^+^ →) spectra of (E) brassicasterol; (F) campesterol; (G) stigmasterol; and (H) sitosterol. Fragmentation nomenclature is described in (41). Data acquired on the Orbitrap IQX using a Hypersil Gold C_18_ column. Some chromatograms have been re-aligned to correct for retention-time drift.

#### Algorithm for diagnosis

To gain maximum amount of information about a given sample the EADSA protocol with positive-ion LC-MS(MS^n^) of fractions-A and -B combined, derived from both the oxysterol (Fr1) and cholesterol (Fr3) rich fractions with also a separate analysis for bile acids in fraction-C prepared in the absence of derivatisation and analysed in the negative-ion mode was performed (see Figure 4). However, the protocol can be greatly simplified depending on the information requirements (Figure 16 and Supplemental Table S2).

1. By omitting preparation of fraction-C and analysing fraction-B instead in the negative-ion mode, information on non-amidated bile acids and glucuronidated-bile acids, -bile alcohols and -oxysterols is still provided although sulfated and amidated bile acids are lost. However, information on amidation is only essential for diagnosis of BACS and BAAT deficiencies (Figure 16).
2. If only cholesterol and its precursors or phytostrols are of interest, the first SPE step (SPE1) in the EADSA protocol can be omitted and the 70% ethanol single phase plasma-extract vacuum concentrated prior to treatment with cholesterol oxidase, GP-reagent and analysis by LC-MS(MS)^n^. Preferably, excess GP-reagent is removed by a reversed-phase SPE step or via a trap column as part of the LC injection system. This allows the definition of lathosterolosis, SLOS, phytosterolemia and in theory MEND and desmosterolosis. It is not necessary to separate A- and B-fractions as the cholesterol precursors monitored do not contain oxo groups so will only be derivatised in the presence of cholesterol oxidase.
3. For a number of disorders, diagnosis can be made from oxysterols with a natural 3-oxo-4-ene function e.g. CTX, HSD3B7, AMACR, ACOX2 deficiencies and in theory AKR1D1, DBP and SCP2 deficiencies, in which case after the first SPE step it is not necessary to divide the eluate into A- and B-fractions, but simply dry down and incubate the with GP-reagent in the absence of cholesterol oxidase.
4. On the other hand, some oxysterols exist almost completely in the 3β-hydroxy form in which case their analysis does not require generation of separate A- and B-fractions, so the entire dried eluate from SPE1 is incubated with cholesterol oxidase followed by GP-derivatisation. This allows the diagnosis of e.g. SPG5A, NPC, ASMD, Wolman disease, Wolman disease with CH25H deficiency and in theory CESD. We have yet to establish the best diagnostic molecules to differentiate NPC, ASMD, Wolman disease and CESD, although the ratio of PPCS to SM(d18:1/0:0) has been suggested to differentiate NPC from ASMD.
5. The EADSA protocol is not applicable to the amidation disorders, i.e. BACS and BAAT deficiencies, where bile acid analysis is necessary. This can be performed in the negative-ion mode on fraction-C (blue or red boxes in Figure 4) to confirm an absence of both glycine and taurine bile acid conjugates.
6. From the above, it is clear that diagnosis of a suspected metabolic disorder does not require the full EADSA protocol with analysis of fractions-A, -B and -C. However, this is not the case where the cause of disease is unknown or in instances of late onset disease where there may be only partial enzyme deficiency in which case the more metabolic information available the better. It should also be noted that cholestasis can lead to a misdiagnosis in the absence of complete metabolic information, as cholestasis can lead to a build-up of multiple bile acids and their precursors in blood confusing data interpretation. However, a simple switch from positive to negative-ion analysis for analysis of fraction-B will indicate elevated bile acids and the presence of cholestasis or not.

**Figure 16.**
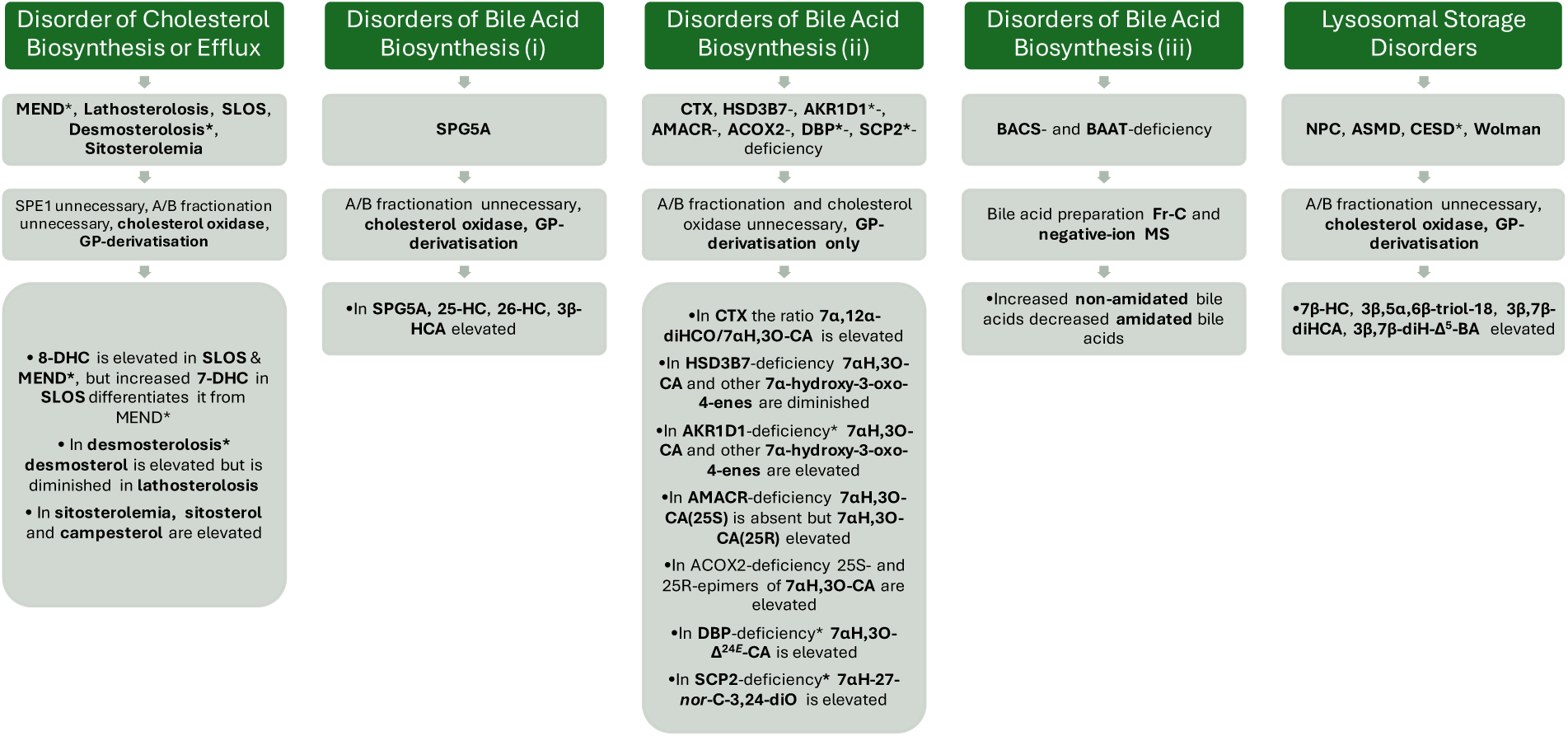
Simplest EADSA protocol for targeted analysis of cholesterol related in-born disorders of metabolism. The asterisk (*) symbol is used to indicate predicted patterns of metabolites.

## Conclusion

Here we describe how the EASDA protocol can be exploited to identify multiple cholesterol related disorders. Where a specific disorder is suspected the method can be simplified to allow analysis of defined analytes. However, where the disorder is not known or in cases of late on-set disorder where there may be only a partial loss in enzyme activity a full EADSA protocol is recommended. This is also true for cases of suspected variants of uncertain significance.

This is the first iteration of our mass spectrometry sterolomic library, we intend to publish up-dates in future years updating “predicted” metabolic patterns to “published” patterns.

## Data Availability

All data produced in the present study are available upon reasonable request to the authors.

## Acknowledgement

We are indebted to patients for generously donating samples.

## Abbreviations

3α,7α-diHCa: 3α,7α-dihydroxy-5β-cholestan-26-oic acid
3α,7α-diHCa-(25R/S)26-Tau: 3α,7α-dihydroxy-5β-cholestan-(25R/S)26-oyl-taurine
3α,7α,12α-triH,24O-Ca(25R): 3α,7α,12α-trihydroxy-24-oxo-5β-cholestan-(25R)26-oic acid
3α,7α,12α-triH-27-*nor*-C-24O: 3α,7α,12α-trihydroxy-27-*nor*-5β-cholestan-24-one
3α,7α,12α-triHCa: 3α,7α,12α-trihydroxy-5β-cholestan-26-oic acid
3α,7α,12α-triHCa-(25R/S)26-Tau: 3α,7α,12α-trihydroxy-5β-cholestan-(25R/S)26-oyl-taurine
3α,7α,12α-triH-Δ^24*E*^-Ca-26-Tau: 3α,7α,12α-trihydroxycholest-24*E*-en-26-oyl-taurine
3α,7α,12α,24R-tetraHCa(25R): 3α,7α,12α,24R-tetrahydroxy-5β-cholestan(25R)26-oic acid
3α,7α,12α,24S-tetraHCa(25S): 3α,7α,12α,24S-tetrahydroxy-5β-cholestan(25S)26-oic acid
3β,5α,6β-triHCa: 3β,5α,6β-trihydroxycholestan-26-oic
3β,5α,6β-triHBA: 3β,5α,6β-trihydroxycholan-24-oic acid
3β,5α,6β-triHBA-24-Gly: 3β,5α,6β-trihydroxycholan-24-oyl-glycine
3β,5α,6β-triHBA-24-Tau: 3β,5α,6β-trihydroxycholan-24-oyl-taurine
3β,5α,6β-triol: cholestane-3β,5α,6β-triol
3β,7α,24R-triHCA(25R): 3β,7α,24R-trihydroxycholest-5-en-(25S)26-oic
3β,7α,24R-triHCA(25S): 3β,7α,24S-trihydroxycholest-5-en-(25S)26-oic
3β,7α-diHCA(25R/S): 3β,7α-dihydroxycholest-5-en-(25R/S)26-oic acid
3β,7α-diH-Δ^5^-BA: 3β,7α-dihydroxychol-5-en-24-oic acid
3β,7α-diH-Δ^24*E*^-CA: 3β,7α-dihydroxycholesta-5,24*E*-dien-26-oic
3β,7α,12α-triH-Δ^5^-BA: 3β,7α,12α-trihydroxychol-5-en-24-oic
3β,7β-diHCA(25R/S): 3β,7β-dihydroxycholest-5-en-(25R/S)26-oic acid
3β,7β-diH-Δ^5^-BA: 3β,7β-dihydroxychol-5-en-24-oic acid
3βH,7β-GlcNAc-Δ^5^-BA: 3β-hydroxy,7β-*N*-acetylglucosaminyl-chol-5-en-24-oic acid
3βH,7β-GlcNAc-Δ^5^-BA-24-Gly: 3β-hydroxy,7β-*N*-acetylglucosaminyl-chol-5-en-24-oyl-glycine
3βH,7O-CA(25R/S): 3β-hydroxy-7-oxocholest-5-en-(25R/S)-oic acid
3βH,7O-Δ^5^-BA: 3β-hydroxy-7-oxochol-5-en-24-oic acid
3β-HCA: 3β-hydroxycholest-5-en-(25R)26-oic acid
3βH-Δ^5^-BA: 3βH-Δ^5^-BA
3βS,7β-GlcNAc-Δ^5^-BA: 3β-sulfooxy7β-*N*-acetylglucosaminyl-chol-5-en-24-oic acid
3βS,7β-GlcNAc-Δ^5^-BA-24-Gly: 3β-sulfooxy-7β-*N*-acetylglucosaminyl-chol-5-en-24-oyl-glycine
3βS,7β-GlcNAc-Δ^5^-BA-24-Tau: 3β-sulfooxy-7β-*N*-acetylglucosaminyl-chol-5-en-24-oyl-taurine
7-DHC: 7-dehydrocholesterol
7-OC: 7-oxocholesterol
7α-HC: 7α-hydroxycholesterol
5,6-EC: 5,6-epoxycholesterol
7α-HCO: 7α-hydroxycholest-4-en-3-one
7αH,3,24-diO-CA(25R): 7αH,3,24-diO-CA(25R)
7αH,3O-CA(25R/S): 7α-hydroxy-3-oxocholest-4-en-(25R/S)26-oic
7αH,3O-CA(25S): 7α-hydroxy-3-oxocholest-4-en-(25S)26-oic acid
7αH,3O-Δ^4^-BA: 7α-hydroxy-3-oxochol-4-en-24-oic
7αH,3O-Δ^24*E*^-CA: 7α-hydroxy-3-oxocholesta-4,24*E*-dien-26-oic
7α,12α-diHCO: 7α,12α-dihydroxycholest-4-en-3-one
7α,12α-diH,3O-CA(25S): 7α,12α-dihydroxy-3-oxocholest-4-en-(25S)26-oic acid
7α,12α-diH,3O-Δ^4^-BA: 7α,12α-dihydroxy-3-oxochol-4-en-24-oic acid
7α,24R-diH,3O-CA(25R): 7α,24R-dihydroxy-3-oxocholest-4-en-(25R)26-oic acid
7α,24S-diH,3O-CA(25S): 7α,24S-dihydroxy-3-oxocholest-4-en-(25S)26-oic acid,
7α,(25S)26-diHC: 7α,(25S)26-dihydroxycholesterol
7αH-27-*nor*-C-3,24-diO: 7α-hydroxy-27-*nor*-cholest-4-en-3,24-dione
7β-HC: 7β-hydroxycholesterol
8-DHC: 8(9)-dehydrocholesterol
24-DHC: desmosterol
25-HC: 25-hydroxycholesterol
26-HC: (25R)26-hydroxycholesterol
ABCG5: ATP-binding cassette sub-family G member 5
ABCG8: ATP-binding cassette sub-family G member 8
ACOX2: branched-chain acyl-CoA oxidase
AKR1C4: 3α-hydroxysteroid dehydrogenase, type I
AKR1D1: Δ^4^-3-oxosteroid 5β-reductase
AMACR: α-methylacyl-CoA racemase
ASMD: acid sphingomyelinase deficiency
BAAT: bile acid-CoA: amino acid *N*-acyl transferase
BACS: bile acyl-CoA synthetase
CDCA: cholesterol ester storage disease, chenodeoxycholic acid
CDPX2: X-linked dominant chondrodysplasia punctata-2
CESD: CH25H, cholesterol 25-hydroxylase
ChEH: cholesterol-5,6-oxide hydrolase
CHO: cholesterol oxidase
CTX: cerebrotendinous xanthomatosis
CYP7A1: cholesterol 7α-hydroxylase
CYP7B1: oxysterol 7α-hydroxylase
CYP8B1: sterol 12α-hydroxylase
CYP27A1: sterol 27-hydroxylase
DHCR7: 7-dehydrocholesterol reductase
DHCR24: 24-dehydrocholesterol reductase
diH-Δ^5^-BA: dihydroxychol-5-enoic acid
DI-MS: direct infusion-MS
EADSA: enzyme-assisted derivatisation for sterol analysis
EBP: emopamil-binding protein
ESI: electrospray ionisation
GC-MS: gas chromatography-MS
GCA: glycocholic acid
GlcA: glucuronic acid
GlcNAc: *N*-acetylglucosamine
GP: Girard P
HSD3B7: 3β-hydroxy-Δ^5^-C_27_-steroid oxidoreductase
HSD11B1: hydroxysteroid 11-beta dehydrogenase 1
HSD11B2: hydroxysteroid 11-beta dehydrogenase 2
LAL: lysosomal acid lipase
LALD: lysosomal acid lipase deficiency
LC-MS: liquid chromatography-MS
LIPA: lysosomal acid lipase
MEND: male EBP disorder with neurological abnormalities
LIT: linear ion trap
MRM: multiple reaction monitoring
MS: mass spectrometry
MS^2^: MS/MS, tandem MS
MS^n^: MS with multistage fragmentation
NPB: Niemann-Pick type B
NPC: Niemann-Pick type C
O7AHD: oxysterol 7α-hydroxylase deficiency
PPCS: *N*-palmitoyl-*O*-phosphocholineserine
Py: pyridine
RIC: reconstructed ion-chromatogram
SC5D: sterol C-5 desaturase
SLC27A2: long-chain fatty acid transport protein 2
SLC27A5: VLCS-homolouge2
SLOS: Smith-Lemli-Opitz syndrome
SMPD1: acid sphingomyelinase
SPC: sphingosine-1-phosphocholine
SPE: solid phase extraction
SPG5A: spastic paraplegia type 5A
SULT2A1: sulfotransferase family 2A member 1
TCA: taurocholic acid
UDCA: ursodeoxycholic acid
UGT3A1: UDP *N*-acetylglucosaminyl transferase 3A1
VLACS: very long chain acyl-CoA synthetase
VLCSH2: VLCS-homolouge2.

