## Supplemental for "Rare Cholesterol Related Disorders – A Sterolomic Library for Diagnosis and Monitoring of Diseases"

Short title: Sterolomic Library for Diagnosis and Monitoring of Diseases

**SUPPLEMENTAL EXPERIMENTAL METHODS FOR CONSTRUCTION OF THE LIBRARY**

**Enzyme-assisted derivatisation for sterol analysis - cholesterol precursors, oxysterols including bile acid precursors: Sample preparation**

**Plasma**

Plasma (100 µL) was added drop-wise under ultrasonication to 1.05 mL of absolute ethanol containing isotope labelled standard. Typically the isotope labelled standards were from Avanti Polar Lipids (now Avanti Research) and included [26,26,26,27,27,27-$^2H_6$]24R/S-hydroxycholesterol ([$^2H_6$]24R/S-HC, LM-4110), [25,26,26,26,27,27,27-$^2H_7$]7α-hydroxycholesterol ([$^2H_7$]7α-HC, LM-4103), [25,26,26,26,27,27,27-$^2H_7$]7-oxocholesterol ([$^2H_7$]7-OC, LM-4107), [26,26,26,27,27,27-$^2H_6$]7α,25-dihydroxycholesterol, [25,26,26,26,27,27,27-$^2H_7$]22R-hydroxycholest-4-en-3-one ([$^2H_7$]22R-HCO) or [25,26,26,26,27,27,27-$^2H_7$]22S-hydroxycholest-4-en-3-one ([$^2H_7$]22S-HCO), both generated from their respective 22-hydroxycholesterols (700052P, 700051P) by treatment with cholesterol oxidase from Streptomyces sp (Merck), and [25,26,26,26,27,27,27-$^2H_7$]cholesterol ([$^2H_7$]cholesterol, LM4100). In some cases [26,26,26,27,27,27-$^2H_6$]desmosterol (LM-4108), [24,24,27,27,27-$^2H_5$]3β-hydroxycholest-5-ene-(25R)26-oic acid ([$^2H_5$]3β-HCA, 700151P) and [27,27,27-$^2H_3$]7α-hydroxy-3-oxocholest-4-en-(25R/S)26-oic acid ([$^2H_3$]7αH,3O-CA(25R/S), 700194P) were added to the mixture, or the mixture was replaced by the OxysterolSPLASH™ (330700W) supplemented with [$^2H_7$]cholesterol. After 5 min sonication, 0.35 mL of water was added to give a 70% ethanol solution which was sonicated for a further 5 min then centrifuge at 14,000 g at 4°C for 30 min.

| Isotope Labelled Standards | | |
|---|---|---|
| Compound | Avanti Code | Typical Concentration ng/100 µL plasma |
| [$^2H_6$]24R/S-HC | LM-4110 | 20[a], 10[b] |
| [$^2H_7$]7α-HC | LM-4103 | 20[a], 10[b] |
| [$^2H_7$]22R-HCO | NA | 20[a] |
| [$^2H_7$]22S-HCO | NA | 5[b] |
| [$^2H_6$]7α,25-diHC | 700078P | 10[a], 4.82[b] |
| [$^2H_7$]7-OC | LM-4107 | 20[a], 10[b] |
| [$^2H_7$]Cholesterol | LM4100 | 20,000[a], 32,280[b] |
| [$^2H_5$]3β-HCA | 700151P | 10[b] |
| [$^2H_3$]7αH,3O-CA | 700194P | 10[b] |
| [$^2H_6$]Desmosterol | LM-4108 | 60[b] |

[a,b] Typical internal standard concentrations used.

A $tC_{18}$, 200 mg Sep-Pak column (Waters Inc) was washed with 4 mL of absolute ethanol and preconditioned with 6 mL of 70% ethanol. Plasma, from above, in 70% ethanol (1.5 mL) was added to the column and allowed to flow at 0.25 mL/min. The flow-through was collected and combined with column wash of 5.5 mL of 70% ethanol. Oxysterols including bile acid precursors elute in this SPE1 fraction 1 (SPE1-Fr1, see Figure 4). The column was then wash with another 4 mL of 70% ethanol, this constitutes SPE1 fraction 2 (SPE1-Fr2). Cholesterol and sterols with similar lipophilicity were eluted in 2 mL of absolute ethanol, SPE1 fraction 3 (SPE1-Fr3).

The fractions SPE1-Fr1 and SPE1-Fr3 were each divided into two equal volumes (A) and (B) and dried under vacuum. Each sub-fraction was reconstitute in 100 µL of propan-2-ol and thoroughly mixed by vortex. To fraction SPE1-Fr1A, 1 mL of 50 mM phosphate buffer ($KH_2PO_4$) pH 7 containing 3.0 µL of cholesterol oxidase from Streptomyces sp (2 mg/mL in $H_2O$, 44 units/mg of protein, Merck) was added. The reaction mixture was incubated at 37°C for 1 hr then quenched by the addition of 2 mL of methanol. Fraction SPE1-Fr3A was treated in an identical manner, while the procedure was repeated for fractions SPE1-Fr1B and SPE1-Fr3B but in the absence of cholesterol oxidase. To each fraction 150 µL of glacial acetic acid was added and thoroughly vortexed.

To the A-fractions 190 mg of [$^2H_5$]GP reagent as the bromine salt was added while 150 mg [$^2H_0$]GP as the chlorine salt (TCI Europe) was added to the B-fractions. After a thorough vortex mix, hydrazone formation was allowed to proceed overnight in the dark (see Supplemental Figure S1A).

Each of the reaction mixtures i.e. SPE1-Fr1A, SPE1-Fr1B, SPE1-Fr3A, SPE1-Fr3B was applied to an individual 60 mg Oasis HLB (Waters Inc.) column which had previously been washed with 6 mL 100% methanol followed by 6 mL 10% methanol and conditioned with 4 mL 70% methanol. The flow-through was collected and the column washed with 1 mL of 70% methanol which was combined with the flow-through to give 4.25 mL of ~70% organic solution. 4 mL of water was added to give a solution 8.25 mL of ~35% organic. The column was washed/conditioned with 1 mL of 35% methanol which was added to the previous collected eluent to give 9.25 mL of 35% organic. This solution was then re-applied to the column, collected, and diluted with 9 mL of water to give 18.25 mL of ~17.5% organic. The column was washed with 1 mL of 17.5% methanol, this was combined with the collected 18.25 mL to give 19.25 mL of 17.5% organic. The 19.25 mL was re-applied to the column and the effluent discarded. At this point derivatised oxysterols, bile acids precursors (derived from SPE1-Fr1) and cholesterol (derived from SPE1-Fr3) were bound (note SPE1-Fr1 and SPE1-Fr3 were applied to separate OASIS HLB columns). The column was washed with 6 mL 10% methanol and the wash discarded. The derivatised molecules were then eluted in 3 x 1 mL of methanol. Oxysterols and bile acid precursors elute in the first 2 mL of methanol, cholesterol and similarly lipophilic sterols elute across the 3 mL of methanol (see Figure 4, SPE2-FrA and SPE2-FrB).

**Dried blood spots**

A dried blood spot (100 µL blood) in 500 µL of absolute ethanol was sonicated for 15 min at room temperature. A further 550 µL of absolute ethanol containing isotope labelled internal standards was added under sonication and sonicated for a further 15 min. The mixture was centrifuged and the supernatant diluted with 450 µL of water and thoroughly vortexed. The resulting 1.5 mL of 70% ethanol was added to a preconditioned $tC_{18}$, 200 mg Sep-Pak column and the procedure continued as above.

**Bile acids and bile alcohols: Sample preparation**

Unconjugated bile acids elute in the oxysterol fraction from SPE2 (Figure 4). However, SPE2-FrA is contaminated with cholic acid, a component of commercial cholesterol oxidase, so this must be considered. SPE2-FrB does not contain cholesterol oxidase so this problem is avoided. SPE2-FrB is devoid of bile acids possessing an oxo group, now derivatised with GP reagent, but bile acids with a 3α- or 3β-hydroxy group are still present. However, amidated bile acids and sulfated bile acids/alcohols may be lost in the derivatisation process in which case it is preferable to prepare bile acids separately from oxysterols. This was done by dividing SPE1-Fr1 into three equal fractions A, B and $C^1$, where fraction SPE1-Fr1$C^1$ was processed in an identical manner to fraction SPE1-Fr1B (above) but in the absence of both cholesterol oxidase and GP reagent, as shown in the blue box in Figure 4. Alternatively, where bile acids and bile alcohols were of particular interest and where plasma quantities were not limited bile acids/alcohols were prepared in the dedicated protocol described below (see red box in Figure 4).

Plasma (100 µL) was added dropwise under sonication to 300 µL of methanol containing 20 µL of internal standard mix (Deuterated Bile Acids MaxSpec® Discovery Mixture, Cayman Chemical Company) and sonicated for 5 min. Water (1.6 mL) was added dropwise and the solution sonicated for a further 5 min. The solution was centrifuged for 30 min at 14,000 x g at 4$^{o}$C and the supernatant collected. The supernatant was loaded onto a 60 mg OASIS HLB column (Waters Inc) pre-washed with 4 mL of methanol and conditioned with 4 mL of 15% methanol. The column was washed with 4 mL of water and bile acids eluted in 4 x 1 mL of methanol. Primary bile acids eluted in the first 2 mL of methanol.

| Deuterated Bile Acids MaxSpec® Discovery Mixture, Cayman Chemical Company | | | |
|---|---|---|---|
| Compound | Stock Concentration (µM) | Concentration (pmol)/100 µL plasma | Concentration (ng)/100 µL plasma |
| [2,2,4,4-$^{2}H_{4}$]Cholic acid | 1.15 | 23 | 9.49 |
| [2,2,4,4-$^{2}H_{4}$]Deoxycholic acid | 1.65 | 33 | 13.09 |
| [2,2,4,4-$^{2}H_{4}$]Glycocholic acid | 1.15 | 23 | 10.08 |
| [2,2,4,4-$^{2}H_{4}$]Lithocholic acid | 0.25 | 5 | 1.90 |
| [2,2,4,4-$^{2}H_{4}$]Taurochenodeoxycholic acid | 1.00 | 20 | 10.07 |
| [2,2,4,4-$^{2}H_{4}$]Glycochenodeoxycholic acid | 3.80 | 76 | 34.48 |
| [2,2,4,4-$^{2}H_{4}$]Chenodeoxycholic acid | 1.15 | 23 | 9.12 |
| [2,2,4,4-$^{2}H_{4}$]Ursodeoxycholic acid | 0.30 | 6 | 2.38 |
| [2,2,4,4-$^{2}H_{4}$]Taurocholic acid | 1.00 | 20 | 10.39 |
| [2,2,4,4-$^{2}H_{4}$]Glycodeoxycholic acid | 1.28 | 25.6 | 11.61 |
| [2,2,4,4-$^{2}H_{4}$]Glycolithocholic acid | 0.25 | 5 | 2.19 |
| [2,2,4,4-$^{2}H_{4}$]Glycoursodeoxycholic acid | 0.60 | 12 | 5.44 |
| [2,2,4,4-$^{2}H_{4}$]Taurodeoxycholic acid | 1.00 | 20 | 10.07 |
| [2,2,4,4-$^{2}H_{4}$]Tauroursodeoxycholic acid | 0.5 | 10 | 5.04 |
| [2,2,4,4-$^{2}H_{4}$]Taurolithocholic acid | 0.25 | 5 | 2.44 |

**LC-MS(MS$^{n}$) and LC-MS(MS/MS)**

**Oxysterols and bile acid precursors - 17 min gradient**

Samples in 60% methanol were injected onto a reversed-phase Hypersil Gold $C_{18}$ column (1.9 µm particle size, 50 x 2.1 mm, Thermo Fisher) using an Ultimate 3000 LC system (Thermo Fisher Scientific). Mobile phase composition was initially 80% A (33.3% methanol, 16.7% acetonitrile, 0.1% formic acid), 20% B (63.3% methanol, 31.7% acetonitrile, 0.1% formic acid) and held for 1 min, changed over 7 min to 20% A, 80% B and maintained for 4 min. The mobile phase was returned to 80% A, 20% B in 6 s and the column reconditioned for a further 4 min 54 s. The flow-rate was 200 µL/min and the eluate introduced by ESI to an Orbitrap mass spectrometer (Thermo Fisher Scientific).

**Oxysterols, bile acid precursors, bile acids and bile alcohols - 37 min gradient**

Samples in 60% methanol were injected onto a reversed-phase Hypersil Gold $C_{18}$ column (1.9 µm particle size, 50 x 2.1 mm, Thermo Fisher) using an Ultimate 3000 LC system (Thermo Fisher Scientific). The mobile phase composition initially at 80% A (33.3% methanol, 16.7% acetonitrile, 0.1% formic acid) and 20% B (63.3% methanol, 31.7% acetonitrile, 0.1% formic acid) was held for 10 min then changed over 10 min to 50% A, 50% B, and maintained for 6 min. The proportion of B increased to 80% over 3 min and maintained for a further 3 min. The mobile phase was returned to 80% A, 20% B in 6 s and the column reconditioned for a further 4 min 54 s. The flow-rate was 200 µL/min and the eluate introduced by ESI to an Orbitrap mass spectrometer (Thermo Fisher Scientific).

**Sterols - 24 min gradient**

Samples in 75% methanol were injected onto a reversed-phase ACE $C_{18}$ column (10 cm x 2.1 mm, 2 µm, Advanced Chromatography Technologies Ltd) using an Ultimate 3000 LC system (Thermo Fisher Scientific). The mobile phase composition was initially 22% A (33.3% methanol, 16.7% acetonitrile, 0.1% formic acid) and 78% B (63.3% methanol, 31.7% acetonitrile, 0.1% formic acid) and changed over 8 min to 20% A, 80% B, then changed to 15% A, 85 %B over 3 min and held at this composition for a further 6 min. The mobile phase was returned to 22% A, 78% B in 6 s and the column reconditioned for a further 6 min 54 s. The flow-rate was 200 µL/min and the eluate introduced by ESI to an Orbitrap mass spectrometer (Thermo Fisher Scientific).

**Positive-ion MS($MS^3$)**

Mass spectra in the positive-ion mode were typically recorded at 120,000 or 240,000 resolution (at *m/z* 400) over the *m/z* range 400 – 610 or 300 – 800. Using the Orbitrap Elite $MS^3$ spectra were recorded for the transitions $[M]^+ \rightarrow [M-Py]^+ \rightarrow$, where $[M]^+$ is the GP-derivatised molecule and $[M-Py]^+$ its fragment ion where pyridine has been lost (see Supplemental Figure S1C & D). Fragmentation was performed in the linear ion trap (LIT) with the isolation window set at 1 and at collision energies of 30% and 35% with ion detection at the ion-trap detector. Similarly, $MS^3$ spectra were recorded on the Orbitrap IDX and IQX but with precursor ion selection window of the quadrupole mass filter at 1.6 u and the isolation window of the LIT at 2 u. MRM-like reconstructed-ion chromatograms (RICs) were recorded with an $MS^3$ fragment-ion *m/z* window of ± 0.35.

**Negative-ion MS(MS/MS)**

Mass spectra were recorded on the Orbitrap IDX and IQX instruments at 120,000 resolution (at *m/z* 400) typically over the *m/z* ranges 370 – 800 or 270 – 800. MS/MS spectra were recorded with precursor ion selection in the quadrupole mass filter with isolation width set at 0.8 u, fragmentation in the ion routing multipole, and *m/z* recorded in the Orbitrap at 7,500 resolution (at *m/z* 400). Collision voltage was at 50 V, 60 V or 80 V depending on the precursor ion (see below). Typically, 4 – 6 MS/MS scan events were included per injections with multiple injections for each sample. This allowed the fragmentation of a wide range of precursor *m/z* without prescription of retention time.

| Bile Acid/Alcohol | Target Collision V |
|---|---|
| Non-amidated bile acid | 50 or 60 |
| Glycine conjugated bile acid | 60 |
| Taurine conjugated bile acid | 80 |
| Sulfate conjugated bile acid | 80 |
| N-Acetylglucosamine conjugated bile acid | 50, 60[a], 80[b] |
| Glucuronidated bile alcohol | 50 |

[a] If doubly conjugated with glycine.

[b] If doubly or triply conjugated with taurine and/or sulfuric acid.

**SUPPLEMENTAL TABLE CAPTIONS**

**Supplemental Table S1.** Cholesterol Related Disorders, Dysregulated Metabolites and Mass Spectrometry Analysis.

**Supplemental Table S2.** Cholesterol Related Disorders, Dysregulated Metabolites and Simplest Mass Spectrometry Analysis.

**SUPPLEMENTAL FIGURE CAPTIONS**

**Figure S1.** Enzyme-assisted derivatisation for sterol analysis (EADSA). (A) Fractions-A are treated with cholesterol oxidase followed by [$^2H_5$]GP, this results in derivatisation of native 3-oxosterols and those enzymatically formed *ex vivo* from 3β-hydroxy substrates by cholesterol oxidase. (B) Fractions-B are treated with [$^2H_0$]GP in the absence of cholesterol oxidase, resulting in the derivatisation of only native oxosterols. $MS^n$ fragmentation of (C) [$^2H_5$]GP and (D) [$^2H_0$]GP derivatised sterols.

**Figure S2.** Side-chain shortening starting with formation of a CoA thioester by very long-chain acyl-CoA synthetase (VLACS) also known as solute carrier family 27 member 2 (SLC27A2) or by bile acyl-CoA ligase (SLC27A5). Metabolites are coloured as in Figure 2. Shown in the dashed-box is a pathway starting with cerebral 24S-hydroxycholesterol. Abbreviations and disorders are as in Figure 2 or defined below: AMACR, α-methylacyl-CoA racemase, (AMACR deficiency); ACOX2, branched-chain acyl-CoA oxidase, (ACOX2 deficiency); DBP, D-bifunctional protein, (DBP deficiency); SCP2, sterol carrier protein-2, (leukoencephalopathy with dystonia and motor neuropathy); CYP39A1, cytochrome P450 family 39 subfamily A member 1, (24S-hydroxycholesterol 7α-hydroxylase deficiency). Other enzymes: LBP, L-bifunctional protein.

**Figure S3.** EADSA-LC-MS($MS^n$) analysis of plasma samples. LC-MS RICs (± 5 ppm) of plasma showing cholesterol and its isomer lathosterol from (A) a patient suffering from mild lathosterolosis (upper panel) and of NIST SRM 1950 doped with lathosterol (lower panel). Note cholesterol was derivatised with [$^2H_5$]GP and lathosterol dopant with [$^2H_0$]GP. $MS^3$ ($[M]^+\rightarrow[M\text{-}Py]^+\rightarrow$) spectra of (B) cholesterol in plasma and (C) lathosterol doped into plasma. Fragment ion nomenclature is described in (1). (D) RICs of plasma from a lathosterolosis patient (upper panel) and of NIST SRM 1950 (lower panel) showing dehydrocholesterol (DHC) isomers. Representative $MS^3$ spectra of desmosterol, 7-DHC and 8-DHC are shown in Figure 5. Data was acquired on an Orbitrap IQX using an Ace $C_{18}$ column. Note, there is some difference in retention time between chromatograms which were recorded on different days and in some cases using different columns although of the same type.

**Figure S4.** Acidic pathway and CTX shunt pathway. Colouring as in Figure 2. Where enzymes are suggested rather than known the reactions are indicated by broken arrows.

**Figure S5.** Analysis of CTX (upper panels) and NIST SRM1950 (lower panels) plasma by EADSA-LC-MS($MS^n$). (A) LC-MS RIC of *m/z* 564.3796 ± 5 ppm corresponding to 7αH,3O-CA(25R/S). The upper and lower panels are plotted on the same y-axis, but the peaks in the upper panel are magnified by a factor of 10. (B) $MS^3$ ($[M]^+\rightarrow[M\text{-}Py]^+\rightarrow$) spectra of 7αH,3O-CA(25S) from the CTX (upper panel) and NIST (lower panel) plasma samples. (C) $MS^3$ ($[M]^+\rightarrow[M\text{-}Py]^+\rightarrow$) spectra of 7αH,3O-CA(25R) from the CTX (upper panel) and NIST (lower panel) plasma samples. Data acquired on the Orbitrap Elite using a Hypersil Gold $C_{18}$ column. See Supplemental S4L in reference (1) for an interpretation of fragmentation patterns. Data for NIST SRM 1950 plasma has been previously reported in reference (1).

**Figure S6**. Identification of CTX from a dried blood spot. After extraction into ethanol and derivatisation with [$^2H_0$]GP, LC-MS(MS$^n$) analysis reveals high concentrations of 7α,12α-diHCO in CTX. (A) MRM-like chromatogram (550.4→471.4→392.3) targeting 7α,12α-diHCO. For comparative data from CTX plasma, see Figure 6D. (B) MS$^3$ spectra of 7α,12α-diHCO from the dried blood spot (upper panel) and from the CTX plasma sample (lower panel). Data acquired on the Orbitrap IDX using a Hypersil Gold $C_{18}$ column. Interpretation of MS$^3$ spectra can be found in Supplemental Figure S4K in reference (1). See Supplemental information for full details of experimental method. CTX patients were previously studies in references (2, 3).

**Figure S7**. Analysis of a plasma sample from a CTX patient treated with CDCA and of the NIST SRM1950 plasma by negative-ion LC-MS(MS/MS) following sample preparation via a single SPE step (see Figure 4, red box, Fr1C$^2$). In chromatograms (A), (C) and (G) data from CTX plasma is shown in the upper panel and from NIST SRM 1950 plasma in the lower panel. (A) LC-MS RICs of *m/z* 611.3801 ± 5 ppm. (B) MS/MS spectra of the two most abundant cholestanetetrol glucuronides in the CTX sample. (C) LC-MS RIC of *m/z* 627.3750 ± 5 ppm. (D – F) MS/MS spectra of the five most abundant cholestanepentol glucuronides in the CTX sample. (G) LC-MS RICs of *m/z* 643.3699 ± 5 ppm. (H) MS/MS spectra of the two most abundant cholestanehexol glucuronides in the CTX sample. Data acquired on the Orbitrap IQX using a Hypersil Gold $C_{18}$ column.

**Figure S8**. O7AHD / SPG5A shunt pathway. Cholesterol metabolites are coloured as in earlier Figures.

**Figure S9**. Analysis of SPG5A (upper panels) and control (lower panels) plasma by negative-ion LC-MS(MS/MS) without enzyme or GP-treatment. The upper and lower chromatograms are plotted on the same y-axis. LC-MS RICs of (A) *m/z* 453.2316 ± 5 ppm, (C) *m/z* 480.2789 ± 5 ppm and (D) 510.2531 ± 5 ppm. LC-MS/MS chromatograms selecting $[M-H]^-$ ions at (B) *m/z* 453.2, and (E) *m/z* 510.3 as the precursor ion, and *m/z* 96.9601 ± 20 ppm, as the target product ion. (F) MS/MS spectrum of *m/z* 510.3. Data acquired on the Orbitrap IQX using a Hypersil Gold $C_{18}$ column. Some chromatograms have been re-aligned to correct for retention-time drift.

**Figure S10**. Analysis of plasma from a patient with apparent HSD3B7 deficiency. EADSA-LC-MS RICs corresponding to 3β,7α-diHCA(25R/S) plus 7αH,3O-CA(25R/S) labelled 3β,7α-diHCA(25R/S)$^‡$, upper panel) and 7αH,3O-CA(25R/S) alone (lower panel) from plasma from (A) a patient with apparent HSD3B7 deficiency and (B) NIST SRM 1950. The y-axis in the upper and lower panels are plotted on the same scale. Note Fractions-A and -B (Figure 4) were both derivatised with [$^2H_0$]GP in this patient sample and analysed in separate LC-MS runs. Data acquired on the Orbitrap Elite using a Hypersil Gold $C_{18}$ column. Chromatograms have not been realigned to correct for retention time drift.

**Supplemental Figure S11.** Analysis of plasma from a patient with apparent HSD3B7 deficiency. (A) Negative-ion DI-MS prepared by a single SPE step without enzyme or GP-treatment (see Figure 4). (B) The *m/z* range 275-350 shown on an expanded x-axis. The second isotopes of doubly charged ions are indicated by blue arrows. The identity of the labelled peaks is given in Supplemental Table S1. (C) MS$^2$ of doubly charged ions at *m/z* 280.12 and 296.12 corresponding to doubly sulfated monohydroxycholesterol and trihydroxycholesterol, respectively. (D) MS$^2$ of doubly charged ions at *m/z* 328.16 and 336.16 corresponding to mono- and di-hydroxycholesterols doubly conjugated with sulfuric and glucuronic acids. Data acquired on an Orbitrap XL. $MS^2$ spectra were recorded in the linear ion-tap. One sulfate group is probably at C-3 and the location of the second sulfate or glucuronic acid group is unknown. However, previous analysis of plasma from a patient with HSD3B7 deficiency suggests the site of the second conjugating group is C-7 (4). (E) Probable structures of sulfate conjugates. (F) Probable structures of sugar conjugates. Some of the data for this patient has previously been reported in (5).

**Supplemental Figure S12.** EASDA-LC-MS($MS^n$) analysis of a plasma from a patient with apparent $\Delta^4$-3-oxosteroid-5β-reductase deficiency, likely to be a secondary effect (upper panels) and NIST SRM 1950 (lower panels) plasma. The upper and lower chromatograms are plotted on the same y-axis. LC-MS RICs of (A) *m/z* 564.3796 ± 5 ppm; (B) *m/z* 534.3690 ± 5 ppm; (C) 522.3326 ± 5 ppm; and (D) 550.4003. $MS^3$ spectra of (E) 7αH,3O-CA(25R); (F) 7αH-27-*nor*-C-3,24-diO; and (G) 7αH,3O-$\Delta^5$-BA, from the patient plasma. Data acquired on the Orbitrap IQX using a Hypersil Gold $C_{18}$ column.

**Supplemental Figures S13.** EADSA-LC-MS($MS^n$) analysis of plasma from the *AMACR*$^{-/-}$ mouse (upper panel) and wild type control (lower panel). LC-MS RICs corresponding to (A) *m/z* 564.3796 ± 5 ppm, 7αH,3O-CA(25R/S) and (B) 580.3745 ± 5 ppm, 7α,12α-diH,3O-CA(25R). The y-axis in the upper and lower panels are plotted on the same scale, but peaks are magnified by a factor of 10 in the lower panels. $MS^3$ spectra of (C) 7αH,3O-CA(25R) and (D) 7α,12α-diH,3O-CA(25R) from the *AMACR*$^{-/-}$ mouse. Data acquired on the Orbitrap Velos using a Hypersil Gold $C_{18}$ column. Data has been reported previously in (6). For fragmentation nomenclature see Supplemental Figure S4 in reference (1).

**Supplemental Figure S14.** EADSA- LC-MS($MS^3$) analysis of 25S and 25R cholestenoic acids in ACOX2 plasma. (A) $MS^3$ spectra of 3β,7β-diHCA(25S) (upper panel) and 3β,7β-diHCA(25R) (lower panel). (B) $MS^3$ spectra of 7αH,3O-CA(25S) (upper panel) and 7αH,3O-CA(25R) (lower panel). (C) $MS^3$ spectra of 7α,12α-diH,3O-CA(25S) (upper panel) and 7α,12α-diH,3O-CA(25R) (lower panel). Data acquired on the Orbitrap Elite using a Hypersil Gold $C_{18}$ column. The spectra are from the chromatograms shown in Figure 11.

**Supplemental Figure S15.** Analysis by negative-ion LC-MS(MS/MS) of ACOX2 (upper panels) and NIST SRM 1950 (lower panels) plasma, prepared without enzyme or GP-treatment. The upper and lower panels are plotted on the same y-axis. LC-MS RICs of $[M-H]^-$ ions (A) *m/z* 429.3010 ± 5 ppm corresponding to 7αH,3O-CA(25R/S); (B) *m/z* 431.3167 corresponding to 3β,7α-diHCA(25R/S); (C) *m/z* 433.3323 ± 5 ppm corresponding to 3α,7α-diHCa(25R/S); (D) *m/z* 449.3272 ± 5 ppm corresponding to 3α,7α,12α-triHCa(25R/S); (E) *m/z* 490.3538 ± 5 ppm corresponding to 3α,7α-diHCa(25R/S)-Gly; (F) *m/z* 506.3487 ± 5 ppm corresponding to 3α,7α,12α-triHCa(25R/S)-Gly; (G) *m/z* 540.3364 ± 5 ppm corresponding to 3α,7α-diHCa(25R/S)-Tau; (H) *m/z* 556.3313 ± 5 ppm corresponding to 3α,7α,12α-triHCa(25R/S)-Tau; (I) *m/z* 572.3263 ± 5 ppm corresponding to tetraHCa(25R/S)-Tau of probable structure 3α,7α,12α,24- tetraHCa(25R/S)-Tau. Data acquired on the Orbitrap IQX using a Hypersil Gold $C_{18}$ column. Note, authentic standards were not available to confirm the identifications made in (C), (E), (F), (G), (H) and (I) which were made by accurate mass, retention time and MS/MS spectra (see Supplemental Figure S16), where signal intensity was sufficient.

**Supplemental Figure S16.** LC-MS(MS/MS) analysis of 25R and 25S cholestenoic and cholestanoic acids in ACOX2 plasma. Upper chromatograms show ACOX2 plasma, and lower panels NIST SRM 1950. Plasma was prepared in the absence of cholesterol oxidase or GP treatment. MRM-like chromatograms

$[M\text{-}H]^- \rightarrow [M\text{-}H\text{-}H_2O]^-$, (A) *m/z* 429.3→411.2905 ± 20 ppm corresponding to 7αH,3O-CA(25R/S); (C) *m/z* 433.3→415.3218 ± 20 ppm corresponding to 3α,7α-diHCa(25R/S); (E) *m/z* 449.3→431.3167 ± 20 ppm corresponding to 3α,7α,12α-triHCa(25R/S); and $[M\text{-}H]^- \rightarrow SO_3^-$ (H) *m/z* 556.3→79.9574 ± 20 ppm corresponding to 3α,7α,12α-triHCa(25R/S)-Tau. MS/MS spectra from ACOX2 plasma of (B) *m/z* 429.3 at 21.0 and 21.6 min corresponding to 7αH,3O-CA(25S) and 7αH,3O-CA(25R), respectively; (D) *m/z* 433.3 at 29.3 and 30.0 min, corresponding to 3α,7α-diHCa(25S) and 3α,7α-diHCa(25R), respectively; (F) *m/z* 449.3 at 22.1 and 22.6 min, corresponding to 3α,7α,12α-triHCa(25S) and 3α,7α,12α-triHCa(25R), respectively; (G) *m/z* 449.3 from ACOX2 plasma and an authentic standard of 3α,7α,12α-triHCa(25S), respectively; (I) *m/z* 556.3 at 12.4 and 14.0 min corresponding to 3α,7α,12α-triHCa(25S)-Tau and 3α,7α,12α-triHCa(25R)-Tau, respectively. Data acquired on the Orbitrap IQX using a Hypersil Gold $C_{18}$ column. Note, authentic standards were not available to confirm the identifications of 3α,7α-diHCa(25R/S), 3α,7α-diHCa(25R/S)-Gly, 3α,7α,12α-triHCa(25R/S)-Gly, 3α,7α-diHCa(25R/S)-Tau and 3α,7α,12α-triHCa(25R/S)-Tau which were made by accurate mass, retention time and MS/MS spectra.

**Supplemental Figure S17.** Analysis of Wolman (upper panels) and NIST SRM 1950 (lower panels) plasma by EADSA-LC-MS(MS)$^n$. The upper and lower panels are plotted on the same y-axis. (A) LC-MS RICs of *m/z* 539.4368 ± 5 ppm corresponding to monohydroxycholesterols including 7β-HC and dehydrated 3β,5α,6β-triol (3β,5α,6β-triol*). (B) LC-MS RICs of *m/z* 534.4054 ± 5 ppm corresponding to monohydroxycholestenones including 7-OC. (C) LC-MS RICs of *m/z* 569.4110 ± 5 ppm corresponding to isomers of 3β,7β-diHCA and 3β,7α-diHCA. (D) MRM-like chromatograms *m/z* 569.4→485.3→413.3 targeting dehydrated 3β,5α,6β-triHCa (3β,5α,6β-triHCa*). The fragment ion at *m/z* 413.3 is enriched but not unique to the $MS^3$ spectrum of dehydrated 3β,5α,6β-triHCa (7). (E) MRM-like chromatograms *m/z* 564.4→485.3→426.3 targeting 3βH,7O-CA(25R/S) (7). (F) LC-MS RICs of *m/z* 527.3640 ± 5 ppm corresponding to isomers of 3β,7β-diH-$\Delta^5$-BA, 3β,7α-diH-$\Delta^5$-BA and dehydrated 3β,5α,6β-triHBA (3β,5α,6β-triHBA*). (G) LC-MS RICs of *m/z* 522.3326 ± 5 ppm corresponding to hydroxyoxocholenoic acids. Data acquired on the Orbitrap Elite using a Hypersil Gold $C_{18}$ column. The symbol (‡) indicates that 3β-hydroxy-5-ene and 3-oxo-4-ene metabolites are measured in combination. The symbol (*) indicates the dehydrated molecule.

**Supplemental Figure S18.** LC-MS(MS/MS) analysis, without enzyme or GP-treatment, of plasma from NPC and Wolman patient samples. MS/MS ($[M\text{-}H]^- \rightarrow$) spectra of (A) 3β,5α,6β-triHBA-24-Gly; (B) GCA; (C) 3β,5α,6β-triHBA-24-Tau and (D) TCA from an NPC sample, chromatograms illustrated in Figure 13. MS/MS ($[M\text{-}H]^- \rightarrow$) spectra of (E) 3β,5α,6β-triHBA; (F) cholic acid; (G) 3β,5α,6β-triHBA-24-Gly; (H) GCA; (I) 3β,5α,6β-triHBA-24-Tau; (J) TCA from a Wolman patient, chromatograms illustrated in Figure S19. Upper panels are spectra from patient samples, lower panels are from authentic standards or NIST SRM 1950 as indicated. Retention times have been aligned to correct for retention time drift. Data acquired on the Orbitrap IQX using a Hypersil Gold $C_{18}$ column

**Supplemental Figure S19.** Analysis of Wolman disease (upper panels) and NIST SRM 1950 (lower panels) plasma by negative-ion LC-MS(MS/MS) without enzyme or GP-treatment. The upper and lower panels are plotted on the same y-axis. LC-MS RICs of (A) *m/z* 407.2803 ± 5 ppm corresponding to trihydroxycholanoic acids including 3β,5α,6β-triHBA and cholic acid; (B) *m/z* 464.3018 ± 5 ppm corresponding to isomers of trihydroxycholanoyl-glycine including 3β,5α,6β-triHBA-24-Gly and GCA. (C) MRM-like transition $[M\text{-}H]^- \rightarrow$74.0248 ± 20 ppm giving the fragment ion $NH_2CH_2CO_2^-$ from trihydroxycholanoyl glycine isomers. (D) RICs of *m/z* 514.2844 ± 5 ppm corresponding to isomers of trihydroxycholanoyl-taurine including 3β,5α,6β-triHBA-24-Tau and TCA. (E) MRM-like transition $[M\text{-}H]^-$

$\rightarrow$79.9574 ± 20 ppm giving the fragment ion $SO_3^-$ from trihydroxycholanoyl taurine isomers. Chromatograms have been aligned to correct for retention time drift. Fragmentation spectra can be found in Supplemental Figure S18. The patient had been treated with recombinant LAL. Data acquired on the Orbitrap IQX using a Hypersil Gold $C_{18}$ column

**Supplemental Figure S20.** Analysis of ASMD (NPB, upper panels) and NIST SRM 1950 (lower panels) plasma by negative-ion LC-MS(MS/MS) prepared without enzyme or GP-treatment. The upper and lower panels are plotted on the same y-axis. LC-MS RICs of (A) *m/z* 407.2803 ± 5 ppm corresponding to trihydroxycholanoic acids including 3β,5α,6β-triHBA and cholic acid; (B) *m/z* 464.3018 ± 5 ppm corresponding to isomers of trihydroxycholanoyl-glycine including 3β,5α,6β-triHBA-24-Gly and GCA. (C) MRM-like transition $[M-H]^- \rightarrow$74.0248 ± 20 ppm giving the fragment ion $NH_2CH_2CO_2^-$ from trihydroxycholanoyl-glycine isomers. (D) RICs of *m/z* 514.2844 ± 5 ppm corresponding to isomers of trihydroxycholanoyl-taurine including 3β,5α,6β-triHBA-24-Tau and TCA. (E) MRM-like transition $[M-H]^- \rightarrow$79.9574 ± 20 ppm giving the fragment ion $SO_3^-$ from trihydroxycholanoyl-taurine isomers. Data acquired on the Orbitrap IQX using a Hypersil Gold $C_{18}$ column.

**Supplemental Figure S21.** LC-MS(MS/MS) analysis of plasma from patients with lysosomal storage disorders. MS/MS ($[M-H]^- \rightarrow$) spectra from ASMD and Wolman patients and where available of authentic standards. (A) (i & ii) 3βH,7β-GlcNAc-$\Delta^5$-BA, (iii) the isomer 3β-glucosyl-chol-5-en-24-oyl-glycine (3βGlc-$\Delta^5$-BA-24-Gly) and (iv) the authentic standard 3αH,7β-GlcNAc-BA. (B) (i & ii) 3βS,7β-GlcNAc-$\Delta^5$-BA, (iii & iv) the isomers of probable structures 3β-glucuronyl-7-hydroxychol-5-en-24-oyl-taurine (3βGlcA,7H-$\Delta^5$-BA-24-Tau), (v) the authentic standard 3βS,7β-GlcNAc-$\Delta^5$-BA; (C) 3βH,7β-GlcNAc-$\Delta^5$-BA-24-Tau; (D) (i & ii) Singly and (iii) doubly charged 3βS,7β-GlcNAc-$\Delta^5$-BA-24-Gly and (iv) authentic standard 3βS,7β-GlcNAc-$\Delta^5$-BA-24-Gly. (E) (i &ii) Doubly charged 3βS,7β-GlcNAc-$\Delta^5$-BA-24-Tau. The identifications of 3βGlc-$\Delta^5$-BA-24-Gly and 3βGlcA,7βH-$\Delta^5$-BA-24-Tau are based on accurate mass and fragmentation patterns, but other isomers are possible. Data was acquired on an Orbitrap IQX using a Hypersil Gold $C_{18}$ column. Relevant chromatograms can be found in Figure 14 and Supplemental Figure S22. Further information on fragmentation patterns can be found in (8).

**Figure S22.** Analysis of Wolman patient with the CH25H deletion post stem cell transplant (upper panels) and control QC (lower panels) plasma by negative-ion LC-MS(MS/MS) without prior enzyme or GP-treatment. The upper and lower panels are plotted on the same y-axis. (A) LC-MS RICs of *m/z* 592.3491 ± 5 ppm corresponding to 3βH,7β-GlcNAc-$\Delta^5$-BA and its isomer 3βGlc-$\Delta^5$-BA-24-Gly; (B) MRM-like transition $[M-H]^- \rightarrow [M-H-203.0794]^-$ ± 20 ppm corresponding to the loss of the GlcNAc conjugating group from 3βH,7β-GlcNAc-$\Delta^5$-BA; (C) LC-MS RIC of *m/z* 649.3706 ± 5 ppm corresponding to 3βH,7β-GlcNAc-$\Delta^5$-BA-24-Gly; (D) LC-MS RIC of *m/z* 672.3059 ± 5 ppm corresponding to 3βS,7β-GlcNAc-$\Delta^5$-BA and 3β-GlcA,7H-$\Delta^5$-BA-24-Tau; (E) MRM-like transition $[M-H]^- \rightarrow$96.9601 ± 20 ppm giving the $HSO_4^-$ ion from 3βS,7β-GlcNAc-$\Delta^5$-BA; (F) LC-MS RICs of *m/z* 335.6493 ± 5 ppm corresponding to doubly charged 3β-GlcA,7βH-$\Delta^5$-BA-24-Tau; (G) MRM-like transition $[M-2H]^{2-} \rightarrow$79.9574 ± 20 ppm giving the $SO_3^-$ ion from 3β-GlcA,7βH-$\Delta^5$-BA-24-Tau; (H) LC-MS RICs of *m/z* 699.3532 ± 5 ppm corresponding to 3βH,7β-GlcNAc-$\Delta^5$-BA-24-Tau; (I) MRM-like transition $[M-H]^- \rightarrow$79.9574 ± 20 ppm giving the $SO_3^-$ ion from 3βH,7β-GlcNAc-$\Delta^5$-BA-24-Tau; (J) LC-MS RICs of *m/z* 729.3274 ± 5 ppm corresponding to 3βS,7β-GlcNAc-$\Delta^5$-BA-24-Gly; (K) MRM-like transition $[M-H]^- \rightarrow$96.9601 ± 20 ppm giving the $HSO_4^-$ ion from 3βS,7β-GlcNAc-$\Delta^5$-BA-24-Gly. (L) LC-MS RICs of *m/z* 364.1601 ± 5 ppm corresponding to doubly charged 3βS,7β-GlcNAc-$\Delta^5$-BA-24-Gly. (M) LC-MS RIC of *m/z* 779.3100 ± 5 ppm corresponding to 3βS,7β-GlcNAc-$\Delta^5$-BA-24-Tau. (N) LC-MS RICs of *m/z* 389.1514 ± 5 ppm corresponding to doubly charged 3βS,7β-GlcNAc-$\Delta^5$-BA-24-Tau. (O) MRM-like transition $[M-H]^-$ →96.9601 ± 20 ppm giving the $HSO_4^-$ ion from 3βS,7β-GlcNAc-$\Delta^5$-BA-24-Tau. Fragmentation spectra can be found in Supplemental Figure S21. The identifications of 3βGlc-$\Delta^5$-BA-24-Gly, and 3βGlcA,7βH-$\Delta^5$-BA-24-Tau and 3βGlcA,7βH-$\Delta^5$-BA-24-Tau are based on accurate mass and fragmentation patterns, but other isomers are possible. Data was acquired on an Orbitrap IQX using a Hypersil Gold $C_{18}$ column.

# S1A-B

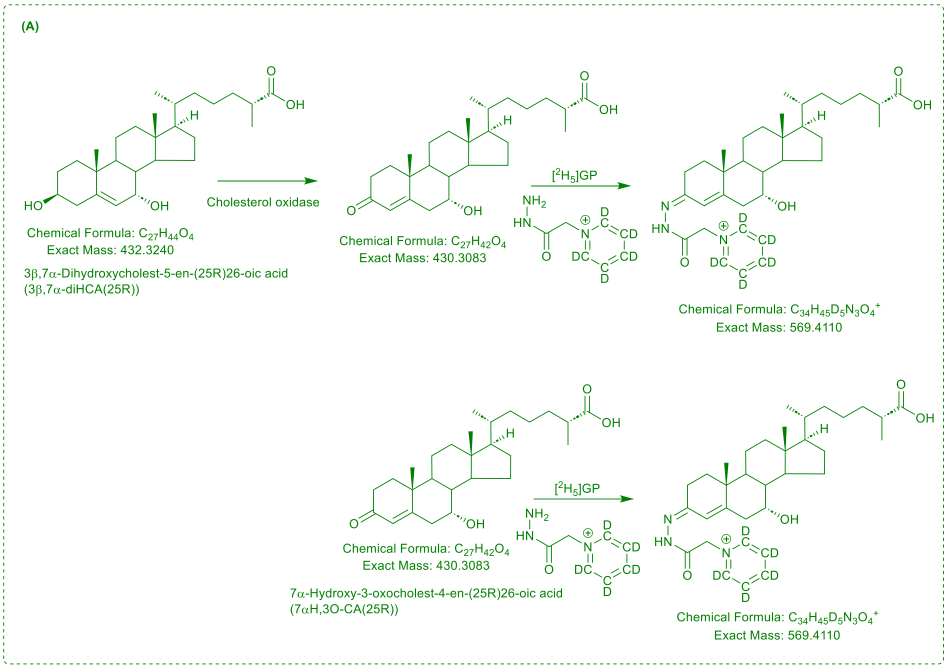
(A)
Cholesterol oxidase
Chemical Formula: C27H44O4
Exact Mass: 432.3240
3β,7α-Dihydroxycholest-5-en-(25R)26-oic acid
(3β,7α-diHCA(25R))
Chemical Formula: C27H42O4
Exact Mass: 430.3083
[2H5]GP
Chemical Formula: C34H45D5N3O4+
Exact Mass: 569.4110
Chemical Formula: C27H42O4
Exact Mass: 430.3083
7α-Hydroxy-3-oxocholest-4-en-(25R)26-oic acid
(7αH,3O-CA(25R))
[2H5]GP
Chemical Formula: C34H45D5N3O4+
Exact Mass: 569.4110

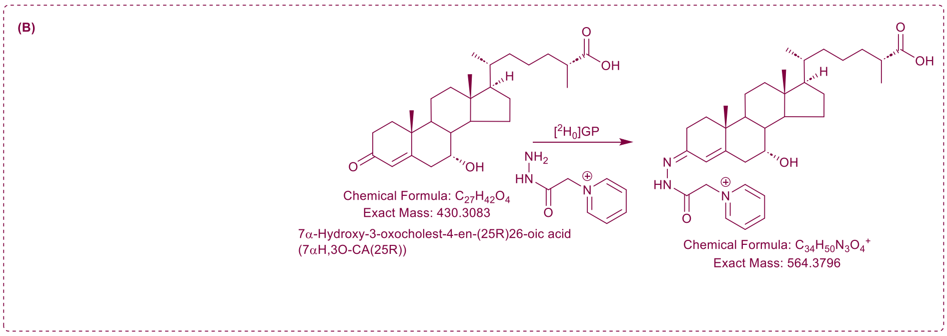
(B)
[2H0]GP
Chemical Formula: C27H42O4
Exact Mass: 430.3083
7α-Hydroxy-3-oxocholest-4-en-(25R)26-oic acid
(7αH,3O-CA(25R))
Chemical Formula: C34H50N3O4+
Exact Mass: 564.3796

# S1C-D

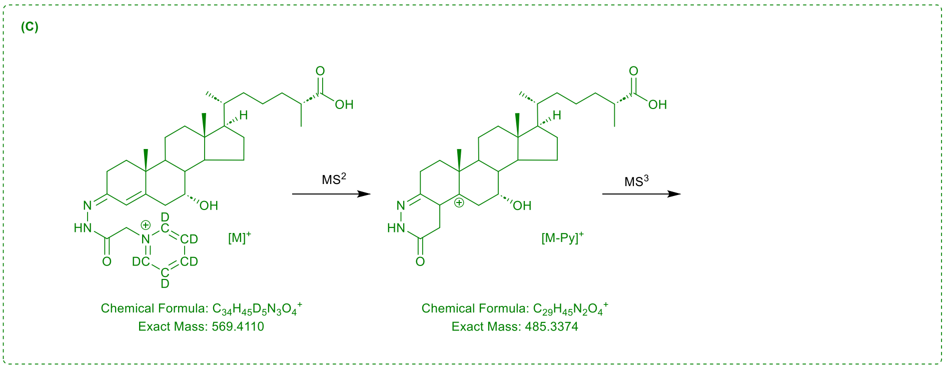
(C)
MS2
MS3
[M]+
[M-Py]+
Chemical Formula: C34H45D5N3O4+
Exact Mass: 569.4110
Chemical Formula: C29H45N2O4+
Exact Mass: 485.3374

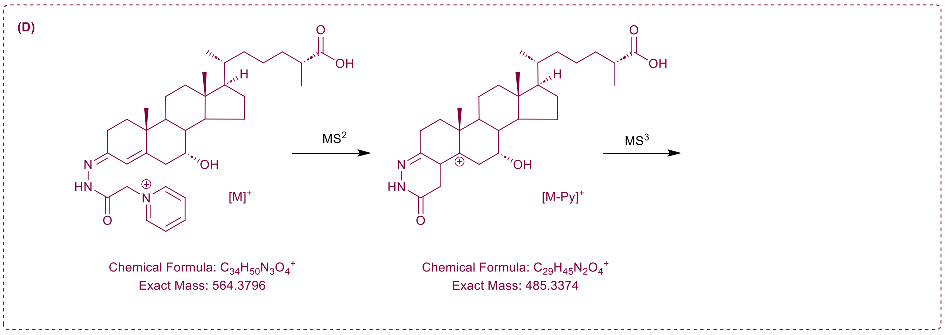
(D)
MS2
MS3
[M]+
[M-Py]+
Chemical Formula: C34H50N3O4+
Exact Mass: 564.3796
Chemical Formula: C29H45N2O4+
Exact Mass: 485.3374

# S2

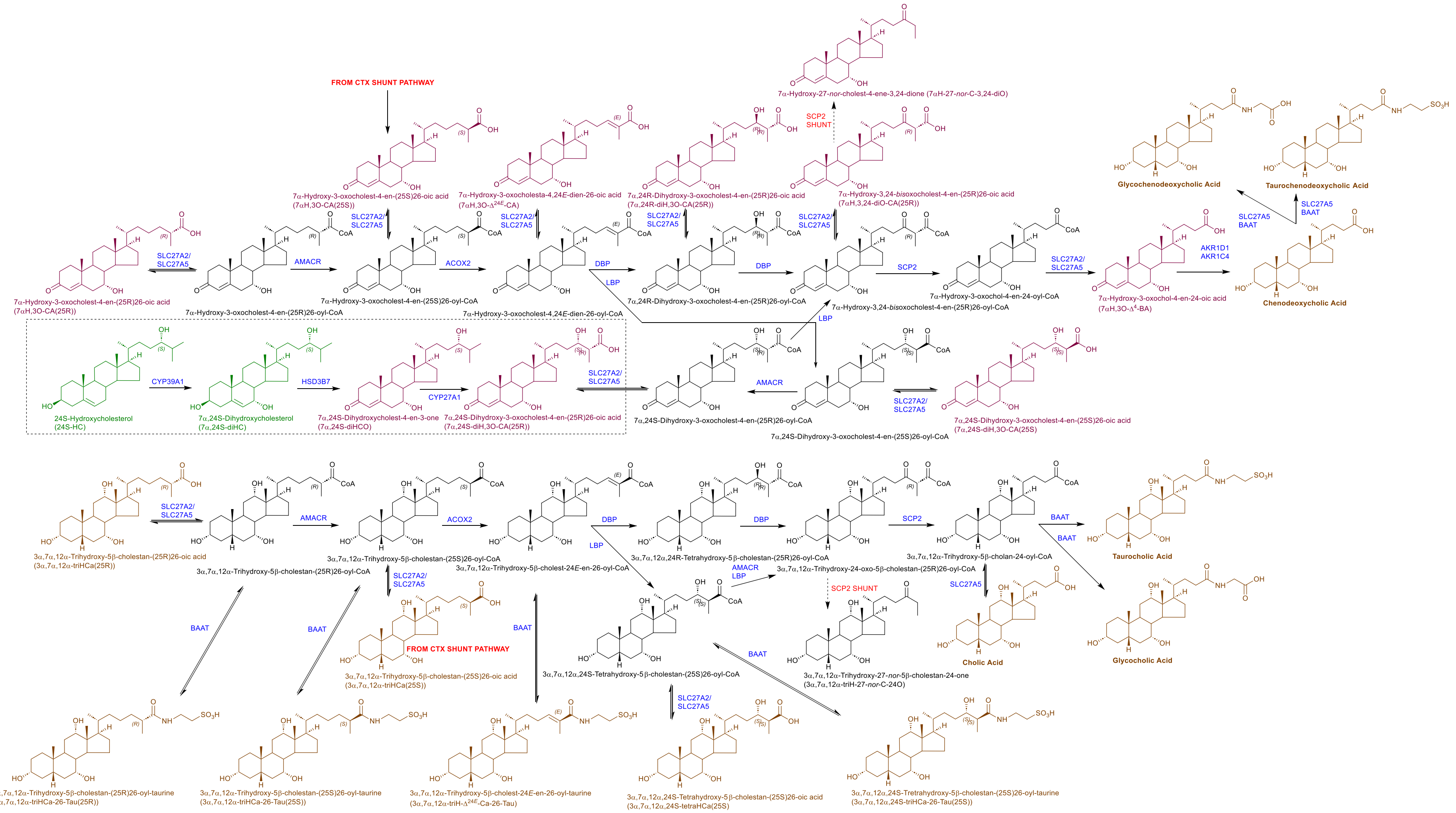

FROM CTX SHUNT PATHWAY
7α-Hydroxy-27-nor-cholest-4-ene-3,24-dione (7αH-27-nor-C-3,24-diO)
SCP2 SHUNT
7α-Hydroxy-3-oxocholest-4-en-(25S)26-oic acid (7αH,3O-CA(25S))
7α-Hydroxy-3-oxocholesta-4,24E-dien-26-oic acid (7αH,3O-Δ24E-CA)
7α,24R-Dihydroxy-3-oxocholest-4-en-(25R)26-oic acid (7α,24R-diH,3O-CA(25R))
7α-Hydroxy-3,24-bisoxocholest-4-en-(25R)26-oic acid (7αH,3,24-diO-CA(25R))
Glycochenodeoxycholic Acid
Taurochenodeoxycholic Acid
SLC27A5 BAAT
SLC27A2/ SLC27A5
AMACR
ACOX2
DBP
LBP
SCP2
AKR1D1 AKR1C4
7α-Hydroxy-3-oxocholest-4-en-(25R)26-oic acid (7αH,3O-CA(25R))
7α-Hydroxy-3-oxocholest-4-en-(25R)26-oyl-CoA
7α-Hydroxy-3-oxocholest-4-en-(25S)26-oyl-CoA
7α-Hydroxy-3-oxocholest-4,24E-dien-26-oyl-CoA
7α,24R-Dihydroxy-3-oxocholest-4-en-(25R)26-oyl-CoA
7α-Hydroxy-3,24-bisoxocholest-4-en-(25R)26-oyl-CoA
7α-Hydroxy-3-oxochol-4-en-24-oyl-CoA
7α-Hydroxy-3-oxochol-4-en-24-oic acid (7αH,3O-Δ4-BA)
Chenodeoxycholic Acid
CYP39A1
HSD3B7
CYP27A1
24S-Hydroxycholesterol (24S-HC)
7α,24S-Dihydroxycholesterol (7α,24S-diHC)
7α,24S-Dihydroxycholest-4-en-3-one (7α,24S-diHCO)
7α,24S-Dihydroxy-3-oxocholest-4-en-(25R)26-oic acid (7α,24S-diH,3O-CA(25R))
7α,24S-Dihydroxy-3-oxocholest-4-en-(25R)26-oyl-CoA
7α,24S-Dihydroxy-3-oxocholest-4-en-(25S)26-oyl-CoA
7α,24S-Dihydroxy-3-oxocholest-4-en-(25S)26-oic acid (7α,24S-diH,3O-CA(25S))
3α,7α,12α-Trihydroxy-5β-cholestan-(25R)26-oic acid (3α,7α,12α-triHCa(25R))
3α,7α,12α-Trihydroxy-5β-cholestan-(25R)26-oyl-CoA
3α,7α,12α-Trihydroxy-5β-cholestan-(25S)26-oyl-CoA
3α,7α,12α-Trihydroxy-5β-cholest-24E-en-26-oyl-CoA
3α,7α,12α,24R-Tetrahydroxy-5β-cholestan-(25R)26-oyl-CoA
3α,7α,12α-Trihydroxy-24-oxo-5β-cholestan-(25R)26-oyl-CoA
3α,7α,12α-Trihydroxy-5β-cholan-24-oyl-CoA
BAAT
Taurocholic Acid
Glycocholic Acid
Cholic Acid
AMACR LBP
SCP2 SHUNT
3α,7α,12α-Trihydroxy-5β-cholestan-(25S)26-oic acid (3α,7α,12α-triHCa(25S))
3α,7α,12α,24S-Tetrahydroxy-5β-cholestan-(25S)26-oyl-CoA
3α,7α,12α-Trihydroxy-27-nor-5β-cholestan-24-one (3α,7α,12α-triH-27-nor-C-24O)
3α,7α,12α-Trihydroxy-5β-cholestan-(25R)26-oyl-taurine (3α,7α,12α-triHCa-26-Tau(25R))
3α,7α,12α-Trihydroxy-5β-cholestan-(25S)26-oyl-taurine (3α,7α,12α-triHCa-26-Tau(25S))
3α,7α,12α-Trihydroxy-5β-cholest-24E-en-26-oyl-taurine (3α,7α,12α-triH-Δ24E-Ca-26-Tau)
3α,7α,12α,24S-Tetrahydroxy-5β-cholestan-(25S)26-oic acid (3α,7α,12α,24S-tetraHCa(25S))
3α,7α,12α,24S-Tretrahydroxy-5β-cholestan-(25S)26-oyl-taurine (3α,7α,12α,24S-triHCa-26-Tau(25S))

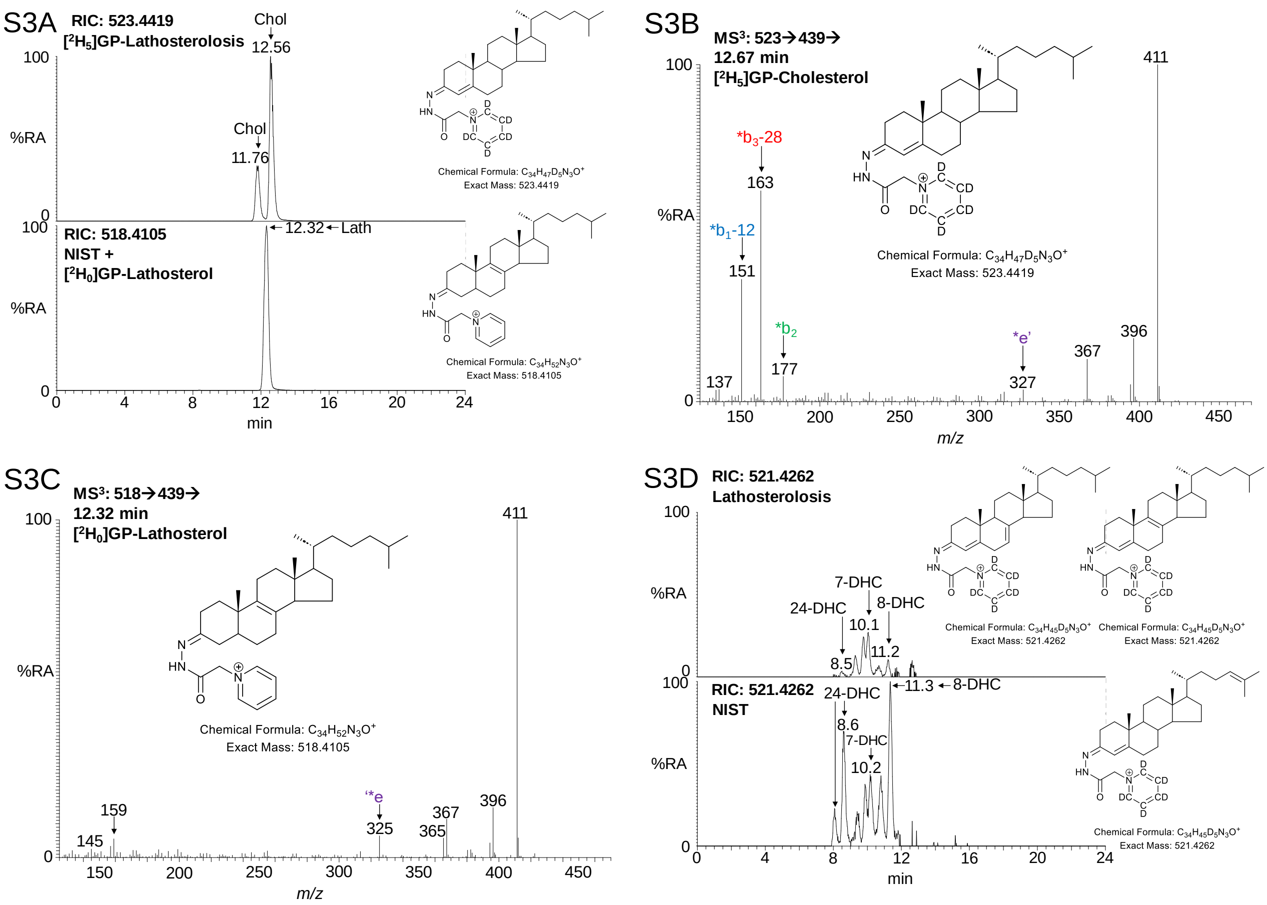
S3A
RIC: 523.4419
[2H5]GP-Lathosterolosis
Chol
12.56
Chol
11.76
100
%RA
0
Chemical Formula: C34H47D5N3O+
Exact Mass: 523.4419
100
RIC: 518.4105
NIST +
[2H0]GP-Lathosterol
12.32
Lath
%RA
0
Chemical Formula: C34H52N3O+
Exact Mass: 518.4105
0
4
8
12
16
20
24
min
S3B
MS3: 523→439→
12.67 min
[2H5]GP-Cholesterol
100
%RA
0
411
*b3-28
163
*b1-12
151
*b2
177
137
*e'
327
367
396
Chemical Formula: C34H47D5N3O+
Exact Mass: 523.4419
150
200
250
300
350
400
450
m/z
S3C
MS3: 518→439→
12.32 min
[2H0]GP-Lathosterol
100
%RA
0
411
Chemical Formula: C34H52N3O+
Exact Mass: 518.4105
159
145
*e
325
367
365
396
150
200
250
300
350
400
450
m/z
S3D
RIC: 521.4262
Lathosterolosis
100
%RA
0
7-DHC
24-DHC
8-DHC
10.1
8.5
11.2
Chemical Formula: C34H45D5N3O+
Exact Mass: 521.4262
Chemical Formula: C34H45D5N3O+
Exact Mass: 521.4262
100
RIC: 521.4262
NIST
24-DHC
11.3
8-DHC
8.6
7-DHC
10.2
%RA
0
Chemical Formula: C34H45D5N3O+
Exact Mass: 521.4262
0
4
8
12
16
20
24
min

S4

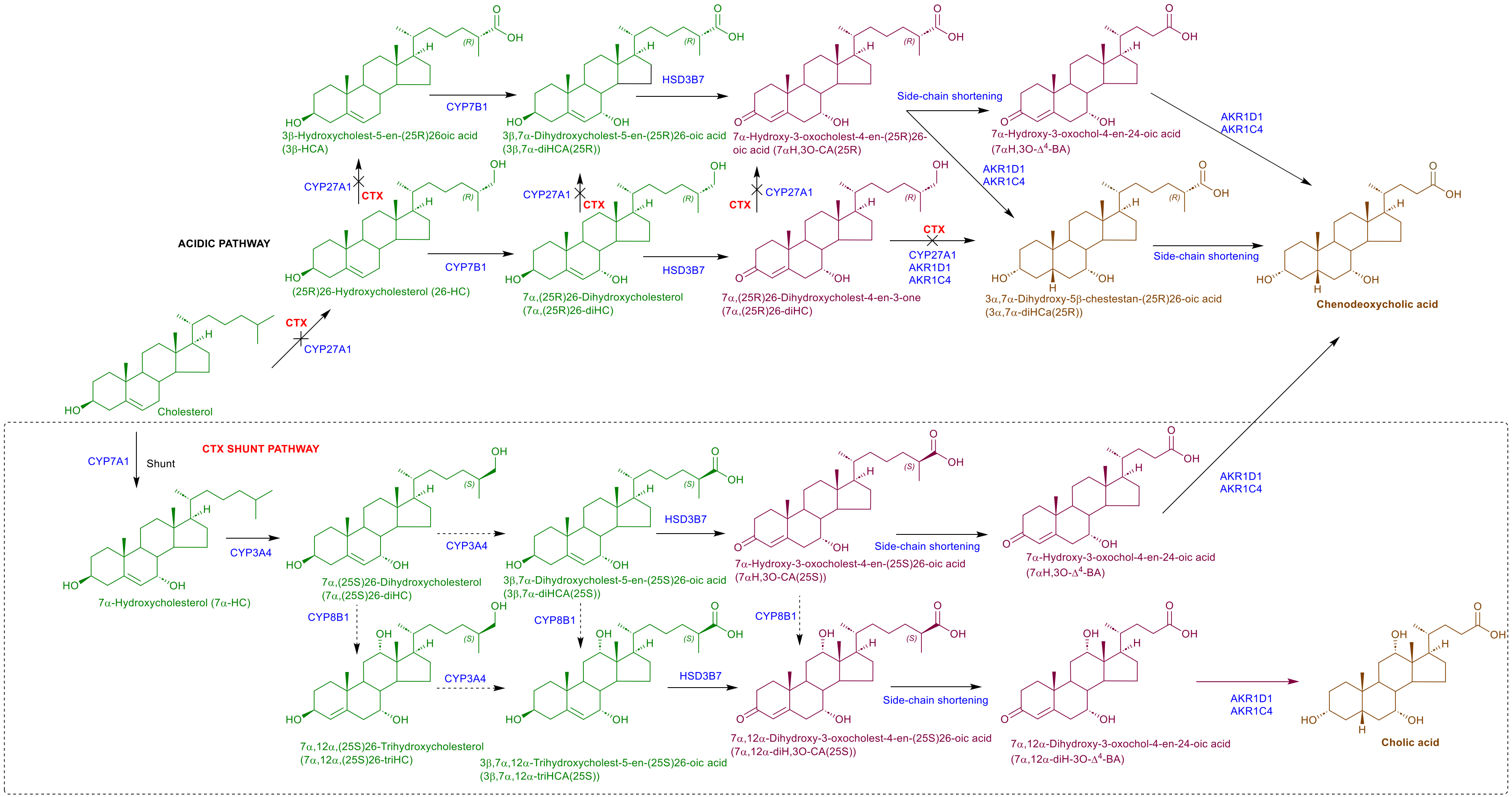

ACIDIC PATHWAY
Cholesterol
CTX
CYP27A1
(25R)26-Hydroxycholesterol (26-HC)
CYP7B1
3β-Hydroxycholest-5-en-(25R)26oic acid
(3β-HCA)
3β,7α-Dihydroxycholest-5-en-(25R)26-oic acid
(3β,7α-diHCA(25R))
HSD3B7
7α-Hydroxy-3-oxocholest-4-en-(25R)26-oic acid (7αH,3O-CA(25R)
Side-chain shortening
7α-Hydroxy-3-oxochol-4-en-24-oic acid
(7αH,3O-Δ4-BA)
AKR1D1
AKR1C4
7α,(25R)26-Dihydroxycholesterol
(7α,(25R)26-diHC)
7α,(25R)26-Dihydroxycholest-4-en-3-one
(7α,(25R)26-diHC)
3α,7α-Dihydroxy-5β-chestestan-(25R)26-oic acid
(3α,7α-diHCa(25R))
Chenodeoxycholic acid
CTX SHUNT PATHWAY
CYP7A1
Shunt
7α-Hydroxycholesterol (7α-HC)
CYP3A4
7α,(25S)26-Dihydroxycholesterol
(7α,(25S)26-diHC)
3β,7α-Dihydroxycholest-5-en-(25S)26-oic acid
(3β,7α-diHCA(25S))
7α-Hydroxy-3-oxocholest-4-en-(25S)26-oic acid
(7αH,3O-CA(25S))
CYP8B1
7α,12α,(25S)26-Trihydroxycholesterol
(7α,12α,(25S)26-triHC)
3β,7α,12α-Trihydroxycholest-5-en-(25S)26-oic acid
(3β,7α,12α-triHCA(25S))
7α,12α-Dihydroxy-3-oxocholest-4-en-(25S)26-oic acid
(7α,12α-diH,3O-CA(25S))
7α,12α-Dihydroxy-3-oxochol-4-en-24-oic acid
(7α,12α-diH-3O-Δ4-BA)
Cholic acid

S5A

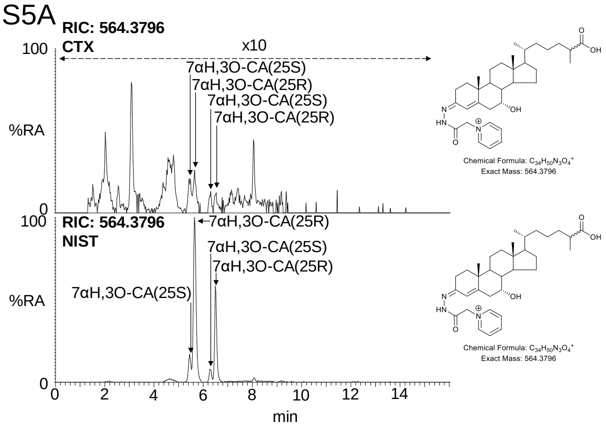

RIC: 564.3796
CTX
x10
7αH,3O-CA(25S)
7αH,3O-CA(25R)
7αH,3O-CA(25S)
7αH,3O-CA(25R)
RIC: 564.3796
NIST
7αH,3O-CA(25R)
7αH,3O-CA(25S)
7αH,3O-CA(25R)
7αH,3O-CA(25S)
%RA
100
0
0 2 4 6 8 10 12 14
min
Chemical Formula: $C_{34}H_{50}N_3O_4^+$
Exact Mass: 564.3796


S5B

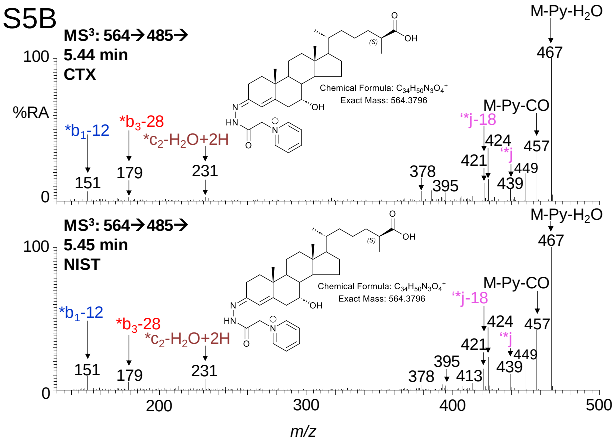

MS³: 564→485→
5.44 min
CTX
M-Py-H₂O
467
M-Py-CO
457
*b₁-12
151
*b₃-28
179
*c₂-H₂O+2H
231
*j-18
*j
378
395
421
424
439
449
MS³: 564→485→
5.45 min
NIST
378
395
413
Chemical Formula: $C_{34}H_{50}N_3O_4^+$
Exact Mass: 564.3796
%RA
100
0
200 300 400 500
m/z


S5C

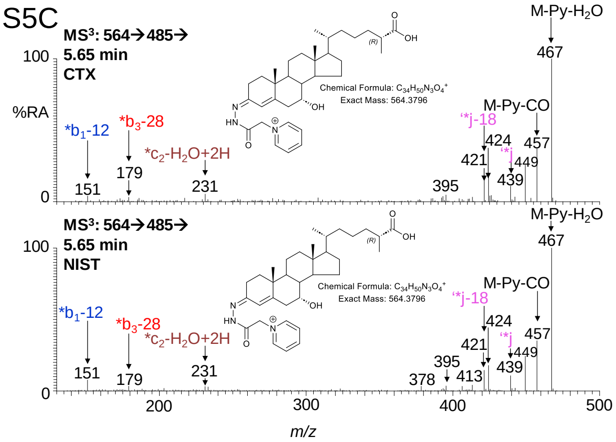

MS³: 564→485→
5.65 min
CTX
M-Py-H₂O
467
M-Py-CO
457
*b₁-12
151
*b₃-28
179
*c₂-H₂O+2H
231
*j-18
*j
395
421
424
439
449
MS³: 564→485→
5.65 min
NIST
378
395
413
Chemical Formula: $C_{34}H_{50}N_3O_4^+$
Exact Mass: 564.3796
%RA
100
0
200 300 400 500
m/z

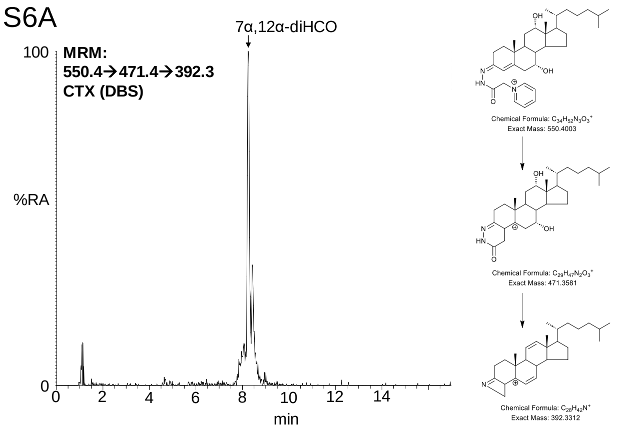

S6A
7α,12α-diHCO
MRM:
550.4→471.4→392.3
CTX (DBS)
100
%RA
0
0 2 4 6 8 10 12 14
min
Chemical Formula: C34H52N3O3+
Exact Mass: 550.4003
Chemical Formula: C29H47N2O3+
Exact Mass: 471.3581
Chemical Formula: C28H42N+
Exact Mass: 392.3312


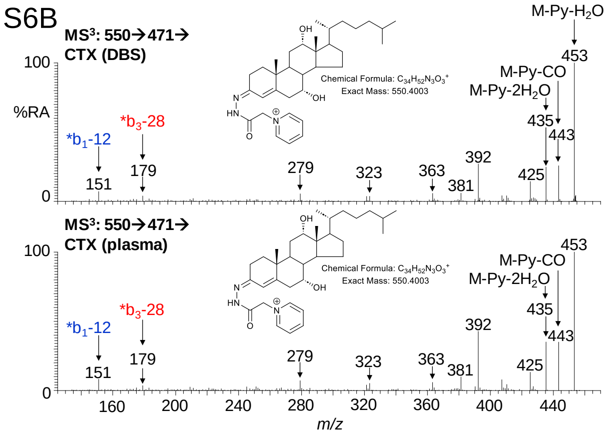

S6B
MS3: 550→471→
CTX (DBS)
Chemical Formula: C34H52N3O3+
Exact Mass: 550.4003
M-Py-H2O
453
M-Py-CO
M-Py-2H2O
435
443
*b1-12
*b3-28
151
179
279
323
363
381
392
425
MS3: 550→471→
CTX (plasma)
100
%RA
0
160 200 240 280 320 360 400 440
m/z

S7A

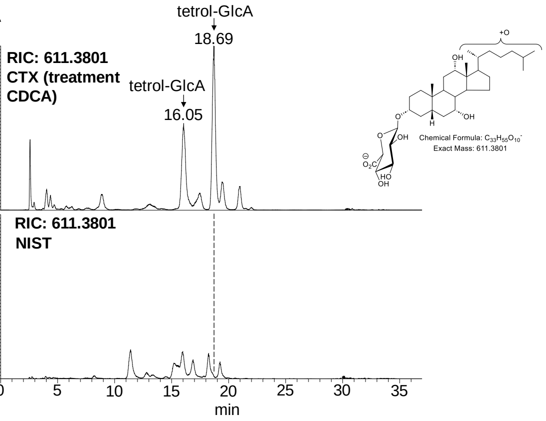


S7B

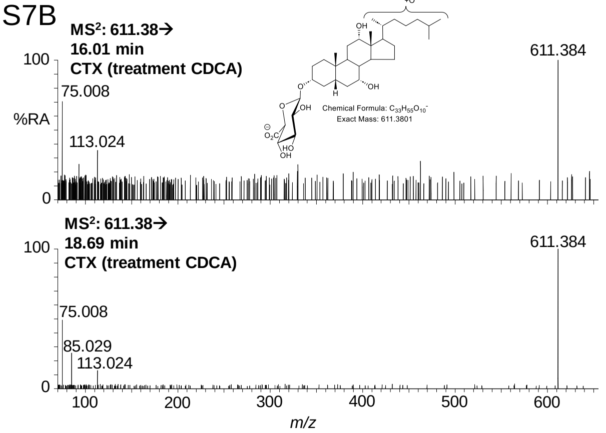


S7C

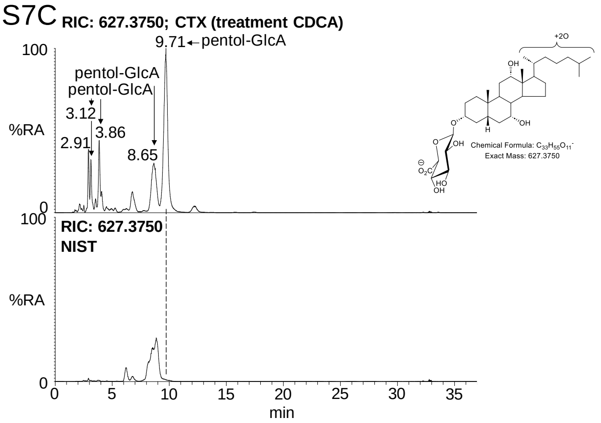


S7D

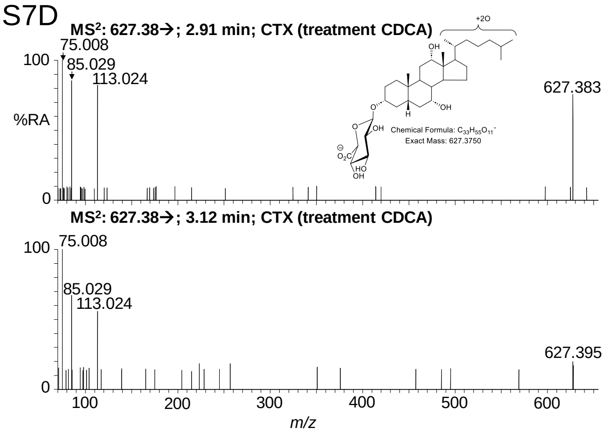


S7E

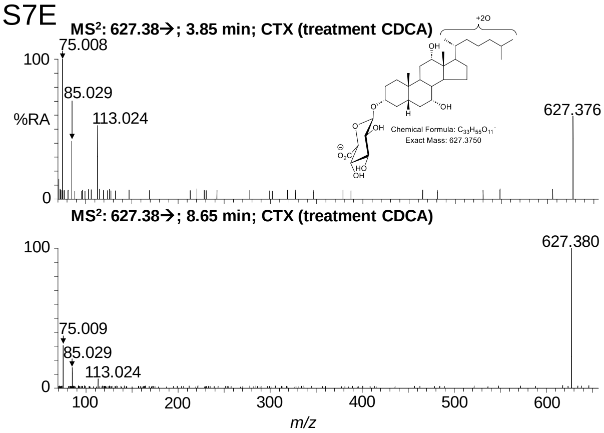


S7F

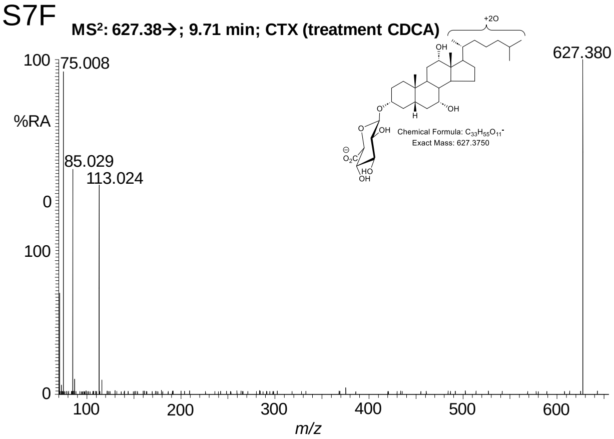


S7G

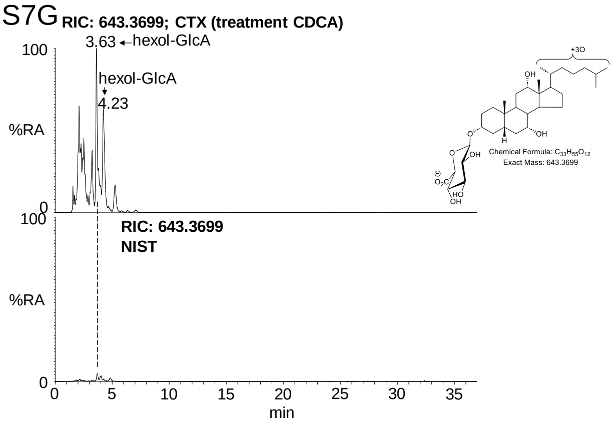


S7H

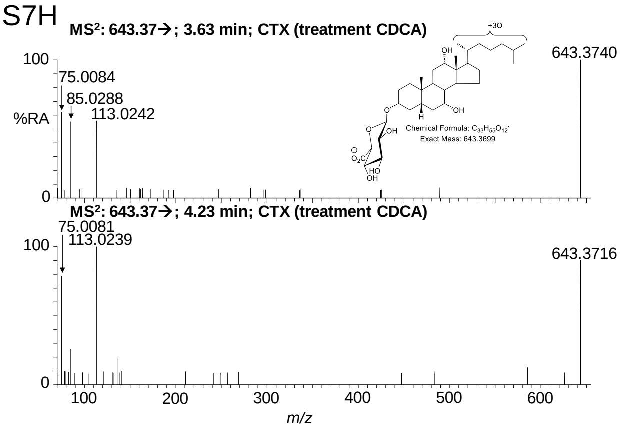

# S8

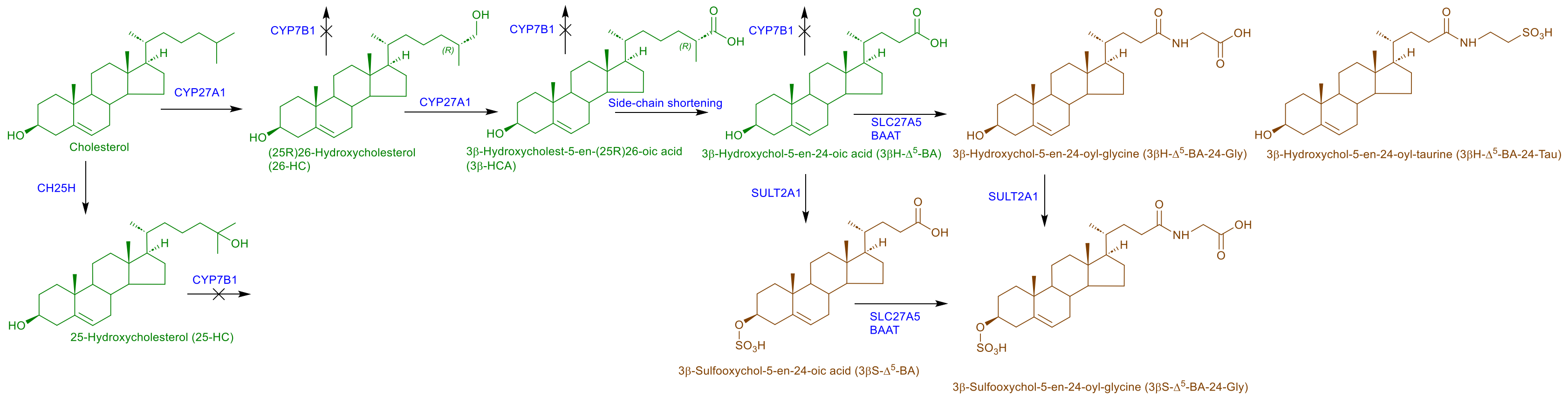

CYP27A1
Cholesterol
CYP7B1
(25R)26-Hydroxycholesterol
(26-HC)
CYP27A1
CYP7B1
3β-Hydroxycholest-5-en-(25R)26-oic acid
(3β-HCA)
Side-chain shortening
CYP7B1
3β-Hydroxychol-5-en-24-oic acid (3βH-Δ5-BA)
SLC27A5
BAAT
3β-Hydroxychol-5-en-24-oyl-glycine (3βH-Δ5-BA-24-Gly)
3β-Hydroxychol-5-en-24-oyl-taurine (3βH-Δ5-BA-24-Tau)
CH25H
25-Hydroxycholesterol (25-HC)
CYP7B1
SULT2A1
SULT2A1
3β-Sulfooxychol-5-en-24-oic acid (3βS-Δ5-BA)
SLC27A5
BAAT
3β-Sulfooxychol-5-en-24-oyl-glycine (3βS-Δ5-BA-24-Gly)

S9A

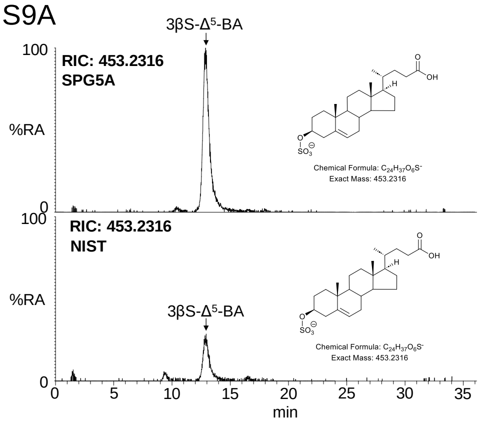


S9D

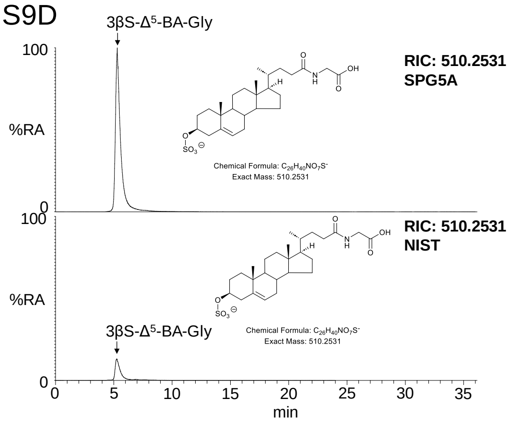


S9B

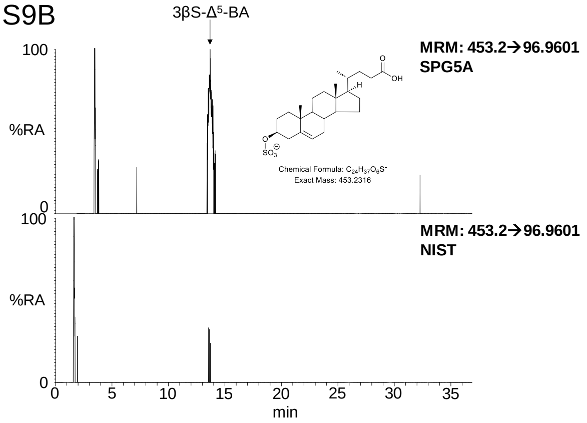


S9E

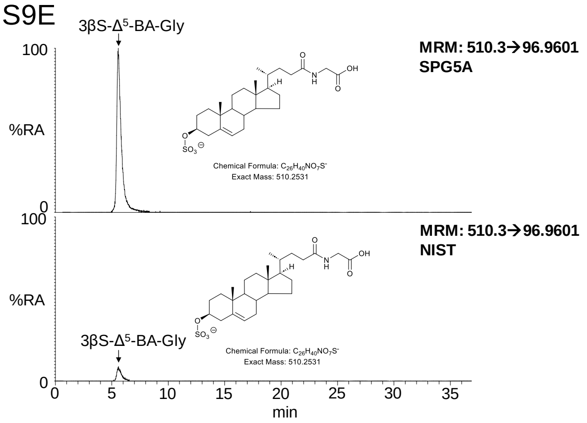


S9C

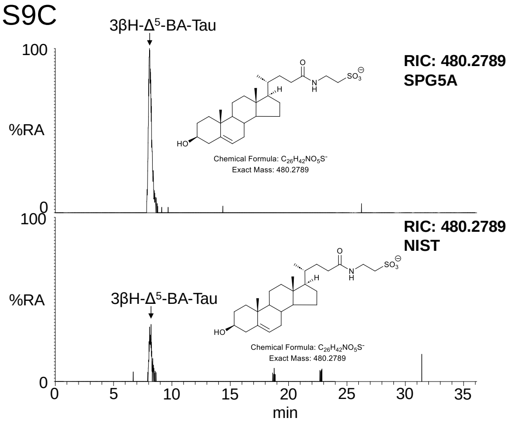


S9F

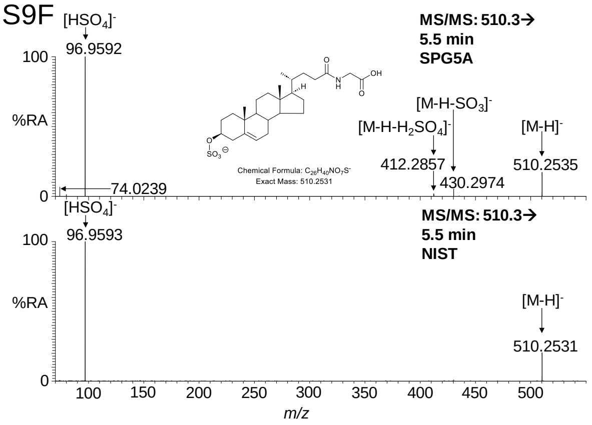

S10A

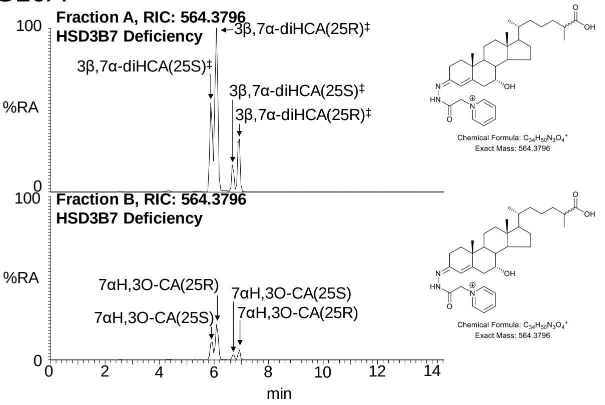
Fraction A, RIC: 564.3796
HSD3B7 Deficiency
3β,7α-diHCA(25R)‡
3β,7α-diHCA(25S)‡
3β,7α-diHCA(25S)‡
3β,7α-diHCA(25R)‡
%RA
100
0
Chemical Formula: C34H50N3O4+
Exact Mass: 564.3796
Fraction B, RIC: 564.3796
HSD3B7 Deficiency
7αH,3O-CA(25R)
7αH,3O-CA(25S)
7αH,3O-CA(25S)
7αH,3O-CA(25R)
0 2 4 6 8 10 12 14
min

S10B

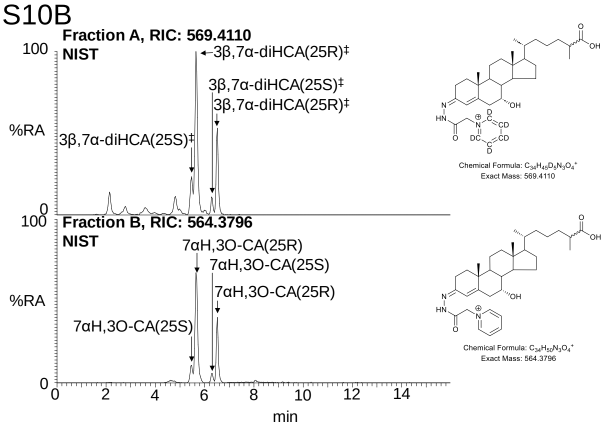
Fraction A, RIC: 569.4110
NIST
3β,7α-diHCA(25R)‡
3β,7α-diHCA(25S)‡
3β,7α-diHCA(25R)‡
3β,7α-diHCA(25S)‡
%RA
100
0
Chemical Formula: C34H45D5N3O4+
Exact Mass: 569.4110
Fraction B, RIC: 564.3796
NIST
7αH,3O-CA(25R)
7αH,3O-CA(25S)
7αH,3O-CA(25R)
7αH,3O-CA(25S)
Chemical Formula: C34H50N3O4+
Exact Mass: 564.3796
0 2 4 6 8 10 12 14
min

S11A

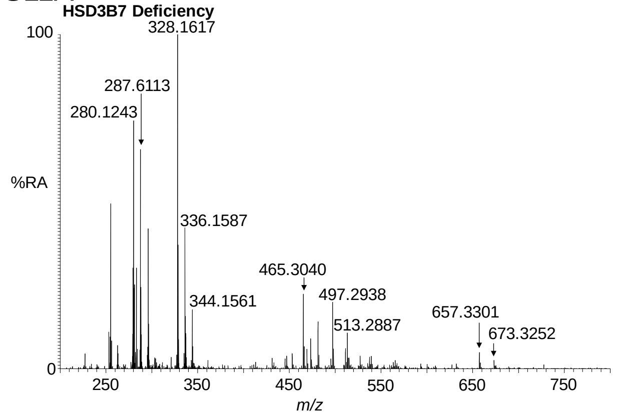


S11B

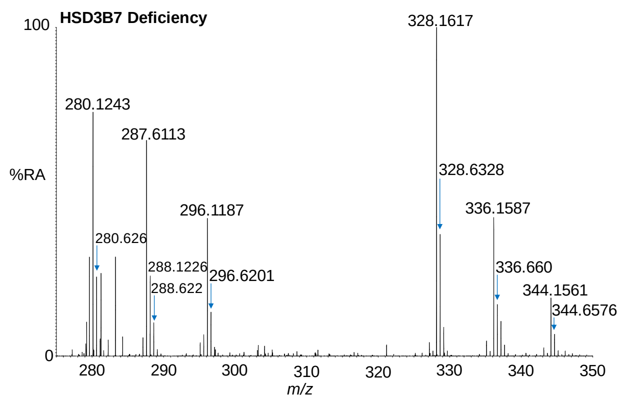


S11C

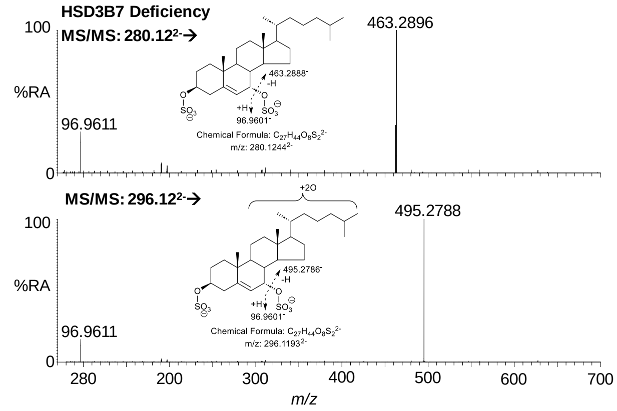


S11D

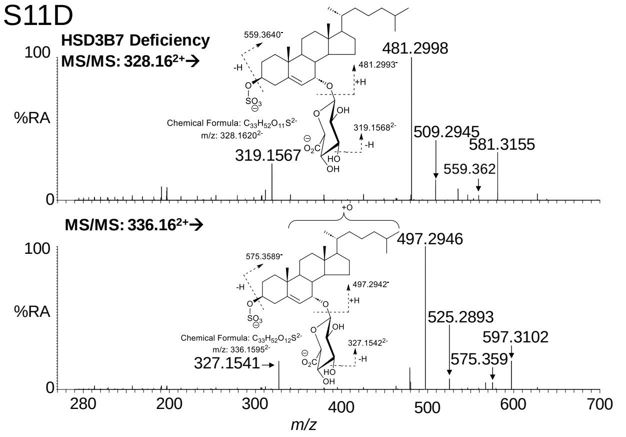


S11E

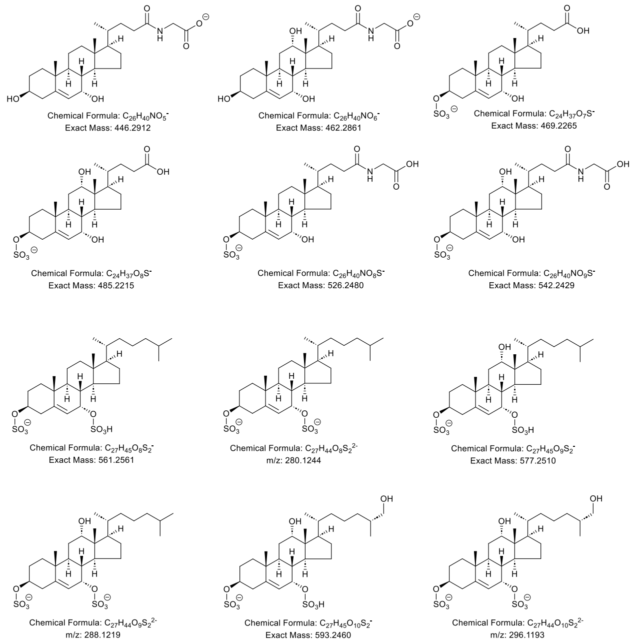


S11F

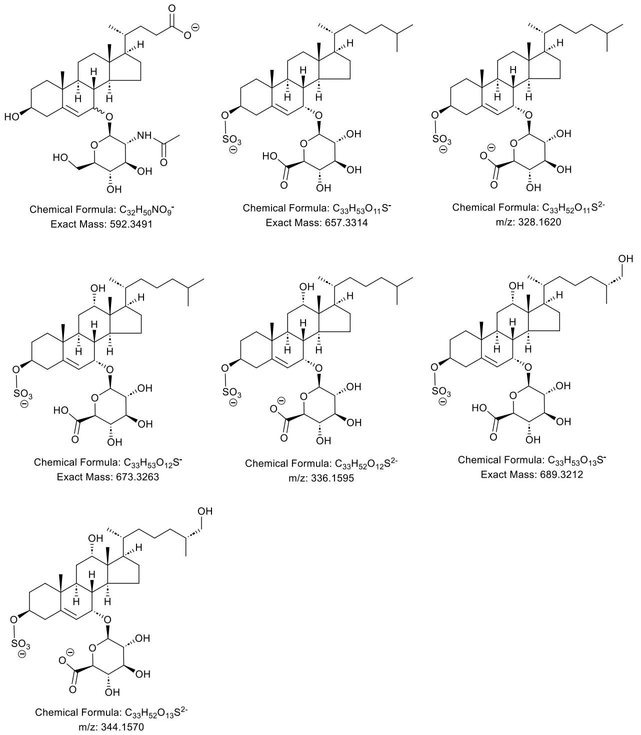

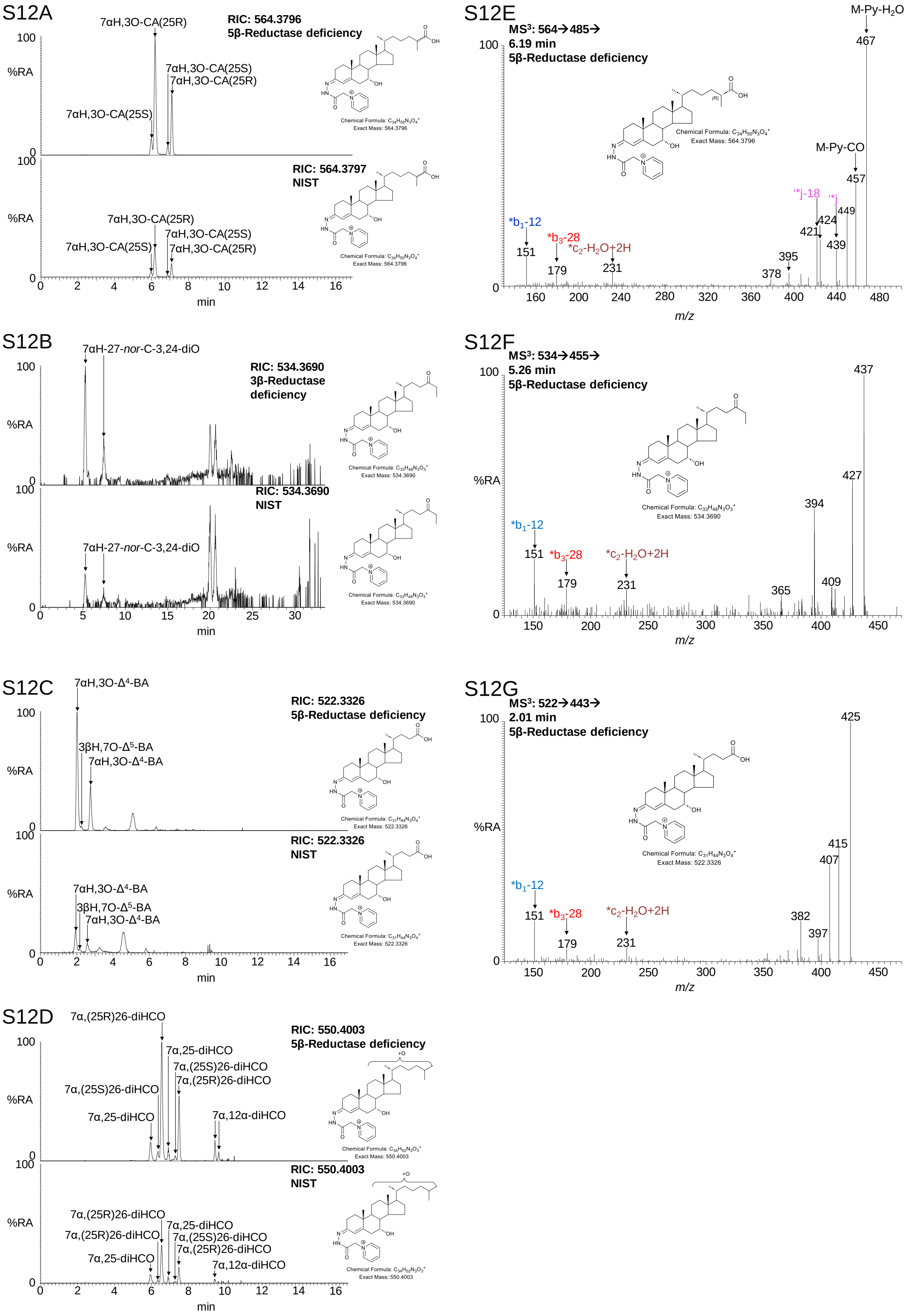
S12A
RIC: 564.3796
5β-Reductase deficiency
7αH,3O-CA(25R)
7αH,3O-CA(25S)
7αH,3O-CA(25S)
7αH,3O-CA(25R)
RIC: 564.3797
NIST
%RA
min
S12B
7αH-27-nor-C-3,24-diO
RIC: 534.3690
3β-Reductase deficiency
RIC: 534.3690
NIST
S12C
7αH,3O-Δ4-BA
3βH,7O-Δ5-BA
7αH,3O-Δ4-BA
RIC: 522.3326
5β-Reductase deficiency
RIC: 522.3326
NIST
S12D
7α,(25R)26-diHCO
7α,25-diHCO
7α,(25S)26-diHCO
7α,(25R)26-diHCO
7α,(25S)26-diHCO
7α,25-diHCO
7α,12α-diHCO
RIC: 550.4003
5β-Reductase deficiency
RIC: 550.4003
NIST
S12E
MS3: 564→485→
6.19 min
5β-Reductase deficiency
M-Py-H2O
467
M-Py-CO
457
*b1-12
151
*b3-28
179
*c2-H2O+2H
231
'*j-18
'*j
421
424
439
449
395
378
m/z
S12F
MS3: 534→455→
5.26 min
5β-Reductase deficiency
437
427
394
365
409
*b1-12
151
*b3-28
179
*c2-H2O+2H
231
S12G
MS3: 522→443→
2.01 min
5β-Reductase deficiency
425
415
407
382
397
*b1-12
151
*b3-28
179
*c2-H2O+2H
231
Chemical Formula: C34H50N3O4+
Exact Mass: 564.3796
Chemical Formula: C33H48N3O3+
Exact Mass: 534.3690
Chemical Formula: C31H44N3O4+
Exact Mass: 522.3326
Chemical Formula: C34H52N3O3+
Exact Mass: 550.4003

S13A

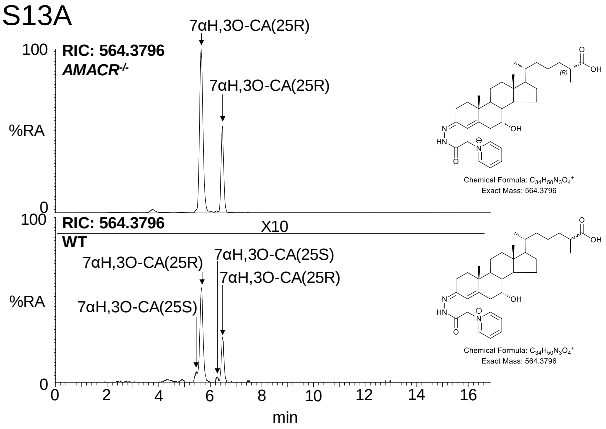


S13C

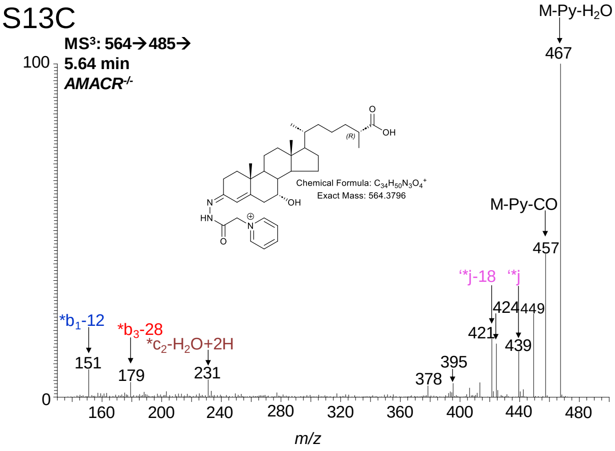


S13B

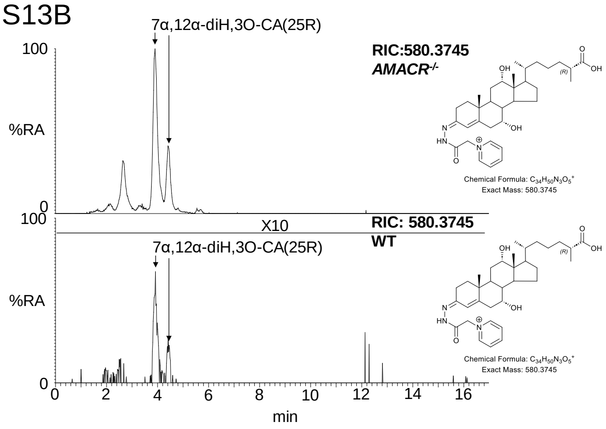


S13D

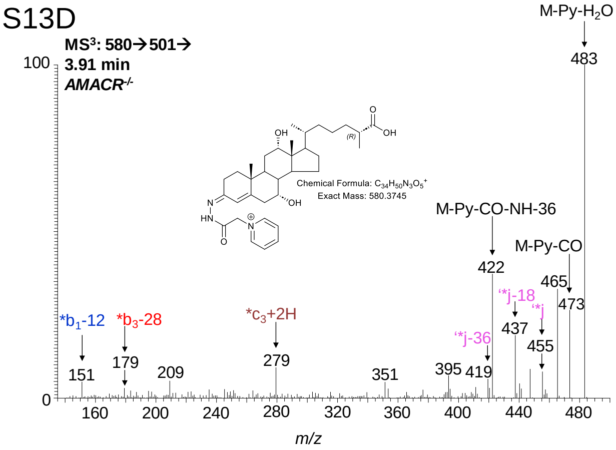

S14A

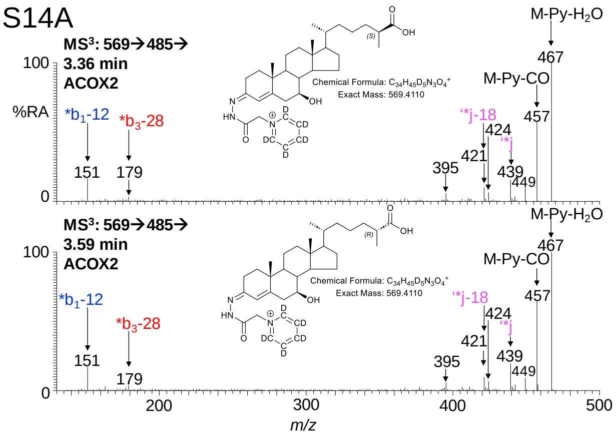


S14B

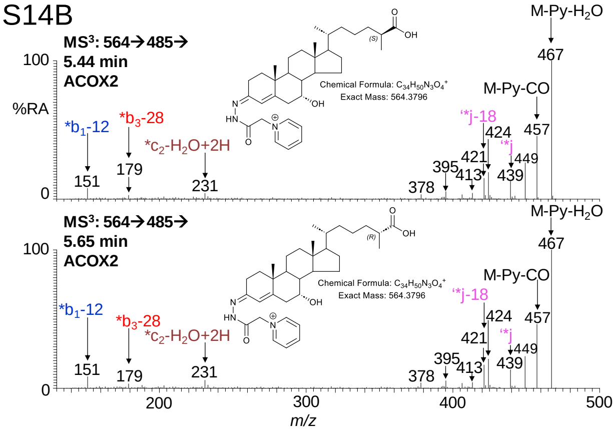


S14C

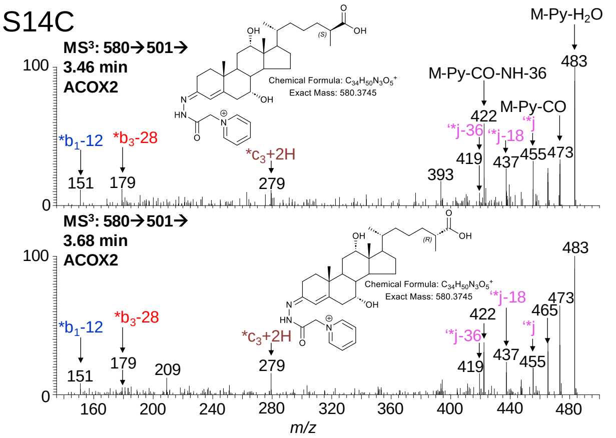

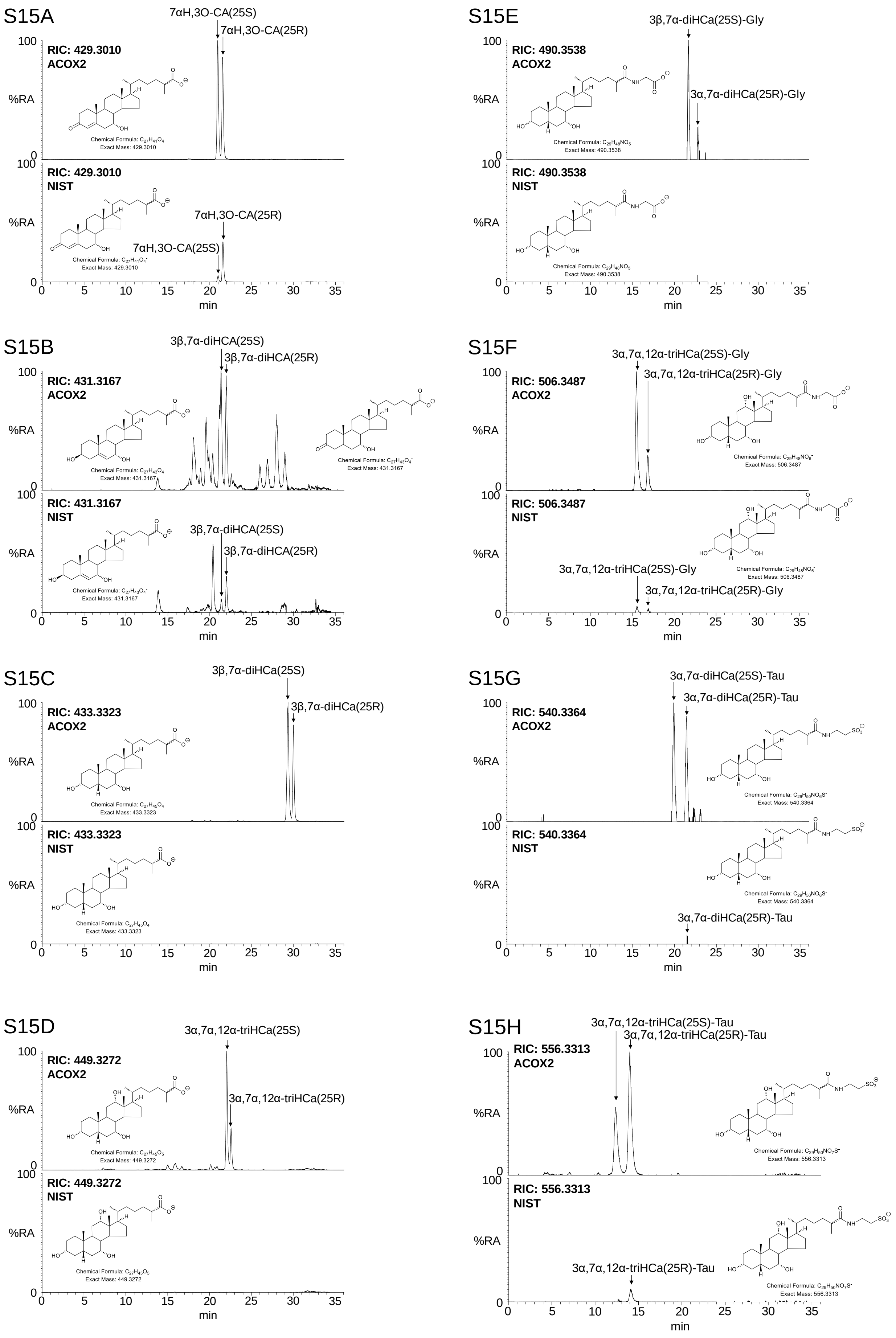
S15A
7αH,3O-CA(25S)
7αH,3O-CA(25R)
RIC: 429.3010
ACOX2
Chemical Formula: $C_{27}H_{41}O_4^-$
Exact Mass: 429.3010
RIC: 429.3010
NIST
7αH,3O-CA(25R)
7αH,3O-CA(25S)
Chemical Formula: $C_{27}H_{41}O_4^-$
Exact Mass: 429.3010
%RA
min
S15E
3β,7α-diHCa(25S)-Gly
3α,7α-diHCa(25R)-Gly
RIC: 490.3538
ACOX2
Chemical Formula: $C_{29}H_{48}NO_5^-$
Exact Mass: 490.3538
RIC: 490.3538
NIST
Chemical Formula: $C_{29}H_{48}NO_5^-$
Exact Mass: 490.3538
S15B
3β,7α-diHCA(25S)
3β,7α-diHCA(25R)
RIC: 431.3167
ACOX2
Chemical Formula: $C_{27}H_{43}O_4^-$
Exact Mass: 431.3167
Chemical Formula: $C_{27}H_{43}O_4^-$
Exact Mass: 431.3167
RIC: 431.3167
NIST
3β,7α-diHCA(25S)
3β,7α-diHCA(25R)
Chemical Formula: $C_{27}H_{43}O_4^-$
Exact Mass: 431.3167
S15F
3α,7α,12α-triHCa(25S)-Gly
3α,7α,12α-triHCa(25R)-Gly
RIC: 506.3487
ACOX2
Chemical Formula: $C_{29}H_{48}NO_6^-$
Exact Mass: 506.3487
RIC: 506.3487
NIST
Chemical Formula: $C_{29}H_{48}NO_6^-$
Exact Mass: 506.3487
3α,7α,12α-triHCa(25S)-Gly
3α,7α,12α-triHCa(25R)-Gly
S15C
3β,7α-diHCa(25S)
3β,7α-diHCa(25R)
RIC: 433.3323
ACOX2
Chemical Formula: $C_{27}H_{45}O_4^-$
Exact Mass: 433.3323
RIC: 433.3323
NIST
Chemical Formula: $C_{27}H_{45}O_4^-$
Exact Mass: 433.3323
S15G
3α,7α-diHCa(25S)-Tau
3α,7α-diHCa(25R)-Tau
RIC: 540.3364
ACOX2
Chemical Formula: $C_{29}H_{50}NO_6S^-$
Exact Mass: 540.3364
RIC: 540.3364
NIST
Chemical Formula: $C_{29}H_{50}NO_6S^-$
Exact Mass: 540.3364
3α,7α-diHCa(25R)-Tau
S15D
3α,7α,12α-triHCa(25S)
3α,7α,12α-triHCa(25R)
RIC: 449.3272
ACOX2
Chemical Formula: $C_{27}H_{45}O_5^-$
Exact Mass: 449.3272
RIC: 449.3272
NIST
Chemical Formula: $C_{27}H_{45}O_5^-$
Exact Mass: 449.3272
S15H
3α,7α,12α-triHCa(25S)-Tau
3α,7α,12α-triHCa(25R)-Tau
RIC: 556.3313
ACOX2
Chemical Formula: $C_{29}H_{50}NO_7S^-$
Exact Mass: 556.3313
RIC: 556.3313
NIST
3α,7α,12α-triHCa(25R)-Tau
Chemical Formula: $C_{29}H_{50}NO_7S^-$
Exact Mass: 556.3313

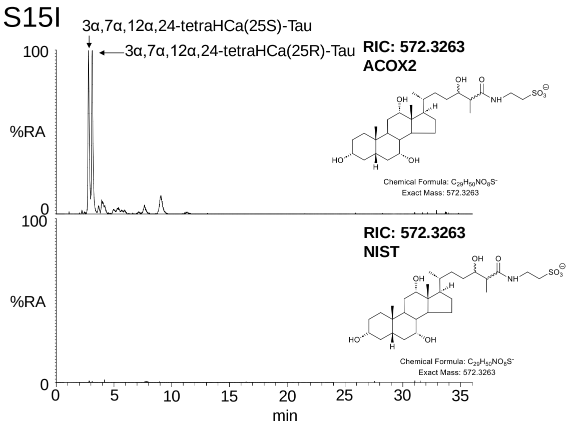

S15I
3α,7α,12α,24-tetraHCa(25S)-Tau
3α,7α,12α,24-tetraHCa(25R)-Tau
RIC: 572.3263
ACOX2
100
%RA
0
Chemical Formula: C29H50NO8S-
Exact Mass: 572.3263
RIC: 572.3263
NIST
100
%RA
0
Chemical Formula: C29H50NO8S-
Exact Mass: 572.3263
0 5 10 15 20 25 30 35
min

S16A

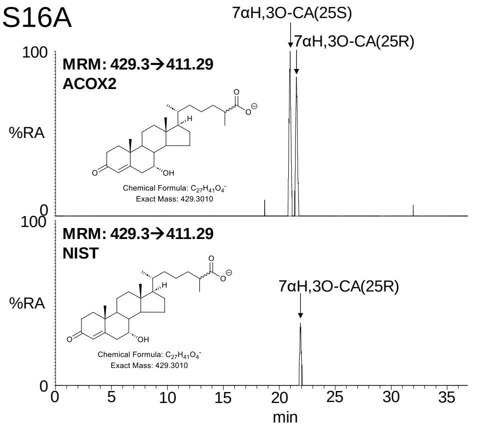


S16E

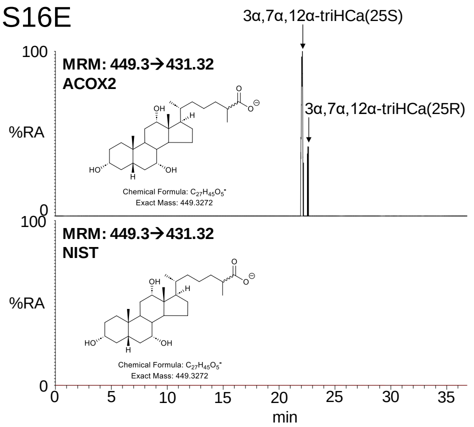


S16B

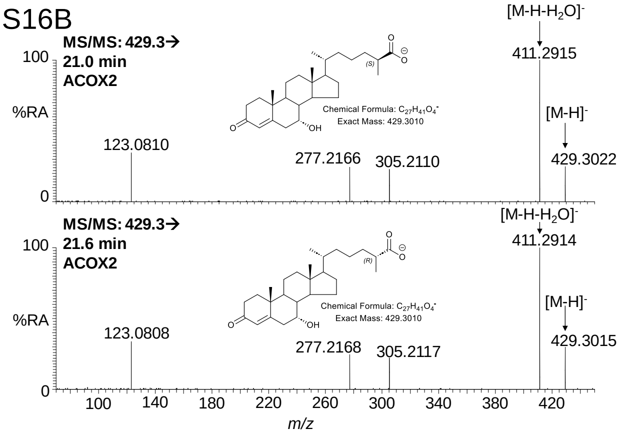


S16F

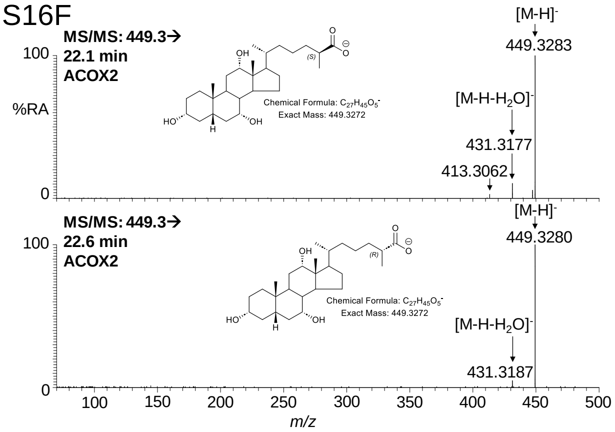


S16C

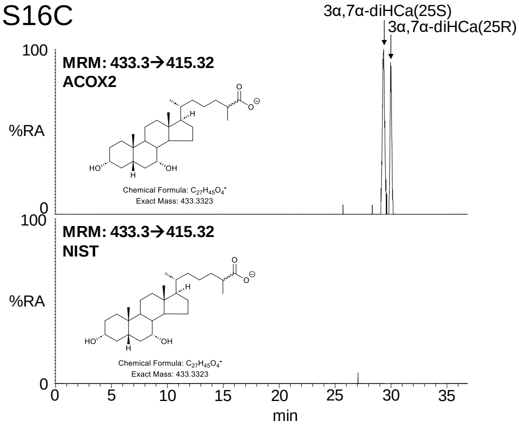


S16G

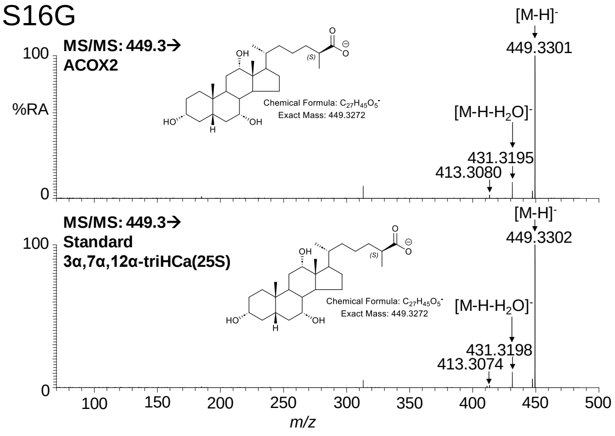


S16D

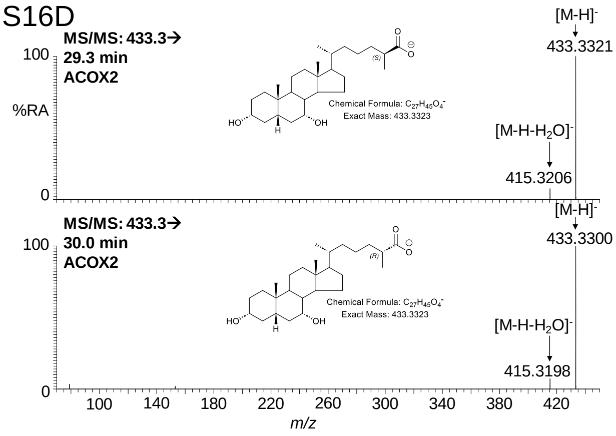


S16H

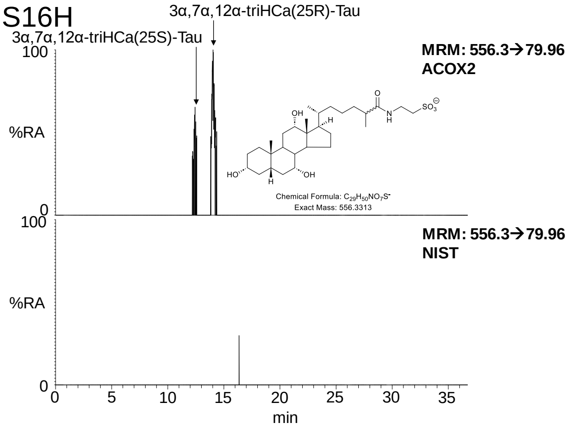

S16I

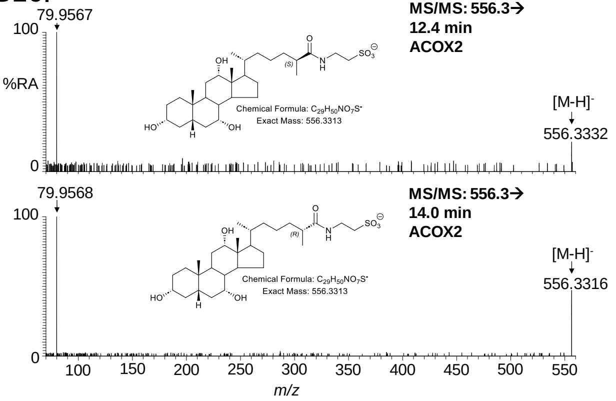

79.9567
100
%RA
0
MS/MS: 556.3→
12.4 min
ACOX2
Chemical Formula: $C_{29}H_{50}NO_7S^-$
Exact Mass: 556.3313
[M-H]-
556.3332
79.9568
100
0
MS/MS: 556.3→
14.0 min
ACOX2
Chemical Formula: $C_{29}H_{50}NO_7S^-$
Exact Mass: 556.3313
[M-H]-
556.3316
100 150 200 250 300 350 400 450 500 550
m/z

S17A

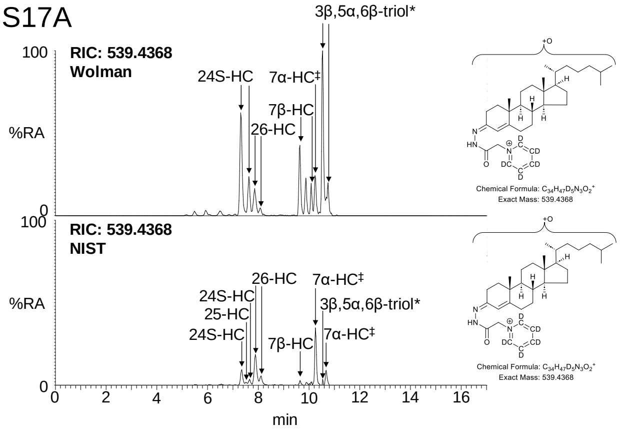


S17E

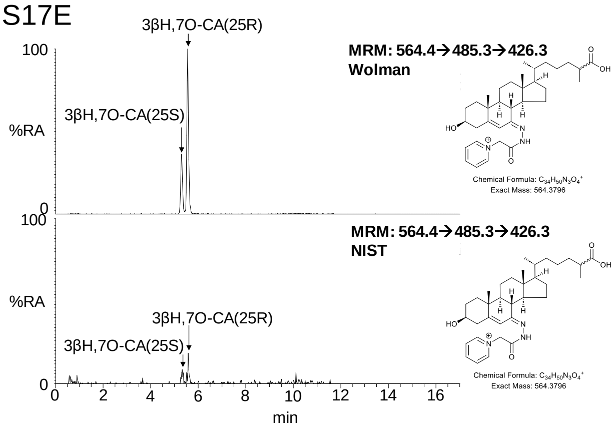


S17B

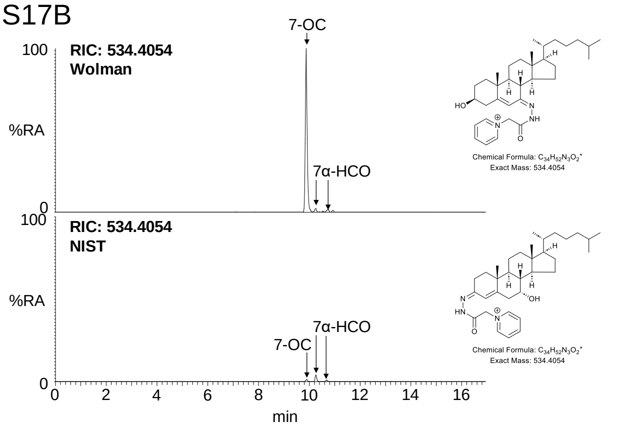


S17F

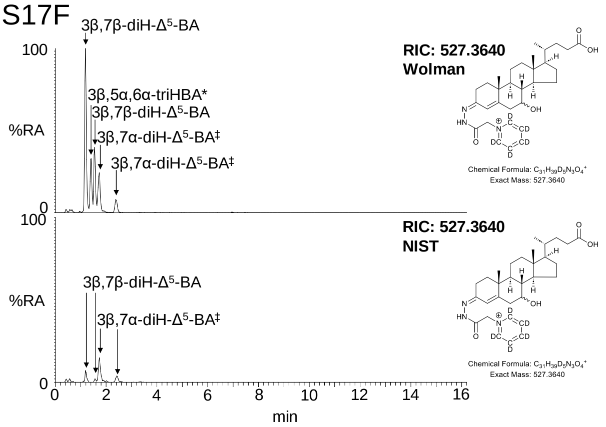


S17C

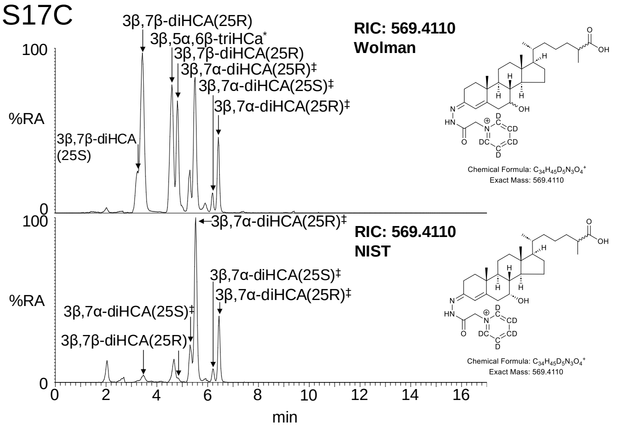


S17G

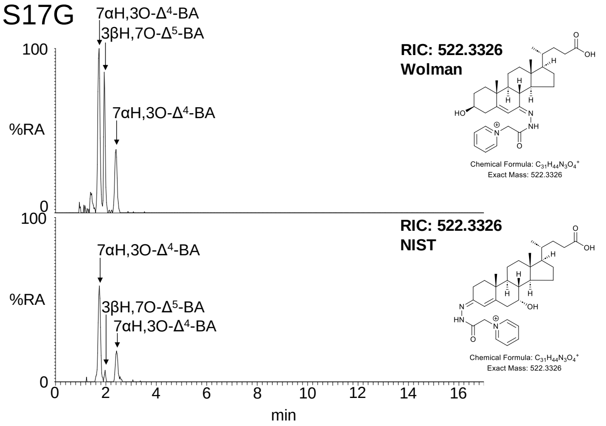


S17D

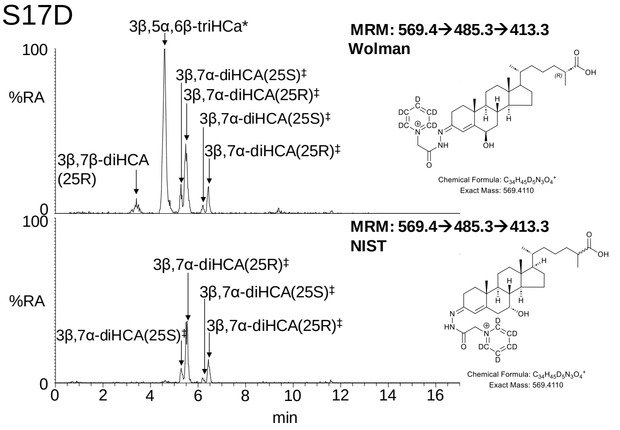

S18A

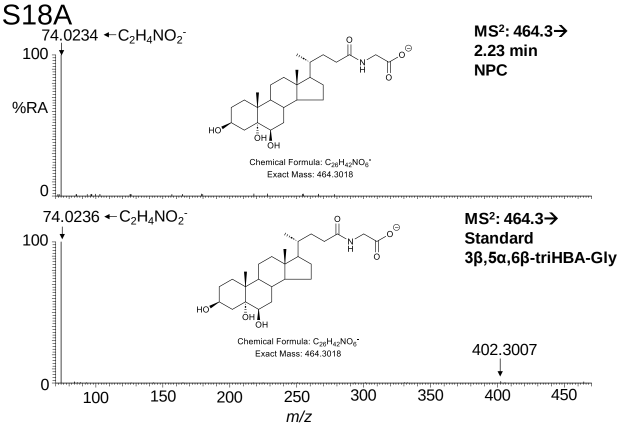


S18E

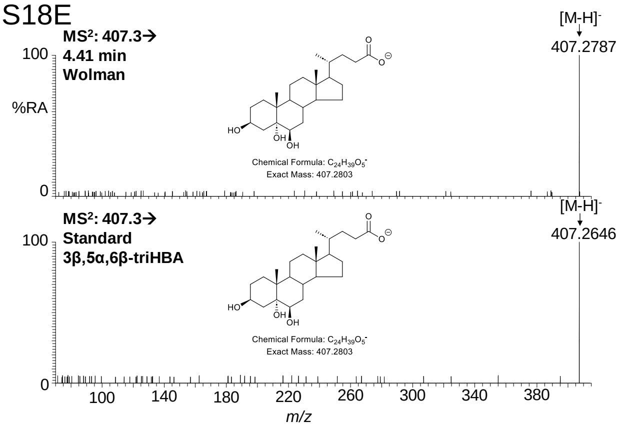


S18B

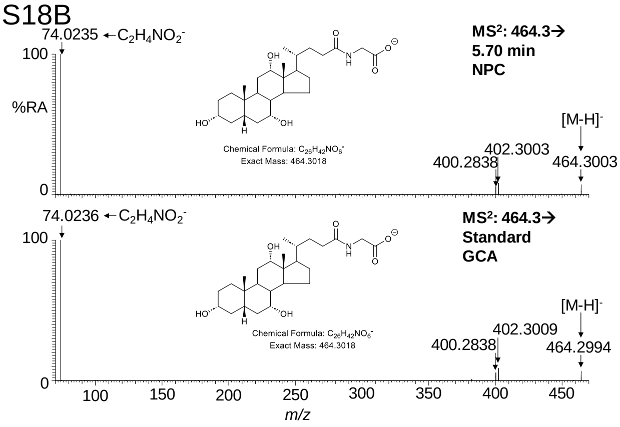


S18F

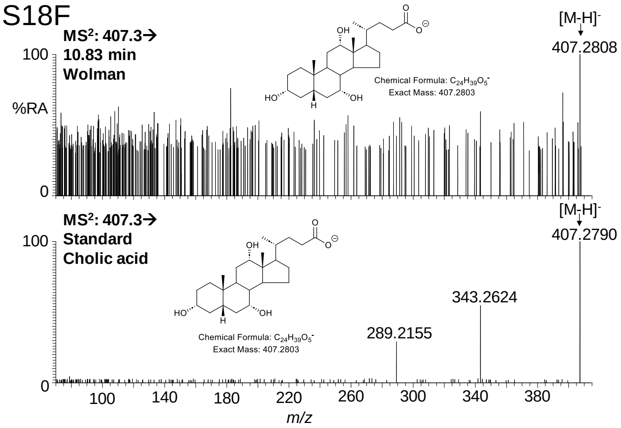


S18C

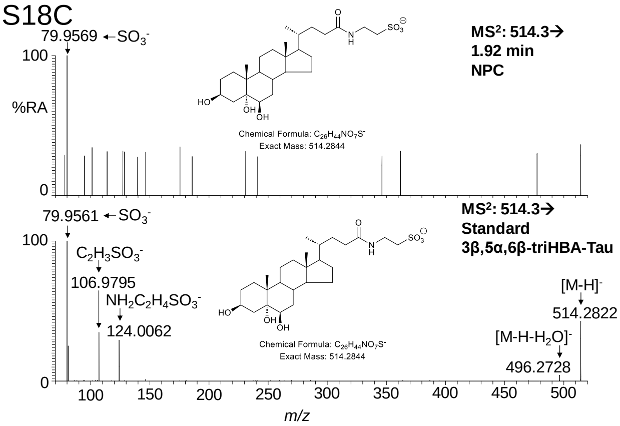


S18G

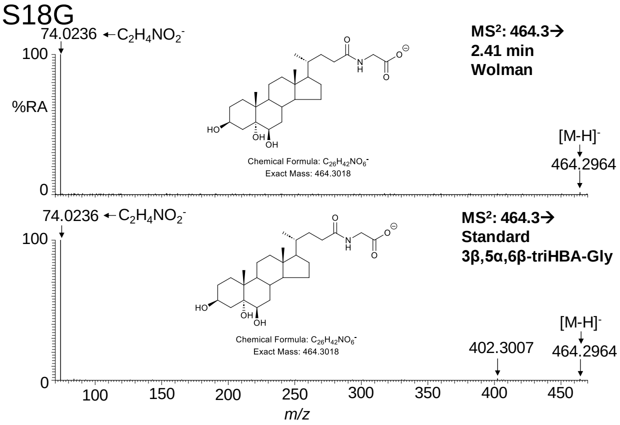


S18D

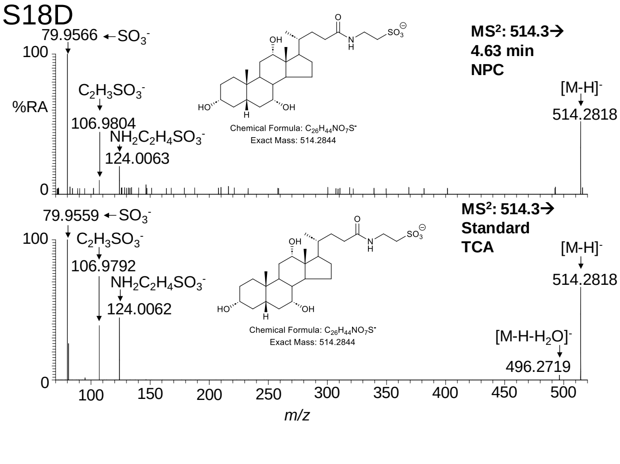


S18H

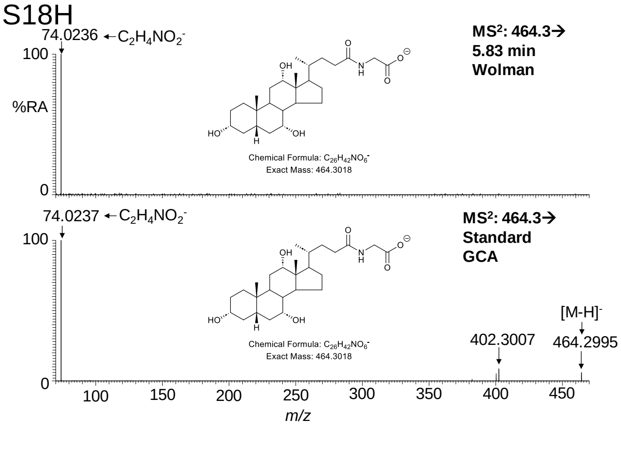

S18I

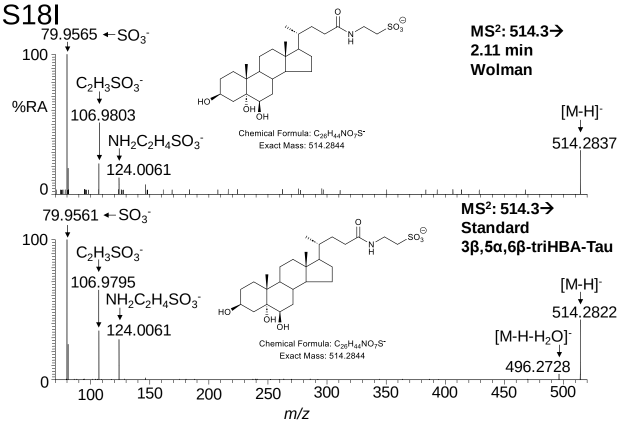
79.9565 ← $SO_3^-$
100
%RA
0
$C_2H_3SO_3^-$
106.9803
$NH_2C_2H_4SO_3^-$
124.0061
Chemical Formula: $C_{26}H_{44}NO_7S^-$
Exact Mass: 514.2844
MS²: 514.3→
2.11 min
Wolman
[M-H]⁻
514.2837
79.9561 ← $SO_3^-$
100
0
$C_2H_3SO_3^-$
106.9795
$NH_2C_2H_4SO_3^-$
124.0061
Chemical Formula: $C_{26}H_{44}NO_7S^-$
Exact Mass: 514.2844
MS²: 514.3→
Standard
3β,5α,6β-triHBA-Tau
[M-H]⁻
514.2822
$[M-H-H_2O]^-$
496.2728
100 150 200 250 300 350 400 450 500
m/z

S18J

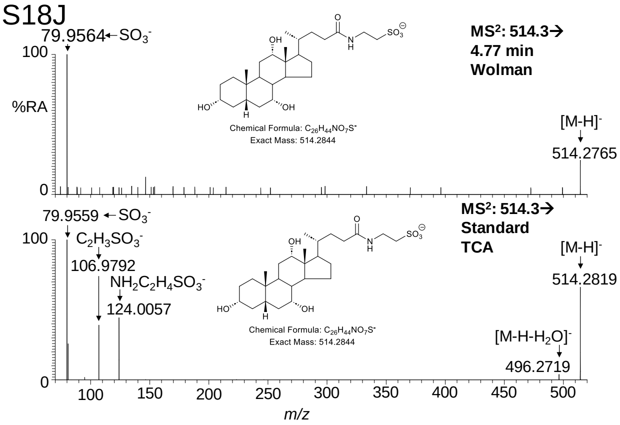
79.9564 ← $SO_3^-$
100
%RA
0
Chemical Formula: $C_{26}H_{44}NO_7S^-$
Exact Mass: 514.2844
MS²: 514.3→
4.77 min
Wolman
[M-H]⁻
514.2765
79.9559 ← $SO_3^-$
100
0
$C_2H_3SO_3^-$
106.9792
$NH_2C_2H_4SO_3^-$
124.0057
Chemical Formula: $C_{26}H_{44}NO_7S^-$
Exact Mass: 514.2844
MS²: 514.3→
Standard
TCA
[M-H]⁻
514.2819
$[M-H-H_2O]^-$
496.2719
100 150 200 250 300 350 400 450 500
m/z

S19A

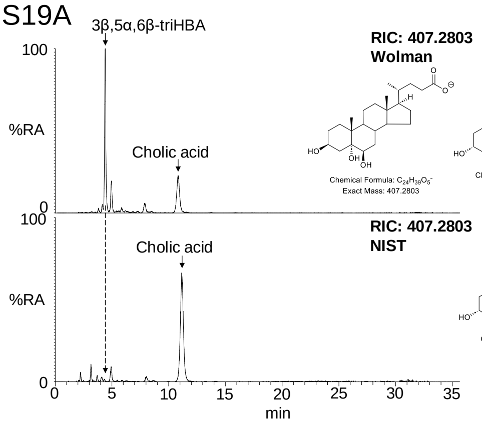


S19D

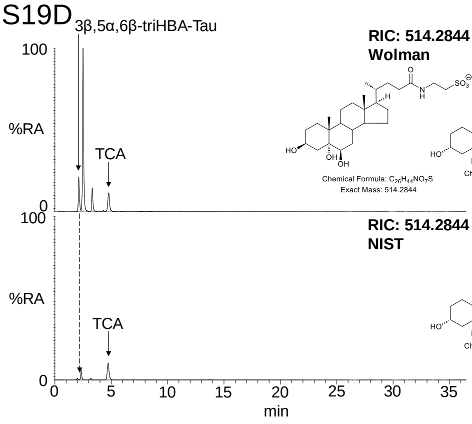


S19B

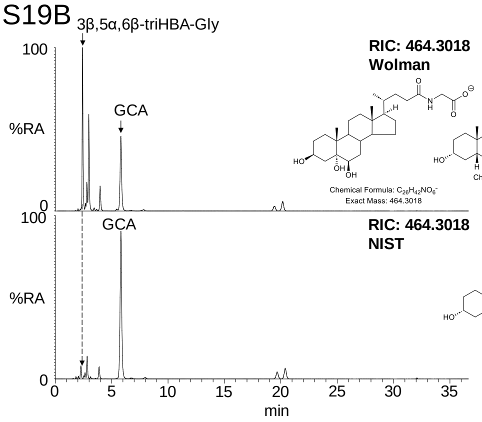


S19E

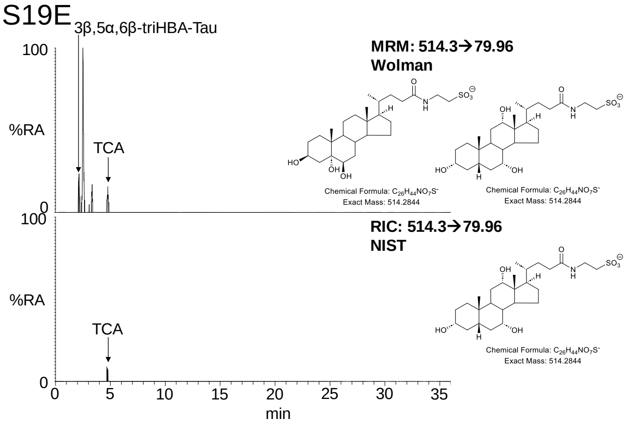


S19C

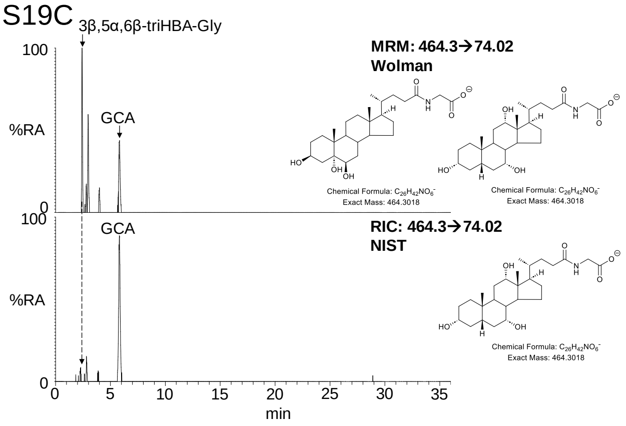

S20A

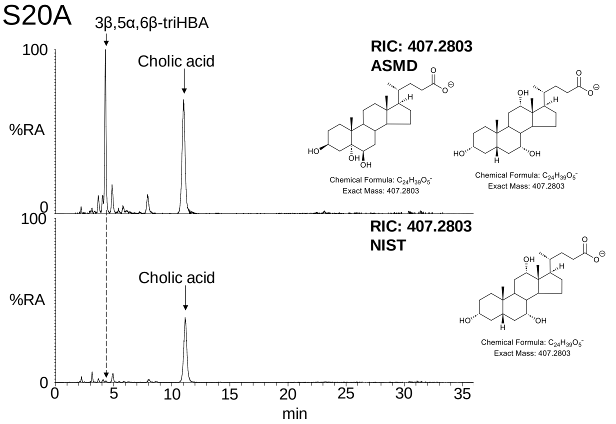


S20D

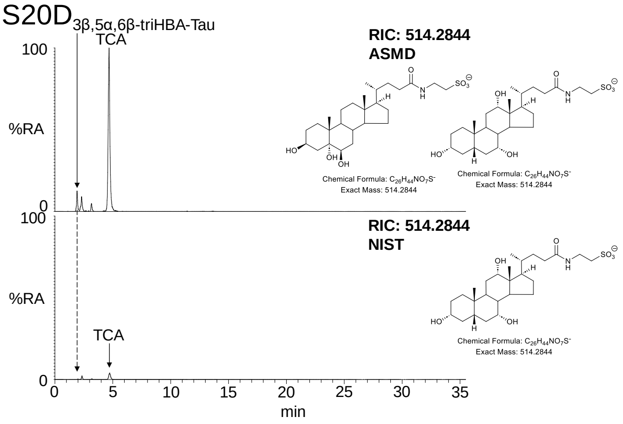


S20B

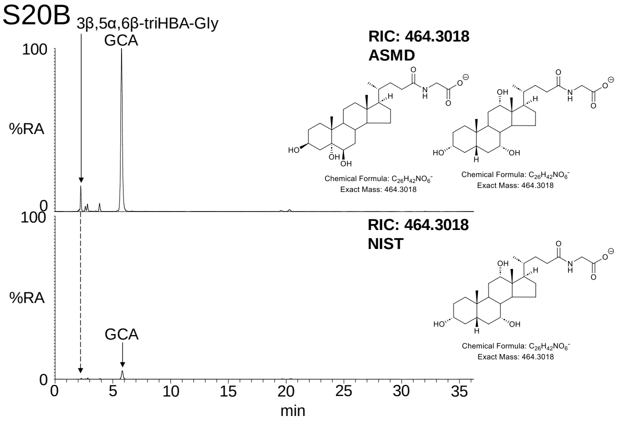


S20E

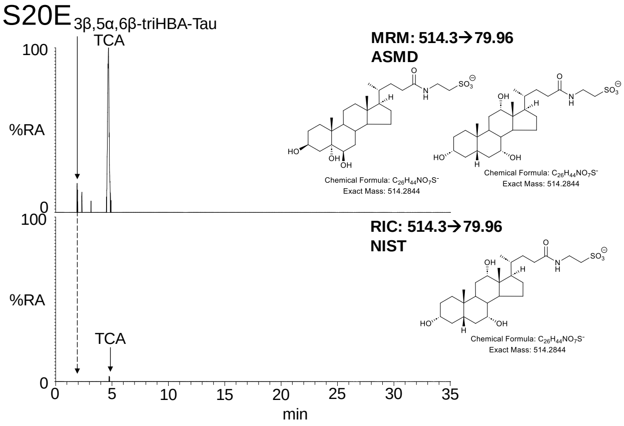


S20C

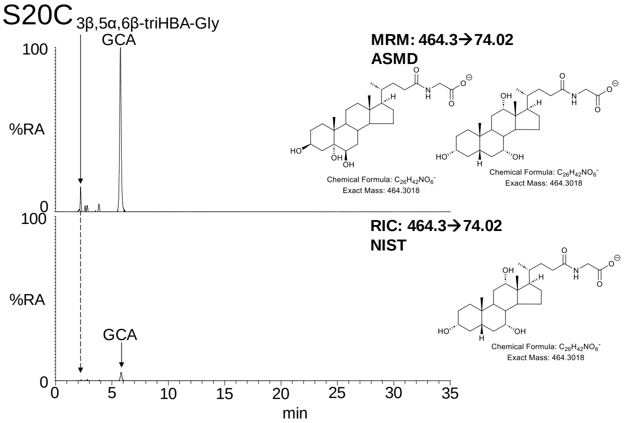

S21A(i)

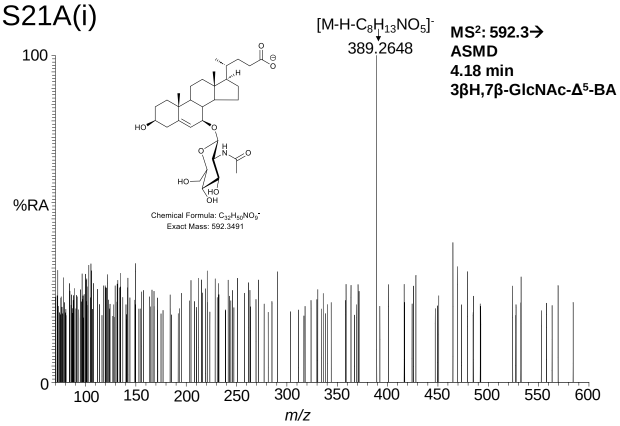


S21A(ii)

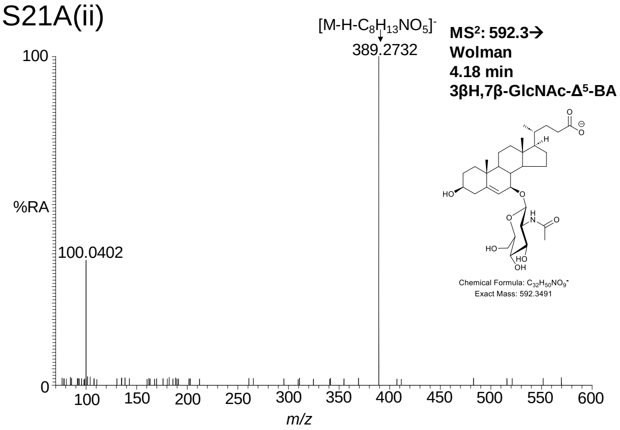


S21A(iii)

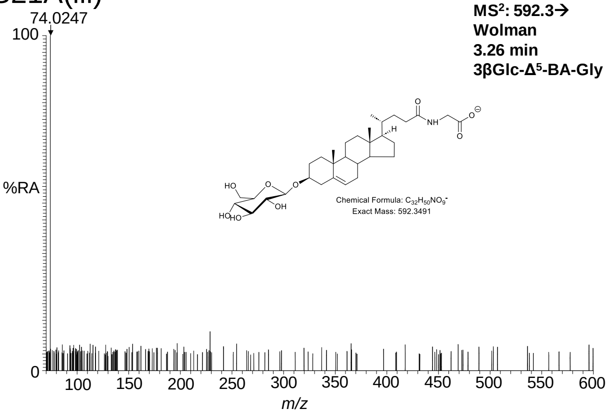


S21A(iv)

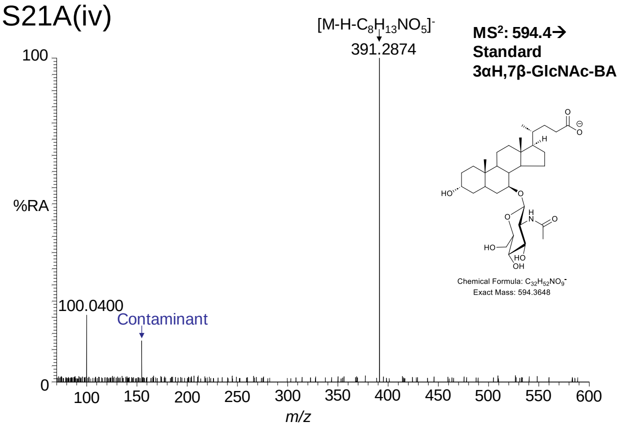

S21B(i)

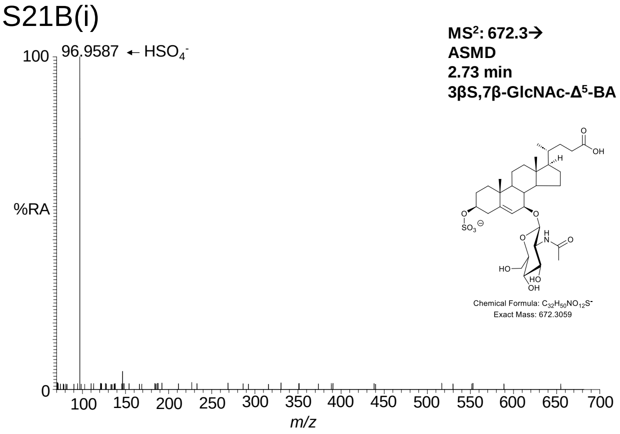


S21B(v)

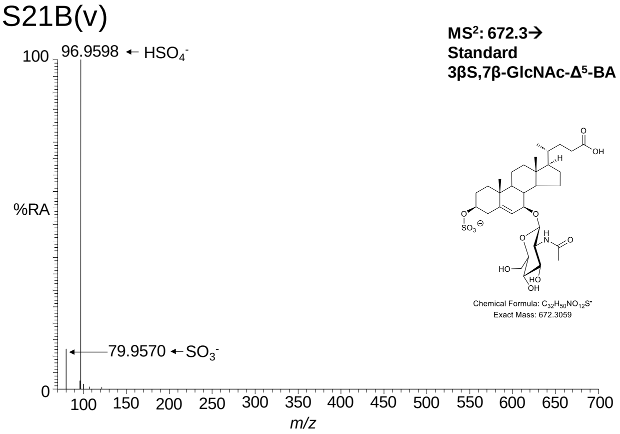


S21B(ii)

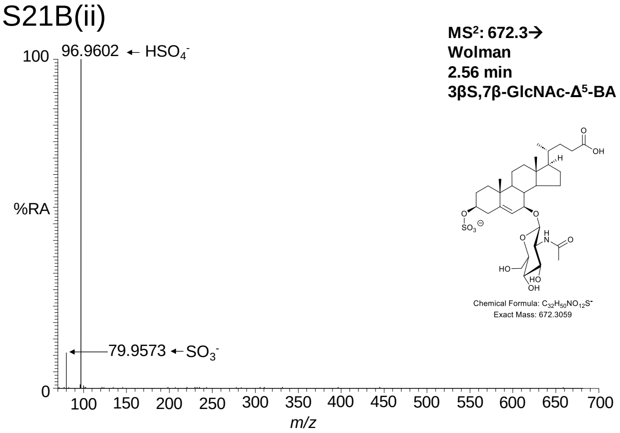


S21B(iii)

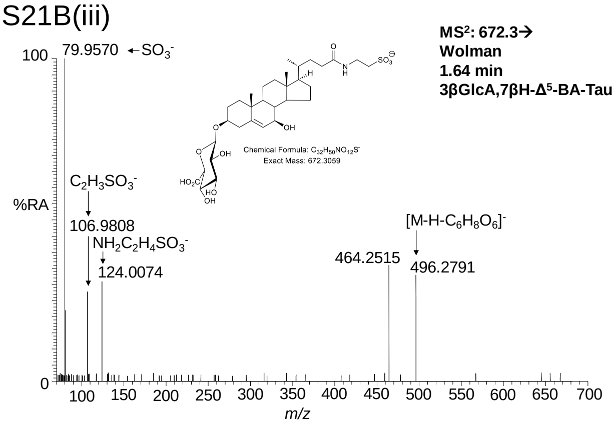


S21B(iv)

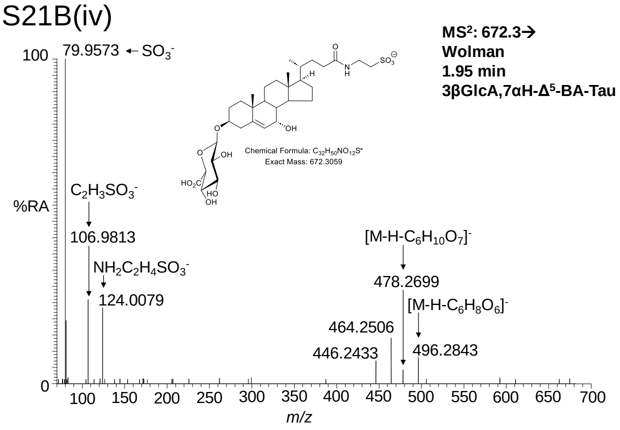

S21C(i)

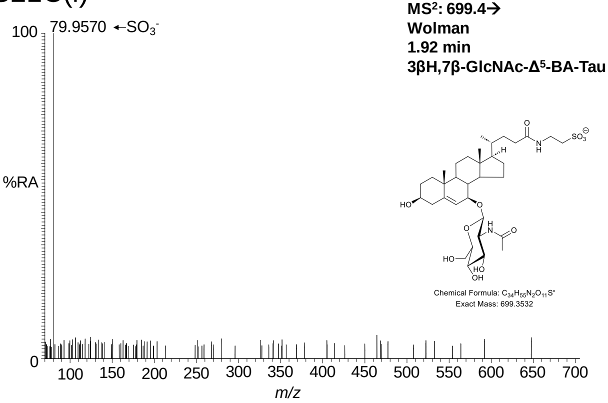


S21D(i)

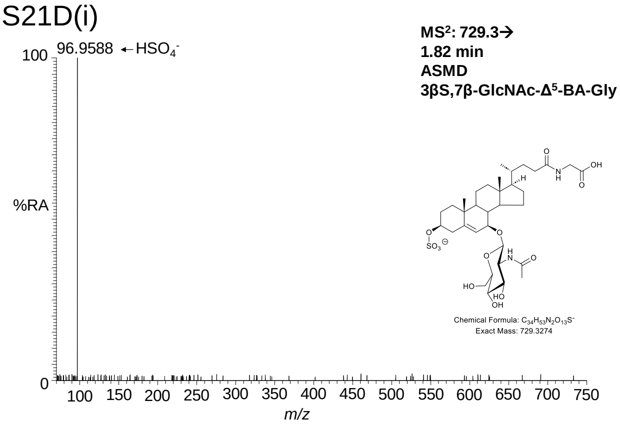


S21D(iv)

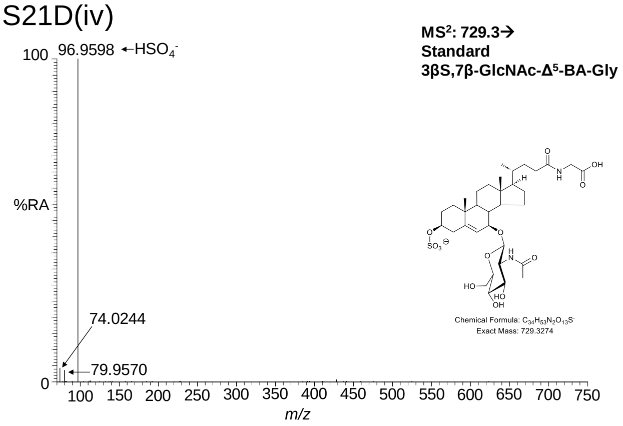


S21D(ii)

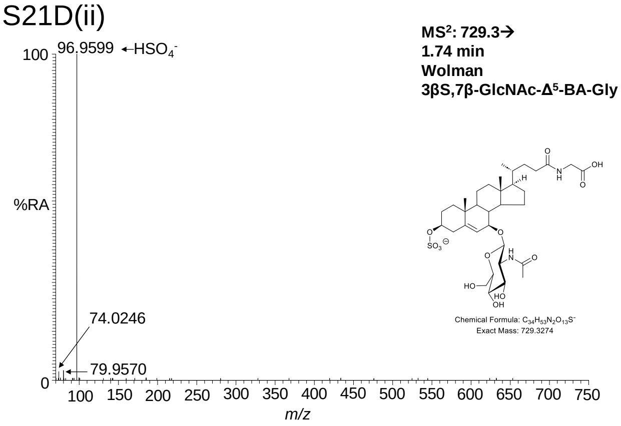


S21D(iii)

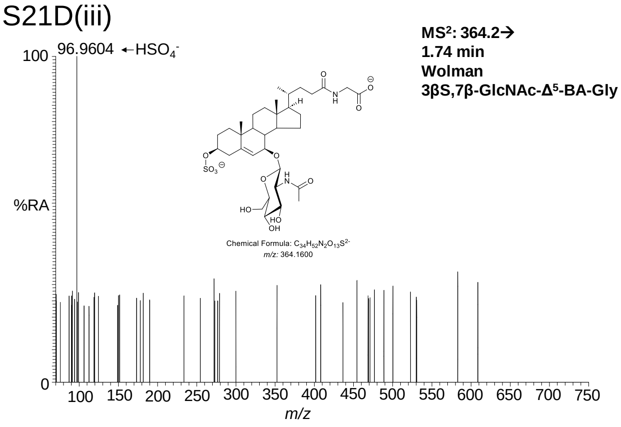

S21E(i)

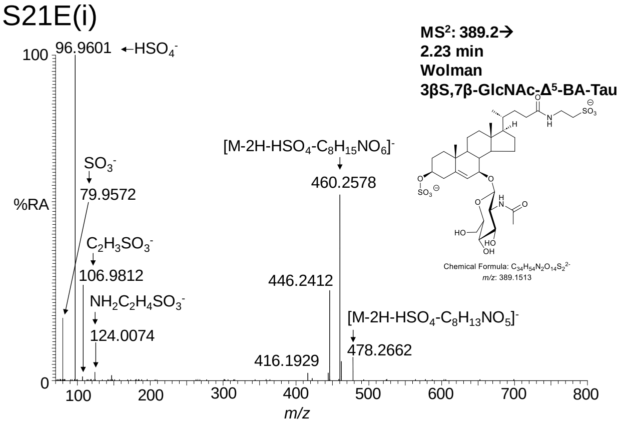


S21E(ii)

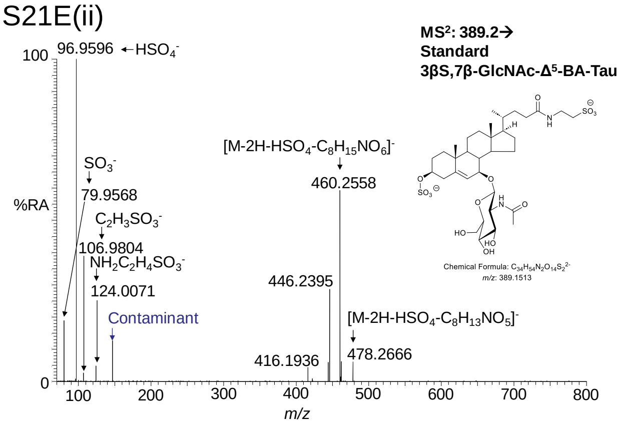

S22A

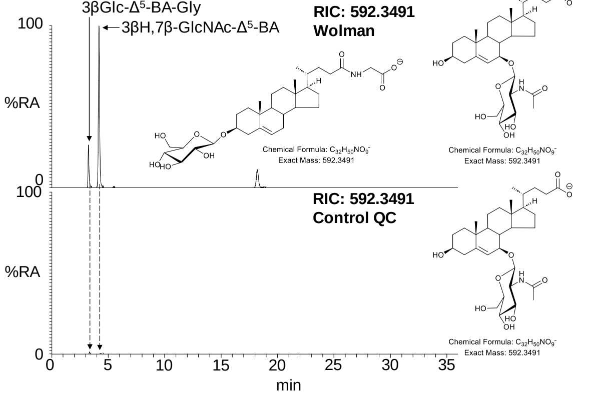


S22D

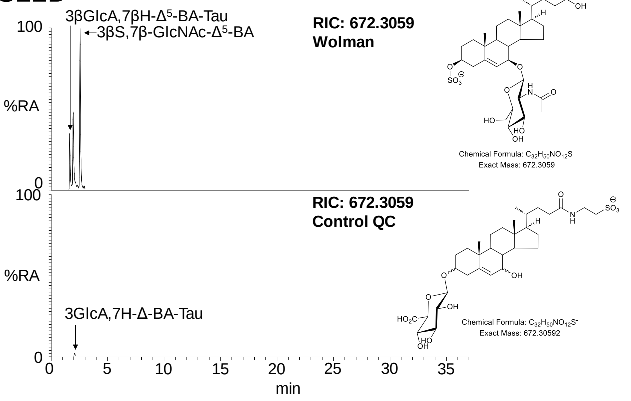


S22B

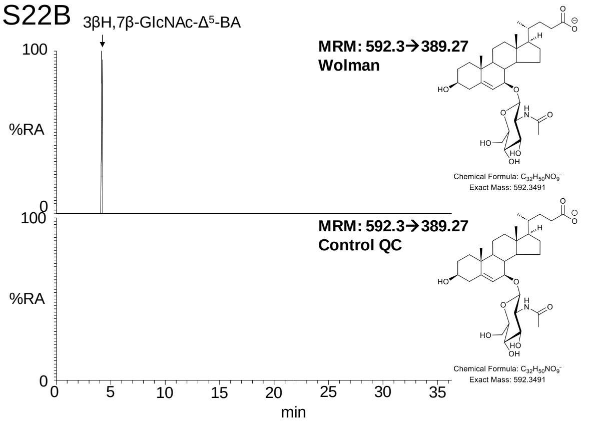


S22E

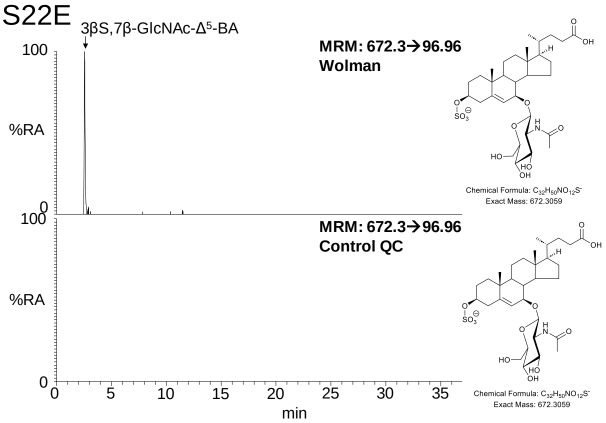


S22C

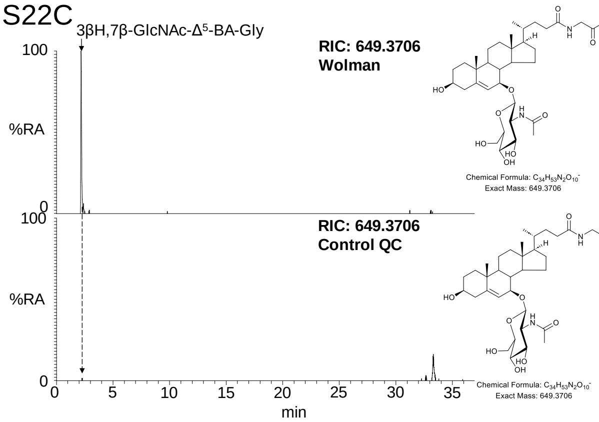


S22F

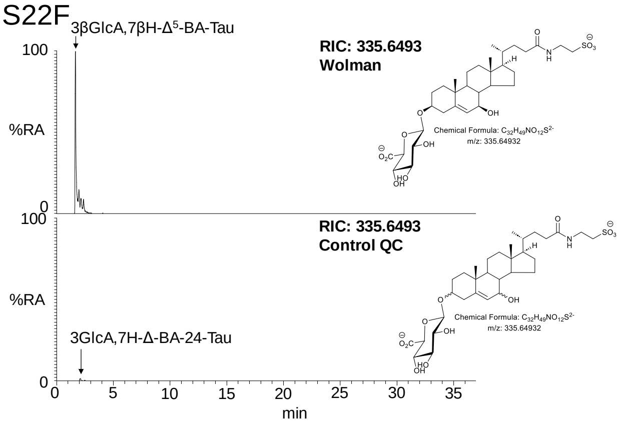


S22G

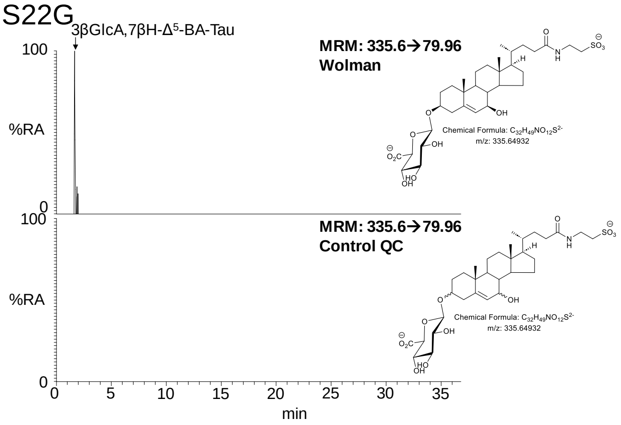

S22H

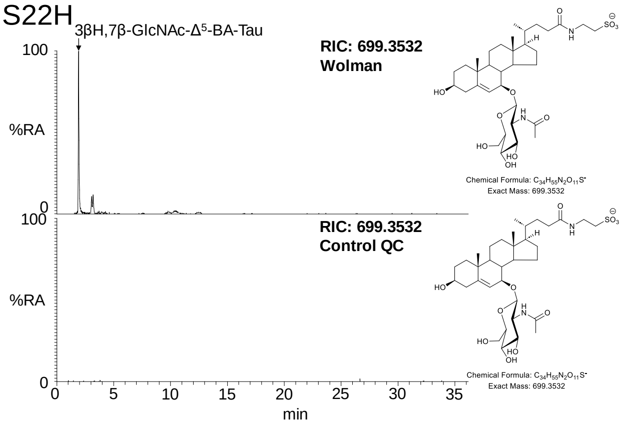


S22L

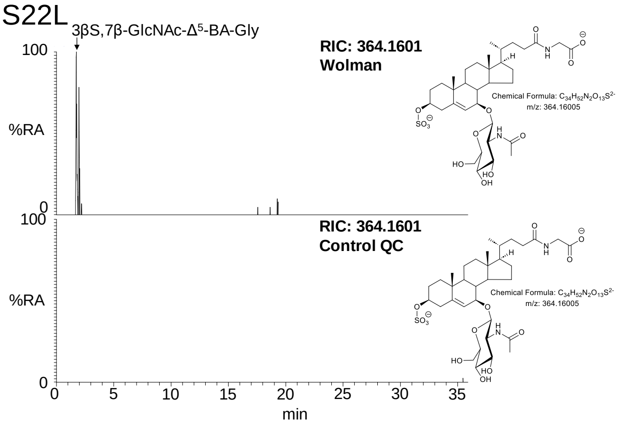


S22I

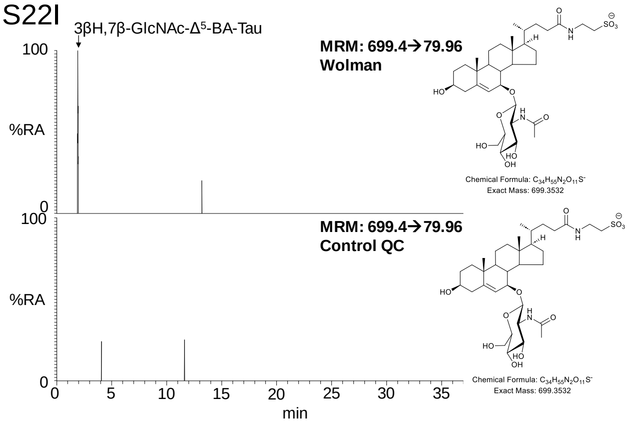


S22M

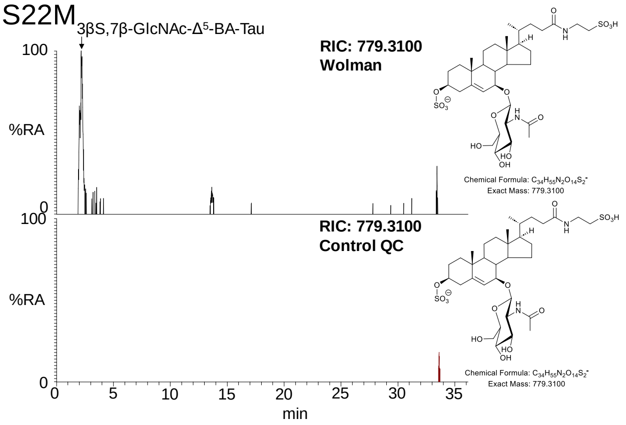


S22J

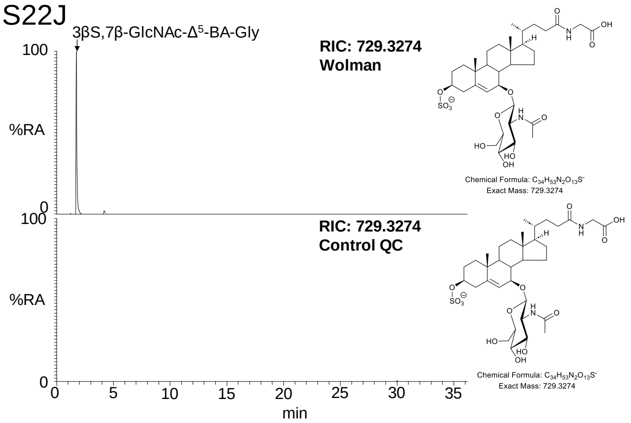


S22N

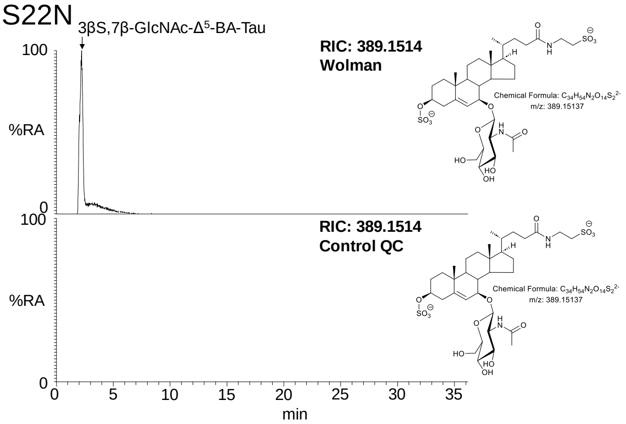


S22K

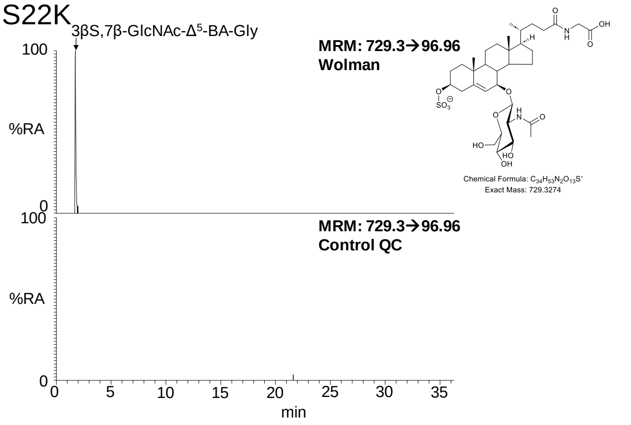


S22O

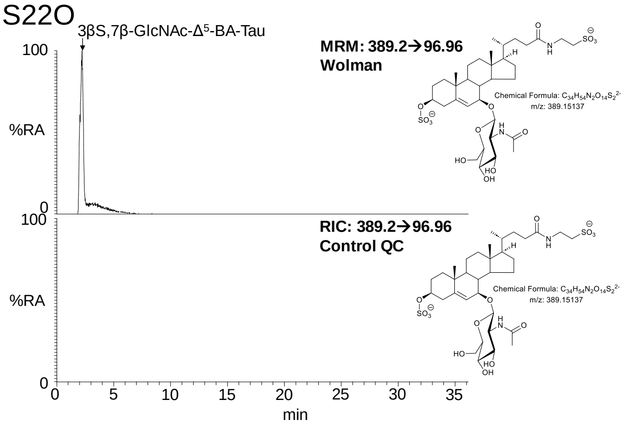

Table S1. Cholesterol Related Disorders, Dysregulated Metabolites and Mass Spectrometry Analysis

| Section | OMIM Disorder | Gene | OMIM Gene | Disorder (Short name, Abbreviation) | Changed (elevated or diminished) Sterol/Oxysterol/Bile acid Precursor/Bile acid Systematic Name (Common Name) | Changed (elevated or diminished) Sterol/Oxysterol/Bile acid Precursor/Bile acid Abbreviation (Other Abbreviation / Name) | Observed in Current Method, *m/z* | Formula of ion in (A)+, (B)+, (B)+, (B)- or (C)- | LipidMaps ID | Observed /predicted in Fraction, polarity | Predicted to be Observed in Current Method, *m/z* | Typical Concentration Ratio [Disease]/[Control] | Comment | Key Reference (origin of quantitative ratio data) |
|---|---|---|---|---|---|---|---|---|---|---|---|---|---|---|
| 3.1.3 | 302960 and 300960 | *EBP* | *300205* | Chondrodysplasia punctata 2 X-linked dominant (CDPX2) and Male EBP disorder with neurologic defects (MEND) | | | Positive-ion | | | | Positive-ion | | | |
| | | | | | Cholesta-5,8(9)-dien-3β-ol (8(9)-Dehydrocholesterol) | 8-DHC↑ | See prediction | [+]C34H45D5N3O | LMST01010242 | (A)+ | [$^2H_5$]GP 521.42624↑ | >500* | *Concentration of ratio of fetus, term birth or child to neonate. | (*Kelley 1999) |
| | | | | | 5α-Cholest-8-en-3β-ol (Zymostenol) | Zymostenol↑ | See prediction | [+]C34H47D5N3O | LMST01010096 | (A)+ | [$^2H_5$]GP 523.44189↑ | >2000* | May be difficult to resolve from much more abundant cholesterol by LC-MS. *Concentration of ratio of fetus, term birth or child to neonate. | (*Kelley 1999) |
| | | | | | | | Positive-ion | | | | Positive-ion | | | |
| 3.1.2 | 607330 | *SCD5* | *602286* | Lathosterolosis | Cholesta-5,8(9)-dien-3β-ol (8(9)-Dehydrocholesterol) | 8-DHC↓ | [$^2H_5$]GP 521.42624↓ | [+]C34H45D5N3O | LMST01010242 | (A)+ | | 0.4 - 0.05† | †Concentration ratio range for three probands to NIST SRM 1950. For two of the three probands concentrations below 2.5th percentile of adult controls. | Anderson 2019, (†Present work) |
| | | | | | Cholesta-5,24-dien-3β-ol (Desmosterol) | 24-DHC (Des)↓ | [$^2H_5$]GP 521.42624↓ | [+]C34H45D5N3O | LMST01010016 | (A)+ | | 0.15 - 0.05† | †Concentration ratio range for three probands to NIST SRM 1950. Absolute concentrations below 2.5th percentile of adult controls for all three probands. | Anderson 2019, (†Present work) |
| | | | | | 5α-Cholest-7-en-3β-ol (Lathosterol) | (Lath)↑ | See prediction | [+]C34H47D5N3O | LMST01010089 | (A)+ | [$^2H_5$]GP 523.44189↑ | >10* | Difficult to resolve lathosterol from much more abundant cholesterol by LC-MS. *Concentration ratio of child proband to maximum of control range. | (*Anderson 2019) |
| 3.1.1 | 270400 | *DHCR7* | *602858* | Smith-Lemli-Opitz syndrome (SLOS) | | | | | | | | | Positive-ion | |
| | | | | | Cholesta-5,7-dien-3β-ol (7-Dehydrocholesterol) | 7-DHC ↑ | [$^2H_5$]GP 521.42624↑ | [+]C34H45D5N3O | LMST01010069 | (A)+ | | 330‡ | ‡Mean concentration ratio, sum of 7-DHC and 8-DHC, adult controls. | Kelley 1995, (‡Present work) |
| | | | | | Cholesta-5,8(9)-dien-3β-ol (8(9)-Dehydrocholesterol) | 8-DHC ↑ | [$^2H_5$]GP 521.42624↑ | [+]C34H45D5N3O | LMST01010242 | (A)+ | | 330‡ | ‡Mean concentration ratio, sum of 7-DHC and 8-DHC, adult controls. | Liu 2014, (‡Present work) |
| | | | | | Cholest-5-en-3β-ol (Cholesterol) | (Chol)↓ | [$^2H_5$]GP 523.44189↓ | [+]C34H47D5N3O | LMST01010001 | (A)+ | | N/A | | Abdel-Khalik 2020 |
| | | | | | 3β,7β-Dihydroxychol-5-en-24-oic acid | 3β,7β-diH-Δ$^5$-BA↑ | [$^2H_5$]GP 527.36403↑ | [+]C31H39D5N3O4 | LMST04010280 | (A)+ | | 20‡ | ‡Mean concentration ratio, adult controls. | Abdel-Khalik 2020, (‡Present work) |
| | | | | | 3β-Hydroxycholest-5-en-7-one (7-Oxocholesterol or 7-Ketocholesterol) | 7-OC (7-KC)↑ | [$^2H_0$]GP 534.4054↑ | [+C34H52N3O2 | LMST01010049 | (B)+ | | 10‡ | ‡Mean concentration ratio, adult controls. | Abdel-Khalik 2020, (‡Present work) |
| | | | | | Cholest-5-ene-3β,7β-diol (7β-Hydroxycholesterol) | 7β-HC↑ | [$^2H_5$]GP 539.4368↑ | [+]C34H47D5N3O2 | LMST01010047 | (A)+ | | 15‡ | ‡Mean concentration ratio, adult controls. | Abdel-Khalik 2020, (‡Present work) |
| | | | | | 3β,7β-Dihydroxycholest-5-en-(25R/S)26-oic acid | 3β,7β-diHCA(25R/S)↑ | [$^2H_5$]GP 569.41098↑ | [+]C34H45D5N3O4 | LMST04030235 | (A)+ | | 15‡ | LipidMaps ID is for the (25R)26-epimer. ‡Mean concentration ratio, adult controls. | Abdel-Khalik 2020, (‡Present work) |
| 3.1.3 | 602398 | *DHCR24* | *606418* | Desmosterolosis | | | Positive-ion | | | | Positive-ion | | | |
| | | | | | Cholesta-5,24-dien-3β-ol (Desmosterol) | 24-DHC (Des)↑ | See prediction | [+]C34H45D5N3O | LMST01010016 | (A)+ | [$^2H_5$]GP 521.42624↑ | >100* | | Clayton 1996, (*Andersson 2002) |
| 3.2.2 | | *CYP7A1* | *118455* | Cholesterol 7α-hydroxylase (CYP7A1) deficiency | | | Positive-ion | | | | Positive-ion | | | |
| | | | | | 7α-Hydroxycholest-4-en-3-one | 7α-HCO (C4)↓ | See prediction | [+]C34H52N3O2 | LMST04030123 | (B)+ | [$^2H_0$]GP 534.4054↓ | NA | Also observed in (A)+ at 539.4368 in combination with 7α-HC. | Pullinger 2002, Present work |
| | | | | | Cholest-5-en-3β-ol (7α-Hydroxycholesterol) | 7α-HC ↓ | See prediction | [+]C34H47D5N3O2 | LMST01010013 | (A+) | [$^2H_5$]GP 539.4368↓ | NA | In (A)+ observed in combination with 7αHCO. | Pullinger 2002, Present work |
| 3.2.1 | 213700 | *CYP27A1* | *606530* | Cerebrotendinous xanthomatosis (CTX) | | | Positive-ion | | | | | | | |
| | | | | | 7α-Hydroxy-3-oxo-chol-4-en-24-oic acid | 7αH,3O-Δ$^4$-BA↓ | [$^2H_0$]GP 522.33263↓ | [+]C31H44N3O4 | LMST04010216 | (B)+ | | 0.03‡ | Also observed in (A)+ at 527.3640 in combination with 3β,7α-diH-Δ$^5$-BA. ‡Mean concentration ratio for CTX against adult controls. | Crick Clin Chem 2015, (‡Present work) |
| | | | | | 5α-cholestan-3β-ol (Cholestanol) | Cholestanol↑ | [$^2H_5$]GP 525.45754↑ | [+]C34H49D5N3O | LMST01010077 | (A)+ | | 10* | Can be difficult to resolve from the M+2 isotopic peak of cholesteol by LC-MS. *Typical CTX to normal (Oregon Health & Science University laboratory) ratio. | Salen 1971, (*deBarber 2024) |
| | | | | | 7α-Hydroxycholest-4-en-3-one | 7α-HCO↑ | [$^2H_0$]GP 534.4054↑ | [+]C34H52N3O2 | LMST04030123 | (B)+ | | 25‡- 180* | Also observed in (A)+ at 539.4368 in combination with 7α-HC. ‡Mean concentration ratio for CTX against adult controls. *Ratio of typical CTX to normal adult. | deBarber 2010, (*deBarber 2024), (‡Present work) |
| | | | | | Cholest-5-ene-3β(25R)26-diol (26-Hydroxycholesterol or 27-hydroxycholesterol) | 26-HC↓ | [$^2H_5$]GP 539.4368↓ | [+]C34H47D5N3O2 | LMST01010088 | (A)+ | | 0.05‡ | 26-HC also called 27-HC. ‡Mean concentration ratio for CTX against adult controls. | Griffiths 2013, (‡Present work) |
| | | | | | 7α,12α-Dihydroxycholest-4-en-3-one | 7α,12α-diHCO↑ | [$^2H_0$]GP 550.40032↑ | [+]C34H52N3O3 | LMST04030114 | (B)+ | | >150* - 5000‡ | Also observed in (A)+ at 555.4317 in combination with 7α,12α-diHC. ‡Mean concentration ratio for CTX against adult controls. *Ratio of typical CTX to normal adult. | deBarber 2010, (*deBarber 2024), (‡Present work) |
| | | | | | 3β-Hydroxycholest-5-en-(25R)26-oic acid (Cholrestenoic acid) | 3β-HCA↓ | [$^2H_5$]GP 553.41607↓ | [+]C34H45D5N3O3 | LMST04030072 | (A)+ | | 0.01‡ | ‡Mean concentration ratio for CTX against adult controls. | Griffiths 2013, (‡Present work) |
| | | | | | 7α-Hydroxy-3-oxocholest-4-en-(25R/S)26-oic acid | 7αH,3O-CA(25R/S) (HOCA)↓ | [$^2H_0$]GP 564.37958↓ | [+]C34H50N3O4 | LMST04030242 | (B)+ | | 0.02‡ | Also observed in (A)+ at 569.4110 in combination with 3β,7α-diHCA(25R/S). LipidMaps ID is for the (25R)26-epimer. ‡Mean concentration ratio for CTX against adult controls. | Griffiths 2013. (‡Present work) |
| | | | | | 3β,7β-Dihydroxycholest-5-en-(25R/S)26-oic acid | 3β,7β-diHCA(25R/S)↓ | [$^2H_5$]GP 569.4110↓ | [+]C34H45D5N3O4 | LMST04030235 | (A)+ | | 0.02‡ | LipidMaps ID is for the (25R)26-epimer. ‡Mean concentration ratio for CTX against adult controls. | Crick 2015, (‡Present work) |
| | | | | | | | Negative-ion | | | | | | | |
| | | | | | 3α,7α-Dihydroxy-5β-cholan-24-oic acid (Chenodeoxycholic acid) | 3α,7α-diHBA (CDCA)↓ | [-]391.2854↓ | [-]C24H39O4 | LMST04010032 | (B)- or (C)- | | <0.05† | †Ratio for CTX against NIST SRM 1950. | Salen 1971, (†Present work) |
| | | | | | 3α,7α-Dihydroxy-5β-cholan-24-oyl-glycine (Glycochenodeoxycholic acid) | 3α,7α-diHBA-24-Gly (GCDCA)↓ | [-]448.3068↓ | [-]C26H42NO5 | LMST05030008 | (C)- | | 0.05† | †Ratio for CTX against NIST SRM 1950. | Salen 1971, (†Present work) |
| | | | | | 3α,7α-Dihydroxy-5β-cholan-24-oyl-taurine (Taurochenodeoxycholic acid) | 3α,7α-diHBA-24-Tau (TCDCA)↓ | [-]498.2895↓ | [-]C26H44NO6S | LMST05040005 | (C)- | | NA | Taurine conjugates are lost in fractions (A) and (B) following GP derivatisation. | Salen 1971 |
| | | | | | Trihydroxy-5β-cholestan-26-oyl-taurine | triHCa-26-Tau↓ | [-]556.3313↓ | [-]C29H50NO7S | NA | (C)- | | NA | Taurine conjugates are lost in fractions (A) and (B) following GP derivatisation. | Vaz 2023 |
| | | | | | 7α,12α,25-Trihydroxy-5β-cholestane-3α-*O*-β-D-glucuronide (5β-Cholestane-3α,7α,12α,25-tetrol-3-*O*-β-D-glucuronide) | 7α,12α,25-triHc-3αGlcA (3α,7α,12α,25-tetrol-3αGlcA)↑ | [-]611.3801↑ | [-]C33H55O10 | LMST05010042 | (B)- or (C)- | | >30* | *Ratio for typical CTX to unaffected dried blood spot. | Egestad 1985, deBarber 2018, (*deBarber 2024) |
| | | | | | Cholestanetetrol glucuronide | tetrol-GlcA↑ | [-]611.3801↑ | [-]C33H55O10 | NA | (B)- or (C)- | | >100† | †Ratio for CTX against NIST SRM 1950. | Egestad 1985, (†Present work) |
| | | | | | Cholestanepentol glucuronide | pentol-GlcA↑ | [-]627.375↑ | [-]C33H55O11 | NA | (B)- or (C)- | | >20† | †Ratio for CTX against NIST SRM 1950. | Egestad 1985, (†Present work) |
| | | | | | Cholestanehexol glucuronide | hexol-GlcA↑ | [-]643.3699↑ | [-]C33H55O12- | NA | (B)- or (C)- | | >50† | †Ratio for CTX against NIST SRM 1950. | Egestad 1985, (†Present work) |
| 3.2.3 | 270800 and 613812 | *CYP7B1* | *603711* | Spastic paraplegia 5A (SPG5A) and Oxysterol 7α-hydroxylase deficiency (O7AHD) | | | Positive-ion | | | | | | | |
| | | | | | Cholest-5-ene-3β(25R)26-diol (26-Hydroxycholesterol or 27-hydroxycholesterol) | 26-HC↑ | [$^2H_5$]GP 539.4368↑ | [+]C34H47D5N3O2 | LMST01010088 | (A)+ | | 5[‡S], 60[‡O] | 26-HC also called 27-HC. [‡S]Mean concentration ratio of SPG5 adults to adult controls. [‡O]Mean concentration ratio of O7AHD infants to adult controls. | Setchell 1998, Schüle 2010, ([‡O]Theofilopoulos 2014), (‡Present work) |
| | | | | | Cholest-5-en-3β,25-diol (25-Hydroxycholesterol) | 25-HC↑ | [$^2H_5$]GP 539.4368↑ | [+]C34H47D5N3O2 | LMST01010018 | (A)+ | | 35[‡S], 260[‡O] | [‡S]Mean concentration ratio of SPG5 adults to adult controls. [‡O]Mean concentration ratio of O7AHD infants to adult controls. | Setchell 1998, Schüle 2010, ([‡O]Theofilopoulos 2014), (‡Present work) |
| | | | | | 3β-Hydroxycholest-5-en-(25R)26-oic acid (Cholrestenoic acid) | 3β-HCA↑ | [$^2H_5$]GP 553.41607↑ | [+]C34H45D5N3O3 | LMST04030072 | (A)+ | | 5[‡S], 35[‡O] | [‡S]Mean concentration ratio of SPG5 adults to adult controls. [‡O]Mean concentration ratio of O7AHD infants to adult controls. | Setchell 1998, ([‡O]Theofilopoulos 2014), (‡Present work) |
| | | | | | 3β-Hydroxychol-5-en-24-oic acid | 3βH-Δ$^5$-BA↑ | [$^2H_5$]GP 511.3691↑ | [+]C31H39D5N3O3 | LMST04010201 | (A)+ | | 25[S], 215[O] | [S]Mean concentration ratio of SPG5 adults to adult controls. [O]Mean concentration ratio of O7AHD infants to adult controls. | Setchell 1998, ([O,S]Theofilopoulos 2014) |
| | | | | | | | Negative-ion | | | | | | | |
| | | | | | 3β-Sulfooxychol-5-en-24-oic acid | 3βS-Δ$^5$-BA↑ | [-]453.2316↑ | [-]C24H37O6S | NA | (C)- | | 5[‡S] | Sulfuric acid conjugates are lost in fractions (A) and (B) following GP derivatisation. [‡S]Ratio of SPG5A adult to NIST SRM 1950. | Setchell 1998, ([‡S]Present work) |
| | | | | | 3β-Hydroxychol-5-en-24-oyl-taurine | 3βH-Δ$^5$-BA-24-Tau↑ | [-]480.2789↑ | [-]C26H42NO5S | NA | (C)- | | 5[‡S] | Taurine conjugates are lost in fractions (A) and (B) following GP derivatisation. [‡S]Ratio of SPG5A adult to NIST SRM 1950. | Setchell 1998, ([‡S]Present work) |
| | | | | | 3β-Sulfooxychol-5-en-24-oyl-glycine | 3βS-Δ$^5$-BA-24-Gly↑ | [-]510.2531↑ | [-]C26H40NO7S | NA | (C)- | | 3 - 10[‡S] | Sulfuric acid conjugates are lost in fractions (A) and (B) following GP derivatisation. [‡S]Ratio of SPG5A adult to NIST SRM 1950. | Setchell 1998, ([‡S]Present work) |
| 3.2.4. | 607765 | *HSD3B7* | *607764* | 3β-Hydroxy-Δ$^5$-C$_{27}$-steroid oxidoreductase (HSD3B7) deficiency (3β-Dehydrogenase deficiency) | | | Positive-ion | | | | | | | |
| | | | | | 7α-Hydroxy-3-oxochol-4-en-24-oic acid | 7αH,3O-Δ$^4$-BA↓ | [$^2H_0$]GP 522.33263↓ | [+]C31H44N3O4 | LMST04010216 | (B)+ | | NA | Also observed in (A)+ at 527.3640 in combination with 3β,7α-diH-Δ$^5$-BA. | Griffiths 2011 |
| | | | | | 7α-Hydroxy-3-oxocholest-4-en-(25R/S)26-oic acid | 7αH,3O-CA(25R/S)↓ | [$^2H_0$]GP 564.37958↓ | [+]C34H50N3O4 | LMST04030242 | (B)+ | | 0.1$ | Also observed in (A)+ at 569.4110 in combination with 3β,7α-diHCA(25R/S). LipidMaps ID is for the (25R)26-epimer. $Concentration of 7αH,3O-CA(25R/S) normalised to 3β,7α-diHCA(25R/S) and compared to NIST SRM 1950. | Griffiths 2011, ($Present work) |
| | | | | | | | Negative-ion | | | | | | | |
| | | | | | 3β,7α-Dihydroxychol-5-en-24-oyl-glycine | 3β,7α-diH-Δ$^5$-BA-24-Gly↑ | [-]446.2912↑ | [-]C26H40NO5 | NA | (C)- | | NA | | Mazzacuva 2016 |
| | | | | | 3β,7α,12α-Trihydroxychol-5-en-24-oyl-glycine | 3β,7α,12α-triH-Δ$^5$-BA-24-Gly↑ | [-]462.2861↑ | [-]C26H40NO6 | NA | (C)- | | NA | | Mazzacuva 2016 |
| | | | | | 3β-Sulfooxy-7α-hydroxychol-5-en-24-oic acid | 3βS,7αH-Δ$^5$-BA↑ | [-]469.2265↑ | [-]C24H37O7S | NA | (C)- | | NA | Sulfuric acid conjugates are lost in fractions (A) and (B) following GP derivatisation. | Ichimiya 1991, Mazzacuva 2016 |
| | | | | | 3β-Sulfooxy-7α,12α-dihydroxychol-5-en-24-oic acid | 3βS,7α,12α-diH-Δ$^5$-BA↑ | [-]485.2215↑ | [-]C24H37O8S | NA | (C)- | | NA | Sulfuric acid conjugates are lost in fractions (A) and (B) following GP derivatisation. | Ichimiya 1991, Mazzacuva 2016 |
| | | | | | 3β-Sulfooxy-7α-hydroxychol-5-en-24-oyl-glycine | 3βS,7αH-Δ$^5$-BA-24-Gly↑ | [-]526.2480↑ | [-]C26H40NO8S | NA | (C)- | | NA | Sulfuric acid conjugates are lost in fractions (A) and (B) following GP derivatisation. | Ichimiya 1991, Mazzacuva 2016 |
| | | | | | 3β-Sulfooxy-7α,12α-dihydroxychol-5-en-24-oyl-glycine | 3βS,7α,12α-diH-Δ$^5$-BA-Gly↑ | [-]542.2429↑ | [-]C26H40NO9S | NA | (C)- | | NA | Sulfuric acid conjugates are lost in fractions (A) and (B) following GP derivatisation. | Ichimiya 1991, Mazzacuva 2016 |
| | | | | | Cholest-5-ene-3β,7α-disulfate (7α-Hydroxycholesterol 3β,7α-disulfate) | C-3β,7α-diS↑** | [-]561.2561↑** | [-]C27H45O8S2 | NA | (C)- | | NA | **Mixture of [M-H]$^-$ and [M-2H]$^{2-}$ ions produced dependent on source parameters. Sulfuric acid conjugates are lost in fractions (A) and (B) following GP derivatisation. | Griffiths 2011 |
| | | | | | | C-3β,7α-diS↑** | [2-]280.12445↑** | [2-]C27H44O8S2 | NA | (C)- | | NA | **Mixture of [M-H]$^-$ and [M-2H]$^{2-}$ ions produced dependent on source parameters. Sulfuric acid conjugates are lost in fractions (A) and (B) following GP derivatisation. | Griffiths 2011 |
| | | | | | x-Hydroxycholest-5-en-3β,y-disulfate (x,y-Dihydroxycholesterol 3β,y-disulfate) | x-HC-3β,y-diS↑** | [-]577.251↑** | [-]C27H45O9S2 | NA | (C)- | | NA | x and y indicate position of functional group not known. **Mixture of [M-H]- and [M-2H]2- ions produced dependent on source parameters. Sulfuric acid conjugates are lost in fractions (A) and (B) following GP derivatisation. | Ichimiya 1991 |
| | | | | | | x-HC-3β,y-diS↑** | [2-]288.1219↑** | [2-]C27H44O9S2 | NA | (C)- | | NA | x and y indicate position of functional group not known. **Mixture of [M-H]- and [M-2H]2- ions produced dependent on source parameters. Sulfuric acid conjugates are lost in fractions (A) and (B) following GP derivatisation. | Ichimiya 1991 |
| | | | | | 3β-Hydroxy-7-*N*-acetylglucosaminyl-chol-5-en-24-oic acid | 3βH,7-GlcNAc-Δ$^5$-BA↑$ | [-]592.3491↑ | [-]C32H50NO9 | NA | (B)- or (C)- | | NA | $GlcNAc conjugates not found in all patient samples. | Fischler 2007, Mazzacuva 2016 |
| | | | | | x,y-Dihydroxycholest-5-en-3β,z-disulfate (x,y,z-Trihydroxycholesterol 3β,z-disulfate) | x,y-diHC,3β,z-diS↑** | [-]593.246↑** | [-]C27H45O10S2 | NA | (C)- | | NA | x, y and ¶ indicate position of functional group not known. **Mixture of [M-H]- and [M-2H]2- ions produced dependent on source parameters. Sulfuric acid conjugates are lost in fractions (A) and (B) following GP derivatisation. | Meng 1997 |
| | | | | | | x,y-diHC,3β,z-diS↑** | [2-]296.11935↑** | [2-]C27H44O10S2 | NA | (C)- | | NA | x, y and ¶ indicate position of functional group not known. **Mixture of [M-H]- and [M-2H]2- ions produced dependent on source parameters. Sulfuric acid conjugates are lost in fractions (A) and (B) following GP derivatisation. | Meng 1997 |
| | | | | | 7α-Glucuronylchol-5-en-3β-sulfate (7α-Hydroxycholesterol 3β-sulfate,7α-glucuronide) | 7α-GlcA-C-3βS↑** | [-]657.3314↑** | [-]C33H53O11S | NA | (C)- | | NA | *Mixture of [M-H]$^-$ and [M-2H]$^{2-}$ ions produced dependent on source parameters. Sulfuric acid conjugates are lost in fractions (A) and (B) following GP derivatisation. | Meng 1997 |
| | | | | | | 7α-GlcA-C-3βS↑** | [2-]328.16205↑** | [2-]C33H52O11S | NA | (C)- | | NA | *Mixture of [M-H]$^-$ and [M-2H]$^{2-}$ ions produced dependent on source parameters. Sulfuric acid conjugates are lost in fractions (A) and (B) following GP derivatisation. | Meng 1997 |
| | | | | | x-Hydroxy-y-glucuronylcholest-5-en-3β-sulfate (x,y-Dihydroxycholesterol 3β-sulfate-x-glucuronide) | x-GlcA,y-HC-3βS↑** | [-]673.3263↑** | [-]C33H53O12S | NA | (C)- | | NA | x and y indicate position of functional group not known. **Mixture of [M-H]- and [M-2H]2- ions produced dependent on source parameters. Sulfuric acid conjugates are lost in fractions (A) and (B) following GP derivatisation. | Meng 1997 |
| | | | | | | x-GlcA,y-HC-3βS↑** | [2-]336.1595↑** | [2-]C33H52O12S | NA | (C)- | | NA | x and y indicate position of functional group not known. **Mixture of [M-H]- and [M-2H]2- ions produced dependent on source parameters. Sulfuric acid conjugates are lost in fractions (A) and (B) following GP derivatisation. | Meng 1997 |
| | | | | | x,y-Dihydroxy-z-glucuronylcholest-5-en-3β-sulfate (x,y,z-Trihydroxycholesterol 3β-sulfate-x-glucuronide) | x-GlcA,y,z-diHC-3βS | [-]689.3212↑** | [-]C33H53O13S | NA | (C)- | | NA | x, y and z indicate position of functional group not known. **Mixture of [M-H]- and [M-2H]2- ions produced dependent on source parameters. Sulfuric acid conjugates are lost in fractions (A) and (B) following GP derivatisation. | Meng 1997 |
| | | | | | | x-GlcA,y,z-diHC-3βS | [2-]344.157↑** | [2-]C27H45O10S | NA | (C)- | | NA | x, y and z indicate position of functional group not known. **Mixture of [M-H]- and [M-2H]2- ions produced dependent on source parameters. Sulfuric acid conjugates are lost in fractions (A) and (B) following GP derivatisation. | Meng 1997 |
| 3.2.5. | 235555 | *AKR1D1* | *604741* | Δ$^4$-3-Oxosteroid-5β-reductase (AKR1D1) deficiency (5β-Reductase deficiency) | | | Positive-ion | | | | Positive-ion | | | |
| | | | | | 7α-Hydroxy-3-oxochol-4-en-24-oic acid | 7αH,3O-Δ$^4$-BA↑ | See prediction | [+]C31H44N3O4 | LMST04010216 | (B)+ | [$^2H_0$]GP 522.33263↑ | 70ʃ | Also observed in (A)+ at 527.3640 in combination with 3β,7α-diH-Δ$^5$-BA. ʃValue determined following enzymatic deconjugation. | Clayton 1996 (ʃLemonde 2003) |
| | | | | | 7α-Hydroxy-27-*nor*-cholest-4-en-3,24-dione | 7αH-27-*nor*-C-3,24-diO↑ | See prediction | [+]C33H48N3O3 | NA | (B)+ | [$^2H_0$]GP 534.36902↑ | NA | Also observed in (A)+ at 539.4004 in combination with 3β,7α-diH-27-*nor*-C-24O. Both 7αH-27-*nor*-C-3,24-diO and 3β,7α-diH-27-*nor*-C-24O are decarboxylation products of 7αH,3,24-diO-CA(25R) and 3β,7α-diH,24O-CA(25), respectively. | Present work |
| | | | | | 7α,12α-Dihydroxy-3-oxochol-4-en-24-oic acid | 7α,12α-diH,3O-Δ$^4$-BA↑ | See prediction | [+]C31H44N3O5 | LMST04010241 | (B)+ | [$^2H_0$]GP 538.32755↑ | 80ʃ | Also observed in (A)+ at 543.3589 in combination with 3β,7α,12α-triH-Δ$^5$-BA. ʃValue determined following enzymatic deconjugation. | (ʃLemonde 2003) |
| | | | | | 7α-Hydroxy-3-oxocholest-4-en-(25R/S)26-oic acid | 7αH,3O-CA(25R/S)↑ | See prediction | [+]C34H50N3O4 | LMST04030242 | (B)+ | [$^2H_0$]GP 564.37958↑ | NA | Also observed in (A)+ at 569.4110 in combination with 3β,7α-diHCA. LipidMaps ID is for the (25R)26-epimer. | Present work |
| | | | | | 7α-Hydroxy-3,24-bisoxocholest-4-en-(25R)26-oic acid | 7αH,3,24-diO-CA(25R)↑ | See prediction | [+] C34H48N3O5 | LMST04030255 | (B)+ | [$^2H_0$]GP 578.35885↑ | NA | Also observed in (A)+ at 583.3902 in combination with 3β,7α-diH,24O-CA(25R). Both 7αH,3,24-diO-CA(25R) and 3β,7α-diH,24O-CA(25) undergo decarboxylation in the EADSA process. | Present work |
| | | | | | 7α,12α-Dihydroxy-3-oxocholest-4-en-(25R/S)26-oic acid | 7α,12α-diH,3O-CA(25R/S)↑ | See prediction | [+] C34H50N3O5 | LMST04030150 | (B)+ | [$^2H_0$]GP 580.37450↑ | NA | Also observed in (A)+ at 585.4059 in combination with 3β,12α,7α-triHCA. LipidMaps ID is for the (25R)26-epimer. | Present work |
| | | | | | | | Negative-ion | | | | | | | |
| | | | | | 7α-Hydroxy-3-oxochol-4-en-24-oyl-glycine | 7αH,3O-Δ$^4$-BA-24-Gly↑ | [-]444.2755↑ | [-]C26H38NO5 | LMST05030020 | (C)- | | 70ʃ | ʃValue determined following enzymatic deconjugation, data for children. | (ʃLemonde 2003) |
| | | | | | 3α,7α-Dihydroxy-5β-cholan-24-oyl-glycine (Glycochenodeoxycholic acid) | 3α,7α-diHBA-24-Gly (GCDCA)↓ | [-]448.3068↓ | [-]C26H42NO5 | LMST05030008 | (C)- | | 0.01ʃ | ʃValue determined by GC-MS following enzymatic deconjugation, data for children. | (ʃLemonde 2003) |
| | | | | | 7α,12α-Dihydroxy-3-oxochol-4-en-24-oyl-glycine | 7α,12α-diH,3O-Δ$^4$-BA-Gly↑ | [-]460.2705↑ | [-]C26H38NO6 | NA | (C)- | | 80ʃ | ʃValue determined following enzymatic deconjugation, data for children. | (ʃLemonde 2003) |
| | | | | | 3α,7α,12α-Trihydroxy-5β-cholan-24-oyl-glycine (Glycocholic acid) | 3α,7α,12α-triHBA-24-Gly (GCA)↓ | [-]464.3018↓ | [-]C26H42NO6 | LMST05030001 | (C)- | | 0.05ʃ | ʃValue determined by GC-MS following enzymatic deconjugation, data for children. | (ʃLemonde 2003) |
| | | | | | 7α-Hydroxy-3-oxochol-4-en-24-oyl-taurine | 7αH,3O-Δ$^4$-BA-24-Tau↑ | [-]494.2582↑ | [-]C26H40NO6S | LMST05040022 | (C)- | | 70ʃ | Taurine conjugates are lost in fractions (A) and (B) following GP derivatisation. ʃValue determined following enzymatic deconjugation, data for children. | (ʃLemonde 2003) |
| | | | | | 3α,7α-Dihydroxy-5β-cholan-24-oyl-taurine (Taurochenodeoxycholic acid) | 3α,7α-diHBA-24-Tau (TCDCA)↓ | [-]498.2895↓ | [-]C26H44NO6S | LMST05040005 | (C)- | | 0.01 | Taurine conjugates are lost in fractions (A) and (B) following GP derivatisation. ʃValue determined by GC-MS following enzymatic deconjugation, data for children. | (ʃLemonde 2003) |
| | | | | | 7α,12α-Dihydroxy-3-oxochol-4-en-24-oyl-taurine | 7α,12α-diH,3O-Δ$^4$-BA-24-Tau↑ | [-]510.2531↑ | [-]C26H40NO7S | NA | (C)- | | 80ʃ | Taurine conjugates are lost in fractions (A) and (B) following GP derivatisation. ʃValue determined following enzymatic deconjugation, data for children. | (ʃLemonde 2003) |
| | | | | | 3α,7α,12α-Trihydroxy-5β-cholan-24-oyl-taurine (Taurocholic acid) | 3α,7α,12α-triHBA-Tau (TCA)↓ | [-]514.2844↓ | [-]C26H44NO7S | LMST05040001 | (C)- | | 0.05ʃ | Taurine conjugates are lost in fractions (A) and (B) following GP derivatisation. ʃValue determined by GC-MS following enzymatic deconjugation, data for children. | (ʃLemonde 2003) |
| | | | | | 7α,12α-Dihydroxy-3-oxocholest-4-en-26-oyl-taurine | 7α,12α-diH,3O-CA-26-Tau↑ | [-]552.3000↑ | [-]C29H46NO7S | NA | (C)- | | NA | Taurine conjugates are lost in fractions (A) and (B) following GP derivatisation. | Vaz 2017 |
| 3.3.1 | | *SLC27A5* | *603314* | Bile acyl CoA synthetase (BACS) deficiency | | | Negative-ion | | | | | | | |
| | | | | | 3α,7α-Dihydroxy-5β-cholan-24-oic acid (Chenodeoxycholic acid) | 3α,7α-diHBA (CDCA)↑ | [-]391.2854↑ | [-]C24H39O4 | LMST04010032 | (B)- or (C)- | | >6ʃ | ʃValue estimated following enzymatic deconjugation, data for infants. | (ʃChong 2012) |
| | | | | | 3α,7β-Dihydroxy-5β-cholan-24-oic acid (Ursodeoxycholic acid) | 3α,7β-diHBA (UDCA)↑ | [-]391.2854↑ | [-]C24H39O4 | LMST04010033 | (B)- or (C)- | | >7ʃ | ʃValue estimated following enzymatic deconjugation, data for infants. | (ʃChong 2012) |
| | | | | | 3α,7α-Dihydroxy-5β-cholan-24-oyl-glycine (Glycochenodeoxycholic acid) | 3α,7α-diHBA-24-Gly (GCDCA)↓ | [-]448.3068↓ | [-]C26H42NO5 | LMST05030008 | (C)- | | 0.5ʃ | ʃValue estimated following enzymatic deconjugation, data for infants. | (ʃChong 2012) |
| | | | | | 3α,7β-Dihydroxy-5β-cholan-24-oyl-glycine (Glycoursodeoxycholic acid) | 3α,7β-diHBA-24-Gly (GUDCA)↓ | [-]448.3068↓ | [-]C26H42NO5 | LMST05030016 | (C)- | | 0.5ʃ | ʃValue estimated following enzymatic deconjugation, data for infants. | (ʃChong 2012) |
| | | | | | 3α,7α,12α-Trihydroxy-5β-cholan-24-oyl-glycine (Glycocholic acid) | 3α,7α,12α-triHBA-24-Gly (GCA)↓ | [-]464.3018↓ | [-]C26H42NO6 | LMST05030001 | (C)- | | 0.5ʃ | ʃValue estimated following enzymatic deconjugation, data for infants. | (ʃChong 2012) |
| | | | | | 3α,7α-Dihydroxy-5β-cholan-24-oyl-taurine (Taurochenodeoxycholic acid) | 3α,7α-diHBA-24-Tau (TCDCA)↓ | [-]498.2895↓ | [-]C26H44NO6S | LMST05040005 | (C)- | | NA | Taurine conjugates are lost in fractions (A) and (B) following GP derivatisation. | Chong 2012 |
| | | | | | 3α,7α,12α-Trihydroxy-5β-cholan-24-oyl-taurine (Taurocholic acid) | 3α,7α,12α-triHBA-24-Tau (TCA)↓ | [-]514.2844↓ | [-]C26H44NO7S | LMST05040001 | (C)- | | NA | Taurine conjugates are lost in fractions (A) and (B) following GP derivatisation. | Chong 2012 |
| 3.3.2 | 614307 | *AMACR* | *604489* | Alpha-methylacyl-CoA recemase (AMACR) deficiency | | | Positive-ion | | | | | | | |
| | | | | | 7α-Hydroxy-3-oxocholest-4-en-(25R)26-oic acid | 7αH,3O-CA(25R)↑ | [$^2H_0$]GP 564.37958↑ | [+]C34H50N3O4 | LMST04030242 | (B)+ | | NA | Extrapolated from analysis of *AMACR$^{-/-}$* mouse. Also observed in (A)+ at 569.4110 in combination with 3β,7α-diHCA(25R). | Autio 2014 |
| | | | | | 7α-Hydroxy-3-oxocholest-4-en-(25S)26-oic acid | 7αH,3O-CA(25S)↓ | [$^2H_0$]GP 564.37958↓ | [+]C34H50N3O4 | NA | (B)+ | | NA | Extrapolated from analysis of *AMACR$^{-/-}$* mouse. Also observed in (A)+ at 569.4110 in combination with 3β,7α-diHCA(25S). | Autio 2014 |
| | | | | | 7α,12α-Dihydroxy-3-oxocholest-4-en-(25R)26-oic acid | 7α,12α-diH,3O-CA(25R)↑ | [$^2H_0$]GP 580.3745↑ | [+]C34H50N3O5 | LMST04030243 | (B)+ | | NA | Extrapolated from analysis of *AMACR$^{-/-}$* mouse. Also observed in (A)+ at 585.4059 in combination with 3β,7α,12α-triHCA(25R). | Present work |
| | | | | | | | Negative-ion | | | | | | | |
| | | | | | 2,6,10,14-Tetramethylpentadecanoic acid (Pristanic acid) | Pristanic acid↑ | [-]297.2799↑ | [-]C19H37O2 | LMPR0104010022 | (B)- or (C)- | | NA | Brabched chain fatty acid. | Ferdinandusse 2001 |
| | | | | | 3α,7α-Dihydroxy-5β-cholestan-(25R)26-oic acid | 3α,7α-diHCa(25R)↑ | [-]433.3323↑ | [-]C27H45O4 | LMST04030153 | (B)- or (C)- | | >300• | •Ratio estimated based on an LOD of 0.06 μM in infant/child cholestasis. | (•Ferdinandusse 2001) |
| | | | | | 3α,7α,12α-Trihydroxy-5β-cholestan-(25R)26-oic acid | 3α,7α,12α-triHCa(25R)↑ | [-]449.3272↑ | [-]C27H45O5 | LMST04030151 | (B)- or (C)- | | >25• | •Ratio estimated based on an LOD of 0.06 μM in infant/child cholestasis. | (•Ferdinandusse 2001) |
| | | | | | Tetrahydroxy-5β-cholestan-26-oic acid↑ | tetraHCa↑ | [-]465.3222↑ | [-]C27H45O6 | NA | (B)- or (C)- | | NA | | Setchell 2003 |
| | | | | | 3α,7α-Dihydroxy-5β-cholestan-(25R)26-oyl-taurine↑ | 3α,7α-diHCa-(25R)26-Tau↑ | [-]540.3364↑ | [-]C29H50NO6S | LMST05040008 | (C)- | | >10• | Taurine conjugates are lost in fractions (A) and (B) following GP derivatisation. •Ratio estimated based on an LOD of 0.06 μM in infant/child cholestasis. | (•Ferdinandusse 2001) |
| | | | | | 3α,7α,12α-Trihydroxy-5β-cholestan-(25R)26-oyl-taurine↑ | 3α,7α,12α-triHCa-(25R)26-Tau↑ | [-]556.3313↑ | [-]C29H50NO7S | LMST05040007 | (C)- | | >20• | Taurine conjugates are lost in fractions (A) and (B) following GP derivatisation. • Ratio estimated against infant/child cholestasis. | (•Ferdinandusse 2001) |
| 3.3.3 | 617308 | *ACOX2* | *601641* | Acyl-CoA oxidase 2 (ACOX2) deficiency | | | Positive-ion | | | | | | | |
| | | | | | 3β-Hydroxycholest-5-en-(25R)26-oic acid (Cholrestenoic acid) | 3β-HCA↑ | [$^2H_5$]GP 553.4161↑ | [+]C34H45D5N3O3 | LMST04030072 | (A)+ | | 5ǁ | ǁRatio of ACOX2 to NIST SRM 1950. | (ǁYutuc 2021) |
| | | | | | 7α-Hydroxy-3-oxocholest-4-en-(25R/S)26-oic acid | 7αH,3O-CA(25R/S)↑ | [$^2H_0$]GP 564.37958↑ | [+]C34H50N3O4 | LMST04030242 | (B)+ | | 5ǁ | Also observed in (A)+ at 569.4110 in combination with 3β,7α-diHCA(25R/S). LipidMaps ID is for the (25R)26-epimer. ǁRatio of ACOX2 to NIST SRM 1950. | (ǁYutuc 2021) |
| | | | | | 3β-Hydroxy,7-oxocholest-5-en-(25R/S)26-oic acid | 3βH,7O-CA(25R/S)↑ | [$^2H_0$]GP 564.3796↑ | [+]C34H50N3O4 | LMST04030239 | (B)+ | | 10† | LipidMaps ID is for the (25R)26-epimer. †Ratio of ACOX2 to NIST SRM 1950. | (†Present work) |
| | | | | | 3β,7α-Dihydroxycholest-5-en-(25R/S)26-oic acid | 3β,7α-diHCA(25R/S)↑ | [$^2H_5$]GP569.4110↑ | [+]C34H45D5N3O4 | LMST04030241 | (A)+ | | 5ǁ | In (A)+ observed in combination with 7αH,3O-CA(25R). LipidMaps ID is for the (25R)26-epimer. ǁRatio of ACOX2 to NIST SRM 1950. | (ǁYutuc 2021) |
| | | | | | 3β,7β-Dihydroxycholest-5-en-(25R/S)26-oic acid | 3β,7β-diHCA(25R/S)↑ | [$^2H_5$]GP 569.4110↑ | [+]C34H45D5N3O4 | LMST04030235 | (A)+ | | 10ǁ | LipidMaps ID is for the (25R)26-epimer. ǁRatio of ACOX2 to NIST SRM 1950. | (ǁYutuc 2021) |
| | | | | | 3β,5α,6β-Trihydroxycholestan-(25R)26-oic acid | 3β,5α,6β-triHCa-H$_2$O↑ | [$^2H_5$]GP 569.4110↑ | [+]C34H45D5N3O4 | LMST04030238 | (A)+ | | 30† | 3β,5α,6β-triHCa observed as the dehydrated molecule. †Ratio of ACOX2 to NIST SRM 1950. | (†Present work) |
| | | | | | 7α,12α-Dihydroxy-3-oxocholest-4-en-(25S)26-oic acid | 7α,12α-diH,3O-CA(25S)↑ | [$^2H_0$]GP 580.37450↑ | [+] C34H50N3O5 | LMST04030150 | (B)+ | | 5† | Also observed in (A)+ at 585.4059 in combination with 3β,12α,7α-triHCA(25S). LipidMaps ID does not give stereochemistry at C-25. †Ratio of ACOX2 to NIST SRM 1950. | (†Present work) |
| | | | | | | | Negative-ion | | | | | | | |
| | | | | | 7α-Hydroxy-3-oxocholest-4-en-(25R/S)26-oic acid | 7αH,3O-CA(25R/S)↑ | [-]429.301↑ | [-]C27H41O4 | LMST04030242 | (B)- or (C)- | | 5† | LipidMaps ID is for the (25R)26-epimer. †Ratio of ACOX2 to NIST SRM 1950. | Yutuc 2021, (†Present work) |
| | | | | | 3β,7α-Dihydroxycholest-5-en-(25R/S)26-oic acid | 3β,7α-diHCA(25R/S)↑ | [-]431.3167↑ | [-]C27H43O4 | LMST04030241 | (B)- or (C)- | | 5† | LipidMaps ID is for the (25R)26-epimer. †Ratio of ACOX2 to NIST SRM 1950. | Yutuc 2021, (†Present work) |
| | | | | | 3α,7α-Dihydroxy-5β-cholestan-(25R/S)26-oic acid | 3α,7α-diHCa(25R/S)↑ | [-]433.3323↑ | [-]C27H45O4 | LMST04030153 | (B)- or (C)- | | >50* - >200† | LipidMaps ID is for the (25R)26-epimer. *Ratio for ACOX2 child to heterozygous unafected father. †Ratio of ACOX2 to NIST SRM 1950. | (*Vilarinho 2016), Monte 2017, (†Present work) |
| | | | | | 3α,7α,12α-Trihydroxy-5β-cholestan-(25R/S)26-oic acid | 3α,7α,12α-triHCa(25R/S)↑ | [-]449.3272↑ | [-]C27H45O5 | LMST04030151 | (B)- or (C)- | | 5* - >200† | LipidMaps ID is for the (25R)26-epimer. *Ratio for ACOX2 child to heterozygous unafected father. †Ratio of ACOX2 to NIST SRM 1950. | (*Vilarinho 2016), Monte 2017, (†Present work) |
| | | | | | 3α,7α-Dihydroxy-5β-cholestan-(25R/S)26-oyl-glycine | 3α,7α-diHCa-(25R/S)26-Gly↑ | [-]490.3538↑ | [-]C29H48NO5 | NA | (C)- | | >5* - > 200† | Presumably both 25R- and 25S-epimers in Vilarinho 2016.*Ratio for ACOX2 child to heterozygous unafected father. †Ratio of ACOX2 to NIST SRM 1950. | (*Vilarinho 2016), (†Present work) |
| | | | | | 3α,7α,12α-Trihydroxy-5β-cholestan-(25R/S)26-oyl-glycine | 3α,7α,12α-triHCa-(25R/S)26-Gly↑ | [-]506.3487↑ | [-]C29H48NO6 | NA | (C)- | | >10* - >20† | Presumably both 25R- and 25S-epimers in Vilarinho 2016. *Ratio for ACOX2 child to heterozygous unafected father. †Ratio of ACOX2 to NIST SRM 1950. | (*Vilarinho 2016), (†Present work) |
| | | | | | 3α,7α-Dihydroxy-5β-cholestan-(25R/S)26-oyl-taurine↑ | 3α,7α-diHCa-(25R/S)26-Tau↑ | [-]540.3364↑ | [-]C29H50NO6S | LMST05040008 | (C)- | | >5* - >100† | Taurine conjugates are lost in fractions (A) and (B) following GP derivatisation. LipidMaps ID is for the (25R)26-epimer. Presumably both 25R and 25S in Vilarinho 2016. *Ratio for ACOX2 child to heterozygous unafected father. †Ratio of ACOX2 to NIST SRM 1950. | (*Vilarinho 2016), Monte 2017, (†Present work) |
| | | | | | 3α,7α,12α-Trihydroxy-5β-cholestan-(25R/S)26-oyl-taurine↑ | 3α,7α,12α-triHCa-(25R/S)26-Tau↑ | [-]556.3313↑ | [-]C29H50NO7S | LMST05040007 | (C)- | | >25* - 20† | Taurine conjugates are lost in fractions (A) and (B) following GP derivatisation. LipidMaps ID is for the (25R)26-epimer. Presumably both 25R and 25S in Vilarinho 2016. *Ratio for ACOX2 child to upper reference value. †Ratio of ACOX2 to NIST SRM 1950. | (*Vilarinho 2016), Monte 2017, (†Present work) |
| | | | | | Tetrahydroxy-5β-cholestan-(25R/S)26-oyl-taurine↑ | tetraHCa-(25R/S)26-Tau↑ | [-]572.3263↑ | [-]C29H50NO8S | NA | (C)- | | >500† | Taurine conjugates are lost in fractions (A) and (B) following GP derivatisation. Presumably both 25R- and 25S-epimers in Ferdinanusse 2018. †Ratio of ACOX2 to NIST SRM 1950. | Ferdinandusse 2018, (†Present work) |
| 3.3.4 | 261515 | *HSD17B4* | *601860* | D-Bifunctional protein (DBP) deficiency type I & II & III | | | Positive-ion | | | | Positive-ion | | | |
| | | | | | 7α-Hydroxy-3-oxocholesta-5,24*E*-dien-26-oic acid | 7αH,3O-Δ$^{24E}$-CA↑ | See prediction | [+]C34H48N3O4 | NA | (B)+ | [$^2H_0$]GP562.3639↑ | NA | Also predicted in (A)+ at 567.3953 in combination with 3β,7α-diH-Δ$^{24}$-CA. | Present work |
| | | | | | 7α-Hydroxy-3-oxocholest-4-en-(25S)26-oic acid | 7αH,3O-CA(25S)↑ | See prediction | [+]C34H50N3O4 | NA | (B)+ | [$^2H_0$]GP564.3796↑ | NA | Also predicted in (A)+ at 569.4110 in combination with 3β,7α-diHCA(25S). | Present work |
| | | | | | 3β,7α-Dihydroxycholesta-5,24*E*-dien-26-oic acid | 3β,7α-diH-Δ$^{24E}$-CA↑ | See prediction | [+]C34H43D5N3O4 | NA | (A)+ | [$^2H_5$]GP567.3953↑ | NA | Predicted in (A)+ in combination with 7αH,3O-Δ$^{24E}$-CA. | Present work |
| | | | | | 3β,7α-Dihydroxycholest-5-en-(25S)26-oic acid | 3β,7α-diHCA(25S)↑ | See prediction | [+]C34H45D5N3O4 | NA | (A)+ | [$^2H_5$]GP569.4110↑ | NA | Predicted in (A)+ in combination with 7αH,3O-CA(25S). | Present work |
| | | | | | 7α,24S-Dihydroxy-3-oxocholest-4-en-(25S)26-oic acid | 7α,24S-diH,3O-CA(25S)↑ | See prediction | [+]C34H48N3O4 | LMST04030260 | (B)+ | [$^2H_0$]GP580.3745↑ | NA | D-Bifunctional protein deficiency types I & II. Also predicted in (A)+ at 585.4059 in combination with 3β,7α,24S-triHCA(25S). | Present work |
| | | | | | 7α,24R-Dihydroxy-3-oxocholest-4-en-(25R)26-oic acid | 7α,24R-diH,3O-CA(25R)↑ | See prediction | [+]C34H48N3O4 | LMST04030257 | (B)+ | [$^2H_0$]GP580.3745↑ | NA | D-Bifunctional protein deficiency type III only. Also predicted in (A)+ at 585.4059 in combination with 3β,7α,24R-triHCA(25R). | Present work |
| | | | | | 3β,7α,24S-Trihydroxycholest-5-en-(25S)26-oic acid | 3β,7α,24S-triHCA(25S)↑ | See prediction | [+]C34H45D5N3O | NA | (A)+ | [$^2H_5$]GP585.4059↑ | NA | D-Bifunctional protein deficiency types I & II. Predicted in (A)+ in combination with 7α,24S-diH,3O-CA(25S). | Present work |
| | | | | | 3β,7α,24R-Trihydroxycholest-5-en-(25R)26-oic acid | 3β,7α,24R-triHCA(25R)↑ | See prediction | [+]C34H45D5N3O | NA | (A)+ | [$^2H_5$]GP585.4059↑ | NA | D-Bifunctional protein deficiency (type III) only. Predicted in (A)+ in combination with 7α,24R-diH,3O-CA(25R). | Present work |
| | | | | | | | Negative-ion | | | | Negative-ion | | | |
| | | | | | 2,6,10,14-Tetramethylpentadecanoic acid (Pristanic acid) | Pristanic acid↑ | [-]297.2799↑ | [-]C19H37O2 | LMPR0104010022 | (B)- or (C)- | | | Brabched chain fatty acid. | Vaz 2017 |
| | | | | | 3,7R,11R,15-tetramethyl-hexadecanoic acid (Phytanic acid) | Phytanic acid↑ | [-]311.2956↑ | [-]C20H39O2 | LMPR0104010004 | (B)- or (C)- | | | Brabched chain fatty acid. | |
| | | | | | 3α,7α-Dihydroxy-5β-cholestan-(25R/S)26-oic acid | 3α,7α-diHCa(25R/S)↑ | [-]433.3323↑ | [-]C27H45O4 | LMST04030153 | (B)- or (C)- | | NA | Presumably both 25R- and 25S-epimers are elevated. LipidMaps ID is for the (25R)26-epimer. | Vaz 2017 |
| | | | | | 3α,7α,12α-Trihydroxy-5β-cholestan-(25R/S)26-oic acid | 3α,7α,12α-triHCa(25R/S)↑ | [-]449.3272↑ | [-]C27H45O5 | LMST04030151 / LMST04030152 | (B)- or (C)- | | NA | Presumably both 25R- and 25S-epimers are elevated. | Vaz 2017 |
| | | | | | 3α,7α,12α,24S-Tetrahydroxy-5β-cholestan-(25S)26-oic acid | 3α,7α,12α,24S-tetraHCa(25S)↑ | [-]465.3222↑ | [-]C27H45O6 | LMST04030196 | (B)- or (C)- | | NA | D-Bifunctional protein deficiency types I & II. | Une 1997, Vreken 1998, |
| | | | | | 3α,7α,12α,24R-Tetrahydroxy-5β-cholestan-(25R)26-oic acid | 3α,7α,12α,24R-tetraHCa(25R)↑ | [-]465.3222↑ | [-]C27H45O6 | LMST04030195 | (B)- or (C)- | | NA | D-Bifunctional protein deficiency type III only. | Une 1997, Vreken 1998 |
| | | | | | 3α,7α-Dihydroxy-5β-cholestan-(25R/S)26-oyl-taurine | 3α,7α-diHCa-(25R/S)26-Tau↑ | [-]540.3364↑ | [-]C29H50NO6S | LMST05040008 | (C)- | | NA | Taurine conjugates are lost in fractions (A) and (B) following GP derivatisation. Presumably both 25R- and 25S-epimers are elevated. LipidMaps ID is for the (25R)26-epimer. | Vaz 2017 |
| | | | | | 3α,7α,12α-Trihydroxy-5β-cholest-24*E*-en-26-oyl-taurine | 3α,7α,12α-triH-Δ$^{24E}$-Ca-26-Tau↑ | [-]554.3157↑ | [-]C29H48NO7S | NA | (C)- | | NA | Taurine conjugates are lost in fractions (A) and (B) following GP derivatisation. | Une 1997, Vaz 2017 |
| | | | | | 3α,7α,12α-Trihydroxy-5β-cholestan-(25R/S)26-oyl-taurine | 3α,7α,12α-triHCa-(25R/S)26-Tau↑ | [-]556.3313↑ | [-]C29H50NO7S | LMST05040007 | (C)- | | NA | Taurine conjugates are lost in fractions (A) and (B) following GP derivatisation. Presumably both 25R- and 25S-epimers are elevated. LipidMaps ID is for the (25R)26-epimer. | Vaz 2017 |
| | | | | | Tetrahydroxy-5β-cholestenoyl-taurine | tetraH-Δ$^{24E}$-Ca-26-Tau↑ | [-]570.3106↑ | [-]C29H48NO8S | NA | (C)- | | NA | Taurine conjugates are lost in fractions (A) and (B) following GP derivatisation. | Vaz 2017 |
| | | | | | 3α,7α,12α,24S-Tetrahydroxy-5β-cholestan-(25S)26-oyl-taurine | 3α,7α,12α,24S-tetraHCa-(25S)26-Tau↑ | [-]572.3263↑ | [-]C29H50NO8S | NA | (C)- | | NA | D-Bifunctional protein deficiency types I & II. Taurine conjugates are lost in fractions (A) and (B) following GP derivatisation. Extrapoltated from GC-MS data. | Une 1997, Vreken 1998 |
| | | | | | 3α,7α,12α,24R-Tetrahydroxy-5β-cholestan-(25R)26-oyl-taurine | 3α,7α,12α,24R-tetraHCa-(25R)26-Tau↑ | [-]572.3263↑ | [-]C29H50NO8S | NA | (C)- | | NA | D-Bifunctional protein deficiency (type III) only. Taurine conjugates are lost in fractions (A) and (B) following GP derivatisation. Extrapoltated from GC-MS data. | Une 1997, Vreken 1998 |
| 3.3.5 | 613724 | *SCP2* | *184755* | Sterol carrier protein 2 (SCP2) deficiency | | | Positive-ion | | | | Positive-ion | | | |
| | | | | | 7α-Hydroxy-27-*nor*-cholest-4-ene-3,24-dione | 7αH-27-*nor*-C-3,24-diO↑ | See prediction | [+]C33H48N3O3 | NA | (B)+ | [$^2H_0$]GP534.369↑ | | Also predicted in (A)+ at 539.4004 in combination with 3β,7α-diH-27-*nor*-C-24-O. Both 7αH-27-*nor*-C-3,24-diO and 3β,7α-diH-27-*nor*-C-24O are decarboxylation products of 7αH,3,24-diO-CA(25R) and 3β,7α-diH,24O-CA(25), respectively. | Present work |
| | | | | | 7α-Hydroxy-3,24-*bis*oxocholest-4-en-(25R)26-oic acid | 7αH,3,24-diO-CA(25R)↑ | See prediction | [+] C34H48N3O5 | LMST04030255 | (B)+ | [$^2H_0$]GP 578.35885↑ | | Also observed in (A)+ at 583.3902 in combination with 3β,7α-diH,24O-CA(25R). Both 7αH,3,24-diO-CA(25R) and 3β,7α-diH,24O-CA(25) undergo decarboxylation in the EADSA process. | Present work |
| | | | | | | | Negative-ion | | | | | | | |
| | | | | | 2,6,10,14-Tetramethylpentadecanoic acid (Pristanic acid) | Pristanic acid↑ | [-]297.2799↑ | [-]C19H37O2 | LMPR0104010022 | (B)- or (C)- | | | Brabched chain fatty acid. | Ferdinandusse 2006, Vaz 2017 |
| | | | | | 3,7R,11R,15-Tetramethyl-hexadecanoic acid (Phytanic acid) | Phytanic acid↑ | [-]311.2956↑ | [-]C20H39O2 | LMPR0104010004 | (B)- or (C)- | | | Brabched chain fatty acid. | Ferdinandusse 2006, Vaz 2017 |
| | | | | | w,x,y-Trihydroxy-z-glucuronyl-27-*nor*-5β-cholestan-24-one (Tetrahydroxy-27-*nor*-5β-cholestan-24-one glucuronide) | w,x,y-triH,z-GlcA-27-*nor*-C-24O↑ | [-]611.3437↑ | [-]C32H51O11 | NA | (B)- or (C)- | | NA | Extrapolated from analysis of the *SCP2$^{-/-}$* mouse plasma and human urine. | Kannenberg 1999, Ferdinandusse 2006, Vaz 2017 |
| | | | | | v,w,x,y-Tetrahydroxy-27-*nor*-5β-cholestan-z-glucuronide (27-*nor*-5β-cholestanpentol glucuronide) | v,w,x,y-tetraH-27-*nor*-C-z-GlcA↑ | [-]613.3593↑ | [-]C32H53O11 | NA | (B)- or (C)- | | NA | Extrapolated from analysis human urine. | Ferdinandusse 2006, Vaz 2017 |
| | | | | | v,w,x,y-Tetrahydroxy-z-glucuronyl-27-*nor*- 5β-cholestan-24-one (Pentahydroxy-27-*nor*- 5β-cholestan-24-one glucuronide) | v,w,x,y-tetraH,z-GlcA-27-*nor*-C-24O↑ | [-]627.3386↑ | [-]C32H51O12 | NA | (B)- or (C)- | | NA | Extrapolated from analysis of the *SCP2$^{-/-}$* mouse plasma and human urine. | Kannenberg 1999, Ferdinandusse 2006, Vaz 2017 |
| | | | | | u,v,w,x,y-Pentahydroxy-27-*nor*-5β-cholestane-z-glucuronide (27-*nor*-5β-cholestanhexol glucuronide) | u,v,w,x,y-pentaH-27-*nor*-C-z-GlcA↑ | [-]629.3543↑ | [-]C32H53O12 | NA | (B)- or (C)- | | NA | Extrapolated from analysis human urine. | Ferdinandusse 2006, Vaz 2017 |
| | | | | | u,v,w,x,y-Pentahydroxy-z-glucyronyl-27-nor-5β-cholestane-24-one (Hexahydroxy-27-*nor*-5β-cholestan-24-one glucuronide) | u,v,w,x,y-pentaH,z-GlcA-27-*nor*-C-24O↑ | [-] 643.3335↑ | [-] C32H51O13 | NA | (B)- or (C)- | | NA | Extrapolated from analysis of the *SCP2$^{-/-}$* mouse plasma and human urine. | Kannenberg 1999, Ferdinandusse 2006, Vaz 2017 |
| | | | | | t,u,v,w,x,y-Hexahydroxy-27-*nor*-5β-cholestan-z-glucuronide (27-*nor*-5β-cholestanheptol glucuronide) | t,u,v,w,x,y-hexaH-27-*nor*-C-z-GlcA↑ | [-]645.3492↑ | [-]C32H53O13 | NA | (B)- or (C)- | | NA | | Ferdinandusse 2006, Vaz 2017 |
| 3.4 | 619232 | *BAAT* | *602938* | Bile acid-CoA: amino acid *N*-acyl transferase (BAAT) deficiency | | | Negative-ion | | | | | | | |
| | | | | | 3α,7α,12α-Trihydroxy-5β-cholan-24-oic acid (Cholic acid) | 3α,7α,12α-triHBA (Cholic acid)↑ | [-]407.2803↑ | [-]C24H39O5 | LMST04010001 | (B)- or (C)- | | 50* | | Clayton 2011, (*Setchell 2013) |
| | | | | | 3α,7α-Dihydroxy-5β-cholan-24-oyl-glycine (Glycochenodeoxycholic acid) | 3α,7α-DiHBA-24-Gly (GCDCA)↓ | [-]448.3068↓ | [-]C26H42NO5 | LMST05030008 | (C)- | | NA | | Clayton 2011, Setchell 2013 |
| | | | | | 3β-Sulfooxychol-5-en-24-oic acid | 3βS-Δ$^5$-BA↑ | [-]453.2316↑ | [-]C24H37O6S | NA | (C)- | | NA | Sulfuric acid conjugates are lost in fractions (A) and (B) following GP derivatisation. | Setchell 2013 |
| | | | | | 3α,7α,12α-Trihydroxy-5β-cholan-24-oyl-glycine (Glychocholic acid) | 3α,7α,12α-triHBA-24-Gly (GCA)↓ | [-]464.3018↓ | [-]C26H42NO6 | LMST05030001 | (C)- | | NA | | Clayton 2011, Setchell 2013 |
| | | | | | 3α-Sulfooxy-7α-hydroxy-5β-cholan-24-oic acid (Chenodeoxycholic acid sulfate) | 3αS,7αHBA (CDCA-S)↑ | [-]471.2422↑ | [-]C24H39O7S | LMST05020024 | (C)- | | NA | The sulfooxy group is presumed to be at C-3. Sulfuric acid conjugates are lost in fractions (A) and (B) following GP derivatisation. | Clayton 2011, Setchell 2013 |
| | | | | | 3α-Sulfooxy-7α,12-dihydroxy-5β-cholan-24-oic acid (Cholic acid sulfate) | 3αS,7α,12α-diBA (Cholic acid-S)↑ | [-]487.2371↑ | [-]C24H39O8S | NA | (C)- | | NA | The sulfooxy group is presumed to be at C-3. Sulfuric acid conjugates are lost in fractions (A) and (B) following GP derivatisation. | Clayton 2011 |
| | | | | | 3α,7α-Dihydroxy-5β-cholan-24-oyl-taurine (Taurochenodeoxycholic acid) | 3α,7α-diHBA-24-Tau (TCDCA)↓ | [-]498.2895↓ | [-]C26H44NO6S | LMST05040005 | (C)- | | NA | Taurine conjugates are lost in fractions (A) and (B) following GP derivatisation. | Clayton 2011, Setchell 2013 |
| | | | | | 3α,7α,12α-Trihydroxy-5β-cholan-24-oyl-taurine (Taurocholic acid) | 3α,7α,12α-triHBA (TCA)↓ | [-]514.2844↓ | [-]C26H44NO7S | LMST05040001 | (C)- | | NA | Taurine conjugates are lost in fractions (A) and (B) following GP derivatisation. | Clayton 2011, Setchell 2013 |
| | | | | | x-Hydroxy-y-glucuronyl-5β-cholan-24-oic acid (Chenodeoxycholic acid glucuronide) | GlcA-HBA (CDCA-GlcA)↑ | [-] 567.3175↑ | [-]C30H47O10 | LMST05010028 | (B)- or (C)- | | NA | LipidMaps ID for the 3-GlcA | Clayton 2011, Setchell 2013 |
| | | | | | x,y-Diydroxy-z-glucuronyl-5β-cholan-24-oic acid (Cholic acid glucuronide) | GlcA-diHBA (Cholic acid-GlcA)↑ | [-]583.3124↑ | [-]C30H47O11 | LMST05010044 | (B)- or (C)- | | NA | LipidMaps ID for the 3-GlcA | Clayton 2011, Setchell 2013 |
| 3.5 | 257220 and 607625 and 607616 and 620151 and 278000 | *NPC1 and NPC2 and SMPD1 and LIPA and LIPA* | *607623 and 601015 and 607608 and 613497 and 613497* | Niemann-Pick type C1 disease (NPC1) and Niemann-Pick type C2 disease (NPC2) and Acid sphingomyelinase deficiency (ASMD, Niemann Pick type B disease, NPB) and Wolman disease and Cholesterol ester storage disease (CESD) | | | Positive-ion | | | | Positive-ion | | | |
| | | | | | Sphing-4-enine-1-phosphocholine (Sphingosine-1-phosphocholine (Lysosphingomyelin) | SM(d18:1/0:0)↑ | [+]465.3452↑ | C23H50N2O5P(+) | LMSP01060001 | (B)+ | | | Not consistently elevated in NPC. Insufficient data for Wolman disease or CESD. Analysed as the underivatised [M+H]$^+$ ion. | Kuchar 2017 |
| | | | | | *N*-Hexadecanoyl-*O*-phosphocholine-L-serine (*N*-palmitoyl-*O*-phosphocholineserine, Lysosphingomyelin-509) | PPCS (lyso-SM509)↑ | [+]509.3350↑ | C24H50N2O7P(+) | LMFA08020401 | (B)+ | | | Elevated in NPC and NPB. Insufficient data for Wolman disease or CESD. Analysed as the underivatised [M+H]$^+$ ion. | Sidhu 2019 |
| | | | | | 3β-Hydroxy-7-oxocholen-5-en-24-oic acid | 3βH,7O-Δ$^5$-BA↑ | [$^2H_0$]522.33263↑ | [+]C31H44N3O4 | LMST04010394 | (B)+ | [$^2H_0$]522.33263↑# | >5[n,a,w] | #Predicted for CESD. [n,a,w]Concentration ratios for NPC, ASMD and Wolman to NIST SRM 1950. | ([n,a,w]Griffiths 2018, 2019), Present work |
| | | | | | 3β,5α,6β-Trihydroxycholan-24-oic acid | 3β,5α,6β-triHBA-18↑ | [$^2H_5$]GP 527.36403↑ | [+]C31H39D5N3O4 | LMST04010339 | (A)+ | [$^2H_5$]GP 527.36403↑# | >10[n,a,w] | In (A)+ 3β,5α,6β-triHBA is observed as the [M-18]$^+$ ion at *m/z* 527.36403 (3β,6β-diH-Δ$^5$-BA). #Predicted for CESD. [n,a,w]Concentration ratios for NPC, ASMD and Wolman to NIST SRM 1950. | Jiang 2016, ([n,a,w]Griffiths 2018, 2019), Present work |
| | | | | | 3β,7β-Dihydroxychol-5-en-24-oic acid | 3β,7β-diH-Δ$^5$-BA↑ | [$^2H_5$]GP 527.36403↑ | [+]C31H39D5N3O4 | LMST04010280 | (A)+ | [$^2H_5$]GP 527.36403↑# | >5[n,a,w] | #Predicted for CESD. [n,a,w]Concentration ratios for NPC, ASMD and Wolman to NIST SRM 1950. | ([n,a,w]Griffiths 2018, Griffiths 2019), Present work |
| | | | | | 3β-Hydroxycholest-5-en-7-one (7-Oxocholesterol or 7-Ketocholesterol) | 7-OC (7-KC)↑ | [$^2H_0$]GP 534.4054↑ | [+]C34H52N3O2 | LMST01010049 | (B)+ | [$^2H_0$]GP 534.4054↑# | >5[n] > 10[n,a,w] | #Predicted for CESD. [n,a,w]Concentration ratios for NPC, ASMD and Wolman to NIST SRM 1950. [n]Concentration ratio for NPC infant to adult. | ([n]Jiang 2011), ([n,a,w]Griffiths 2018, 2019), Present work |
| | | | | | Cholestane-3β,5α,6β-triol | 3β,5α,6β-triol-18↑ | [$^2H_5$]GP 539.4368↑ | [+]C34H47D5N3O2 | LMST01010052 | (A)+ | [$^2H_5$]GP 539.4368↑# | >5[n] - >5[n,a,w] | In (A)+ 3β,5α,6β-triol (5α,6β-diHC) is predominantly observed as the [M-18]$^+$ ion at *m/z* 539.4368 (6β-HC). #Predicted for CESD. [n,a,w]Concentration ratios for NPC, ASMD and Wolman to NIST SRM 1950. [n]Concentration ratio for NPC infant to adult. | ([n]Jiang 2011), ([n,a,w]Griffiths 2018, 2019), Present work |
| | | | | | Cholest-5-ene-3β,7β-diol (7β-Hydroxycholesterol) | 7β-HC↑ | [$^2H_5$]GP 539.4368↑ | [+]C34H47D5N3O2 | LMST01010047 | (A)+ | [$^2H_5$]GP 539.4368↑# | >5[n,a,w] | #Predicted for CESD. [n,a,w]Concentration ratios for NPC, ASMD and Wolman to NIST SRM 1950. | ([n,a,w]Griffiths 2018, 2019), Present work |
| | | | | | Cholestane-3β,5α,6β-triol | 3β,5α,6β-triol (C-triol)↑ | [$^2H_5$]GP 557.44737↑ | [+]C34H49D5N3O3 | LMST01010052 | (A)+ | [$^2H_5$]GP 557.44737↑# | >5[n] - >10[n,a,w] | In (A+) 3β,5α,6β-triol (5α,6β-diHC) is predominantly observed as the [M-18]$^+$ ion at *m/z* 539.4368 (6β-HC). #Predicted for CESD. [n,a,w]Concentration ratios for NPC, ASMD and Wolman to NIST SRM 1950. [n]Concentration ratio for NPC infant to adult. | ([n]Jiang 2011), ([n,a,w]Griffiths 2018, 2019), Present work |
| | | | | | 3β-Hydroxy-7-oxocholest-5-en-(25R/S)26-oic acid | 3βH,7O-CA(25R/S)↑ | [$^2H_0$]GP 564.3796↑ | [+]C34H50N3O4 | LMST04030239 | (B)+ | [$^2H_0$]GP 564.3796↑# | >10[n,a,w] | The epimer 3βH,7O-CA(25S) is of lower abundance than 3βH,7O-CA(25R). The LipidMaps ID is for the (25R)26-epimer. #Predicted for CESD. [n,a,w]Concentration ratios for NPC, ASMD and Wolman to NIST SRM 1950. | ([n,a,w]Griffiths 2018, 2019), Present work |
| | | | | | 3β,5α,6β-Trihydroxycholestan-(25R)26-oic acid | 3β,5α,6β-triHCA-18↑ | [$^2H_5$]GP 569.41098↑ | [+]C34H45D5N3O4 | LMST04010339 | (A)+ | [$^2H_5$]GP 569.41098↑# | >10[n,a,w] | In (A)+ 3β,5α,6β-triCa is observed as the [M-18]$^+$ ion at *m/z* 569.41098 (3β,6β-diHCA). #Predicted for CESD. [n,a,w]Concentration ratios for NPC, ASMD and Wolman to NIST SRM 1950. | Jiang 2016, Mazzacuva 2016, ([n,a,w]Griffiths 2018, 2019) Present work |
| | | | | | 3β,7β-Dihydroxycholest-5-en-(25R/S)26-oic acid | 3β,7β-diHCA(25R/S)↑ | [$^2H_5$]GP 569.41098↑ | [+]C34H45D5N3O4 | LMST04030235 | (A)+ | [$^2H_5$]GP 569.41098↑# | >5[n,a,w] | The epimer 3β,7β-diHCA(25S) is of lower abundance than 3β,7β-diHCA(25R). The LipidMaps ID is for the (25R)26-epimer. #Predicted for CESD. [n,a,w]Concentration ratios for NPC, ASMD and Wolman to NIST SRM 1950. | ([n,a,w]Griffiths 2018, 2019) Present work |
| | | | | | | | Negative -ion | | | | Negative -ion | | | |
| | | | | | 3β,5α,6β-Trihydroxycholan-24-oic acid | 3β,5α,6β-triHBA↑ | [-]407.2803↑ | [-]C24H39O5 | LMST04010339 | (B)- or (C)- | [-]407.2803↑# | >40[fn], >100[fa], >10[fw] | #Predicted for CESD. [fn]Concentration ratios for NPC to NIST SRM 1950. [fa]Concentration ratios for ASMD to NIST SRM 1950. [fw]Concentration ratios for Wolman to NIST SRM 1950. | Jiang 2016, Mazzacuva 2016, Griffiths 2019, ([f,n,a,w]Present work) |
| | | | | | 3β,5α,6β-Trihydroxycholan-24-oyl-glycine | 3β,5α,6β-triHBA-24-Gly↑ | [-]464.3018↑ | [-]C26H42NO6 | LMST05030019 | (C)- | [-]464.3018↑# | >3[nl] - > 4[na] - >20[fn], >30[fa], >2[fw] | #Predicted for CESD. [nl]Concentration ratio for NPC to control newborn dried blood spots. [na]Concentration ratio for NPC to adult control. [fn]Concentration ratio for NPC to NIST SRM 1950. [fa]Concentration ratios for ASMD to NIST SRM 1950. [fw]Concentration ratios for Wolman to NIST SRM. 1950. | ([nl]Jiang 2016), ([na]Motamed-Gorji), Mazzacuva 2016, Griffiths 2019, ([f,n,a,w]Present work) |
| | | | | | 3β,5α,6β-Trihydroxycholan-24-oyl-taurine | 3β,5α,6β-triHBA-24-Tau↑ | [-]514.2844↑ | [-]C26H44NO7S | NA | (C)- | [-]514.2844↑# | >30[fn], >40[fa], >5[fw] | Taurine conjugates are lost in fractions (A) and (B) following GP derivatisation. #Predicted for CESD. [fn]Concentration ratio for NPC to NIST SRM 1950. [fa]Concentration ratios for ASMD to NIST SRM 1950. [fw]Concentration ratios for Wolman to NIST SRM. 1950. | Jiang 2016, ([f,n,a,w]Present work) |
| | | | | | 3β-Hydroxy-7β-*N*- acetylglucosaminyl-chol-5-en-24-oic acid | 3βH,7β-GlcNAc-Δ$^5$-BA (3β,7β-diH-Δ$^5$-BA-7β-GlcNAc)↑$ | [-]592.3491↑ | [-]C32H50NO9 | NA | (B)- or (C)- | [-]592.3491↑# | $, >15[fn], >70[fa], >2[fw] | #Predicted for CESD. $GlcNAc conjugates not found in all patient samples. [fn]Concentration ratio for NPC to NIST SRM 1950. [fa]Concentration ratios for ASMD to NIST SRM 1950. [fw]Concentration ratios for Wolman to NIST SRM. 195 | Alvelius 2001, Mazzacuva 2016, ([f,n,a,w]Present work) |
| | | | | | 3β-Hydroxy-7β-*N*- acetylglucosaminyl-chol-5-en-24-oyl-glycine | 3βH,7β-GlcNAc-Δ$^5$-BA-24-Gly (3β,7β-diH-Δ$^5$-BA-24-Gly-7β-GlcNAc)↑$ | [-]649.3706↑ | [-]C34H53N2O10 | NA | (C)- | [-]649.3706↑# | $, >5[fn], >50[fa], 1[fw] | #Predicted for CESD. $GlcNAc conjugates not found in all patient samples. [fn]Concentration ratio for NPC to NIST SRM 1950. [fa]Concentration ratios for ASMD to NIST SRM 1950. [fw]Concentration ratios for Wolman to NIST SRM. 195 | Alvelius 2001, Mazzacuva 2016, Present work |
| | | | | | 3β-Sulfooxy-7β-*N*- acetylglucosaminyl-chol-5-en-24-oic acid | 3βS,7β-GlcNAc-Δ$^5$-BA (3β,7β-diH-Δ$^5$-BA-3βS-7β-GlcNAc)↑$ | [-]672.3059↑ | [-]C32H50NO12S | LMST05050051 | (C)- | [-]672.3059↑# | $, >2[fn], >30[fa], 1[fw] | Sulfuric acid conjugates are lost in fractions (A) and (B) following GP derivatisation. #Predicted for CESD. $GlcNAc conjugates not found in all patient samples. [fn]Concentration ratio for NPC to NIST SRM 1950. [fa]Concentration ratios for ASMD to NIST SRM 1950. [fw]Concentration ratios for Wolman to NIST SRM. 195. | Alvelius 2001, Mazzacuva 2016, ([f,n,a,w]Present work) |
| | | | | | 3β-Hydroxy-7β-*N*- acetylglucosaminyl-chol-5-en-24-oyl-taurine | 3βH,7β-GlcNAc-Δ$^5$-BA-24-Tau (3β,7β-diH-Δ$^5$-BA-24-Tau-7β-GlcNAc)↑$ | [-]699.3532↑ | [-]C34H55N2O11S | NA | (C)- | [-]699.3532↑# | $ | Taurine conjugates are lost in fractions (A) and (B) following GP derivatisation. #Predicted for CESD. | Mazzacuva 2016, Present work |
| | | | | | 3β-Sulfooxy-7β-*N*- acetylglucosaminyl-chol-5-en-24-oyl-glycine | 3βS,7β-GlcNAc-Δ$^5$-BA-24-Gly (3β,7β-diH-Δ$^5$-BA-24-Gly-3βS-7β-GlcNAc)↑$** | [-]729.3274↑** | [-]C34H53N2O13S | LMST05050052 | (C)- | [-]729.3274↑**# | $, >10[fn], >100[fa], 5[fw] | **Mixture of [M-H]$^-$ and [M-2H]$^{2-}$ ions produced dependent on source parameters. Sulfuric acid conjugates are lost in fractions (A) and (B) following GP derivatisation. #Predicted for CESD. $GlcNAc conjugates not found in all patient samples. | Alvelius 2001, ([f,n,a,w]Present work) |
| | | | | | | 3βS,7β-GlcNAc-Δ$^5$-BA-24-Gly (3β,7β-diH-Δ$^5$-BA-24-Gly-3βS-7β-GlcNAc)↑$** | [2-]364.1601↑** | [2-]C34H52N2O13S | LMST05050052 | (C)- | [2-]364.1601↑**# | $ | **Mixture of [M-H]$^-$ and [M-2H]$^{2-}$ ions produced dependent on source parameters. Sulfuric acid conjugates are lost in fractions (A) and (B) following GP derivatisation. #Predicted for CESD. $GlcNAc conjugates not found in all patient samples. | Alvelius 2001, Present work |
| | | | | | 3β-Sulfooxy-7β-*N*- acetylglucosaminyl-chol-5-en-24-oyl-taurine | 3βS,7β-GlcNAc-Δ$^5$-BA-24-Tau (3β,7β-diH-Δ$^5$-BA-24-Tau-3βS-7β-GlcNAc)↑$** | [-]779.3100↑** | [-]C34H55N2O14S2 | LMST05050053 | (C)- | [-]779.3100↑**# | $ | **Mixture of [M-H]$^-$ and [M-2H]$^{2-}$ ions produced dependent on source parameters. Sulfuric acid conjugates are lost in fractions (A) and (B) following GP derivatisation. #Predicted for CESD. $GlcNAc conjugates not found in all patient | Alvelius 2001, Present work |
| | | | | | | 3βS,7β-GlcNAc-Δ$^5$-BA-24-Tau (3β,7β-diH-Δ5-BA-24-Tau-3βS-7β-GlcNAc)↑$** | [2-]389.1513↑** | [2-]C34H54N2O14S2 | LMST05050053 | (C)- | [2-]389.1513↑**# | $ | **Mixture of [M-H]- and [M-2H]2- ions produced dependent on source parameters. Sulfuric acid conjugates are lost in fractions (A) and (B) following GP derivatisation. #Predicted for CESD. $GlcNAc conjugates not found in all patient | Alvelius 2001, Prersent work |
| 3.5.2 | | *LIPA/CH25H* | *613497/604551* | Lysosomal acid lipase (LAL) deficiency with cholesterol 25-hydroxylase deficiency | | | Positive-ion | | | | | | | |
| | | | | | Cholest-5-ene-3β,25-diol (25-Hydroxycholesterol) see also Wolman disease | 25-HC↓ | [$^2H_5$]GP 539.4368↓ | [+]C34H47D5N3O2 | LMST01010018 | (A)+ | | | 25-HC is absent in Lysosomal acid lipase deficiency with cholesterol 25-hydroxylase deficiency. Otherwise the oxysterol pattern is similar to classical Wolman disease. | Goenka 2017 |
| | | | | | | | Negative -ion | | | | | | | |
| | | | | | See Wolman disease | | | | | | | | Bile acid profile as classical Wolman disease. | |
| 3.6 | 210250 and 618666 | *ABCG8 and ABCG5* | *605460 and 605459* | Sitosterolaemia-1 (phytosterolemia) and Sitosterolaemia-2 (phytosterolemia) | | | Positive-ion | | | | Positive-ion | | | |
| | | | | | 24(R)Methylcholesta-5,22*E*-dien-3β-ol (Brassicasterol) | Brassicasterol↑ | [$^2H_5$]GP 535.4419↑ | [+]C35H47D5N3O | LMST01030098 | (A)+ | | >25† | †Average concentration ratio for two probands to NIST SRM 1950. | Rees 2005, (†Present work) |
| | | | | | 24(R)Methylcholest-5-en-3β-ol (Campesterol) | Campesterol↑ | [$^2H_5$]GP 537.4575↑ | [+]C35H49D5N3O | LMST01030097 | (A)+ | | >15†, >20* | †Average concentration ratio for two probands to NIST SRM 1950. *Concentration ratio of sitosterolemia to laboratory normal. | (*Rees 2005), (†Present work) |
| | | | | | 24(S)Ethylcholesta-5,22*E*-dien-3β-ol (Stigmasterol) | Stigmasterol↑ | [$^2H_5$]GP 549.4575↑ | [+]C36H49D5N3O | LMST01040123 | (A)+ | | 50†, >20* | †Average concentration ratio for two probands to NIST SRM 1950. *Concentration ratio of sitosterolemia to laboratory normal. | (*Rees 2005), (†Present work) |
| | | | | | 24Z-Ethylidene-cholest-5-en-3β-ol (Isofucosterol) | Isofucosterol↑ | [$^2H_5$]GP 549.4575↑ | [+]C36H49D5N3O | LMST01040145 | (A)+ | | >20* | *Concentration ratio of sitosterolemia to laboratory normal. | (*Rees 2005) |
| | | | | | 24(R)Ethylcholest-5-en-3β-ol (Sitosterol) | Sitosterol↑ | [$^2H_5$]GP 551.4732↑ | [+]C36H51D5N3O | LMST01040129 | (A)+ | | >50†, >20* | †Average concentration ratio for two probands to NIST SRM 1950. *Concentration ratio of sitosterolemia to laboratory normal. | (*Rees 2005), (†Present work) |

Table S2. Cholesterol Related Disorders, Dysregulated Metabolites and Simplest Mass Spectrometry Analysis

| Section | OMIM Disorder | Gene | OMIM Gene | Disorder (Short name, Abbreviation) | Simplest EADSA Diagnostic | Simplest Diagnostic Markers | *m/z* using $[^{2}H_{5}]$GP | *m/z* using $[^{0}H_{5}]$GP | LipidMaps ID |
|---|---|---|---|---|---|---|---|---|---|
| 3.1.3 | 302960 and 300960 | *EBP* | *300205* | Chondrodysplasia punctata 2 X-linked dominant (CDPX2) and Male EBP disorder with neurologic defects (MEND) | | | Positive-ion | | |
| | | | | | SPE1 unnecessary, A/B fractionation unnecessary, Cholesterol oxidase, GP-derivatisation | 8-DHC↑ | $[^{2}H_{5}]$GP 521.42624↑ | $[^{2}H_{0}]$GP 516.39484↑ | LMST01010242 |
| | | | | | | Zymostenol↑ | $[^{2}H_{5}]$GP 523.44189↑ | $[^{2}H_{0}]$GP 518.41049↑ | LMST01010096 |
| | | | | | | | Positive-ion | | |
| 3.1.2 | 607330 | *SCD5* | *602286* | Lathosterolosis | SPE1 unnecessary, A/B fractionation unnecessary, Cholesterol oxidase, GP-derivatisation | 8-DHC↓ | $[^{2}H_{5}]$GP 521.42624↓ | $[^{2}H_{0}]$GP 516.39484↓ | LMST01010242 |
| | | | | | | 24-DHC (Des)↓ | $[^{2}H_{5}]$GP 521.42624↓ | $[^{2}H_{0}]$GP 516.39484↓ | LMST01010016 |
| | | | | | | (Lath)↑ | $[^{2}H_{5}]$GP 523.44189↑ | $[^{2}H_{0}]$GP 518.41049↑ | LMST01010089 |
| 3.1.1 | 270400 | *DHCR7* | *602858* | Smith-Lemli-Opitz syndrome (SLOS) | | | Positive-ion | | |
| | | | | | SPE1 unnecessary, A/B fractionation unnecessary, Cholesterol oxidase, GP-derivatisation | 7-DHC ↑ | $[^{2}H_{5}]$GP 521.42624↑ | $[^{2}H_{0}]$GP 516.39484↑ | LMST01010069 |
| | | | | | | 8-DHC ↑ | $[^{2}H_{5}]$GP 521.42624↑ | $[^{2}H_{0}]$GP 516.39484↑ | LMST01010242 |
| | | | | | | (Chol)↓ | $[^{2}H_{5}]$GP 523.44189↓ | $[^{2}H_{0}]$GP 518.41049↓ | LMST01010001 |
| 3.1.3 | 602398 | *DHCR24* | *606418* | Desmosterolosis | | | Positive-ion | | |
| | | | | | SPE1 unnecessary, A/B fractionation unnecessary, Cholesterol oxidase, GP-derivatisation, | 24-DHC (Des)↑ | $[^{2}H_{5}]$GP 521.42624↑ | $[^{2}H_{0}]$GP 516.39484↑ | LMST01010016 |
| 3.2.2 | | *CYP7A1* | *118455* | Cholesterol 7α-hydroxylase (CYP7A1) deficiency | | | Positive-ion | | |
| | | | | | A/B fractionation unnecessary, Cholesterol oxidase, GP-derivatisation, | 7α-HCO (C4)↓ | $[^{2}H_{5}]$GP 539.4368↓ | $[^{2}H_{0}]$GP 534.4054↓ | LMST04030123 |
| | | | | | | 7α-HC ↓ | $[^{2}H_{5}]$GP 539.4368↓ | $[^{2}H_{0}]$GP 534.4054↓ | LMST01010013 |
| 3.2.1 | 213700 | *CYP27A1* | *606530* | Cerebrotendinous xanthomatosis (CTX) | | | Positive-ion | | |
| | | | | | A/B fractionation unnecessary, GP-derivatisation only | 7αH,3O-$\Delta^{4}$-BA↓ | $[^{2}H_{5}]$GP 527.36403↓ | $[^{2}H_{0}]$GP 522.33263↓ | LMST04010216 |
| | | | | | | 7α-HCO↑ | $[^{2}H_{5}]$GP 539.4368↑ | $[^{2}H_{0}]$GP 534.4054↑ | LMST04030123 |
| | | | | | | 7α,12α-diHCO↑ | $[^{2}H_{5}]$GP 555.43172↑ | $[^{2}H_{0}]$GP 550.40032↑ | LMST04030114 |
| | | | | | | 7αH,3O-CA(25R/S) (HOCA)↓ | $[^{2}H_{5}]$GP 569.41098↓ | $[^{2}H_{0}]$GP 564.37958↓ | LMST04030242 |
| | | | | | | | Negative-ion | | |
| | | | | | Switch to negative ion mode | 7α,12α,25-triHc-3αGlcA (3α,7α,12α,25-tetrol-3αGlcA)↑ | [-]611.3801↑ | [-]611.3801↑ | LMST05010042 |
| 3.2.3 | 270800 and 613812 | *CYP7B1* | *603711* | Spastic paraplegia 5A (SPG5A) and Oxysterol 7α-hydroxylase deficiency (O7AHD) | | | Positive-ion | | |
| | | | | | A/B fractionation unnecessary, Cholesterol oxidase, GP-derivatisation | 26-HC↑ | $[^{2}H_{5}]$GP 539.4368↑ | $[^{2}H_{0}]$GP 534.4054↑ | LMST01010088 |
| | | | | | | 25-HC↑ | $[^{2}H_{5}]$GP 539.4368↑ | $[^{2}H_{0}]$GP 534.4054↑ | LMST01010018 |
| | | | | | | 3β-HCA↑ | $[^{2}H_{5}]$GP 553.41607↑ | $[^{2}H_{0}]$GP 548.38467↑ | LMST04030072 |
| | | | | | | 3βH-$\Delta^{5}$-BA↑ | $[^{2}H_{5}]$GP 511.3691↑ | $[^{2}H_{0}]$GP 506.3377 | LMST04010201 |
| 3.2.4. | 607765 | *HSD3B7* | *607764* | 3β-Hydroxy-$\Delta^{5}$-$C_{27}$-steroid oxidoreductase (HSD3B7) deficiency (3β-Dehydrogenase deficiency) | | | Positive-ion | | |
| | | | | | A/B fractionation unnecessary, GP-derivatisation only | 7αH,3O-$\Delta^{4}$-BA↓ | $[^{2}H_{5}]$GP 527.36403↓ | $[^{2}H_{0}]$GP 522.33263↓ | LMST04010216 |
| | | | | | | 7αH,3O-CA(25R/S)↓ | $[^{2}H_{5}]$GP 569.41098↓ | $[^{2}H_{0}]$GP 564.37958↓ | LMST04030242 |
| 3.2.5. | 235555 | *AKR1D1* | *604741* | $\Delta^{4}$-3-Oxosteroid-5β-reductase (ACOX2) deficiency (5β-Reductase deficiency) | | | Positive-ion | | |
| | | | | | A/B fractionation unnecessary, GP-derivatisation only | 7αH,3O-$\Delta^{4}$-BA↑ | $[^{2}H_{5}]$GP 527.36403↑ | $[^{2}H_{0}]$GP 522.33263↑ | LMST04010216 |
| | | | | | | 7αH-27-*nor* -C-3,24-diO↑ | $[^{2}H_{5}]$GP 539.40042↑ | $[^{2}H_{0}]$GP 534.36902↑ | NA |
| | | | | | | 7α,12α-diH,3O-$\Delta^{4}$-BA↑ | $[^{2}H_{5}]$GP 543.35895↑ | $[^{2}H_{0}]$GP 538.32755↑ | LMST04010241 |
| | | | | | | 7αH,3O-CA(25R/S)↑ | $[^{2}H_{5}]$GP 569.41098↑ | $[^{2}H_{0}]$GP 564.37958↑ | LMST04030242 |
| | | | | | | 7αH,3,24-diO-CA(25R)↑ | $[^{2}H_{5}]$GP 583.39025↑ | $[^{2}H_{0}]$GP 578.35885↑ | LMST04030255 |
| | | | | | | 7α,12α-diH,3O-CA(25R/S)↑ | $[^{2}H_{5}]$GP 585.4059↑ | $[^{2}H_{0}]$GP 580.37450↑ | LMST04030150 |
| 3.3.1 | | *SLC27A5* | *603314* | Bile acyl CoA synthetase (BACS) deficiency | | | Negative-ion | | |
| | | | | | Fraction C in negative-ion mode | 3α,7α-diHBA (CDCA)↑ | [-]391.2854↑ | [-]391.2854↑ | LMST04010032 |
| | | | | | | 3α,7β-diHBA (UDCA)↑ | [-]391.2854↑ | [-]391.2854↑ | LMST04010033 |
| | | | | | | 3α,7α-diHBA-24-Gly (GCDCA)↓ | [-]448.3068↓ | [-]448.3068↓ | LMST05030008 |
| | | | | | | 3α,7β-diHBA-24-Gly (GUDCA)↓ | [-]448.3068↓ | [-]448.3068↓ | LMST05030016 |
| | | | | | | 3α,7α,12α-triHBA-24-Gly (GCA)↓ | [-]464.3018↓ | [-]464.3018↓ | LMST05030001 |
| 3.3.2 | 614307 | *AMACR* | *604489* | Alpha-methylacyl-CoA recemase (AMACR) deficiency | | | Positive-ion | Positive-ion | |
| | | | | | A/B fractionation unnecessary, GP-derivatisation only | 7αH,3O-CA(25R)↑ | $[^{2}H_{5}]$GP 569.41098↑ | $[^{2}H_{0}]$GP 564.37958↑ | LMST04030242 |
| | | | | | | 7αH,3O-CA(25S)↓ | $[^{2}H_{5}]$GP 569.41098↓ | $[^{2}H_{0}]$GP 564.37958↓ | NA |
| | | | | | | 7α,12α-diH,3O-CA(25R)↑ | $[^{2}H_{5}]$GP 585.4059↑ | $[^{2}H_{0}]$GP 580.3745↑ | LMST04030243 |
| 3.3.3 | 617308 | *ACOX2* | *601641* | Acyl-CoA oxidase 2 (ACOX2) deficiency | | | Positive-ion | | |
| | | | | | A/B fractionation unnecessary, GP-derivatisation only | 7αH,3O-CA(25R/S)↑ | $[^{2}H_{5}]$GP 569.41098↑ | $[^{2}H_{0}]$GP 564.37958↑ | LMST04030242 |
| | | | | | | 7α,12α-diH,3O-CA(25S)↑ | $[^{2}H_{5}]$GP 585.4059↑ | $[^{2}H_{0}]$GP 580.37450↑ | LMST04030150 |
| 3.3.4 | 261515 | *HSD17B4* | *601860* | D-Bifunctional protein (DBP) deficiency type I & II & III | | | Positive-ion | | |
| | | | | | A/B fractionation unnecessary, GP-derivatisation only | 7αH,3O-$\Delta^{24E}$-CA↑ | $[^{2}H_{5}]$GP 567.39533↑ | $[^{2}H_{0}]$GP562.3639↑ | NA |
| | | | | | | 7αH,3O-CA(25S)↑ | $[^{2}H_{5}]$GP 569.41098↑ | $[^{2}H_{0}]$GP564.3796↑ | NA |
| | | | | | | 7α,24S-diH,3O-CA(25S)↑ | $[^{2}H_{5}]$GP 585.4059↑ | $[^{2}H_{0}]$GP580.3745↑ | LMST04030260 |
| | | | | | | 7α,24R-diH,3O-CA(25R)↑ | $[^{2}H_{5}]$GP 585.4059↑ | $[^{2}H_{0}]$GP580.3745↑ | LMST04030257 |
| 3.3.5 | 613724 | *SCP2* | *184755* | Sterol carrier protein 2 (SCP2) deficiency | | | Positive-ion | | |
| | | | | | A/B fractionation unnecessary, GP-derivatisation only | 7αH-27-*nor* -C-3,24-diO↑ | $[^{2}H_{5}]$GP 539.40042↑ | $[^{2}H_{5}]$GP534.369↑ | NA |
| | | | | | | 7αH,3,24-diO-CA(25R)↑ | $[^{2}H_{5}]$GP 583.39025↑ | $[^{2}H_{0}]$GP 578.35885↑ | LMST04030255 |
| 3.4 | 619232 | *BAAT* | *602938* | Bile acid-CoA: amino acid *N* -acyl transferase (BAAT) deficiency | | | Negative-ion | | |
| | | | | | Fraction C in negative-ion mode | 3α,7α,12α-triHBA (Cholic acid)↑ | [-]407.2803↑ | [-]407.2803↑ | LMST04010001 |
| | | | | | | 3α,7α-DiHBA-24-Gly (GCDCA)↓ | [-]448.3068↓ | [-]448.3068↓ | LMST05030008 |
| | | | | | | 3βS-$\Delta^{5}$-BA↑ | [-]453.2316↑ | [-]453.2316↑ | NA |
| | | | | | | 3α,7α,12α-triHBA-24-Gly (GCA)↓ | [-]464.3018↓ | [-]464.3018↓ | LMST05030001 |
| | | | | | | GlcA-HBA (CDCA-GlcA)↑ | [-] 567.3175↑ | [-] 567.3175↑ | LMST05010028 |
| | | | | | | GlcA-diHBA (Cholic acid-GlcA)↑ | [-]583.3124↑ | [-]583.3124↑ | LMST05010044 |
| 3.5 | 257220 and 607625 and 607616 and 620151 and 278000 | *NPC1 and NPC2 and SMPD1 and LIPA and LIPA* | *607623 and 601015 and 607608 and 613497 and 613497* | Niemann-Pick type C1 disease (NPC1) and Niemann-Pick type C2 disease (NPC2) and Acid sphingomyelinase deficiency (ASMD, Niemann Pick type B disease, NPB) and Wolman disease and Cholesterol ester storage disease (CESD) | | | Positive-ion | | |
| | | | | | A/B fractionation unnecessary, Cholesterol oxidase, GP-derivatisation | SM(d18:1/0:0)↑ | [+]465.3452↑ | [+]465.3452↑ | LMSP01060001 |
| | | | | | | PPCS (lyso-SM509)↑ | [+]509.3350↑ | [+]509.3350↑ | LMFA08020401 |
| | | | | | | 3β,5α,6β-triHBA-18↑ | $[^{2}H_{5}]$GP 527.36403↑ | $[^{2}H_{0}]$GP 522.33263↑ | LMST04010339 |
| | | | | | | 3β,7β-diH-$\Delta^{5}$-BA↑ | $[^{2}H_{5}]$GP 527.36403↑ | $[^{2}H_{0}]$GP 522.33263↑ | LMST04010280 |
| | | | | | | 3β,5α,6β-triol-18↑ | $[^{2}H_{5}]$GP 539.4368↑ | $[^{2}H_{0}]$GP 534.4054↑ | LMST01010052 |
| | | | | | | 7β-HC↑ | $[^{2}H_{5}]$GP 539.4368↑ | $[^{2}H_{0}]$GP 534.4054↑ | LMST01010047 |
| | | | | | | 3β,5α,6β-triol (C-triol)↑ | $[^{2}H_{5}]$GP 557.44737↑ | $[^{2}H_{0}]$GP 552.41597↑ | LMST01010052 |
| | | | | | | 3β,5α,6β-triHCA-18↑ | $[^{2}H_{5}]$GP 569.41098↑ | $[^{2}H_{0}]$GP 564.3796↑ | LMST04010339 |
| | | | | | | 3β,7β-diHCA(25R/S)↑ | $[^{2}H_{5}]$GP 569.41098↑ | $[^{2}H_{0}]$GP 564.3796↑ | LMST04030235 |
| 3.5.2 | | *LIPA/CH25H* | *613497/604551* | Lysosomal acid lipase (LAL) deficiency with cholesterol 25-hydroxylase deficiency | | | Positive-ion | | |
| | | | | | A/B fractionation unnecessary, Cholesterol oxidase, GP-derivatisation | 25-HC↓ | $[^{2}H_{5}]$GP 539.4368↓ | $[^{2}H_{0}]$GP 534.4054↓ | LMST01010018 |
| | | | | | | Plus other Wolman diagnostics | | | |
| 3.6 | 210250 and 618666 | *ABCG8 and ABCG5* | *605460 and 605459* | Sitosterolaemia-1 (phytosterolemia) and Sitosterolaemia-2 (phytosterolemia) | | | Positive-ion | | |
| | | | | | SPE1 unnecessary, A/B fractionation unnecessary, Cholesterol oxidase, GP-derivatisation, | Brassicasterol↑ | $[^{2}H_{5}]$GP 535.4419↑ | $[^{2}H_{0}]$GP 530.4105↑ | LMST01030098 |
| | | | | | | Campesterol↑ | $[^{2}H_{5}]$GP 537.4575↑ | $[^{2}H_{0}]$GP 532.4261↑ | LMST01030097 |
| | | | | | | Stigmasterol↑ | $[^{2}H_{5}]$GP 549.4575↑ | $[^{2}H_{0}]$GP 544.4261↑ | LMST01040123 |
| | | | | | | Isofucosterol↑ | $[^{2}H_{5}]$GP 549.4575↑ | $[^{2}H_{0}]$GP 544.4261↑ | LMST01040145 |
| | | | | | | Sitosterol↑ | $[^{2}H_{5}]$GP 551.4732↑ | $[^{2}H_{0}]$GP 546.4418↑ | LMST01040129 |
